# Genomic profiling defines variable clonal relatedness between invasive breast cancer and primary ductal carcinoma *in situ*

**DOI:** 10.1101/2021.03.22.21253209

**Authors:** Esther H. Lips, Tapsi Kumar, Anargyros Megalios, Lindy L. Visser, Michael Sheinman, Angelo Fortunato, Vandna Shah, Marlous Hoogstraat, Emi Sei, Diego Mallo, Maria Roman-Escorza, Ahmed A. Ahmed, Mingchu Xu, Wim Brugman, Karen Clements, Helen R. Davies, Liping Fu, Anita Grigoriadis, Timothy M. Hardman, Lorraine M. King, Marielle Krete, Petra Kristel, Michiel de Maaker, Carlo C. Maley, Jeffrey R. Marks, Brian Menegaz, Lennart Mulder, Frank Nieboer, Salpie Nowinski, Sarah Pinder, Jelmar Quist, Carolina Salinas-Souza, Michael Schaapveld, Marjanka K. Schmidt, Abeer M. Shaaban, Rana Shami, Mathini Sridharan, John Zhang, Hilary Stobart, Deborah Collyar, Serena Nik-Zainal, Lodewyk F.A. Wessels, E. Shelley Hwang, Nicholas N. Navin, Andrew Futreal, Alastair Thompson, Jelle Wesseling, Elinor J. Sawyer, on behalf of the Grand Challenge PRECISION consortium

## Abstract

Pure ductal carcinoma *in situ* (DCIS) is being diagnosed more frequently through breast screening programmes and is associated with an increased risk of developing invasive breast cancer. We assessed the clonal relatedness of 143 cases of pure DCIS and their subsequent events using a combination of whole exome, targeted and copy number sequencing, supplemented by single cell analysis. Unexpectedly, 18% of all invasive events after DCIS were clonally unrelated to the primary DCIS. Single cell sequencing of selected pairs confirmed our findings. In contrast, synchronous DCIS and invasive disease (n=44) were almost always (93%) clonally related. This challenges the dogma that almost all invasive events after DCIS represent invasive transformation of the initial DCIS and suggests that DCIS could be an independent risk factor for developing invasive disease as well as a precursor lesion. Our findings support a paradigm shift that confirms a more complex role for DCIS than previously recognized, and that the future management of DCIS should take into account both the precursor and risk factor implications of this diagnosis.

## Introduction

Approximately 60% of invasive breast cancers of ductal/no special type are associated with synchronous ductal carcinoma *in situ* (DCIS) ^1^. The majority of molecular studies looking at these two synchronous components suggest that they are clonally related supporting the hypothesis that DCIS is a non-obligate precursor of invasive breast cancer ^2,3^. Recent single cell analysis of DCIS that presented with synchronous invasive disease has shown that most mutations and copy number aberrations have already evolved within DCIS prior to invasion and suggest that multiple clones escape from the *in situ* component into the adjacent tissues to establish invasive carcinomas ^4^.

Less is known about the genomic evolution of invasive disease that occurs after pure DCIS (i.e., DCIS without an invasive component) that has been treated with surgery with or without radiotherapy. Pure DCIS is being diagnosed more frequently through breast screening programmes and now accounts for ∼20% of screen detected breast cancers. It carries a 4-to 10-fold increased risk of invasive breast cancer with the highest risk being in women under the age of 50 ^5-8^. It is presumed that the majority of these invasive lesions are clonally related to the initial DCIS, particularly if treated with surgery alone. However, analysis of clonal relatedness of subsequent invasive cancer arising after a diagnosis of pure DCIS has not been widely performed, due to the difficulty of obtaining samples from a large cohort of women with DCIS with adequate follow-up. Previously tumour grade, morphology and immunohistochemistry have been shown to differ between DCIS and subsequent invasive cancer, but it is not clear whether this is a result of tumour progression or represents the development of a new primary invasive cancer ^9,10^.

It is important to understand the clonal relatedness of DCIS and subsequent invasive disease in order to assess the true recurrence rate of DCIS after different treatment modalities and to be able to design effective strategies, not only to reduce subsequent diagnoses but also overtreatment. To investigate the clonal relatedness between pure DCIS and subsequent events, we pooled samples from three countries resulting in the largest cohort to date of DCIS cases with 5-17 years follow up and thus adequate time to develop subsequent events (Supplementary Table 1).

## Results

143 primary DCIS and their subsequent events were analysed. The median age at diagnosis of primary DCIS was 57 years (range 34-87 years) and median time to subsequent event was 4 years (0.4-17.5years). 54% were high grade, 67% were ER positive (ER+), 24% were ER negative (ER-), 28% Her2 positive and 46% Her2 negative.

We studied three different types of subsequent events after a diagnosis of pure DCIS:

1. Pure DCIS that developed a subsequent ipsilateral invasive event (**DCIS->ipINV**; n=95)
2. Pure DCIS that developed a subsequent ipsilateral DCIS event (**DCIS->ipDCIS**; n=34).
3. Pure DCIS that developed a subsequent contralateral invasive event (**DCIS->contraINV**; n=14)

We also analysed 44 pairs of synchronous DCIS and invasive disease (**synDCIS&INV**) in order to assess whether clonal relatedness rates differed between synchronous and subsequent invasive disease (Figure 1, Supplementary Table 2 for summary of sample pair type, origin and molecular characterisation, Supplementary Tables 3a-c for clinical characteristics).

**Figure 1.**
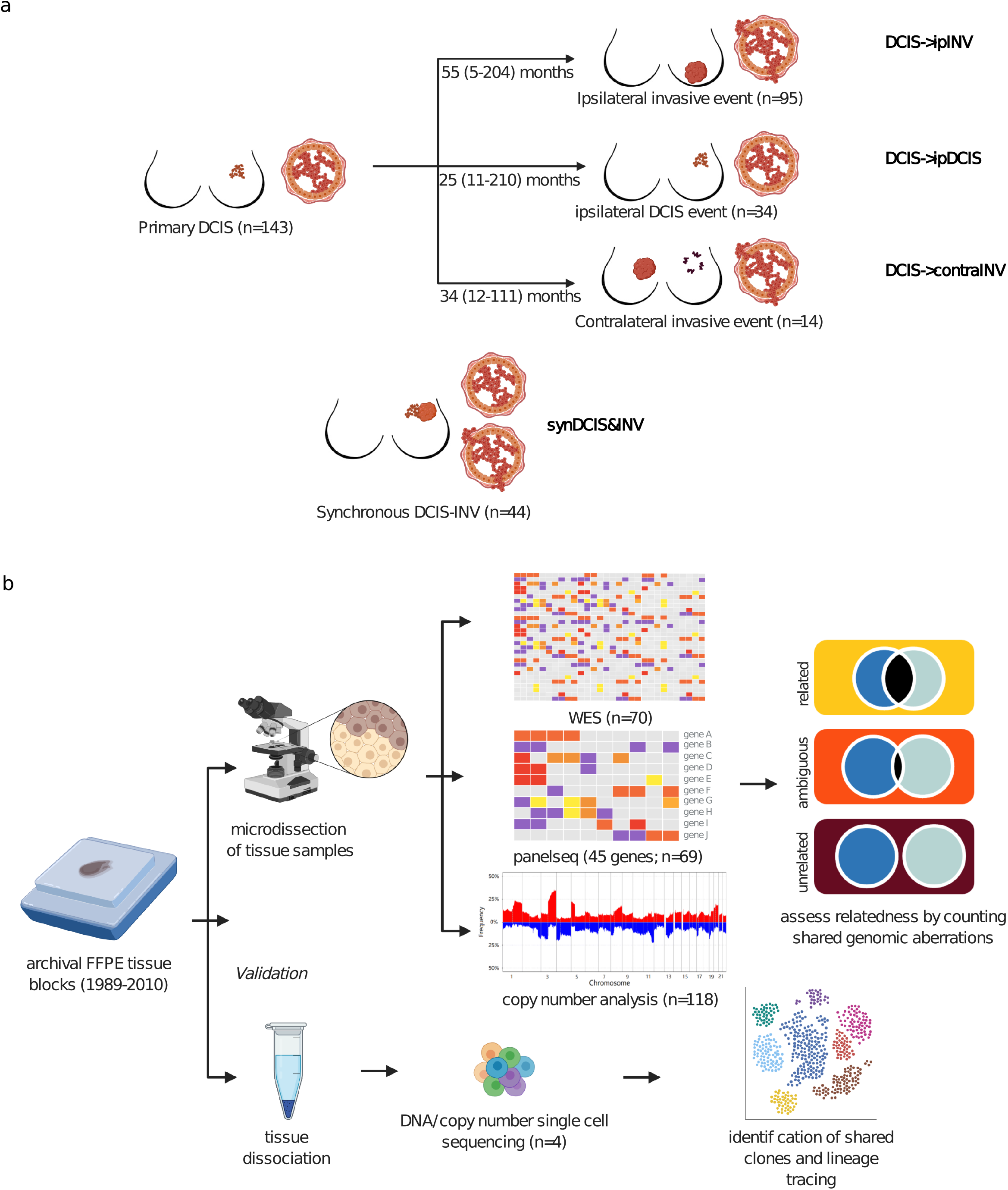
Study design. **a**, Graphical representation of our clinical cohort with long-term follow up to study clonal relatedness between primary DCIS and subsequent disease. The three different groups of subsequent events (DCIS*→*ipINV, DCIS*→*ipDCIS and DCIS*→*contraINV) are shown, together with sample numbers and the median time to follow up for the three respective groups. In addition, a group of synchronous pairs (synDCIS&INV) was studied. **b**, Representation of the two different strategies undertaken to unravel clonality in DCIS with subsequent disease. In the first approach (top), tissue from a large cohort of DCIS-subsequent event pairs was microdissected and analysed with WES, PanelSeq and copy number profiling. In the second approach (bottom), tissue of paired lesions was dissociated, followed by single cell sequencing to study the clonal composition.

### Genomic features of primary DCIS and subsequent invasive disease

Whole exome sequencing (WES) (n=70), targeted sequencing (PanelSeq) (n=69) and copy number analysis (n=118) revealed that pure DCIS and subsequent invasive disease (INV, where INV includes both ipINV and contraINV) share similar genomic profiles (Figure 2).

**Figure 2.**
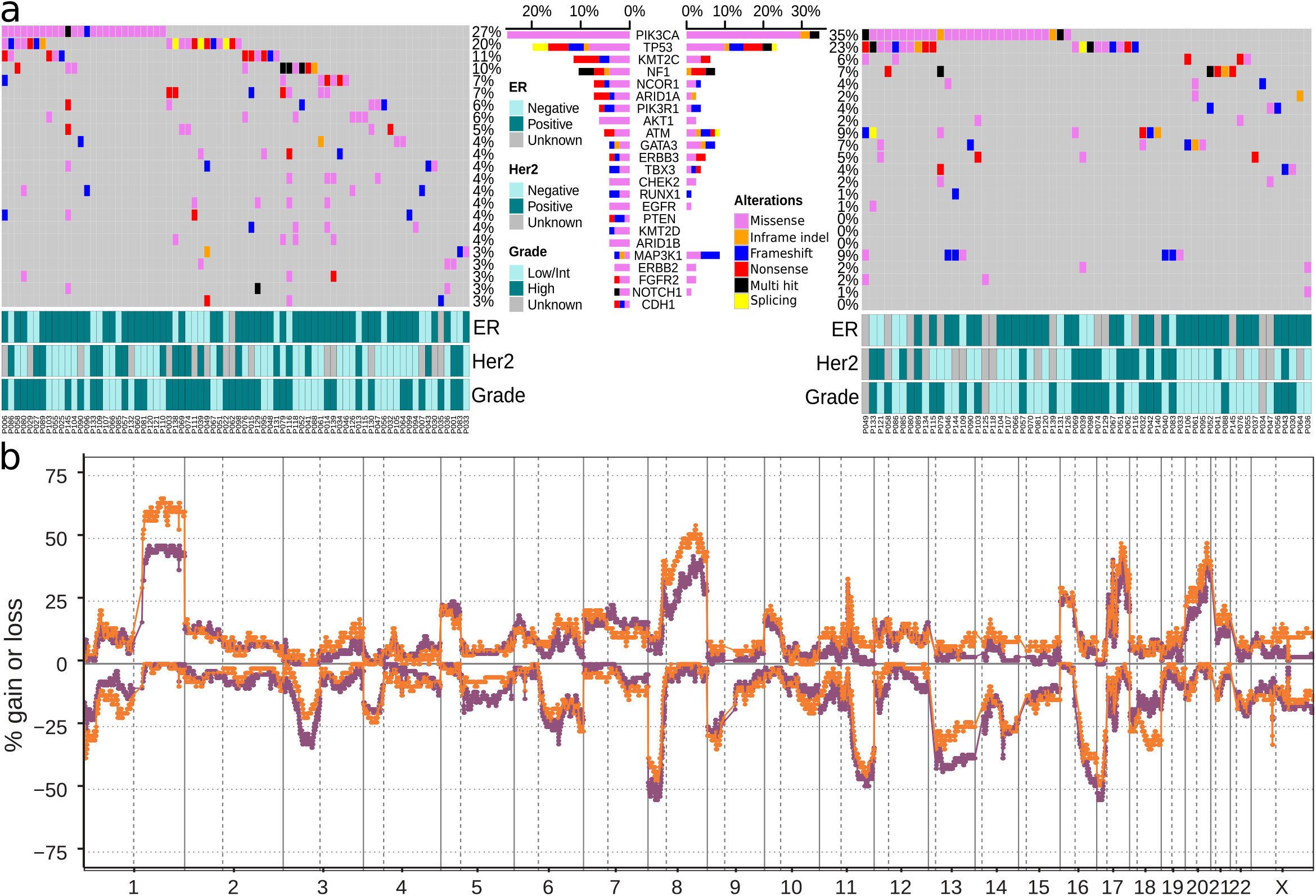
Mutational spectrum and copy number alterations in primary DCIS and subsequent invasive disease. **a**, Oncoplots for primary DCIS samples (left) and invasive 2nd events (right) based on WES and targeted sequencing. Of the 45 genes covered by all sequencing platforms, only genes mutated in more than 3% of the primary DCIS or invasive 2nd event samples are shown. We removed C>T mutations with AF*<* 0.1 and fewer than 3 appearances in the COSMIC database. **b**, Frequency plot of genome wide copy number alterations based on lpWGS (*n* = 128) of primary DCIS samples (purple, *n* = 72) and subsequent invasive disease (orange, *n* = 56). The *y*-axis shows the percentage of samples with gains (positive) and losses (negative). The horizontal axis represents the genomic position with chromosome indices indicated and chromosome boundaries marked by vertical solid lines.

There were no genes that were significantly more frequently mutated in subsequent invasive disease compared to pure DCIS among the 45 driver genes that were common to both targeted sequencing panels and WES (Figure 2a). The most frequently mutated genes were: *PIK3CA* (27% DCIS, 35% INV), *TP53* (20% DCIS, 23% INV), *KMT2C* (11% DCIS, 6% INV) and *NF1*(10% DCIS, 7% INV). *TP53* mutations were more common in ER-negative DCIS (P=0.05, Fisher’s exact test) and HER2 positive DCIS (P=0.02, Fisher’s exact test) and this association was also seen in the subsequent invasive events (P=0.007 and P=0.008, respectively, Fisher’s exact test), Supplementary Table 4. Mutations in *TP53* were associated with high grade DCIS (P= 0.002, Fisher’s exact test). *PIK3CA* mutations were not associated with any specific subtype of DCIS.

Similarly, the frequency plots of gains and losses for DCIS and subsequent invasive disease were almost identical (Figure 2b, low-pass whole genome sequencing (lpWGS), DCIS n=72, INV n=56, and Supplementary Figure 1, SNP array, DCIS n=38, INV n=29) as were recurrent amplifications with 17q12 (*ERBB2*: 33% DCIS, 25% INV), 17q21.1 (*GSDMB, PSMD3*: 25% DCIS, 20% INV) and 11q13 (*CCND1, FGF3&4*: 19% DCIS, 20% INV) being the most common (Supplementary Table 5). The fraction of the genome altered overall did not differ significantly between DCIS and invasive disease (Supplementary Figure 2a). The SNP array data showed that 60% of DCIS and 66% of invasive disease were diploid (Supplementary Figure 2b). The copy number changes that were more frequent in invasive disease at a nominal p-value of 0.05 are listed in Supplementary Tables 6a & 6b, with the most significant differential copy number aberration being gain of 8q12 in samples that underwent lpWGS (DCIS: 11%, 8/72; INV: 34%, 19/56 P=0.002, Fisher’s Exact Test).

### Whole exome sequencing of primary DCIS and paired subsequent ipsilateral invasive events

Clonal relatedness was assessed using WES in 24 DCIS->ipINV pairs and in 34 synchronous DCIS&INV pairs, using Breakclone, an approach we developed to assess clonal relatedness, described in detail in the Methods. Unlike other packages ^11-13^ it incorporates both population frequency and allele frequency when using mutation data, and the position of the individual copy number aberration breakpoints when using copy number data. A reference distribution of concordance scores is calculated by randomly permuting all possible pairs from different patients and is used to calculate p-values for the concordance score of each tumour pair.

The numbers of shared and private mutations were highly variable for the different tumour pairs for both subsequent and synchronous pairs, ranging from 112 to 0 shared mutations (Figure 3a). Shared mutations had significantly higher allele frequencies compared to private mutations, suggesting they are more likely to be driver mutations, with the most common shared mutations being in *TP53* and *PIK3CA* (Supplementary Figure 3a,c). The invasive subsequent events had a higher number of private mutations than primary DCIS, (P=0.024), but this was not seen when we compared DCIS with paired synchronous IBC. There was also no difference in the number of mutations found in primary pure DCIS and DCIS that presents with synchronous invasive disease (Figure 3e).

**Figure 3.**
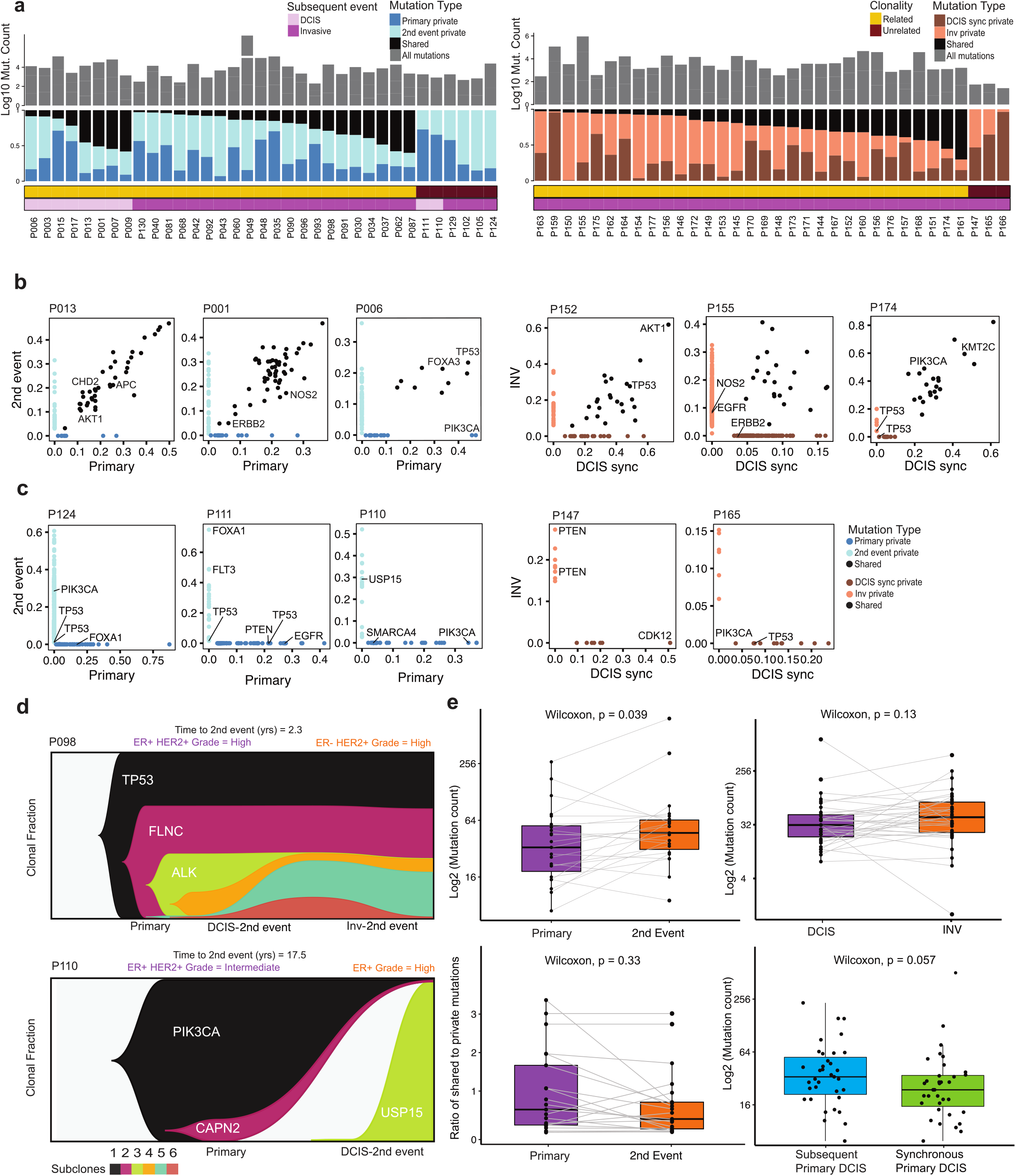
Clonality assessment using whole exome sequencing. **a**, Distribution of mutations across time points in subsequent (left) and synchronous DCIS-INV (right) pairs. The color bars indicate occurrence of mutations according to whether they are unique to the timepoint *i*.*e*. either private to the primary or 2nd event (light purple indicates DCIS is 2nd event and dark purple invasive) or shared by primary-2nd event timepoints for subsequent samples. Right colored bars indicate mutations based on DCIS or INV sites. Top grey bar indicates the log_10_ values of total mutations for each patient. **b**, Scatter plots showing the variant allele frequency of mutations in 3 clonally related pairs (subsequent) and 3 (synchronous). **c**, Similar as **b**, for clonally unrelated pairs. **d**, Clonal lineages showing change in frequency for 1 patient with primary DCIS and synchronous DCIS-INV recurrence and 1 clonally unrelated patient. **e**, (Clockwise) (Top left) Boxplots comparing mutation counts in 1. primary vs invasive 2nd event in subsequent samples (*p* = 0.039), (Top right) 2. DCIS *vs* invasive in synchronous pairs (*p* = 0.13), (Bottom right) 3. subsequent *vs* synchronous primary DCIS (*p* = 0.05) and (Bottom left) ratio of shared to private mutations between primary DCIS *vs* invasive recurrence in clonal related subsequent samples.

Twenty DCIS->ipINV pairs (83%) showed clear evidence of clonal relatedness including three cases of primary DCIS that developed an invasive event despite having undergone a mastectomy (Figure 3b, Supplementary Figure 3a). The remaining 4 DCIS->ipINV pairs (17%) did not carry any shared mutations (Figures 3a (left) and 3c (left)) that we could detect in our analyses, which could indicate that they are clonally unrelated. Evolutionary analysis confirmed this result (Figure 3d).

WES of synchronous DCIS-IBC pairs confirmed that most (31/34, 91%) showed clonal relatedness, with only three pairs not sharing any mutations (Figure 3a, right, Supplementary Figure 3b). Unlike the four unrelated DCIS->ipINV pairs, these three synchronous pairs also had the least number of mutations overall, which may suggest that methodological limitations are preventing us from detecting their common origin. There was no statistically significant association between the clonality score and physical distance between the DCIS and invasive component (Supplementary Figure 4).

**Figure 4.**
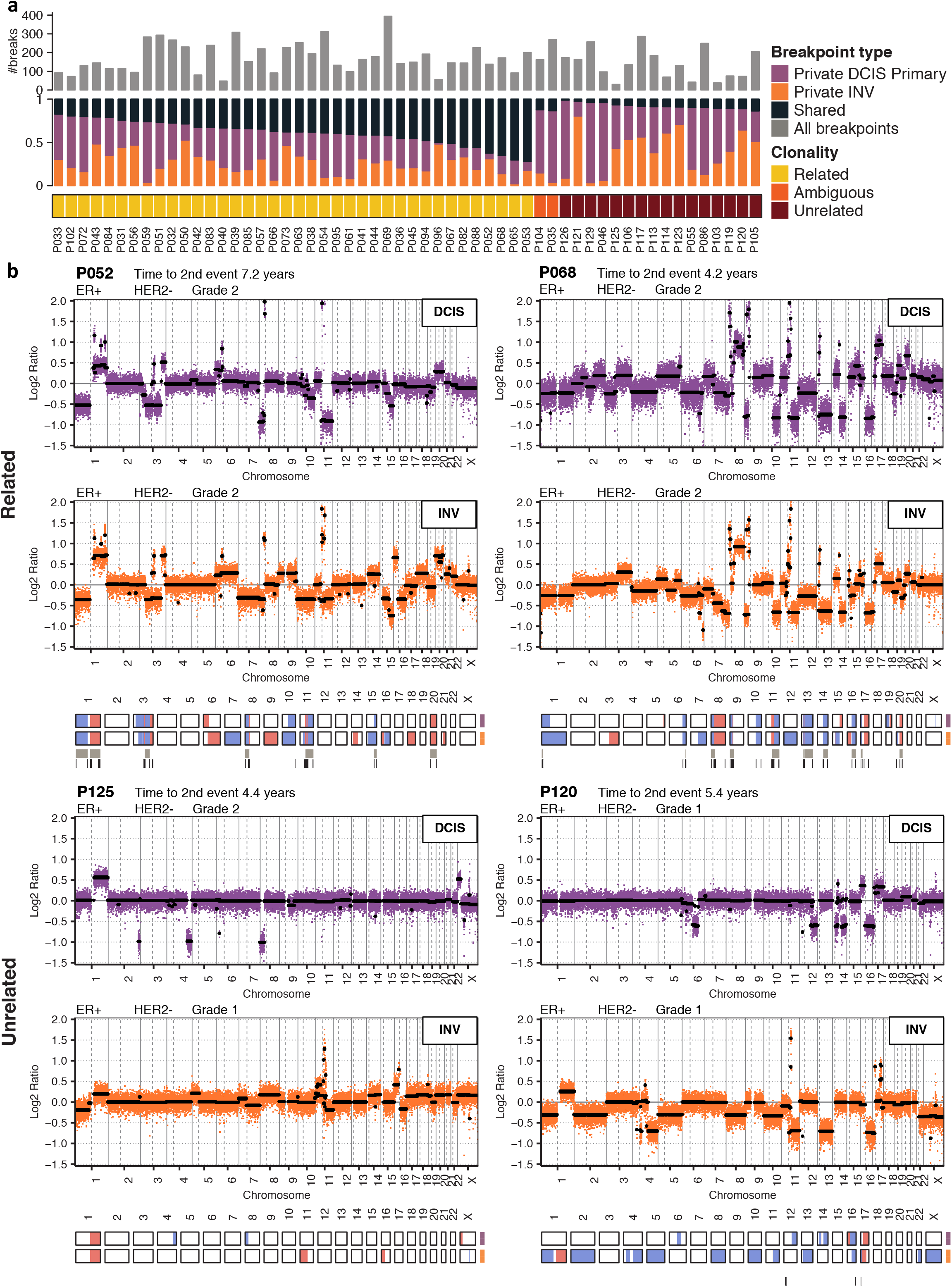
Validation of clonality using copy number profiling. **a**, Distribution of breakpoints in primary DCIS and subsequent invasive pairs (DCIS*→*ipINV) derived from lpWGS copy number profiles. The stacked bar graphs indicate whether the breakpoints were private to the primary (purple) or subsequent invasive tumour (orange) or shared (black). The grey bars at the top indicate the total number of breakpoints for each patient. **b**, Genome-wide segmented copy number profiles and associated heatmaps representing called aberrations (gains in red, losses in blue) of two related and two unrelated pairs. The copy number profiles illustrate the relatedness between the primary DCIS (purple) and its paired subsequent invasive event (orange) based on lpWGS copy number profiles. In the copy number profile plot, raw log ratios are depicted in colour and segmented log ratios in black. Below the heatmaps, shared aberration events (top bar) and shared breakpoints (bottom bar) between pairs are depicted. The genomic position is indicated by chromosome indices.

### Copy number and targeted sequencing confirm unrelated ipsilateral invasive events

In order to increase the sample size, we analysed a further 71 DCIS->ipINV pairs by copy number analysis using either SNP array or lpWGS data (Methods). Of the 62 that passed QC, 71% (44/62) were considered clonally related, 2% (1/62) ambiguous and 27% (17/62) unrelated (Figure 4, Supplementary Figure 5a). In 45 of the 71 pairs that underwent copy number analysis there was enough DNA to also perform targeted sequencing which revealed that 51% (23/45) were considered clonally related (including four considered unrelated by copy number) and 15% (7/45) unrelated (all supported by copy number data). A further 33% (15/45) were considered ambiguous, with only a single mutation identified and shared between both components. In 11 of these cases copy number data confirmed clonal relatedness. There were nine pairs that had both targeted sequencing data and WES, in seven of these, the results were concordant and in two a single mutation was found to be shared on targeted sequencing but not detected on WES. Inspection of raw WES data revealed that these mutations were present but had not passed QC thresholds.

**Figure 5.**
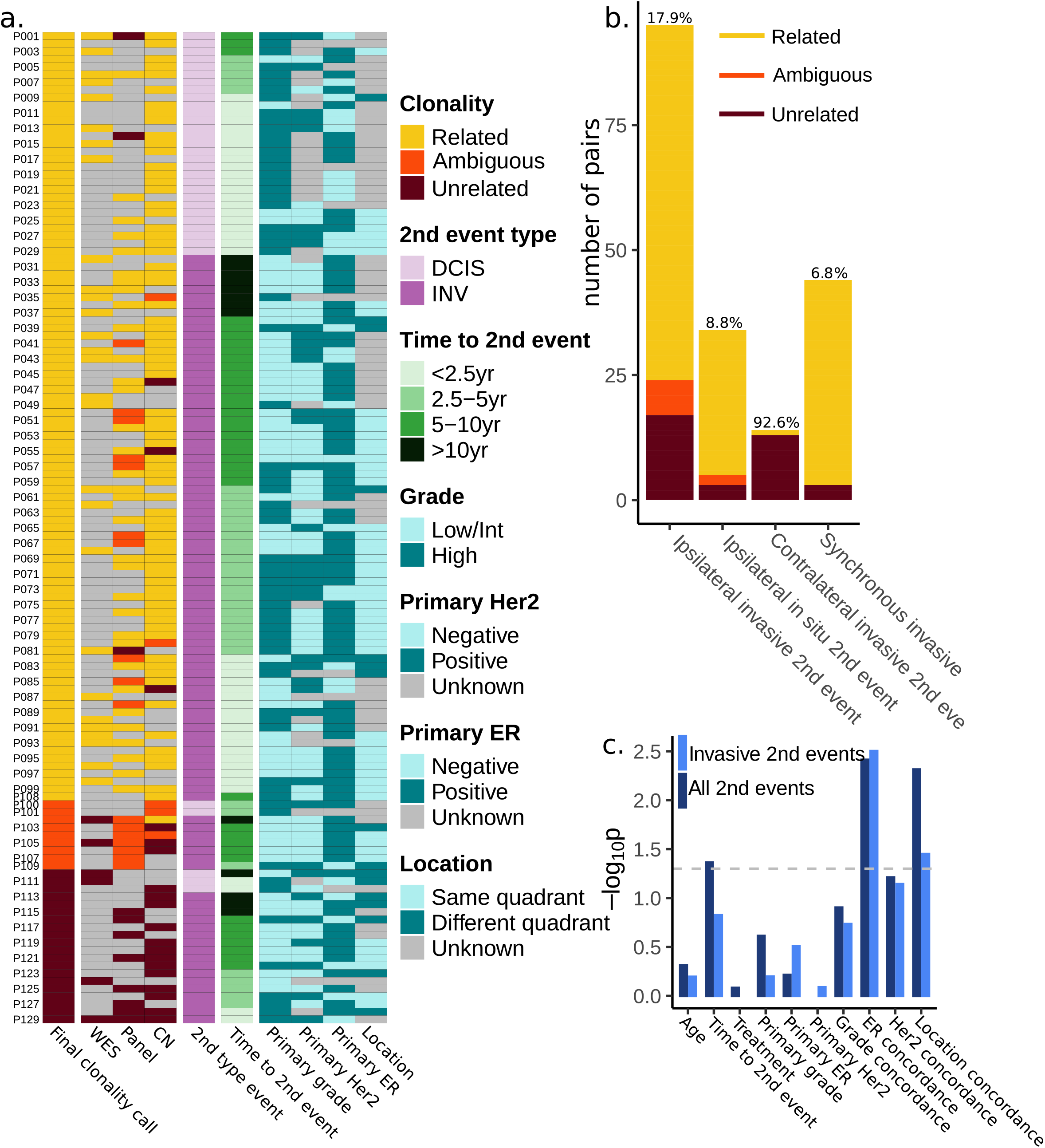
Combined clonality calls and clinical data. **a**, Summary of subsequent pairs with ipsilateral recurrences and their combined clonality calls together with clinical and histological characteristics. **b**, Comparison of clonality calls for all classes of subsequent and synchronous pairs. On top of the bars the percentage unrelated pairs is shown for each category. **c**, Association between clonality calls as well as clinical and histological data (see also Supplementary Table 7).

### Combined calls based on WES, Copy Number and PanelSeq show that 18% may be unrelated

We combined our relatedness calls based on WES, PanelSeq and copy number data to obtain a final verdict on the clonal relatedness of samples (Figure 5a). In case of conflicting data between platforms, relatedness prevailed over unrelatedness. Based on all samples, irrespective of analysis platform, we estimate the percentage of pairs to be clonally related as 75% (71/95), not related as 18% (17/95) and ambiguous as 7% (7/95; Figure 5a & b, Supplementary Table 3a). 70% (61/87) of subsequent ipsilateral invasive events with detailed histological data available presented with synchronous DCIS adjacent to the invasive disease. In 10 there was enough DCIS to analyse separately, (Supplementary Table 3c) and, in all cases, the subsequent synchronous DCIS was clonally related to the adjacent synchronous invasive tumours (8 by copy number, 2 by WES). However, the subsequent synchronous DCIS and invasive disease were only clonally related to the primary DCIS in 6 cases, indicating that new primary invasive tumours may arise from new independent DCIS lesions. Evolutionary analysis of a case where all three components had undergone WES revealed that the primary DCIS comprised four sub-clones, two of which were also seen in the synchronous DCIS and invasive disease, but the dominant clone in the invasive disease was a clone that appeared in the synchronous DCIS (Figure 3d, top panel). Combining the clonality results for these 10 pairs of synDCIS&INV with the 34 that had undergone WES we estimate the percentage of synDCIS&INV to be clonally related as 93% compared to 75% for subsequent ipsilateral invasive events (P=0.07, Fishers Exact Test).

Having identified which pairs were clonally related we were able to compare copy number aberrations (CNAs) and mutations between the primary DCIS and true invasive events and it was striking that the common amplifications and mutations seen in invasive breast cancer are already established in the primary DCIS and there were no clear genomic markers of invasive progression (Supplementary Figure 6).

**Figure 6.**
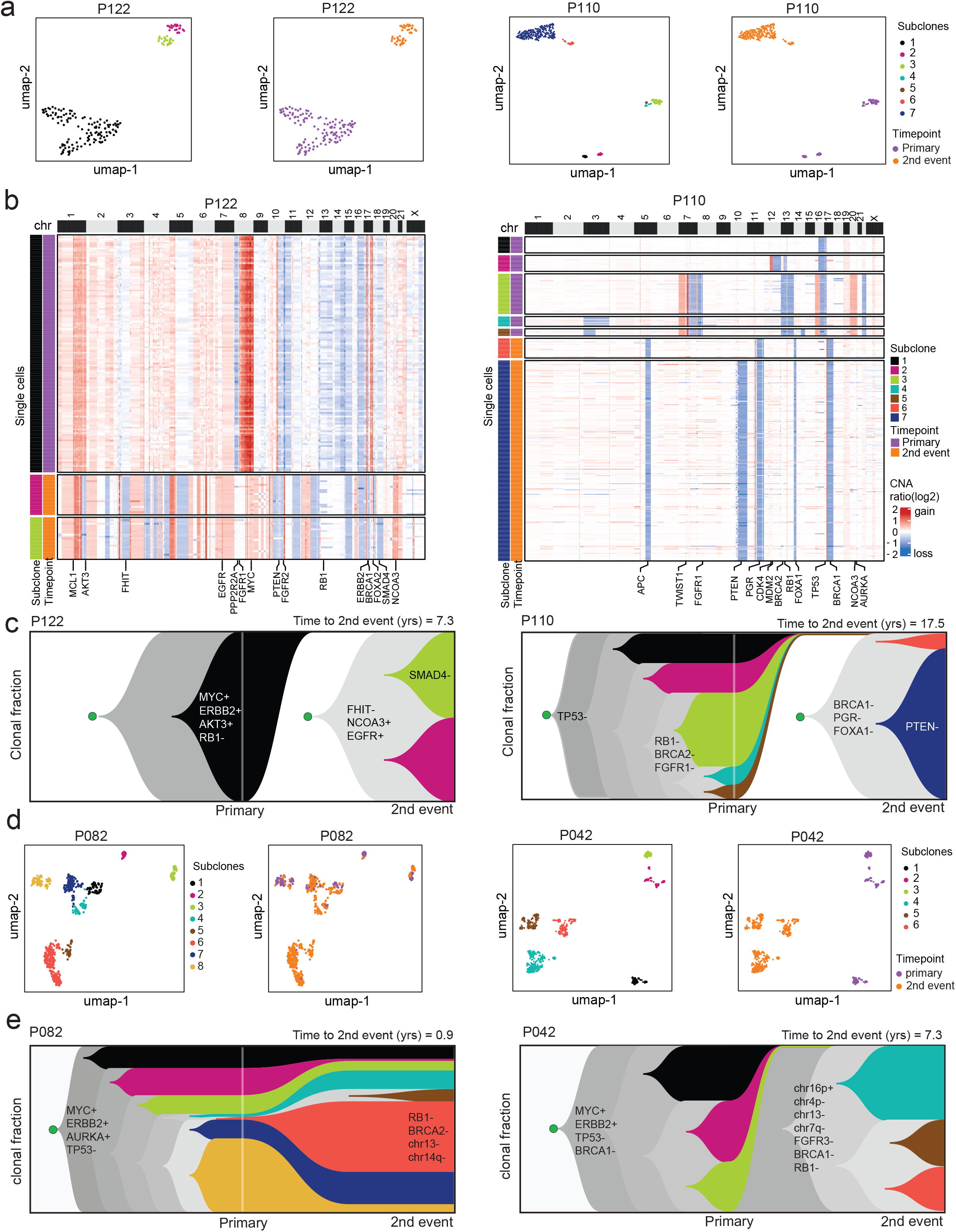
Clonal lineage reconstruction by single cell genome sequencing. **a**, UMAP plots of single cell copy number profiles from FFPE tissue showing clusters of subclones at 2nd event or primary time points for two DCIS patients with independent lineages. **b**, Clustered heatmaps of single cell copy number profiles for two DCIS cases where the 2nd event represents an independent lineage, with selected breast cancer genes annotated below the heatmap. **c**, Mueller plots showing clonal frequencies and lineages reconstructed from neighbour-joining trees using timescape, with selected breast cancer genes annotated, and chromosomal gains and losses indicated by plus and minus signs, respectively. **d**, UMAP plots of single cell copy number profiles from FFPE tissue for two clonally related pairs showing subclones at the primary DCIS and at the 2nd event time points. **e**, Mueller plots of the same two clonally related pairs showing clonal frequencies and lineages reconstructed from neighbour-joining trees using timescape, with selected breast cancer genes annotated, again with gains and losses annotated with plus and minus signs, respectively.

### Clonal analysis of DCIS that recurs as ipsilateral DCIS or as contralateral disease

In total, 34 pairs that recurred as pure DCIS (DCIS->ipDCIS) were analysed, nine by WES and 25 by copy number with or without additional targeted sequencing. 85% (29/34) were found to be related, 9% (3/34) unrelated and 6% (2/34) ambiguous (Figures 3a,5b, Supplementary Figure 5b). DCIS->ipDCIS tended to recur earlier than DCIS->ipINV (mean 36 vs mean 65 months respectively, P=0.0003, t-test) and have a higher rate of clonal relatedness, although not statistically significant (DCIS->ipDCIS 85%, DCIS->ipINV 75%, P=0.2 Fisher’s Exact Test).

As expected, 13 of 14 contralateral invasive events did not show any evidence of relatedness to their primary DCIS. One case, unrelated by copy number, shared a single non-synonymous mutation in HNRNPL (c.G769C) which could represent an early embryonic mutation ^14^.

### Clinical Associations

When considering all ipsilateral events, including DCIS that recurs as DCIS, there was evidence that clonal pairs were more likely to recur in the same quadrant of the original DCIS, and non-clonal pairs in a different quadrant (P=0.004, Fisher’s Exact Test, Supplementary Table 7, Figure 5c). Non-clonal pairs were also more likely to have discordant ER status (P=0.003, Fisher’s Exact Test). In addition, there was a weak but significant association with time to subsequent event with non-clonal pairs having a slightly longer time interval between primary and subsequent event (median time to event 5.0 vs 3.0 years, P=0.04, t-test).

When only pairs that developed a subsequent ipsilateral invasive event were considered non-clonal pairs were still more likely to have discordant ER status (P= 0.003, Fisher’s Exact Test) and occur in a different quadrant of the breast compared to the primary event (P= 0.03, Fisher’s Exact Test) but there was now no association with time to subsequent event. There was also no association with age at diagnosis of primary DCIS or ER/ HER2 receptor status of primary DCIS or grade of primary DCIS in either comparison.

### Reconstructing clonal lineages in DCIS with subsequent invasive disease by single cell genome sequencing

Single cell DNA sequencing (scDNA-seq) was used to profile genomic copy number in 2294 cells from primary and invasive disease from formalin fixed embedded tissue collected from 4 DCIS patients that were classified as clonally unrelated (P122, P110) or clonally related (P082, P042) by the bulk DNA sequencing methods. In the clonally unrelated sample pairs, unbiased clustering identified three major subclones in P122 and seven subclones in P110, in which each of the individual clones was specific to either the primary or subsequent events (Figure 6a). In P122 the clustered heatmaps (Figure 6b) of single cell copy number profiles showed that subclone 1 (c1) was specific to the primary sample and had a number of CNA events including a major amplification of chr8q (*MYC*), loss of chr13 (*RB1*) and gain of a 3.4mb region of chr17q (*ERBB2*), but showed no common CNA events or breakpoints with the recurrent subclones (c2-3), which harbored a large number of CNA events including chromosomal losses of chr3p (*FHIT*), chr10 loss (*PTEN, FGFR2*), chr17q loss (*BRCA1*), loss of chr18 (*SMAD4*) and gains of chr7 (*EGFR*) and chr21 (*NCOA3*). Similarly, in the non-clonal P110 patient, the clustered heatmaps identified common chromosomal losses on 16q and 17p (*TP53*) in all of the subclones (c1-5) from the primary sample but did not share any CNA events or breakpoints with the subclones (c6-c7) that were detected in the invasive disease which had losses on chr5, 11q (*PGR*), chr14p (*FOXA1*), and chr 17q (*BRCA1*). To identify changes in subclonal frequencies during the evolution of the invasive disease, we computed consensus subclone CNA profiles (Supplementary Figure 7c) and reconstructed clonal lineages using neighbour joining trees (Supplementary Figure 7d). The resulting trees were used to reconstruct mueller plots of subclonal frequencies using timescape ^15^ to delineate the order of CNA events that occurred during disease progression in both DCIS tumours (Figure 6c). These data showed that the subclones in the primary samples did not persist or expand in the invasive lesions, suggesting that they represented independent lineages that initiated from different initiating cells in the breast tissue.

We further investigated the clonal substructure of two clonally related patients (P082, P042) that were classified by bulk DNA-seq as having shared mutations between the primary and invasive disease. Unbiased clustering of the single cell copy number profiles identified eight subclones in P082 and six subclones in P042 (Figure 6d). In contrast to the two clonally unrelated pairs, these tumours shared a large number of CNA events between the primary and invasive tissue samples (Supplementary Figure 7a-b). In P082 chromosomal gains in 8q (*MYC*), 17q (*ERBB2*) and 20 (*AURKA*) and losses in 11q (*PGR*), 16q, 17p (*TP53*) were shared among all of the eight subclones, while in P042 chromosomal gains in 1q, 8q (MYC) and 17q (*ERBB2*) and losses in 8p, 11q(*PGR*) 16q, 17p(*TP53*) were present in all six subclones. Furthermore, in P082 multiple subclones (c1, c2, c3, c7, c8) with the same genotypes were detected in both the primary and invasive disease time points. Consensus subclone CNA profiles were computed from the single cell CNA profiles (Supplementary Figure 7b) and used to reconstructed clonal lineages using neighbour joining trees (Supplementary Figure 7d).

Mueller plots of the clonally related tumours identified subclones that expanded in the invasive disease, including clones c4-c7 in P082 and clones c4-c6 in P042 (Figure 6e). In P082 the expanded invasive clones harboured CNA events including losses of chr13 (*BRCA2, RB1*) and 14q (Figure 6e, Supplementary Figure 7a-b). In P042 the expanded invasive clones (c4-c6) harboured losses of chromosomes 2, 4p and 13 (*BRCA2, RB1*) and gains of chr10 (*GATA3*), 11p (*MDM2*) and 12 and 16p (Figure 6e, Supplementary Figure 6a). Collectively, this single cell DNA data validated the clonal classifications (related, unrelated) estimated from the bulk DNA-seq analysis and further resolved direct and independent clonal lineages, revealing chromosomal events and genes associated with subsequent events in these four cases.

## Discussion

Our results show that 18% of subsequent ipsilateral invasive breast cancers that occur after DCIS appear to be clonally unrelated to their primary DCIS and thus are likely to be new primary cancers. Our data are supported by one small study that also used genomic data to assess clonal relatedness and showed that in two of eight pairs of primary DCIS and subsequent invasive disease copy number aberrations were not concordant ^16^. In our study non-clonal ipsilateral invasive disease were more likely to occur in a different quadrant of the breast compared to the primary DCIS but interestingly there was no clear association with time when only subsequent invasive events were considered. Including subsequent DCIS events in the analysis resulted in a weak association with time due to the fact that subsequent pure DCIS events were more likely to occur within 5 years compared to subsequent invasive events and were often clonal. In fact, of the 12 invasive breast cancers that developed >10 years after their primary DCIS, eight still showed evidence of clonal relatedness to the preceding DCIS. With 75% of invasive subsequent events appearing clonally related by at least one method of analysis, our findings are consistent with the model that DCIS is a precursor for invasive breast disease, including in cases where the initial DCIS had been treated by mastectomy.

The “sick lobe” theory and field cancerization concept may explain why, despite wide local excision of DCIS with histologically clear margins and radiotherapy, we still see clonal recurrences ^17,18^. Both concepts imply that the epithelial lining within a lobe can consist of cells which have undergone early genetic events (first “hit”), either during mammary development (sick lobe theory) or at unspecified time points (field cancerization concept) but have not acquired all the changes (second “hit”) necessary for tumour invasion. Better understanding of mammary field cancerization could improve informed decision-making in patient risk stratification and treatment. Ultimately, this may reveal markers that could be applied to identify and create a “map” of the affected lobe(s) within the breast and could be used to guide surgical planning ^19^. Yet, one must bear in mind that choosing optimal resection margins needs to be balanced against the morbidities associated with more radical surgery and the risk of recurrence from a cancerized field left *in situ* ^18^. Moreover, these findings support a greater role for systemic treatments, such as endocrine therapy, particularly when less aggressive surgery is considered.

There is no consensus about which type of data and statistical method is most suitable to distinguish clonal recurrences from independent primary tumours. Bierman and colleagues have shown that the type of molecular data analyzed had a stronger impact on clonality determination than the analytical methods used ^20^. Our data show, as does the data of Bierman *et al*, that assessment of clonal relatedness by different types of molecular data is not always concordant. CNAs which are acquired at early stages of tumorigenesis are thought to be the most stable type of biological data for clonality assessment, in comparison to mutations which evolve gradually over long periods of time, generating extensive clonal diversity ^21,22^. However, they are also more common so could be false positive indicators of clonality (such as chr1q gain, 8q gain and 17p loss) and for genomically stable tumours mutation data is likely to be more informative. False negative results can be attributed to clonal heterogeneity obfuscating the copy number profile in the case of copy number data, or shared variants found outside the scope of the selected gene panel in the case of targeted sequencing, with high depth whole exome or genome sequencing providing more definitive evidence at the mutation level. The pattern found in synchronous DCIS&INV cases, by which the 7% of cases that do not show clonal relatedness also harbour the least number of mutations, is compatible with these technical limitations. Importantly, we did not observe this pattern in the invasive events following DCIS. To that end single cell analysis from both tumours of a pair provides the most robust assessment of shared events. Our single cell data support the findings of our bulk analyses and we see clear evidence that not all ipsilateral invasive events are clonally related to their primary DCIS.

The finding that 18% of invasive events following DCIS are not clonally related indicates that: 1) true recurrence rates after DCIS are likely to be overestimated; and 2) DCIS might not just be a precursor, but could also be a risk lesion for development of further invasive disease, as has been postulated for lobular carcinoma *in situ* (LCIS) ^23^. This may also explain why it has been difficult to develop robust biomarkers for risk stratification of DCIS, as two different risk issues have to be addressed ^24^: first, the risk of the DCIS lesion progressing to invasive disease; second, the risk of developing an unrelated, most likely independent new invasive breast cancer. For the latter, it will be important to assess how germline breast cancer predisposition variants influence the risk of new ipsilateral and contralateral new primaries as known invasive breast cancer risk polymorphisms and rare variants have been shown to also have a strong association with DCIS, particularly ER+ DCIS which has previously been shown to be a highly significant predictor of contralateral breast events ^25-27^.

Our findings are consistent with the use of endocrine therapy in ER+ DCIS, as endocrine therapy has been shown to reduce the risk of both ipsilateral and contralateral events after wide local excision of DCIS and is effective at reducing invasive breast cancer in high-risk women ^26,28,29^. However endocrine therapy comes with potential short and long-term adverse effects. The data showing that lower doses of tamoxifen with less toxicity are likely to be effective at reducing events after DCIS as well as in women with a diagnosis of ADH may make endocrine therapy a more acceptable option for risk reduction and an alternative to radiotherapy ^30^.

In conclusion, comprehensive genomic and molecular data provide evidence that 18% of invasive subsequent invasive events are unrelated to the preceding DCIS lesion, and appear to arise as a genomically distinct entity. These results challenge the current dogma that almost all subsequent ipsilateral invasive breast cancers are due to progression of the initial DCIS. Our findings support a paradigm shift that confirms a more complex role for DCIS than previous recognized, and that the future management of DCIS should take into account both the precursor and risk factor implications of this diagnosis.

## Methods

### Samples

Cases of pure primary DCIS that, after treatment, had subsequently developed recurrent disease were identified from:

1. the Sloane project, a national audit of women with non-invasive neoplasia within the United Kingdom National Health Service Breast Screening Programme (REF 08/S0703/147), median follow up 5.3 years ^31^.
2. the Dutch DCIS cohort study, this is a nation-wide, population-based patient cohort derived from the Netherlands Cancer Registry (NCR), in which all women diagnosed with primary DICS between 1989 and 2004 were included, and has a median follow up time of 12 years ^32^. This cohort was linked to the nationwide network and registry of histology and cytopathology in the Netherlands (PALGA). The study was approved by the review boards of the NCR (ref. no. 12.281) and PALGA (ref.no. LZV990). This study was approved by the institutional review board of the Netherlands Cancer Institute under number CFMPB166, CFMPB393 and CFMPB688.
3. the Duke Hospital cohort, a hospital-based study of women (age 40-75 years) diagnosed with DCIS between 1998 and 2016, with a median follow up of 7.9 years (IRB approvals: Pro00054877, Pro00068646).

In addition, 34 synchronous DCIS-IBC lesions were also included for comparison with published literature, identified from the Duke DCIS cohort. Details of the cohorts can be found in Supplementary Table 1.

Formalin-fixed paraffin-embedded (FFPE) tissue specimens of patient-matched DCIS and subsequent recurrence were retrieved and reviewed by specialist breast pathologists to confirm the diagnosis and exclude confounding features (such as microinvasion).

In total, 143 DCIS-recurrence pairs were included in this study, 95 had developed an ipsilateral invasive recurrence, 34 had an ipsilateral DCIS recurrence and 14 a recurrence in the contralateral breast.

### DNA isolation

For DNA isolation, either macrodissection using a light microscope or laser microdissection (LMD) was performed. 8µm sections were stained using nuclear fast red (macrodissection) or toluidine blue (LMD) and DCIS or invasive disease were separated from the normal tissue. Tumour DNA was extracted using the AllPrep DNA/RNA FFPE Kit (Qiagen).

### Exome Sequencing

Whole exome sequening (WES) of the paired DCIS with subsequent recurrence together with matched normal tissue was performed at the Department of Genomic Medicine, MD Anderson Cancer Center. Genomic DNA (18-300 ng) was used to generate sequencing libraries using the SureSelect^XT^ Low Input library kit. Libraries were sequenced on NovaSeq 6000 multiplexing 16 tumor samples per lane.

WES of the Duke Hospital Cohort of 34 synchronous paired DCIS and invasive disease and matched normal tissue was performed at the McDonnell Genome Institute at Washington University School of Medicine. Genomic DNA (30-150 ng) was sheared, to a mean fragment length of 250 bp and Illumina sequencing libraries were generated as dual-indexed, with unique bar-code identifiers, using the Swift Biosciences library kit. Libraries were sequenced on Illumina HiSeq 2500 1T instrument by multiplexing nine tumor samples per lane.

Data were converted to a fastq format and then aligned to the hg19 reference genome using the Burroughs-Wheeler Aligner (BWA). The aligned BAM files were subjected to mark duplication, re-alignment, and re-calibration using Picard v2.21.9 and GATK v4.1.7.0. The BAM files were then analyzed by MuTect and Pindel against the matched normal sample to detect somatic single nucleotide variants (SNV) and insertions/deletions (indels), respectively.

Individuals with normal sample median target coverage (MTC) > 40x and tumor sample MTC > 80x were included for further investigation. Variants were filtered by the following criteria: (1) Log odds score >= 10. (2) exonic variants. (3) Tumor sample coverage at this site >= 15. (4) Normal sample coverage at this site >= 10. (5) Allele fraction in tumor sample >= 0.02. (6) Allele fraction in normal sample < 0.01. (7) Population frequency < 0.01 in ExAC, ESP6500, and 1000g database. (8) Hotspot mutations in *PIK3Ca* and *TP53* were added back to the dataset, if they did not pass these criteria. (9) For all independent samples, mutations present were manually checked in IGV.

We identified potential sample mismatches using an in house script for computing SNP matching index. For any pair of Platypus vcfs (two samples), we removed the SNPs from random chromosomes as well as SNPs with coverage < 10, and calculated the number (nAB) of overlapping SNP’s (by position), and the number (nGAB) of the same alleles within the overlapping SNPs. The score (match-index %) = nGAB*100/nAB. Using this index, we removed all mismatches with score <90%.”

### Copy number analysis

Somatic copy number aberrations were ascertained using the HumanCytoSNP-12 BeadChip Kit (Illumina) in cases with 100-250ng of DNA available from the Sloane Project. DNA was restored with the Infinium HD FFPE DNA Restore Kit (Illumina). Raw SNP array data was processed with GenomeStudio 2.0 software (Illumina) and subsequently with the ASCAT (Allele-Specific Copy Number Analysis of Tumours) software algorithm (implemented in R), to estimate allele-specific copy number, the aberrant cell fraction and tumour ploidy ^33^. Copy number profiles with number of segments higher than 500 and a LRR noise higher than 0.16 were removed from the analysis. Whole genome duplicated (WGD) events were determined by the mode of the major allele in a sample (Supplementary Figure 2b). Copy number profiles of samples that underwent WGD events were corrected. PLINK v1.07 was used to estimate the pairwise relatedness using the raw SNP genotyping data in order to exclude sample mismatches between primary DCIS and subsequent event.

In cases with limited DNA copy number was ascertained using low pass whole genome analysis. The UK cohort used NEBNext Ultra II DNA Library Prep Kit for Illumina as per manufacturer’s instructions and the Dutch cohort used the KAPA hyper prep kit (KAPA Biosystems), protocol KR0961-v5.16. Agilent S5XT-2 (1-96) adapters with Illumina P5 and P7 sequences were used, containing 8bp Agilent indices.

Libraries were pooled and sequenced single-end on HiSeq 2500 sequencer (Illumina). After demultiplexing, FASTQ files were aligned to the human reference genome GRCh38 (hg38) using BWA 0.7.17 aligner and converted to BAM files with Samtools 1.9. Duplicate reads were marked with Picard 2.18.3 and removed together with reads with mapping qualities lower than 37 using Samtools 1.9. Relative copy number profiles were obtained with QDNAseq 1.22.0 after setting a 100 kb fixed bin size. We filtered out copy number profiles with a number of segments higher than 400 and observed/expected noise ratio higher than

50. In addition, we filtered out profiles that did not show any copy number aberrations and removed noisy patterns by visual inspection. CGHcall 2.48.0 was used for relative copy number calling. For detecting differential copy number variation between groups, absolute copy number calls after tumour cell fraction adjustment obtained with ACE 1.4.0 and Fisher’s Exact Test were used.

The copy number profiles for all pairs can be found in Supplementary File 1.

### Targeted sequencing

For the UK cohort, sequencing of all exons of a custom 121 breast cancer-associated gene panel (Supplementary Table 8) was performed using the SureSelect XT low input Target Enrichment System (Agilent Technologies, Santa Clara, CA, USA). 100bp read paired end sequencing was performed on the HiSeq2500 platform. The sequencing output was aligned to the reference genome hg19 using the Burrows-Wheeler Aligner (BWA-MEM). Variants were called using MuTect2 from the Genome Analysis Toolkit (version 4.1.0.0), using the matched normal tissue to exclude germline variants. Variants with an allele frequency <5%, coverage <30x were excluded. Sequencing reads of tumour and normal pairs were visualized on the Integrative Genomics Viewer (IGV) to exclude germline variants and also potential sequencing artefacts.

The Dutch cohort was sequenced using an IonTorrent AmpliSeq custom 53-gene panel (Supplementary Table 9) and were processed according to the Ion AmpliSeq Library Kit Plus protocol (ThermoFisher Scientific). Reads were aligned to the reference genome GRCh37 (hg19) using the Torrent Mapping Alignment Program, and variant calling was performed using Torrent Variant Caller (TVC) version 5.6. Variant data in VCF format was first translated to GRCh38 and annotated using bedtools, picard (https://broadinstitute.github.io/picard/command-line-overview.html), samtools, bcftools and VEP, and further analyzed in R, employing vcfR, and tidy verse. True somatic variants, identified via filtering during which low quality variants (variant allele frequency, VA) <10%, coverage <100x, and a quality (QUAL) of <1000), artifacts (found in >90% of samples), and germline variants (>5 cases in GNOMAD and GoNL) were removed. Details regarding the amplicon panel design, performance, and filtering QC are provided in Supplementary File 2.

### Single Cell Sequencing

FFPE samples were deparaffinized using the FFPE Tissue Dissociation Kit from MACS (Cat#130-118-052). Nuclear suspensions were prepared from the recovered cell suspensions using a DAPI-NST lysis buffer (800 mL of NST (146 mM NaCl, 10 mM Tris base at pH 7.8, 1 mM CaCl2, 21 mM MgCl2, 0.05% BSA, 0.2% Nonidet P-40)), 200 mL of 106 mM MgCl2, 10 mg of DAPI)1,2. The nuclear suspensions were filtered through a 35 mm mesh and single nuclei were flow sorted (BD FACSMelody) into individual wells of 384-well plates from the aneuploid peak. After sorting single nuclei, direct tagmentation chemistry was performed following the Acoustic Cell Tagmentation (ACT) Protocol (Minussi et al. 2020, under revision). Briefly, nuclei were lysed and tagmentation was performed using TN5 to add dual barcode adapters to the DNA, followed by 12 cycles of PCR. The resulting libraries were QCed for concentration >10ng/ul and pooled for sequencing on the HiSeq4000 (Illumina) instrument at 76 cycles.

To calculate single-cell copy number profiles we demultiplexed sequencing data from each cell into FASTQ files, allowing 1 mismatch of the 8 bp barcode. FASTQ files were aligned to hg19 (NCBI Build 37) using bowtie2 (2.1.0) ^34^ and converted from SAM to BAM files with SAMtools (0.1.16) ^35^. PCR duplicates were removed based on start and end positions. Copy number profiles were calculated at 220kb resolution using the variable binning method ^36^. Single cells with <10 median reads/bin were excluded for downstream copy number analysis. GC normalized read counts were binned into bins of variable size, averaging 200kb, followed by population segmentation with the multipcf ^37^ (gamma = 10) method from the R Bioconductor multipcf package. The log2 copy number ratio were calculated and used for subsequent analysis. We filtered out noisy single cells with mean 9-nearest neighbor correlation less than 0.85. The mean 9-nearest neighbor correlation is calculated as the average of the Pearson correlation coefficients between any single cell and its 9-nearest neighbors. This step removed single cells with poor whole-genome amplification from the subsequent data analysis. Single-cell ratio data was embedded into two dimensions using UMAP ^38^, R package ‘uwot’ (v0.1.8, seed = 31, min dist = 0.2, n_neighbors = 30, distance = “manhattan”). The resulting embedding was used to create an SNN graph with R Bioconductor package scran (v1.14.6) ^39^. Subclones were identified with R package ‘dbscan’ (v1.1-5, k_minor = 0.02*#cells) ^40^. Heatmaps were plotted with R package ComplexHeatmap (v2.2.0) ^41^.

### Clonal relatedness calculation using Breakclone

Breakclone is an in-house package to assess clonal relatedness. Unlike other packages ^11 12^, it incorporates both population frequency and allele frequency when using mutation data for determining clonal relatedness. When using copy number data, it uses the position of the individual copy number aberration breakpoints rather than aberration events at the chromosome arm level to determine clonal relatedness correcting for the frequency of the event within the cohort. These are harder to compare across cohorts analysed with different techniques but, we believe, provide much stronger evidence of clonal relatedness when shared between lesions ^20^. A reference distribution of concordance scores is calculated by randomly permuting all possible pairs from different patients, the number of permutations empirically determined as necessary for the distribution to converge, and is used to calculate p-values for the concordance score of each tumour pair. The threshold for determining clonal relatedness is set as p < 0.01. Clonality scores between 0.05 and 0.01 were called ambiguous. All values above 0.05 were considered as non-clonal. We considered a sample pair as clonally related if at least one of the different methods (WES, copy number, panelseq) gave a clonal score.

### Copy number data

Each breakpoint shared between two tumours is interpreted as evidence of their relatedness, while each breakpoint unique to one tumour is interpreted as evidence of independence. However, given the generally stochastic process of genomic instability, a common aberration provides stronger evidence than an independent once – therefore, the effect of the unique aberrations in the score calculations is weighted down by ½. The concordance score range starts at 0 for samples that share no aberrations and approaches the theoretical limit of 1 as the samples become more similar – the score for any two identical samples will be slightly below 1 due to the population frequency corrections.

Each somatic copy number aberration (SCNA) breakpoint was compared between the pairs of tumours from the same individual. Concordant breakpoints were defined as the same type of aberration, present in the same location ± 5 * *averageinterprobelength* in order to account for technical variation. This figure was empirically determined as the number that captured the most likely concordant breakpoints without compromising their uniqueness – larger values led to the same breakpoints being included in calculations twice. Each concordant breakpoint was adjusted for its frequency in the entire cohort (*f*_*b*_), producing an adjusted breakpoint concordance score (*s*_*b*_) based on the equation:

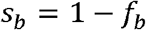

For those samples that carried whole genome duplications a parsimonious approach was used to infer the most likely ancestral copy number state (Supplementary Table 2), which was then used in subsequent calculations.

The final sample concordance score (*s*) was then calculated between the pairs using all of the SCNAs in the samples and taking into account the total number of breakpoints in both samples (*n*_*b*_), using the following formula:

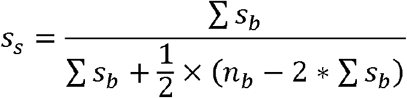

A reference distribution of concordance scores was calculated using all possible tumour pairs from different patients and was used to calculate p-values for the concordance score of each tumour pair.

### Somatic mutation data

The allele frequency is weighted according to the population frequency – the lower the population frequency, the higher the weight of the allele frequency. In the calculations the square root of the population frequency is used to normalise the range of possible values. The range of values for this score is between 0 for samples with no shared mutations and 1 for samples with identical mutation profiles.

Mutation data from each sample was compared and common variants were assigned a score, based on both their allele frequency in each sample (*A*_*1*_ and *A*_*2*_), and their frequency in the population (*P*_*c*_). A higher allele frequency is interpreted as a stronger indicator of clonal relatedness, while a higher population frequency is interpreted as diminishing the predictive value of the variant. The TCGA Pan-Cancer Atlas breast cancer mutation calls were used for this adjustment, in addition to the mutations found in our cohort. The concordance score (*s*_*s*_) was subsequently calculated, taking into account the private variants in both tumour samples and their allele (*A*_*p*_) and population (*P*_*p*_) frequencies, using the following formula:

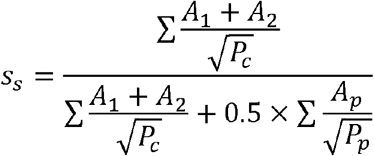

A reference distribution of concordance scores was calculated using all possible tumour pairs from different patients and was used to calculate p-values for the concordance score of each tumour pair.

An implementation of the method is available as an R package at github.com/argymeg/breakclone.

### Comparison of the different clonality algorithms

In order to assess the added usefulness of our method, we applied the relevant copy number and mutation-based functions in the previously published Clonality package to our samples where copy number was ascertained by SNP array (see figure below). We observed with the estimate of the number of clonally related pairs was lower for the clonality package compared to our proposed method, as well as suggesting a number of the contralateral recurrences were clonally related. Visual inspection of the contralateral samples that were called clonal showed that they were generally genomically stable samples, with few major aberrations. When those samples do share aberrations on the same chromosomal arms, they are considered clonally related by the Clonality package which, by design, relies on fewer events, but not by our method which relies on the presence of multiple copy number events.

**Clonal relatedness by Breakclone in SNP array data**

**Clonal relatedness by Clonality Package in SNP array data**

## Data Availability

All data presented in this article will be made available upon publication and are available upon request.

## Acknowledgements

We thank all patients in the US, The Netherlands and the UK who have donated their data and tissue for this work. We also wish to thank all the collaborating hospitals, and in particular, pathology departments, and all persons who have helped in the process of data collection and analysis.

We would like to specifically acknowledge Otto Visser, Annemarie Eeltink, and the registration teams of the Netherlands Comprehensive Cancer Organization for collection of the data for the Netherlands Cancer Registry. We thank Lucy Overbeek and PALGA, the nation-wide histopathology and cytopathology data network and archive, for providing pathology data and for their help in the collection of the residual patient material. We acknowledge the staff of the NKI-AVL Core Facility Molecular Pathology & Biobanking for their technical support and the staff of the NKI-AVL Genomics Core Facility and Ronald van Marion of the Erasmus University Medical Center for their sequencing support.

We thank Thomas Hardiman and Radhika Kataria for their input during development of the Breakclone method. The data for the Sloane Project is based on information collected and quality assured by the PHE Population Screening Programmes. Access to the data was facilitated by the PHE Office for Data Release.

We are also grateful for Thomas Lynch who helped to expertly coordinate the PRECISON team at Duke University.

## Author contributions

EHL, LFAW, ESH, NNN, AF, AT, JW, EJS: Study design, supervision, data interpretation, preparation of the manuscript

TK: data analysis, data interpretation, preparation of the manuscript

AM: Method development, data analysis, data interpretation, preparation of the manuscript

LLV: Study design, data collection, data analysis, data interpretation, preparation of the manuscript

MSche, AF, VS, MH, DM, MRE, AAA, LK, CSS,: Data collection, data analysis, data interpretation, preparation of the manuscript

ES: technical support single cell sequencing

MX: technical support WES

WM, MK, PK, MM, LM, FN, RS, TH, JZ: technical support

KC, LF, BM, AS : Data collection

SN, JQ, MSr, AG: Bioinformatics support

CM, SP: Study design, supervision

HRD, JM, SNZ, CM,: Supervision, data interpretation

MScha, MKS: Study design, data interpretation

HS, DC: manuscript preparation

All authors reviewed the manuscript

