## Supplementary figures for "Genomic profiling defines variable clonal relatedness between invasive breast cancer and primary ductal carcinoma *in situ*"

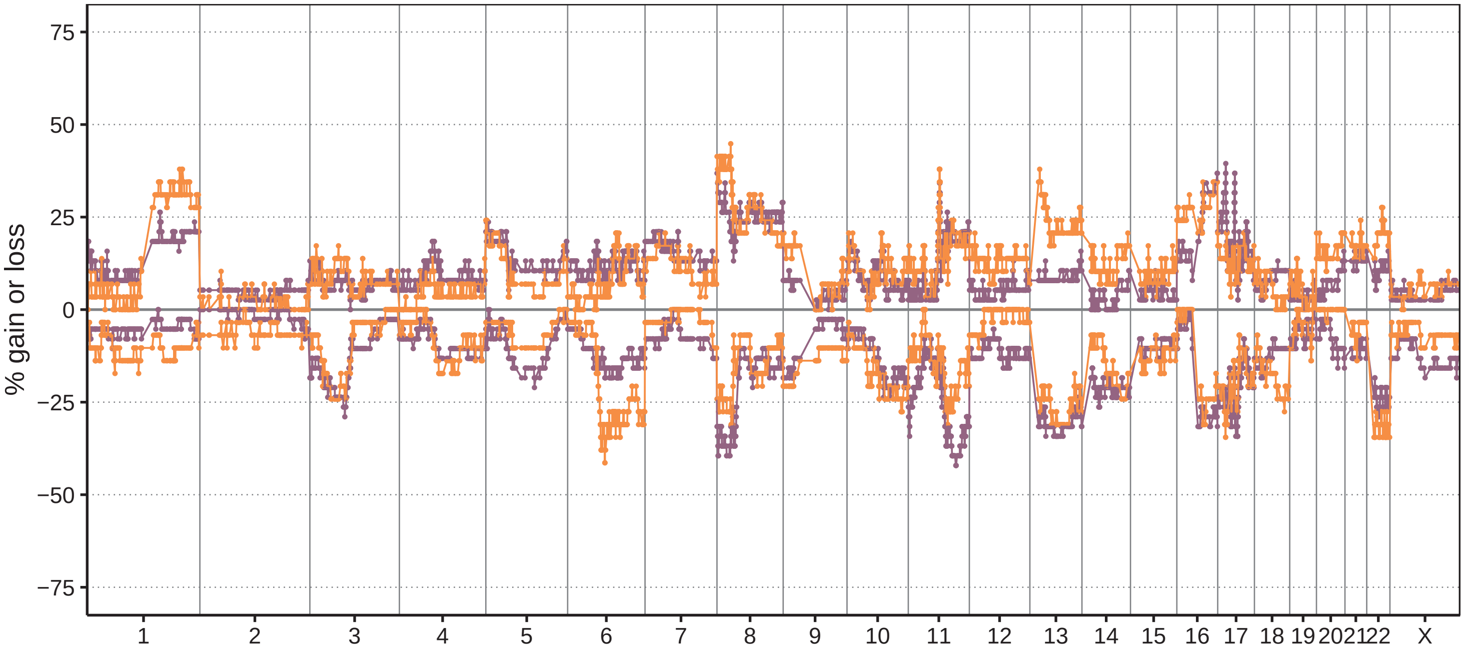


Figure S1. **Copy number alterations in primary DCIS and subsequent invasive disease by SNP-array.** Frequency plot of genome wide copy number assessed by SNP array in primary DCIS (purple, $n = 38$) and subsequent invasive disease (orange, $n = 29$). The $y$-axis shows percentage of samples with gains (above zero line) and losses (below zero line). The genomic position is indicated by chromosome 1 on the left and up to chromosome X on the right with chromosome boundaries indicated by vertical lines.

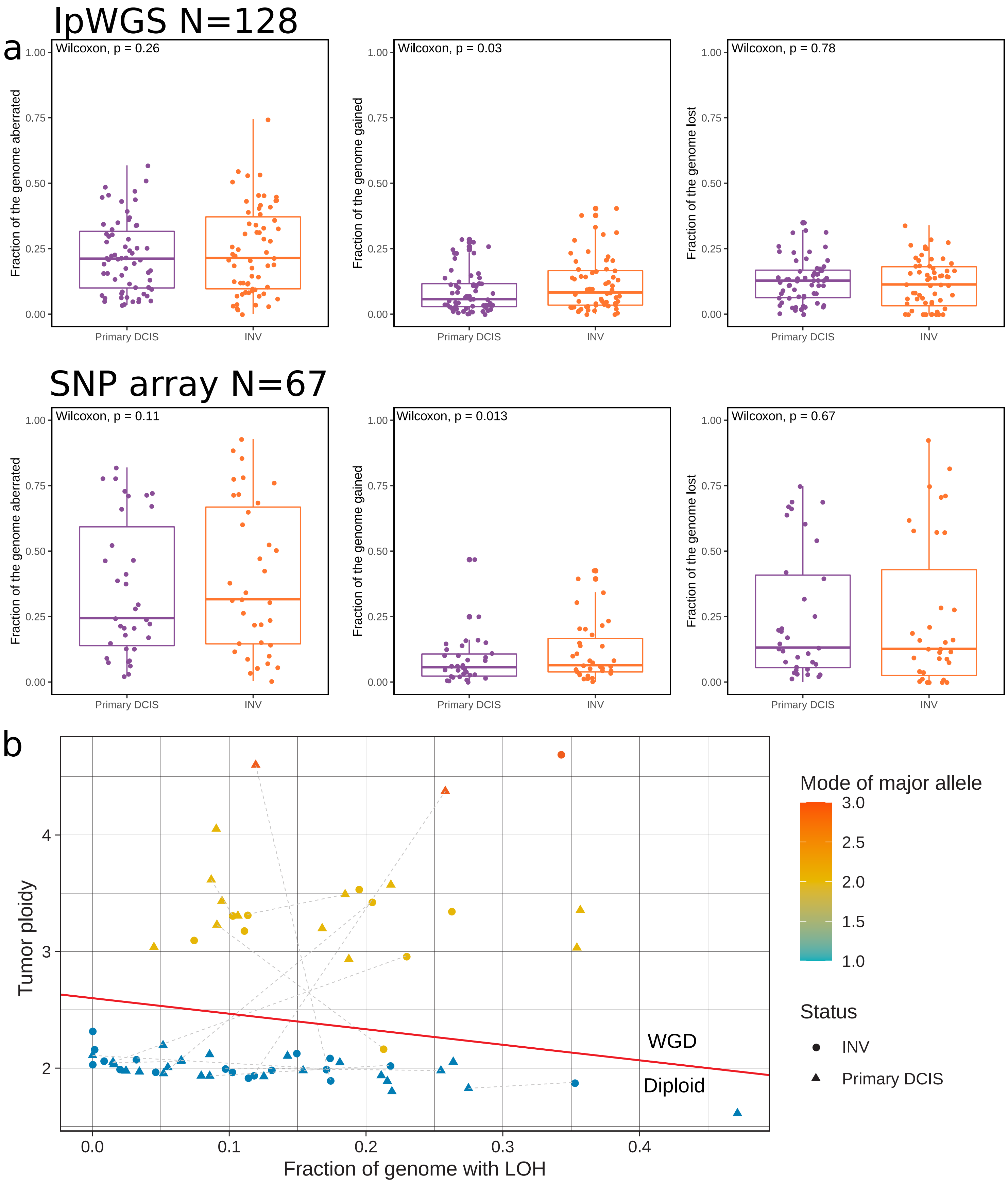
lpWGS N=128
a
Wilcoxon, p = 0.26
Fraction of the genome aberrated
Wilcoxon, p = 0.03
Fraction of the genome gained
Wilcoxon, p = 0.78
Fraction of the genome lost
Primary DCIS
INV
SNP array N=67
Wilcoxon, p = 0.11
Fraction of the genome aberrated
Wilcoxon, p = 0.013
Fraction of the genome gained
Wilcoxon, p = 0.67
Fraction of the genome lost
0.00
0.25
0.50
0.75
1.00
b
Tumor ploidy
Fraction of genome with LOH
0.0
0.1
0.2
0.3
0.4
2
3
4
WGD
Diploid
Mode of major allele
3.0
2.5
2.0
1.5
1.0
Status
INV
Primary DCIS

Figure S2. **Copy number alterations analysis details in lpWGS and SNP-array.** **a**, Fraction of genome aberrated (FGA), gained (FGG) and lost (FGL) in primary DCIS and subsequent invasive disease by lpWGS (top panel, $n = 128$) and SNP-array (bottom panel, $n = 67$). The boxplots present the distribution of FGA, FGG and FGL for all samples in the primary DCIS (purple) and the subsequent invasive disease (orange). Wilcoxon signed-rank test $p$-values are shown. **b**, Whole genome duplication (WGD) events in primary DCIS (triangles) and subsequent invasive disease (dots) by SNP-array samples ($n = 67$) determined by the mode of the major allele in a sample. Dotted grey lines link clonally related primary DCIS samples and its paired subsequent invasive disease. WGD samples found above the red line (*i.e.*, samples with a ploidy higher than $2.4 - \frac{4}{3}FGLOH$ meaning $FGLOH$ fraction of genome with loss of heterozygosity) have a mode of major allele greater than 2, which matches inference from the spread of the samples in the proportion of LOH versus ploidy space.

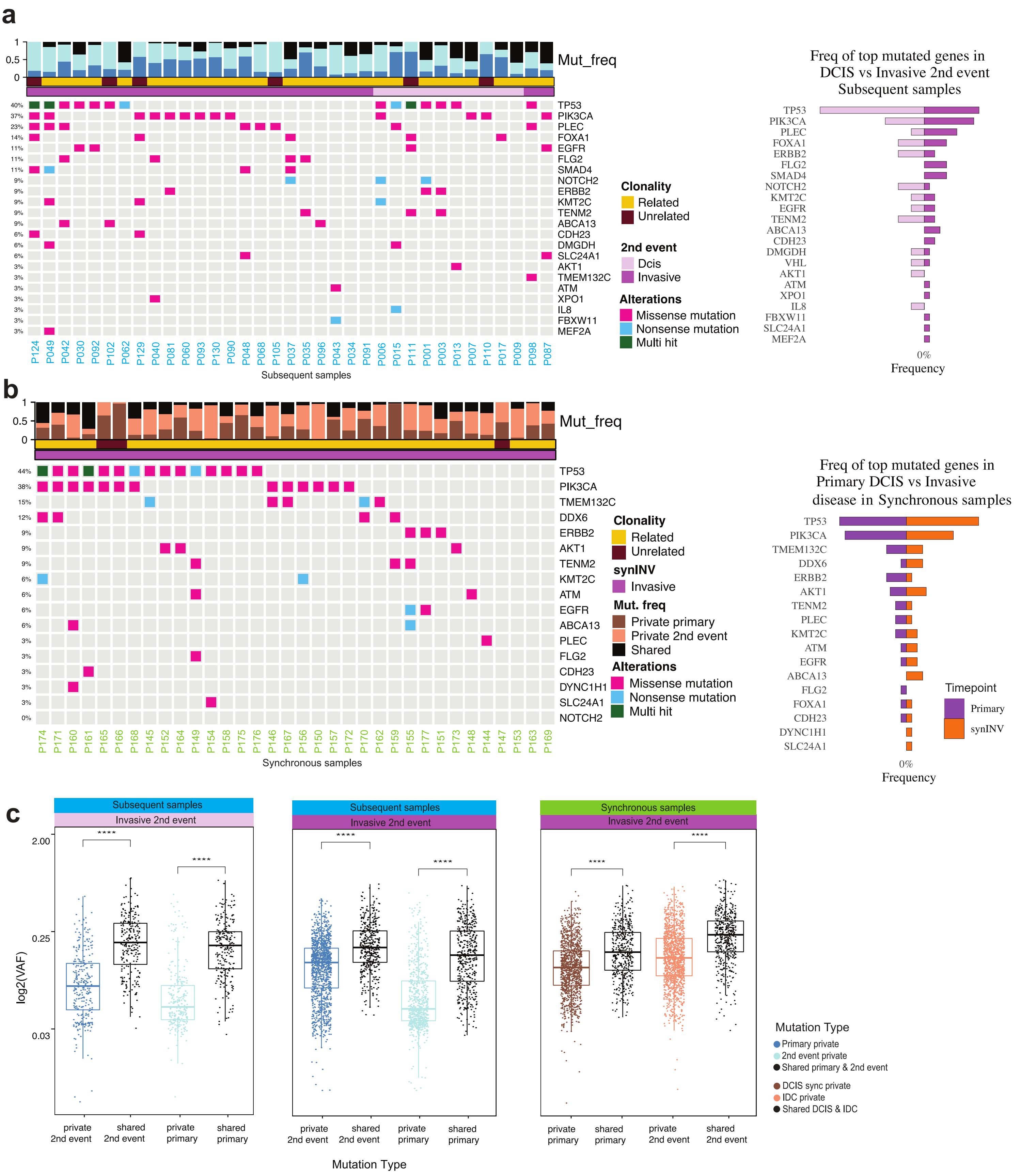
a
1
0.5
0
Mut_freq
40%
37%
23%
14%
11%
11%
11%
9%
9%
9%
9%
9%
6%
6%
6%
3%
3%
3%
3%
3%
3%
3%
TP53
PIK3CA
PLEC
FOXA1
EGFR
FLG2
SMAD4
NOTCH2
ERBB2
KMT2C
TENM2
ABCA13
CDH23
DMGDH
SLC24A1
AKT1
TMEM132C
ATM
XPO1
IL8
FBXW11
MEF2A
P124 P049 P042 P030 P092 P102 P062 P129 P040 P081 P060 P093 P130 P090 P048 P068 P105 P037 P035 P096 P043 P034 P091 P006 P015 P111 P001 P003 P013 P007 P110 P017 P009 P098 P087
Subsequent samples
Clonality
Related
Unrelated
2nd event
Dcis
Invasive
Alterations
Missense mutation
Nonsense mutation
Multi hit
Freq of top mutated genes in DCIS vs Invasive 2nd event Subsequent samples
TP53
PIK3CA
PLEC
FOXA1
ERBB2
FLG2
SMAD4
NOTCH2
KMT2C
EGFR
TENM2
ABCA13
CDH23
DMGDH
VHL
AKT1
ATM
XPO1
IL8
FBXW11
SLC24A1
MEF2A
0%
Frequency
b
1
0.5
0
Mut_freq
44%
38%
15%
12%
9%
9%
9%
6%
6%
6%
6%
3%
3%
3%
3%
3%
0%
TP53
PIK3CA
TMEM132C
DDX6
ERBB2
AKT1
TENM2
KMT2C
ATM
EGFR
ABCA13
PLEC
FLG2
CDH23
DYNC1H1
SLC24A1
NOTCH2
P174 P171 P160 P161 P165 P166 P168 P145 P152 P164 P149 P154 P158 P175 P176 P146 P167 P156 P150 P157 P172 P170 P162 P159 P155 P177 P151 P173 P148 P144 P147 P153 P163 P169
Synchronous samples
Clonality
Related
Unrelated
synINV
Invasive
Mut. freq
Private primary
Private 2nd event
Shared
Alterations
Missense mutation
Nonsense mutation
Multi hit
Freq of top mutated genes in Primary DCIS vs Invasive disease in Synchronous samples
TP53
PIK3CA
TMEM132C
DDX6
ERBB2
AKT1
TENM2
PLEC
KMT2C
ATM
EGFR
ABCA13
FLG2
FOXA1
CDH23
DYNC1H1
SLC24A1
0%
Frequency
Timepoint
Primary
synINV
c
Subsequent samples
Invasive 2nd event
Subsequent samples
Invasive 2nd event
Synchronous samples
Invasive 2nd event
****
****
****
****
****
****
2.00
0.25
0.03
log2(VAF)
private 2nd event
shared 2nd event
private primary
shared primary
private 2nd event
shared 2nd event
private primary
shared primary
private primary
shared primary
private 2nd event
shared 2nd event
Mutation Type
Mutation Type
Primary private
2nd event private
Shared primary & 2nd event
DCIS sync private
IDC private
Shared DCIS & IDC

Figure S3. **Mutational landscape of DCIS samples.** **a**, Distribution of mutations in subsequent samples with DCIS and invasive 2nd event. Mirror plot shows the frequency of the top mutated genes comparing DCIS and invasive 2nd event. **b**, Distribution of mutations in synchronous DCIS and invasive disease. Mirror plot shows the frequency of the top mutated genes comparing DCIS and synchronous invasive disease. **c**, Boxplots comparing variant allele frequency in subsequent samples between different occurrence types left to right, In subsequent and synchronous samples with DCIS and invasive 2nd event, shared mutations have significantly higher VAF's than private mutations.

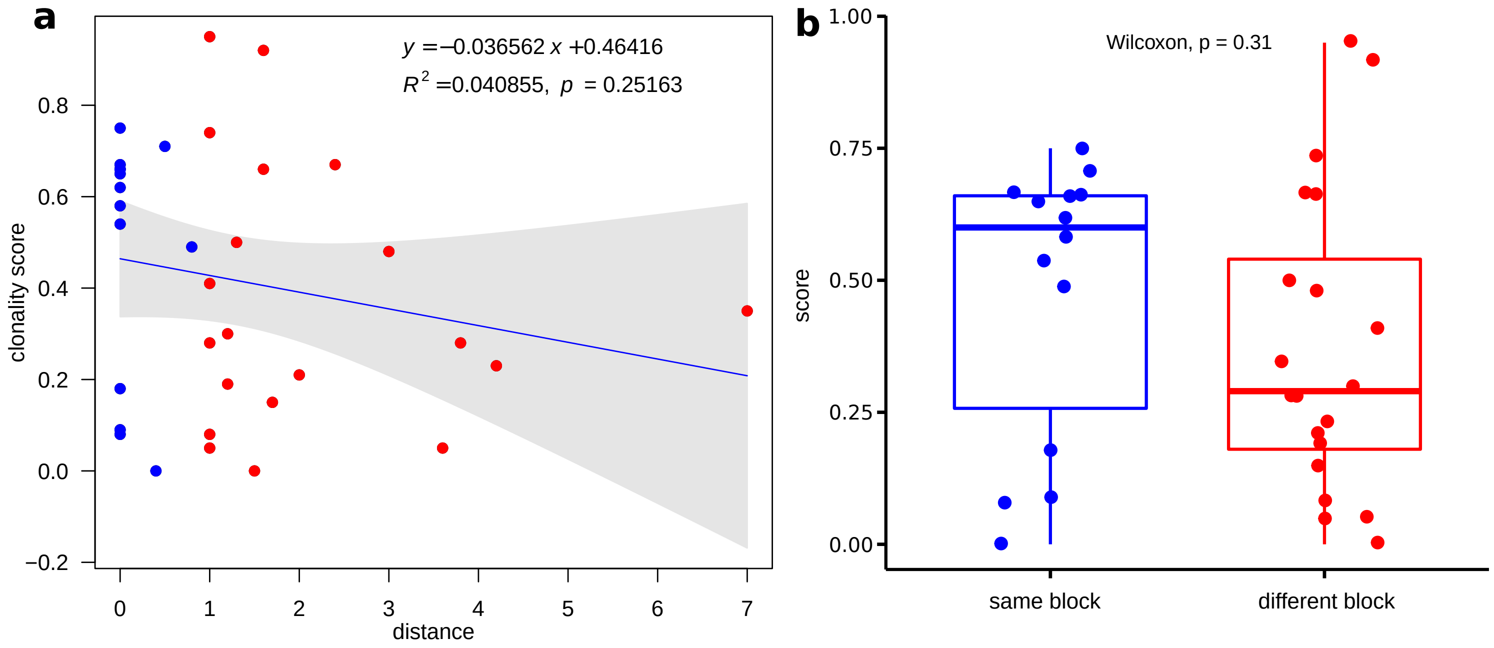


Figure S4. **Distance between DCIS and synchronous invasive disease.** **a**, Clonality score plotted against the distance between the DCIS tissue cells and synchronous invasdive tissue in cm for $n = 34$ synchronous DCIS-INV lesions. **b**, Clonality score plotted for the $n = 34$ synchronous DCIS-INV lesions, depending if both lesions were in the same or a different FFPE block.

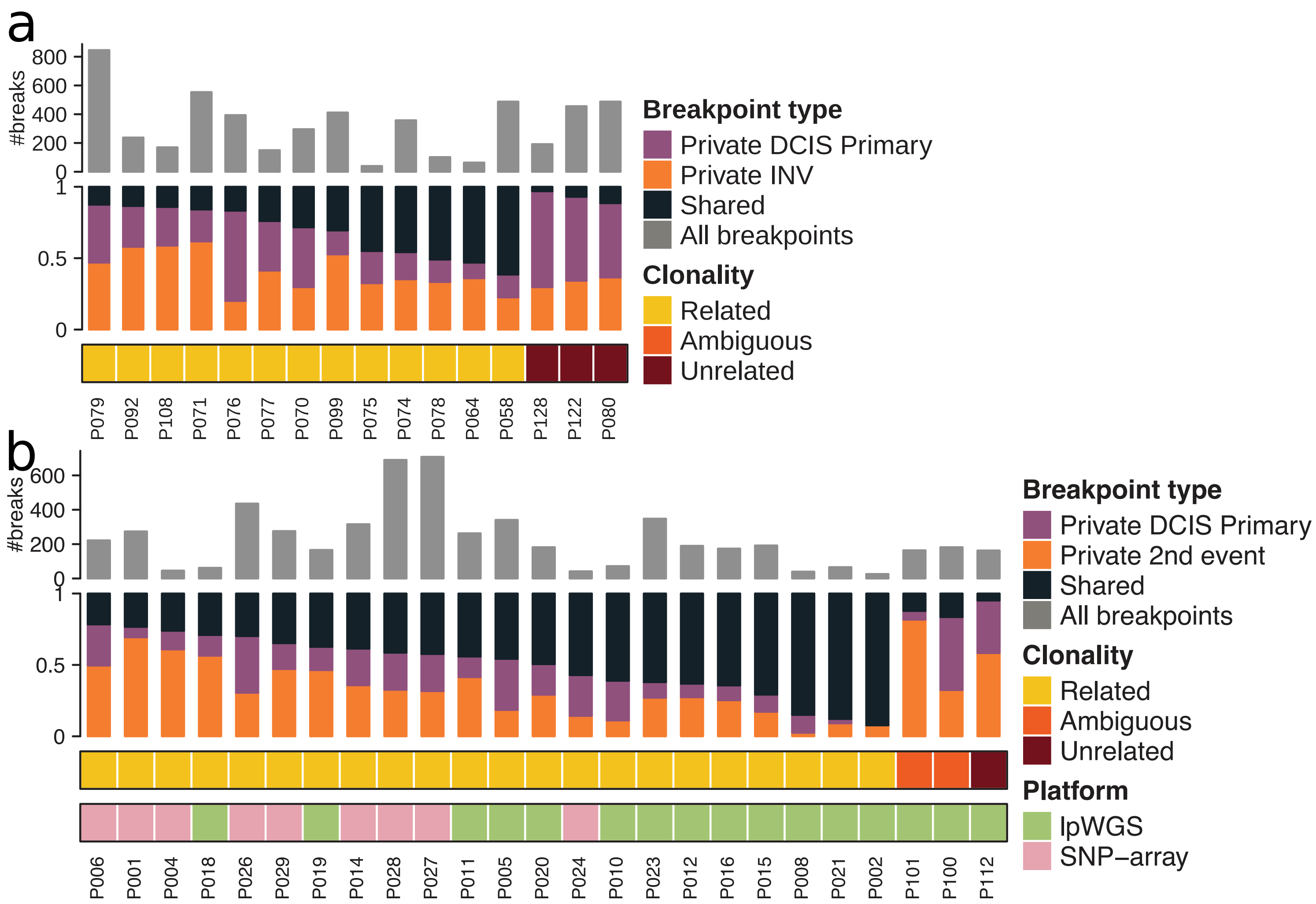


Figure S5. **Validation of clonality using copy number profiling.** **a**, Distribution of breakpoints in primary DCIS and paired subsequent invasive disease in samples analysed by SNP-array. The vertical bars indicate whether the breakpoints were unique (private) to the primary or subsequence or shared. Top grey bar indicates the total number of breakpoints for each patient. **b**, Distribution of breakpoints in 25 primary DCIS where the 2nd event was pure DCIS (DCIS→ipDCIS).

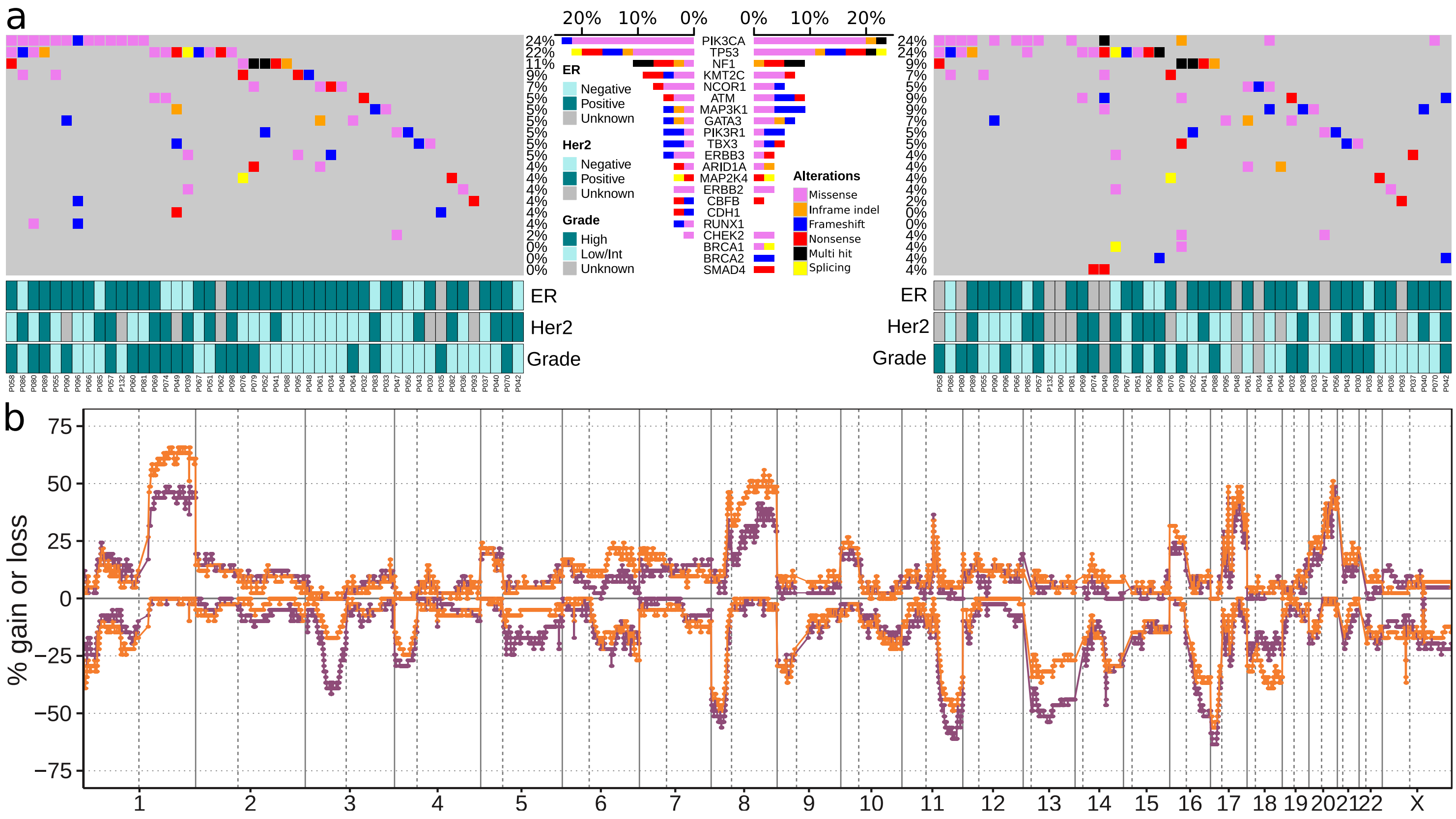


Figure S6. **Mutational spectrum and copy number alterations in clonally related primary DCIS and recurrent invasive disease pairs. a**, Oncoplots for primary DCIS samples (left) and subsequent invasive events (right) based on WES and targeted sequencing for clonally related pairs. Only genes mutated in more than 3% in primary DCIS or invasive events among the 45 sequenced genes covered by all sequencing platforms are kept and samples without mutations in these genes are not shown. C>T mutations with $AF < 0.1$ and less than 3 appearances in the COSMIC70 database were filtered out. **b**, Frequency plot of genome wide copy number alterations of clonally-related DCIS->ipINV pairs analysed by lpWGS ($n = 82$) - primary DCIS (purple) and its paired ipsilateral subsequent invasive disease (orange). The $y$-axis shows percentage of samples with gains (above zero line) and losses (below zero line). The genomic position is indicated by chromosome 1 on the left and up to chromosome X on the right with chromosome boundaries indicated by vertical lines.

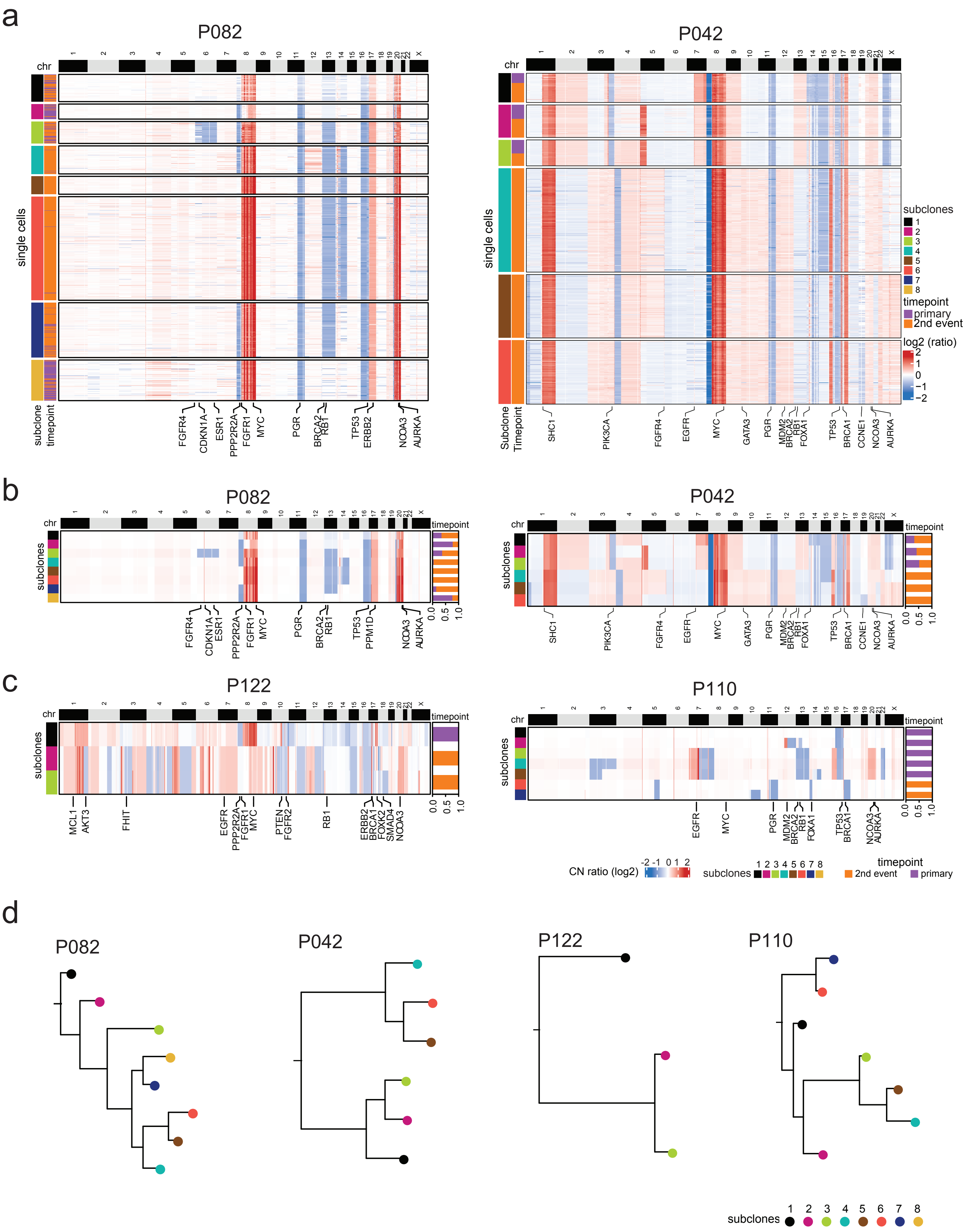
a
P082
P042
chr
single cells
subclone
timepoint
Subclone
Timepoint
FGFR4
CDKN1A
ESR1
PPP2R2A
FGFR1
MYC
PGR
BRCA2
RB1
TP53
ERBB2
NCOA3
AURKA
SHC1
PIK3CA
EGFR
GATA3
MDM2
FOXA1
BRCA1
CCNE1
subclones
timepoint
primary
2nd event
log2 (ratio)
b
subclones
PPM1D
c
P122
P110
MCL1
AKT3
FHIT
PTEN
FGFR2
FOXK2
SMAD4
CN ratio (log2)
d
P082
P042
P122
P110
subclones

Figure S7. **Additional clonal lineages inferred from single cell genome sequencing.** **a**, Clustered heatmaps of single cell copy number profiles in genomic order from two DCIS cases with related clonal lineages, with cluster and timepoint information on the right panels and selected breast cancer genes annotated below. **b-c**, Consensus copy number heatmaps of subclones calculated from clusters of single cell copy number profiles from clonally-related (**b**,) and clonally unrelated (**c**,) patients. **d**, Neighbor-joining trees of clonal lineages constructed from consensus subclones from clonally related and unrelated patients rooted by a diploid node.
