## Supplementary file 1A for "Genomic profiling defines variable clonal relatedness between invasive breast cancer and primary ductal carcinoma *in situ*"

# P001

| Syn/Meta | Time from 1st surgery to 2nd event (Months) | Side 2nd event | Histology 2nd event | Surgery | AdjuvanT Treatment Pri (RT/ HT) | ER Pri | ER 2nd event | Her2 Pri | Her2 2nd event | Grade Pri | Grade 2nd event | Quadrant 2nd event | Margins | Screening | Clonality P value Copy N | Clonality P value Panel seq | Clonality P value WES | Final verdict |
|---|---|---|---|---|---|---|---|---|---|---|---|---|---|---|---|---|---|---|
| metachronous | 63 | Ipsilateral | DCIS only | lumpectomy | RT | - | NA | + | + | 3 | NA | NA | Clear | screen-detected | 0.000466 | 0.05 | 0.00075244 | Related |

### Primary event

**LogR**

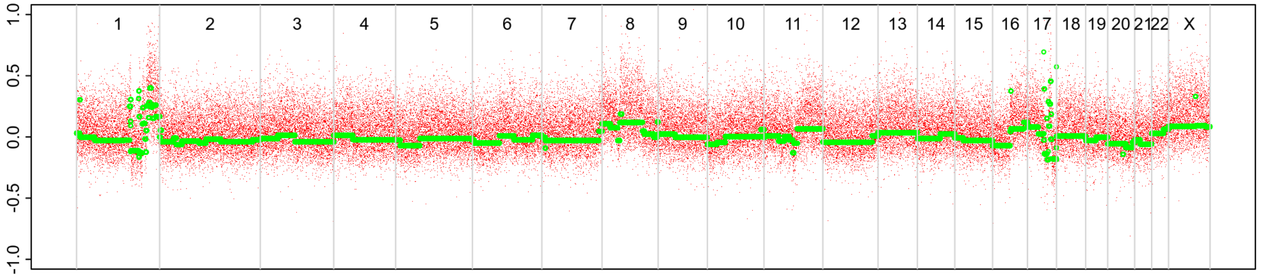


**BAF**

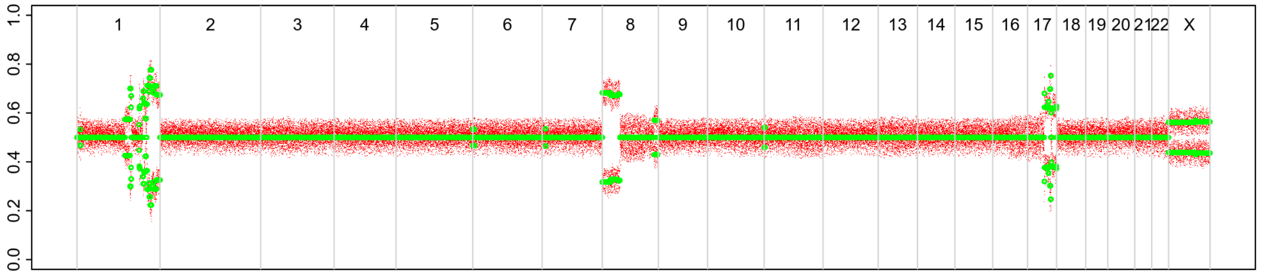


### 2nd event

**LogR**

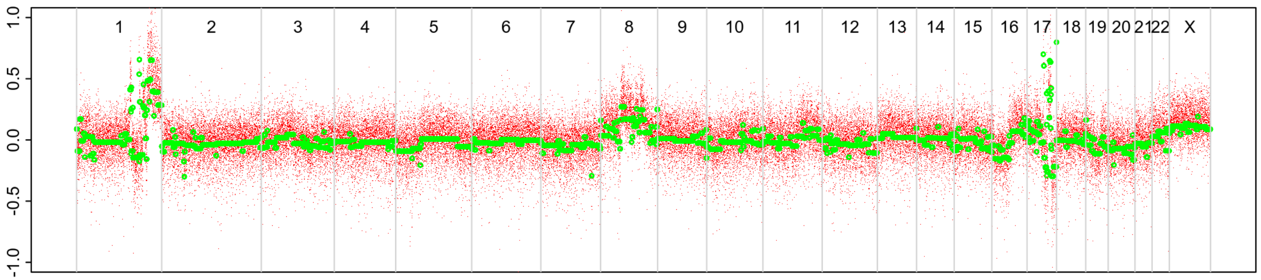


**BAF**

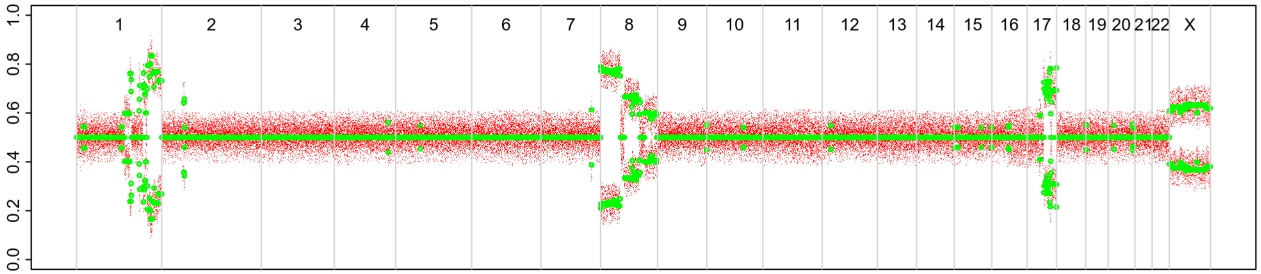

# P002

| Syn/Meta | Time from 1st surgery to 2nd event (Months) | Side 2nd event | Histology 2nd event | Surgery | AdjuvanT Treatment Pri (RT/ HT) | ER Pri | ER 2nd event | Her2 Pri | Her2 2nd event | Grade Pri | Grade 2nd event | Quadrant 2nd event | Margins | Screening | Clonality P value Copy N | Clonality P value Panel seq | Clonality P value WES | Final verdict |
|---|---|---|---|---|---|---|---|---|---|---|---|---|---|---|---|---|---|---|
| metachronous | 72 | Ipsilateral | DCIS only | lumpectomy | None | NA | NA | NA | NA | 3 | 3 | Unknown | Clear | screen-detected | 0.003663 | NA | NA | Related |

### Primary event

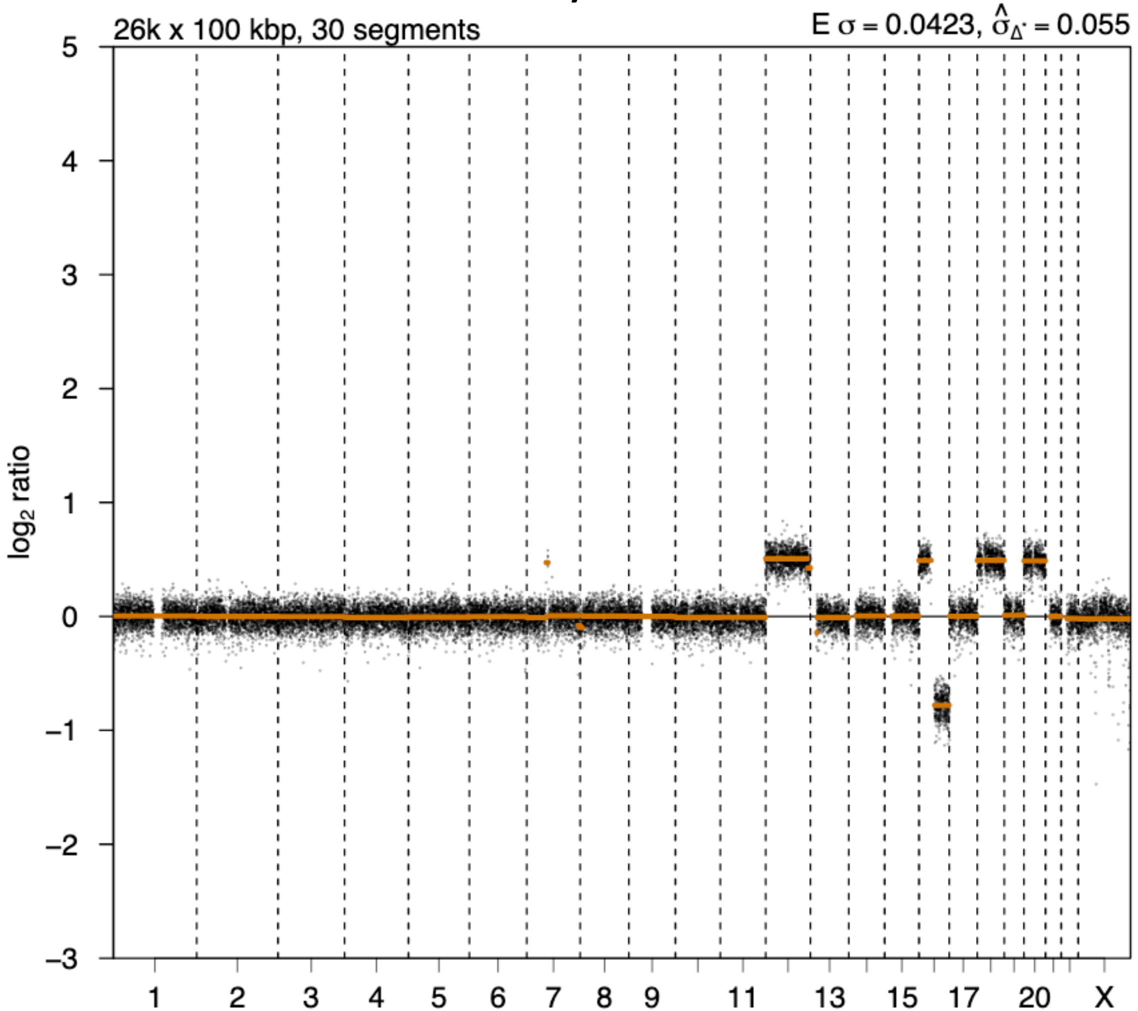


### 2nd event

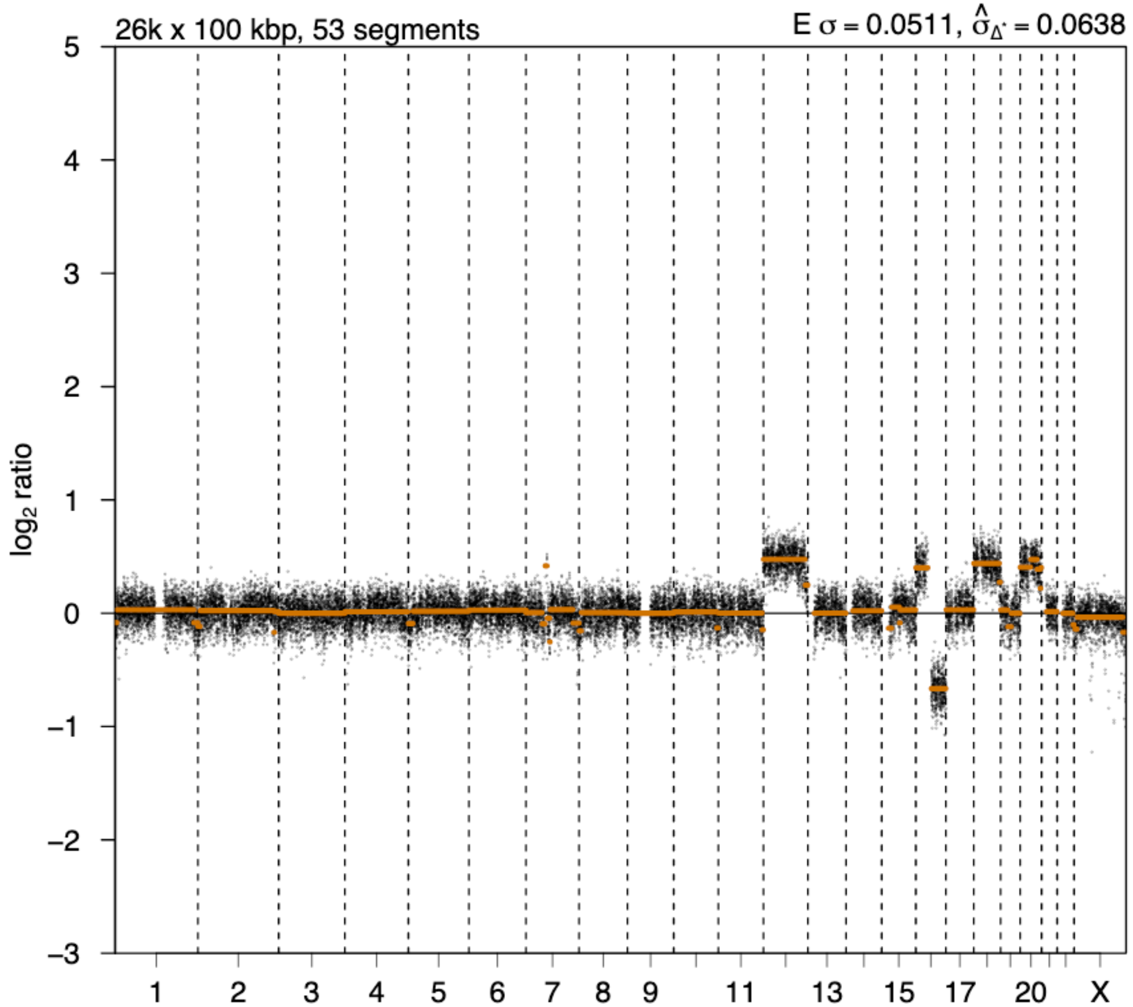

# P004

| Syn/Meta | Time from 1st surgery to 2nd event (Months) | Side 2nd event | Histology 2nd event | Surgery | AdjuvanT Treatment Pri (RT/ HT) | ER Pri | ER 2nd event | Her2 Pri | Her2 2nd event | Grade Pri | Grade 2nd event | Quadrant 2nd event | Margins | Screening | Clonality P value Copy N | Clonality P value Panel seq | Clonality P value WES | Final verdict |
|---|---|---|---|---|---|---|---|---|---|---|---|---|---|---|---|---|---|---|
| metachronous | 37 | Ipsilateral | DCIS only | lumpectomy | RT | + | NA | - | NA | 2 | NA | NA | Clear | screen-detected | 0.002331 | NA | NA | Related |

### Primary event

LogR

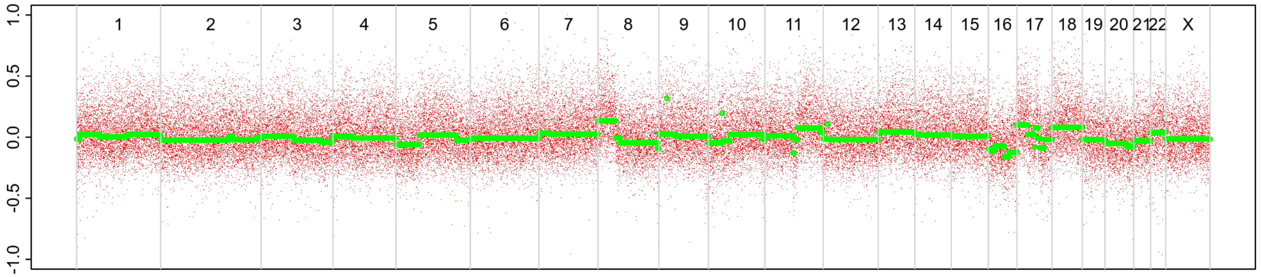


BAF

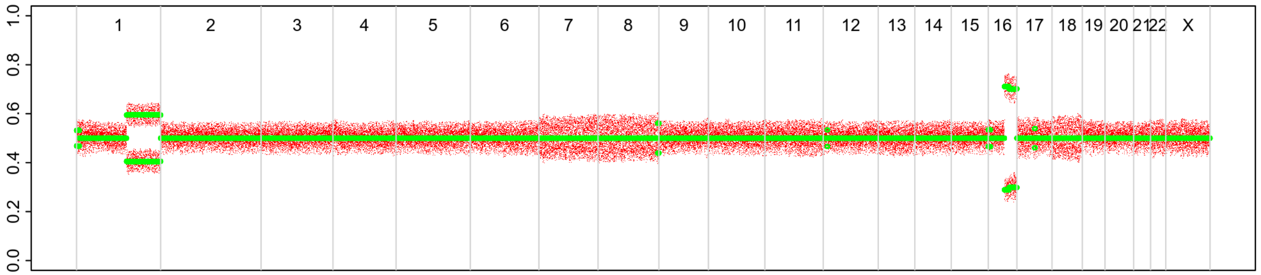


### 2nd event

LogR

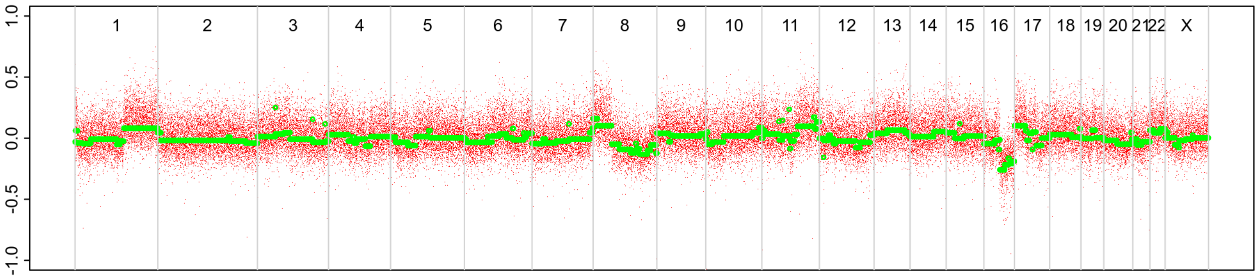


BAF

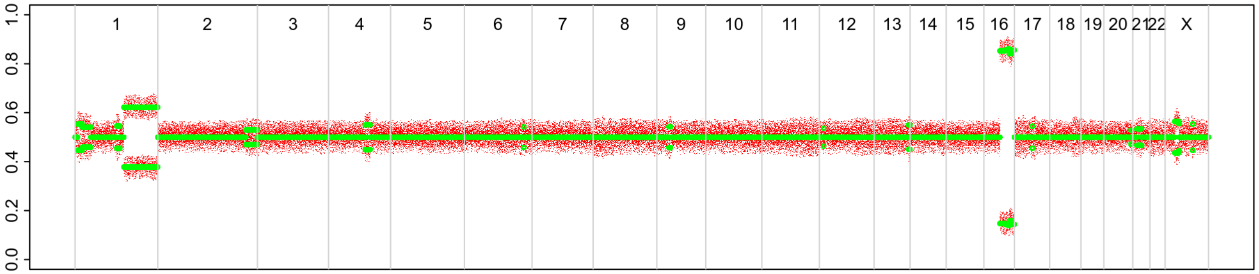

# P005

| Syn/Meta | Time from 1st surgery to 2nd event (Months) | Side 2nd event | Histology 2nd event | Surgery | AdjuvanT Treatment Pri (RT/ HT) | ER Pri | ER 2nd event | Her2 Pri | Her2 2nd event | Grade Pri | Grade 2nd event | Quadrant 2nd event | Margins | Screening | Clonality P value Copy N | Clonality P value Panel seq | Clonality P value WES | Final verdict |
|---|---|---|---|---|---|---|---|---|---|---|---|---|---|---|---|---|---|---|
| metachronous | 35 | Ipsilateral | DCIS only | lumpectomy | RT | NA | NA | + | NA | 3 | 3 | NA | Clear | screen-detected | 0.003663 | NA | NA | Related |

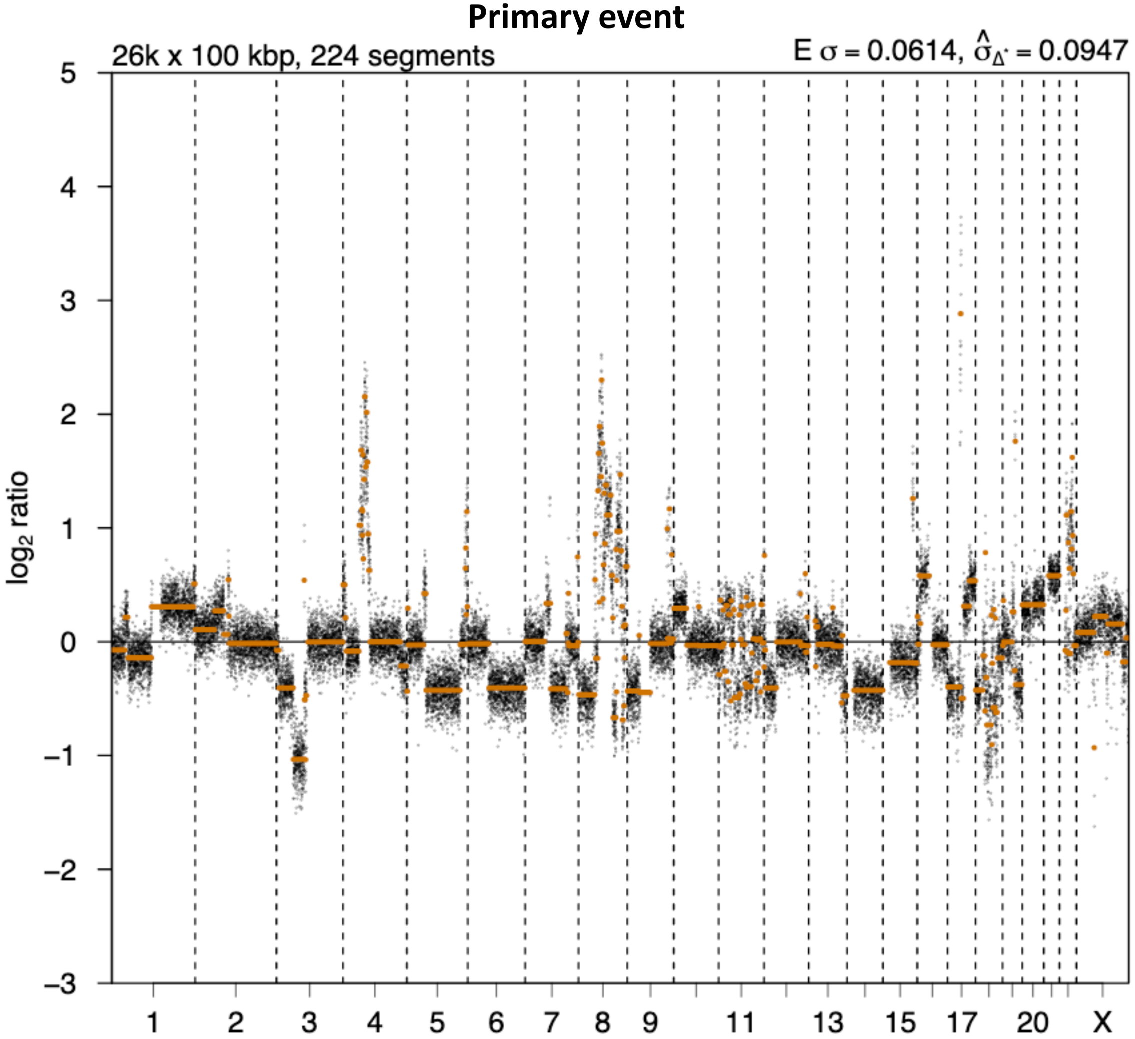


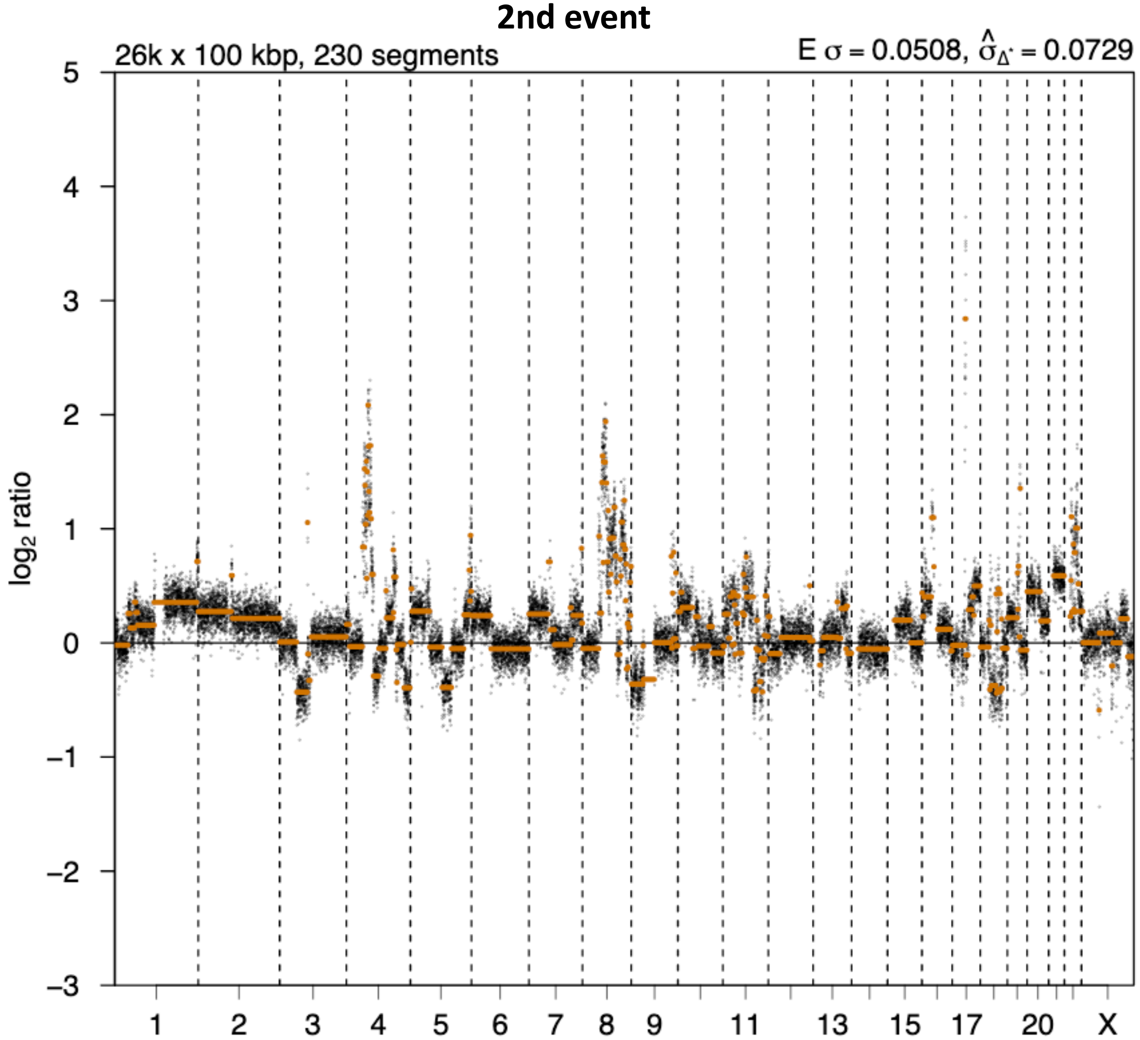

# P006

| Syn/Meta | Time from 1st surgery to 2nd event (Months) | Side 2nd event | Histology 2nd event | Surgery | AdjuvanT Treatment Pri (RT/ HT) | ER Pri | ER 2nd event | Her2 Pri | Her2 2nd event | Grade Pri | Grade 2nd event | Quadrant 2nd event | Margins | Screening | Clonality P value Copy N | Clonality P value Panel seq | Clonality P value WES | Final verdict |
|---|---|---|---|---|---|---|---|---|---|---|---|---|---|---|---|---|---|---|
| metachronous | 46 | Ipsilateral | DCIS only | lumpectomy | RTHT | + | NA | NA | NA | 3 | 3 | NA | <2mm | screen-detected | 0.000466 | 0 | 0.00075244 | Related |

### Primary event

LogR

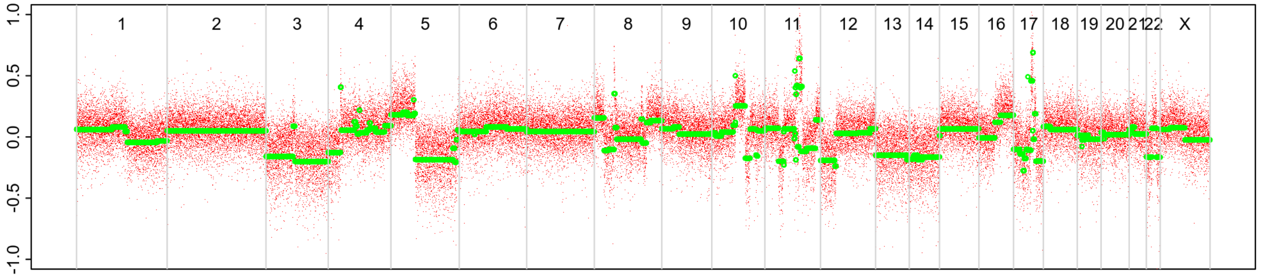


BAF

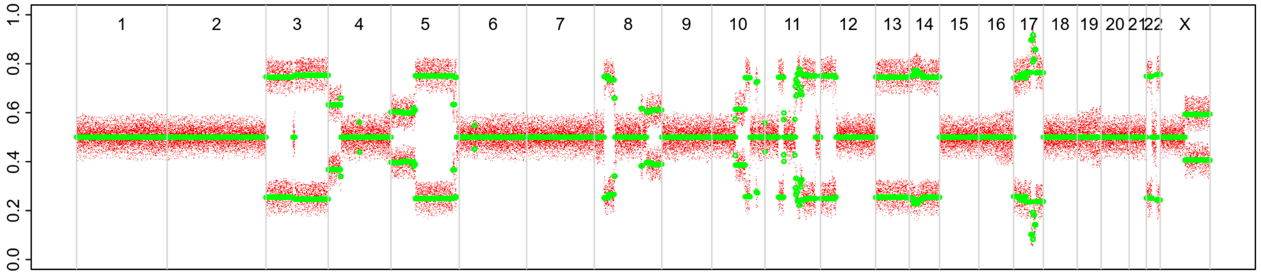


### 2nd event

LogR

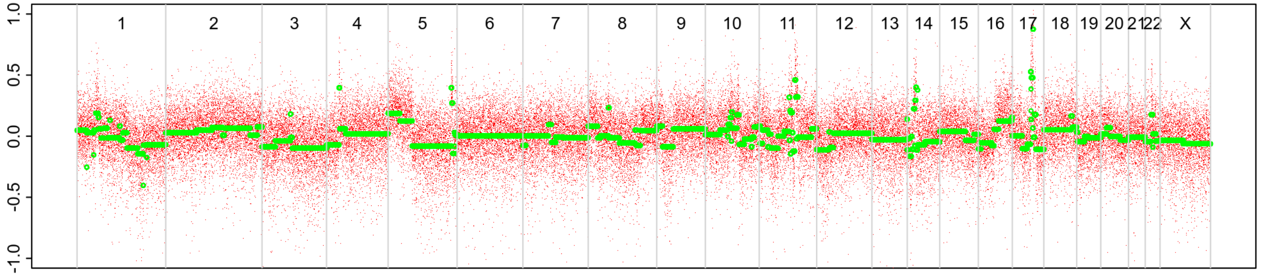


BAF

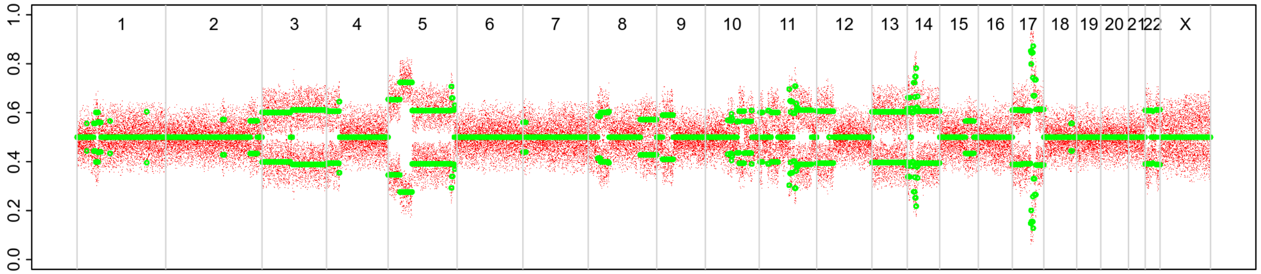

# P008

| Syn/Meta | Time from 1st surgery to 2nd event (Months) | Side 2nd event | Histology 2nd event | Surgery | AdjuvanT Treatment Pri (RT/ HT) | ER Pri | ER 2nd event | Her2 Pri | Her2 2nd event | Grade Pri | Grade 2nd event | Quadrant 2nd event | Margins | Screening | Clonality P value Copy N | Clonality P value Panel seq | Clonality P value WES | Final verdict |
|---|---|---|---|---|---|---|---|---|---|---|---|---|---|---|---|---|---|---|
| metachronous | 48 | Ipsilateral | DCIS only | lumpectomy | HT | + | + | - | NA | 3 | 3 | NA | NA | screen-detected | 0.003663 | NA | NA | Related |

### Primary event

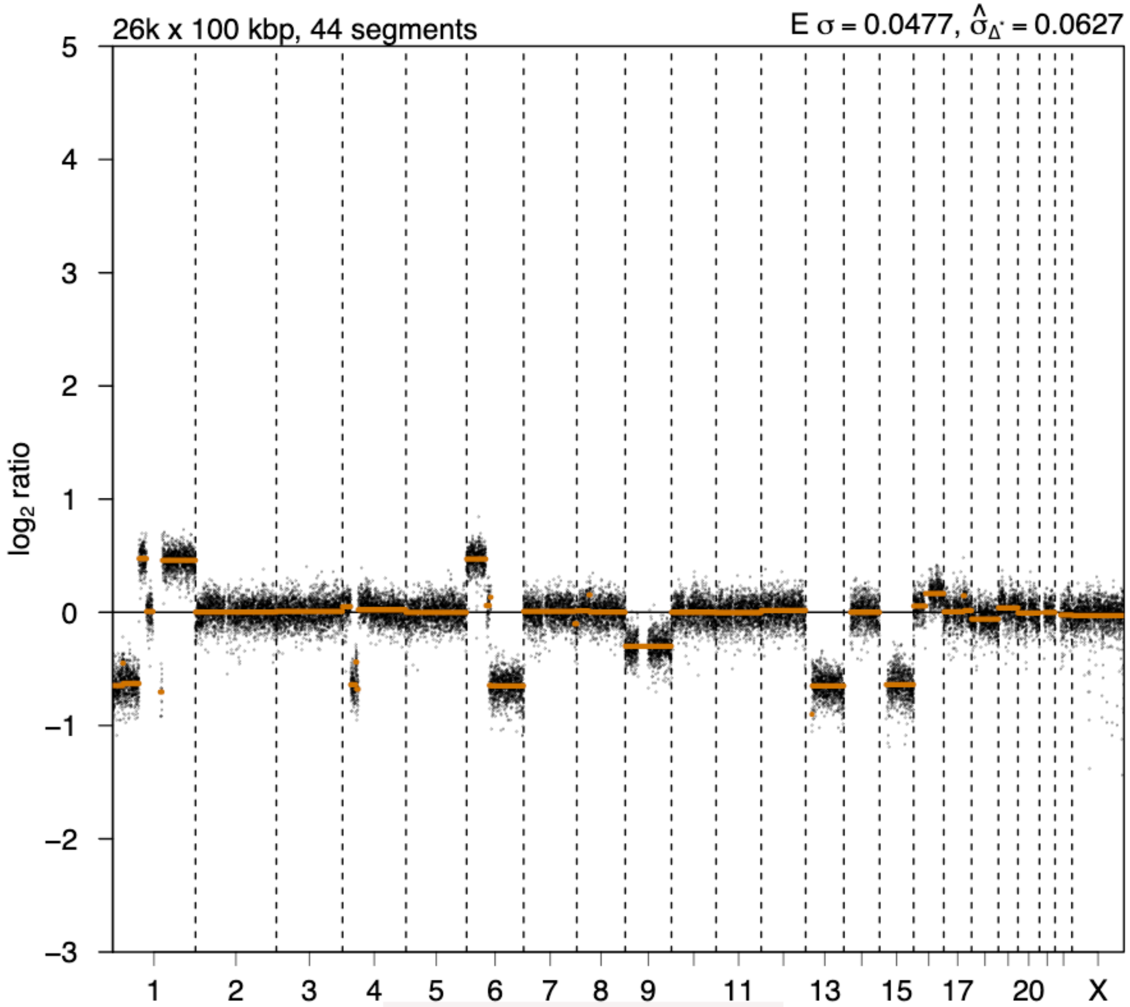


### 2nd event

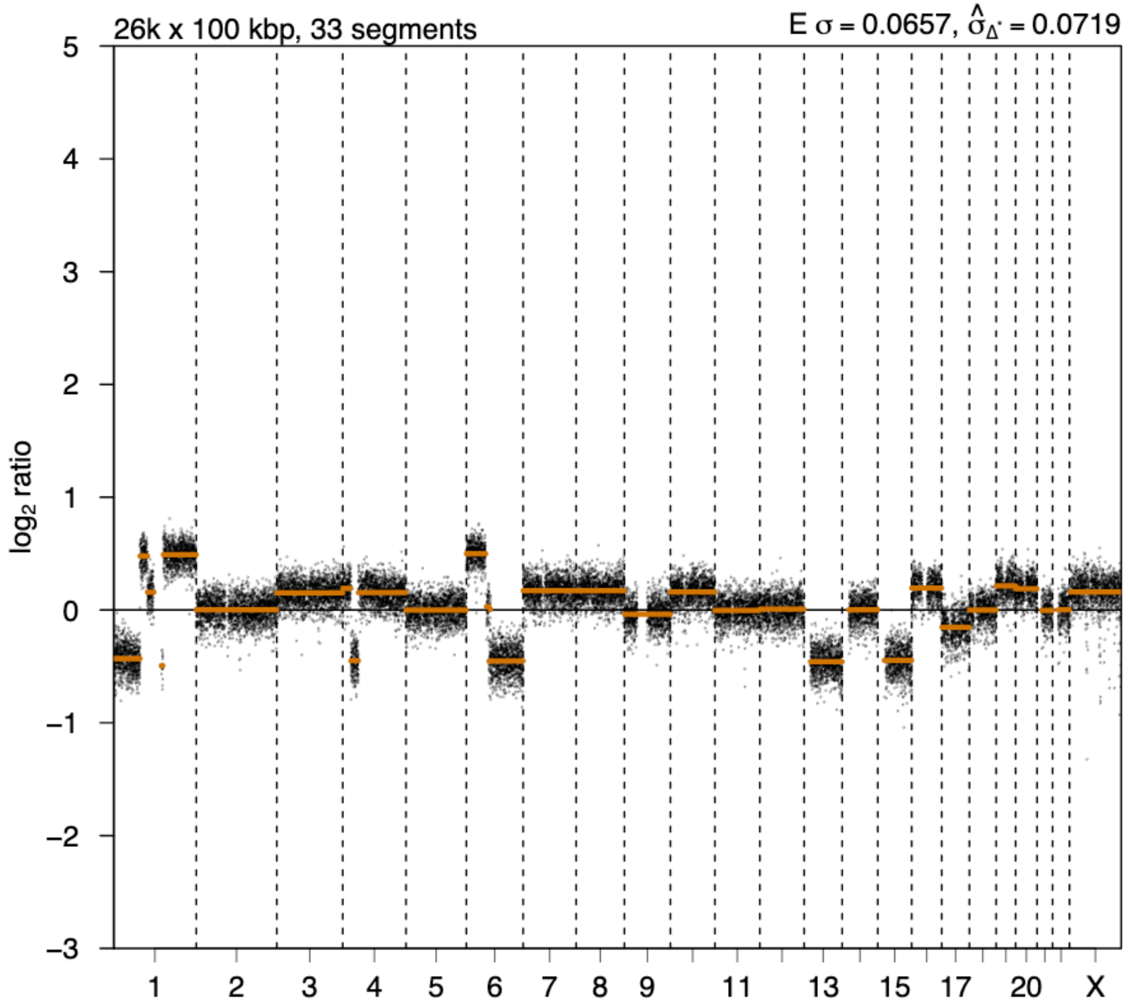

# P010

| Syn/Meta | Time from 1st surgery to 2nd event (Months) | Side 2nd event | Histology 2nd event | Surgery | AdjuvanT Treatment Pri (RT/ HT) | ER Pri | ER 2nd event | Her2 Pri | Her2 2nd event | Grade Pri | Grade 2nd event | Quadrant 2nd event | Margins | Screening | Clonality P value Copy N | Clonality P value Panel seq | Clonality P value WES | Final verdict |
|---|---|---|---|---|---|---|---|---|---|---|---|---|---|---|---|---|---|---|
| metachronous | 11 | Ipsilateral | DCIS only | lumpectomy | RT | + | - | NA | NA | 2 | 3 | NA | Clear | screen-detected | 0.003663 | NA | NA | Related |

**Primary event**

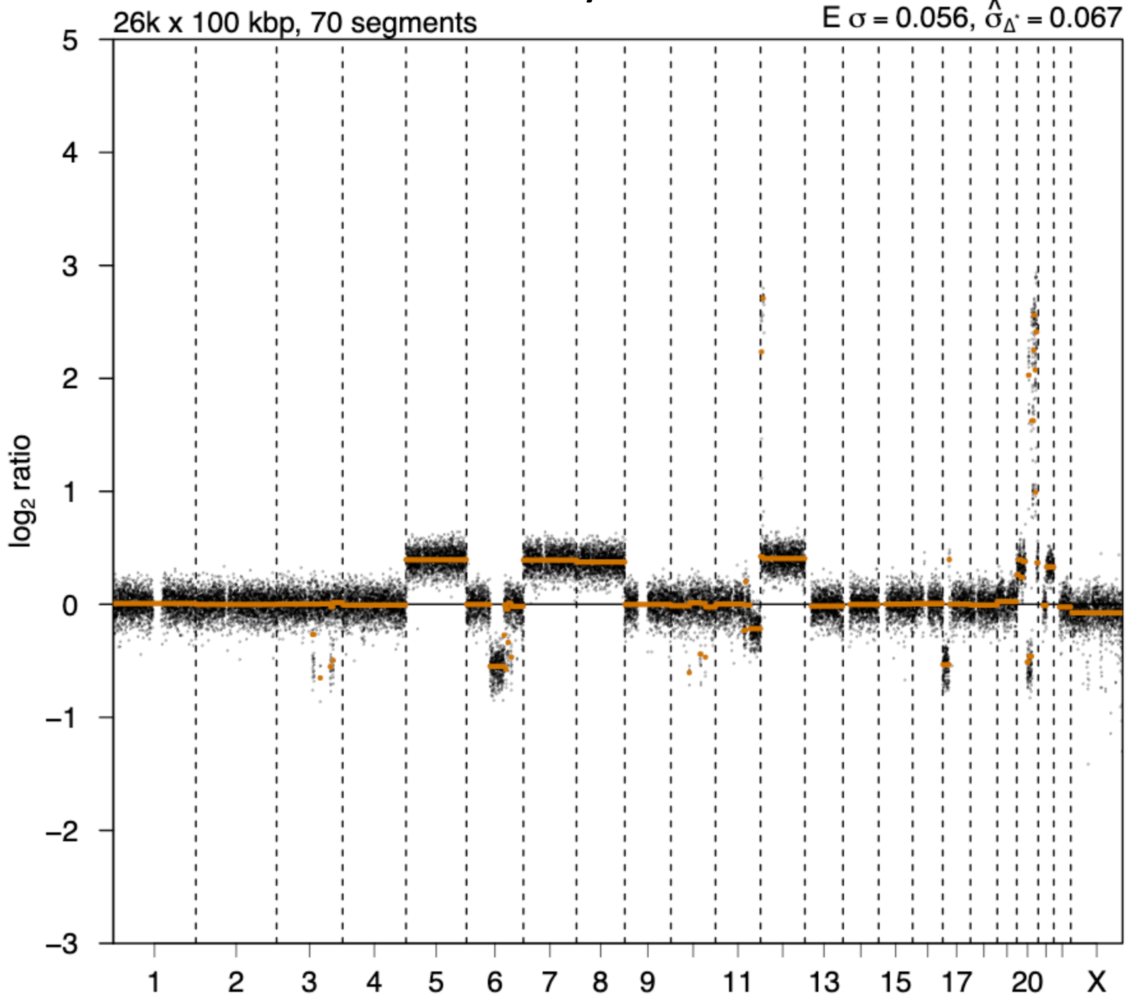


**2nd event**

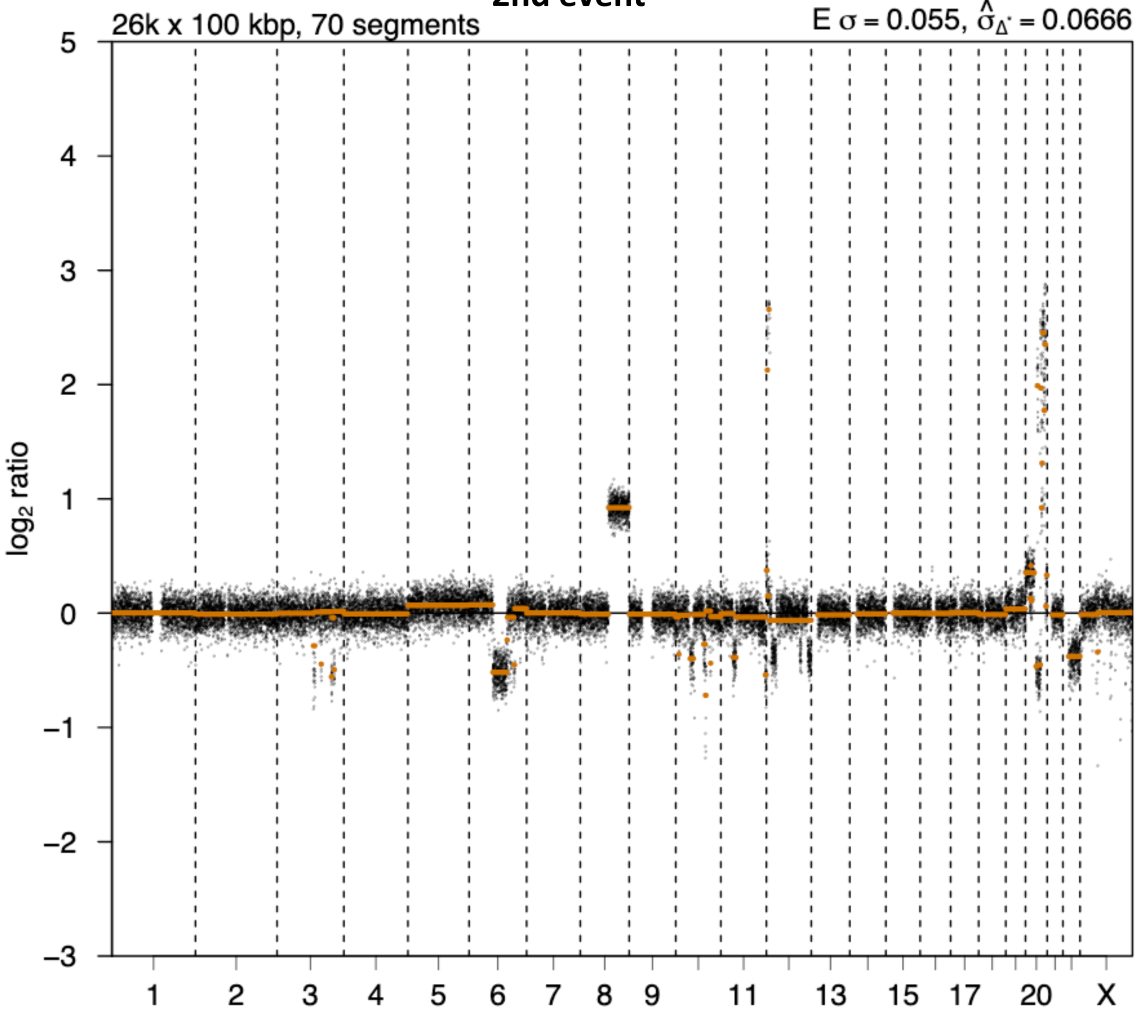

# P011

| Syn/Meta | Time from 1st surgery to 2nd event (Months) | Side 2nd event | Histology 2nd event | Surgery | AdjuvanT Treatment Pri (RT/ HT) | ER Pri | ER 2nd event | Her2 Pri | Her2 2nd event | Grade Pri | Grade 2nd event | Quadrant 2nd event | Margins | Screening | Clonality P value Copy N | Clonality P value Panel seq | Clonality P value WES | Final verdict |
|---|---|---|---|---|---|---|---|---|---|---|---|---|---|---|---|---|---|---|
| metachronous | 25 | Ipsilateral | DCIS only | lumpectomy | RT | - | - | + | NA | 3 | 3 | NA | Clear | screen-detected | 0.003663 | NA | NA | Related |

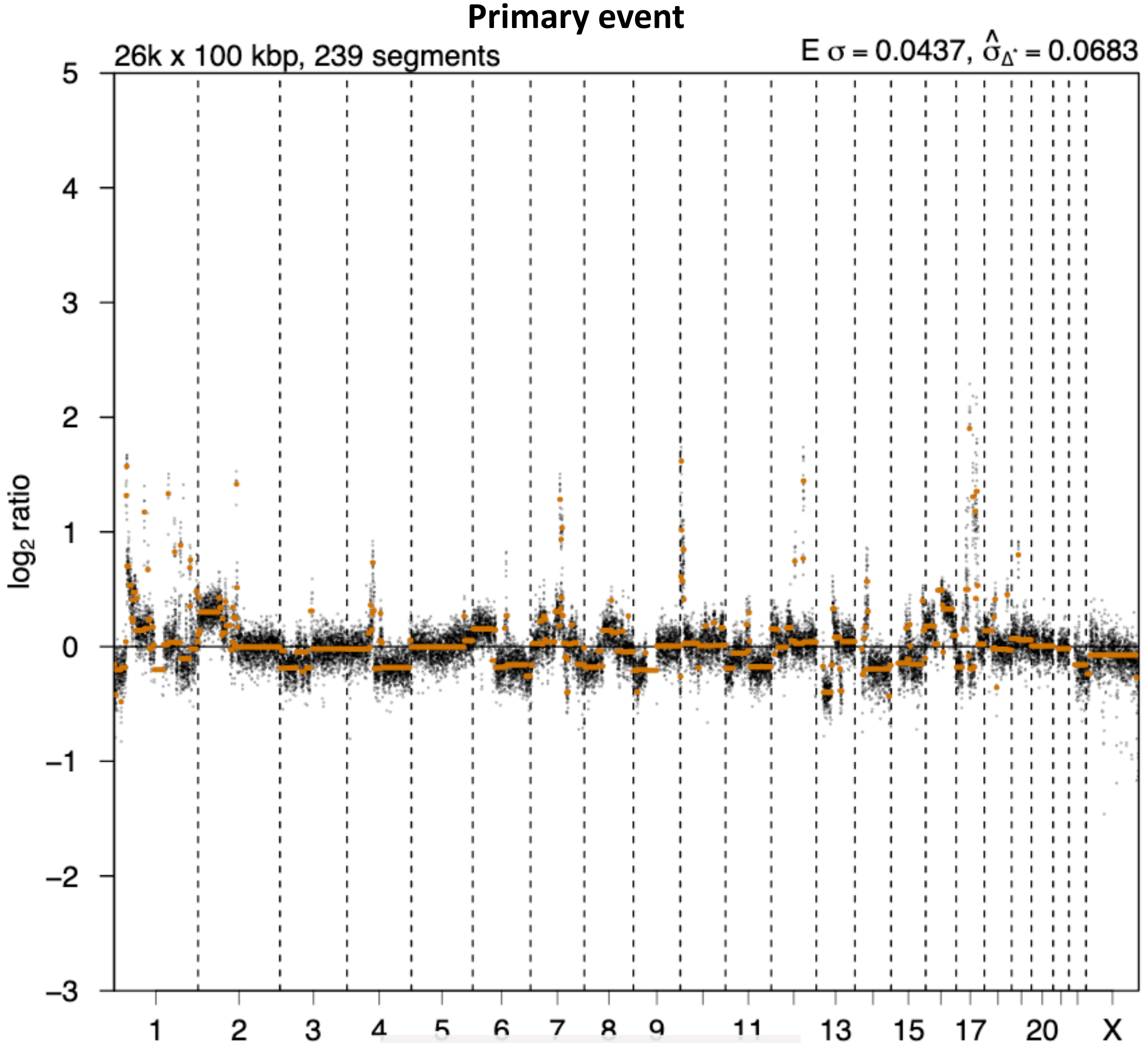


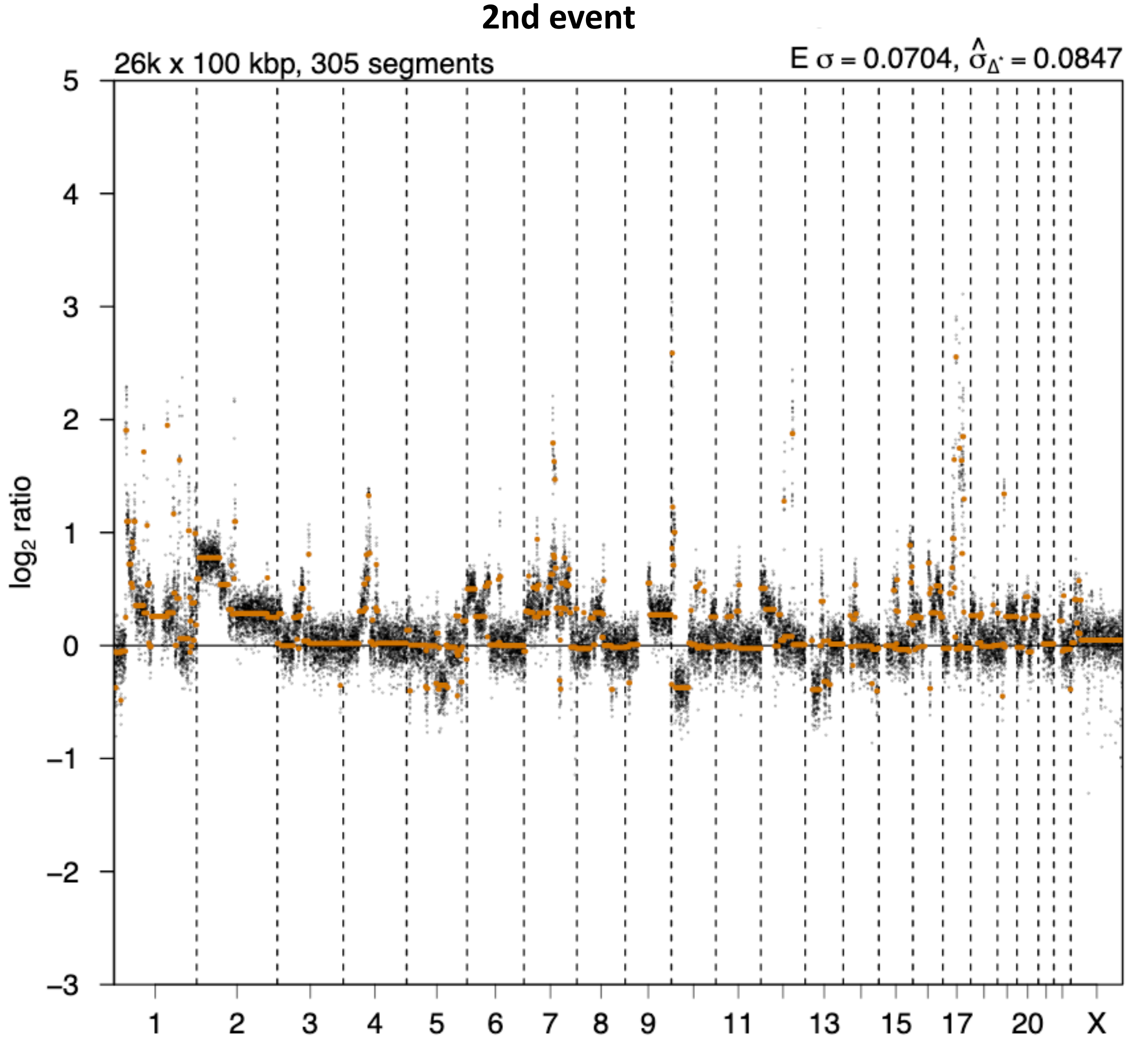

# P012

| Syn/Meta | Time from 1st surgery to 2nd event (Months) | Side 2nd event | Histology 2nd event | Surgery | AdjuvanT Treatment Pri (RT/ HT) | ER Pri | ER 2nd event | Her2 Pri | Her2 2nd event | Grade Pri | Grade 2nd event | Quadrant 2nd event | Margins | Screening | Clonality P value Copy N | Clonality P value Panel seq | Clonality P value WES | Final verdict |
|---|---|---|---|---|---|---|---|---|---|---|---|---|---|---|---|---|---|---|
| metachronous | 24 | Ipsilateral | DCIS only | lumpectomy | RT | - | NA | + | NA | 3 | 3 | NA | <2mm | screen-detected | 0.003663004 | NA | NA | Related |

### Primary event

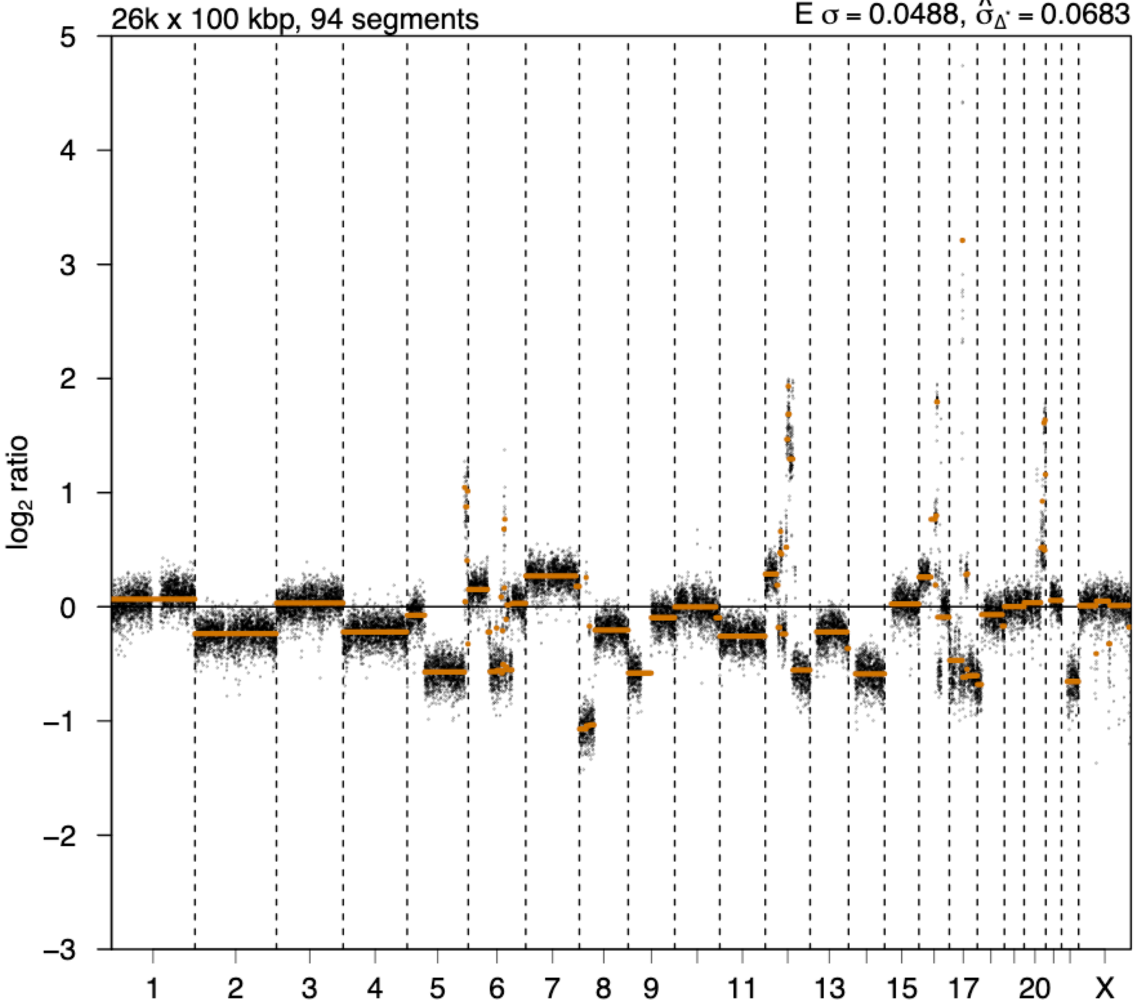


### 2nd event

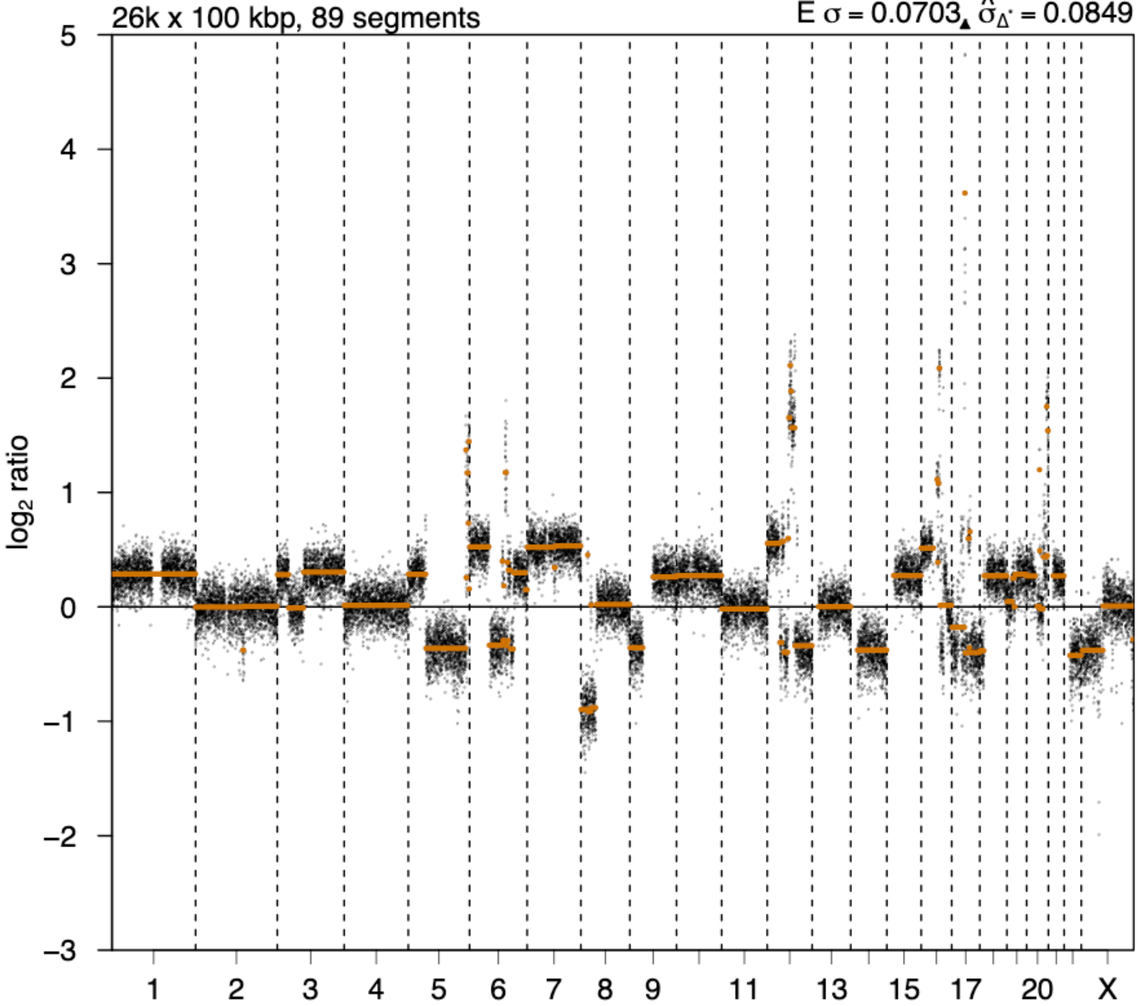

# P014

| Syn/Meta | Time from 1st surgery to 2nd event (Months) | Side 2nd event | Histology 2nd event | Surgery | AdjuvanT Treatment Pri (RT/ HT) | ER Pri | ER 2nd event | Her2 Pri | Her2 2nd event | Grade Pri | Grade 2nd event | Quadrant 2nd event | Margins | Screening | Clonality P value Copy N | Clonality P value Panel seq | Clonality P value WES | Final verdict |
|---|---|---|---|---|---|---|---|---|---|---|---|---|---|---|---|---|---|---|
| metachronous | 20 | Ipsilateral | DCIS only | lumpectomy | None | + | NA | NA | NA | 3 | 3 | NA | Clear | screen-detected | 0.000466 | 0.16 | NA | Related |

### Primary event

**LogR**

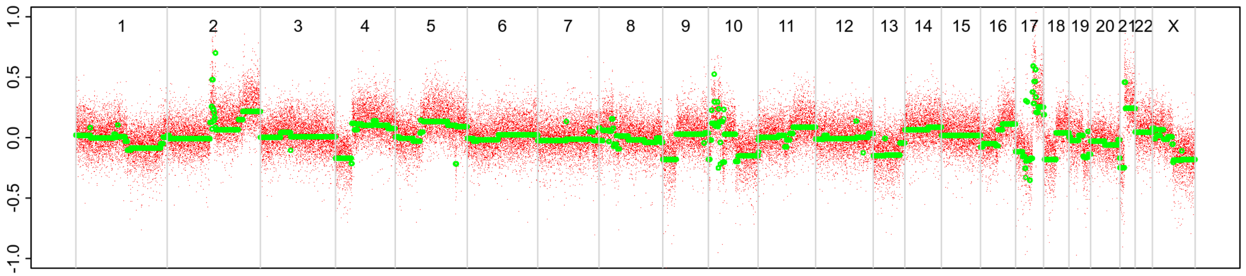


**BAF**

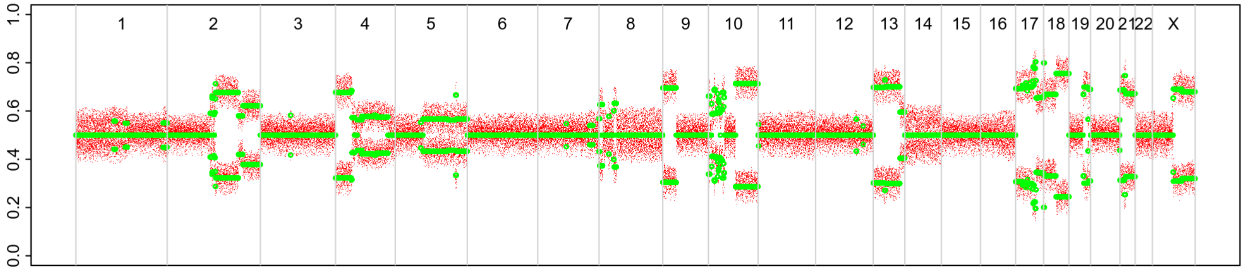


### 2nd event

**LogR**

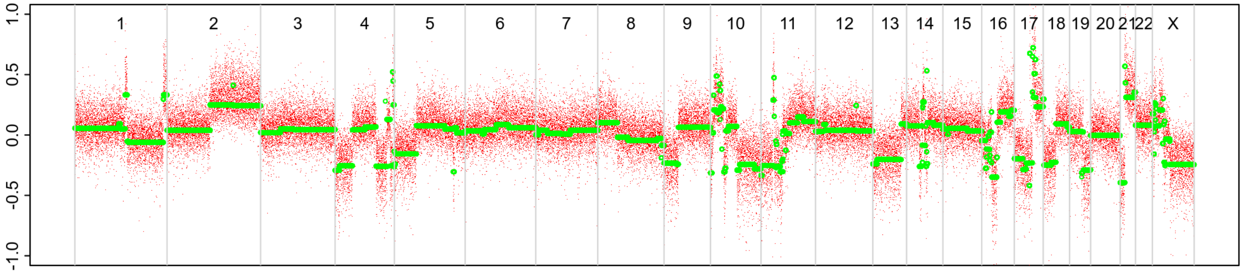


**BAF**

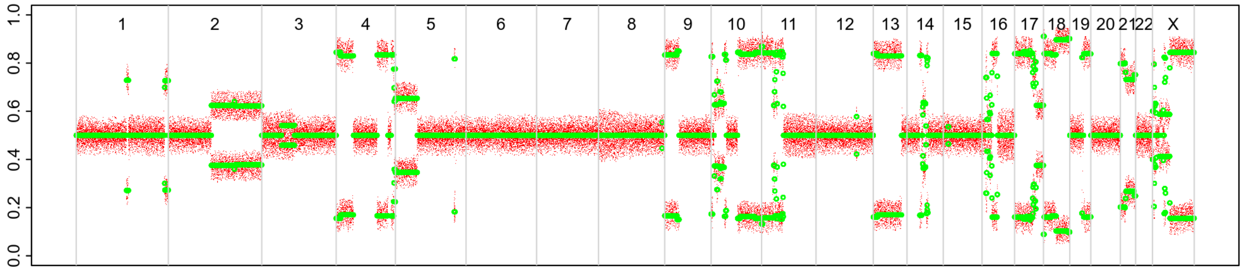

# P015

| Syn/Meta | Time from 1st surgery to 2nd event (Months) | Side 2nd event | Histology 2nd event | Surgery | AdjuvanT Treatment Pri (RT/ HT) | ER Pri | ER 2nd event | Her2 Pri | Her2 2nd event | Grade Pri | Grade 2nd event | Quadrant 2nd event | Margins | Screening | Clonality P value Copy N | Clonality P value Panel seq | Clonality P value WES | Final verdict |
|---|---|---|---|---|---|---|---|---|---|---|---|---|---|---|---|---|---|---|
| metachronous | 13 | Ipsilateral | DCIS only | lumpectomy | None | + | - | NA | NA | 3 | 2 | NA | Clear | screen-detected | 0.003663004 | NA | 0.000752445 | Related |

### Primary event

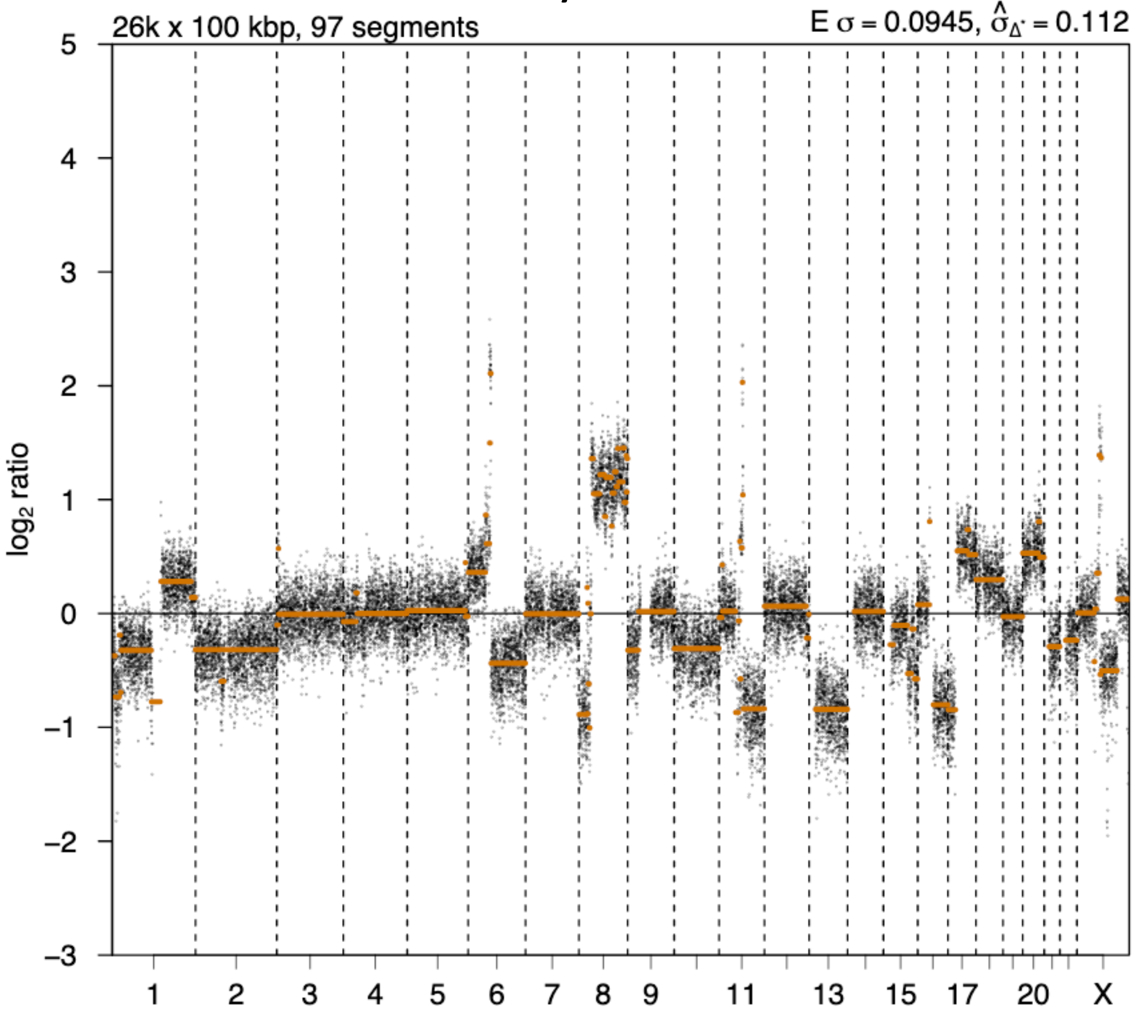


### 2nd event

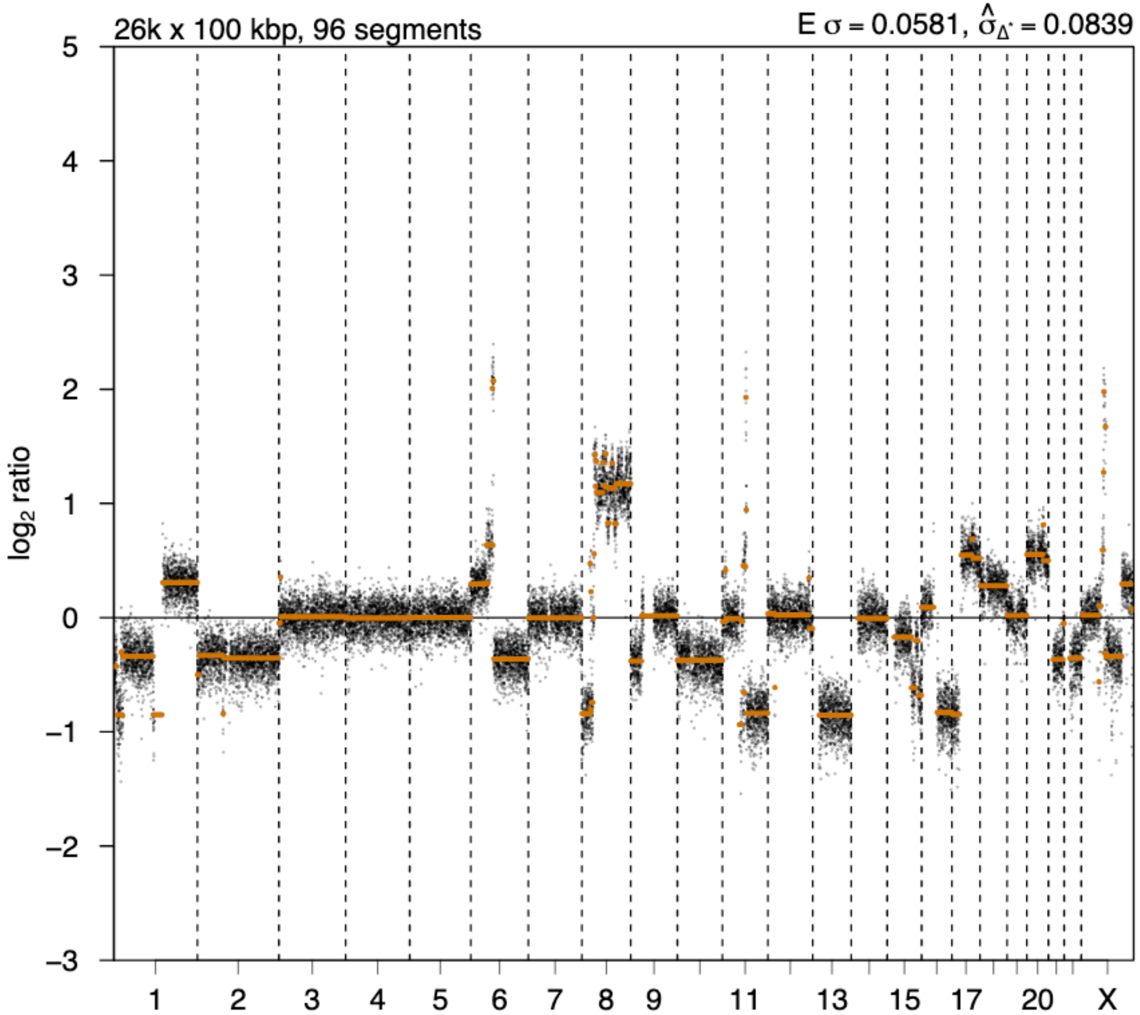

# P016

| Syn/Meta | Time from 1st surgery to 2nd event (Months) | Side 2nd event | Histology 2nd event | Surgery | AdjuvanT Treatment Pri (RT/ HT) | ER Pri | ER 2nd event | Her2 Pri | Her2 2nd event | Grade Pri | Grade 2nd event | Quadrant 2nd event | Margins | Screening | Clonality P value Copy N | Clonality P value Panel seq | Clonality P value WES | Final verdict |
|---|---|---|---|---|---|---|---|---|---|---|---|---|---|---|---|---|---|---|
| metachronous | 20 | Ipsilateral | DCIS only | lumpectomy | RT | + | + | NA | NA | 3 | 3 | NA | Clear | screen-detected | 0.003663004 | NA | NA | Related |

### Primary event

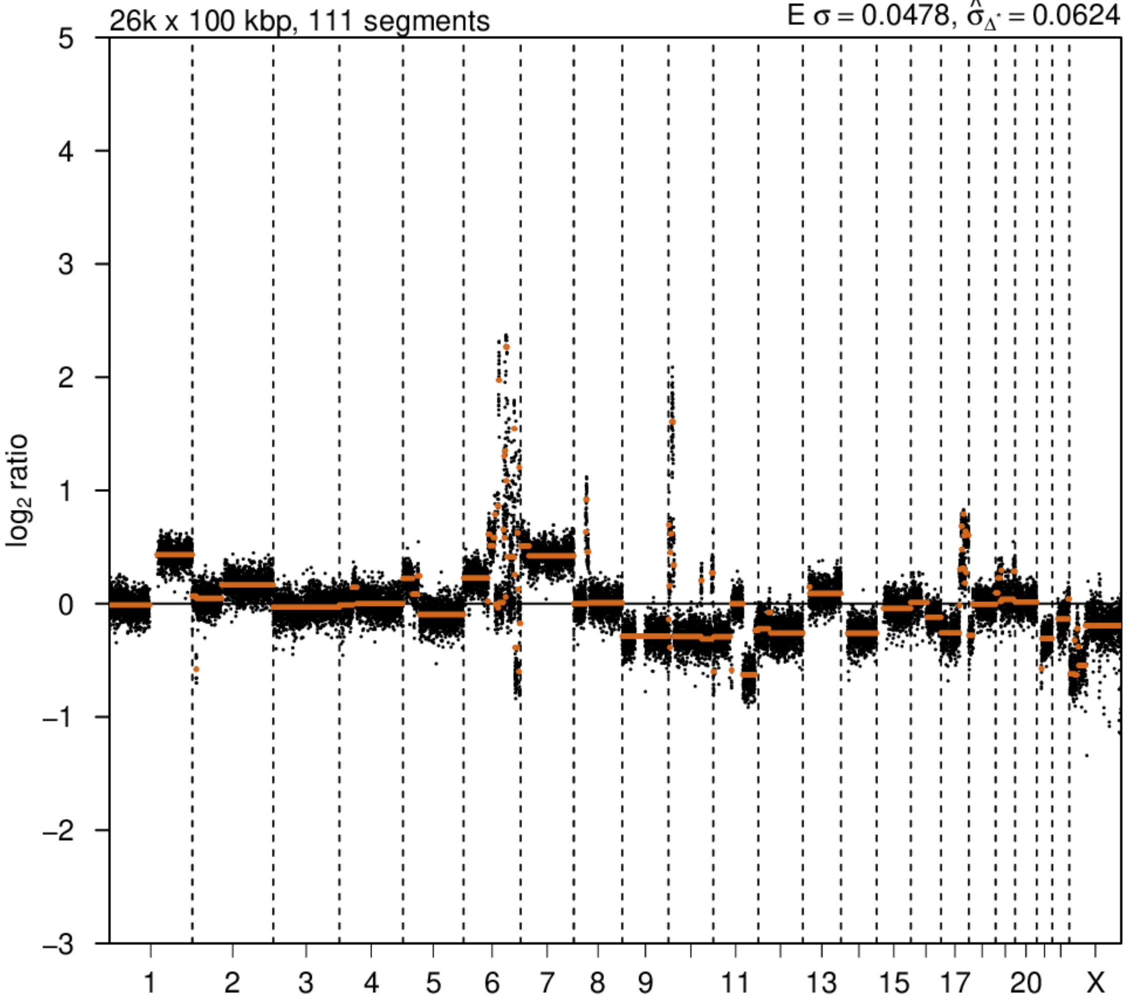


### 2nd event

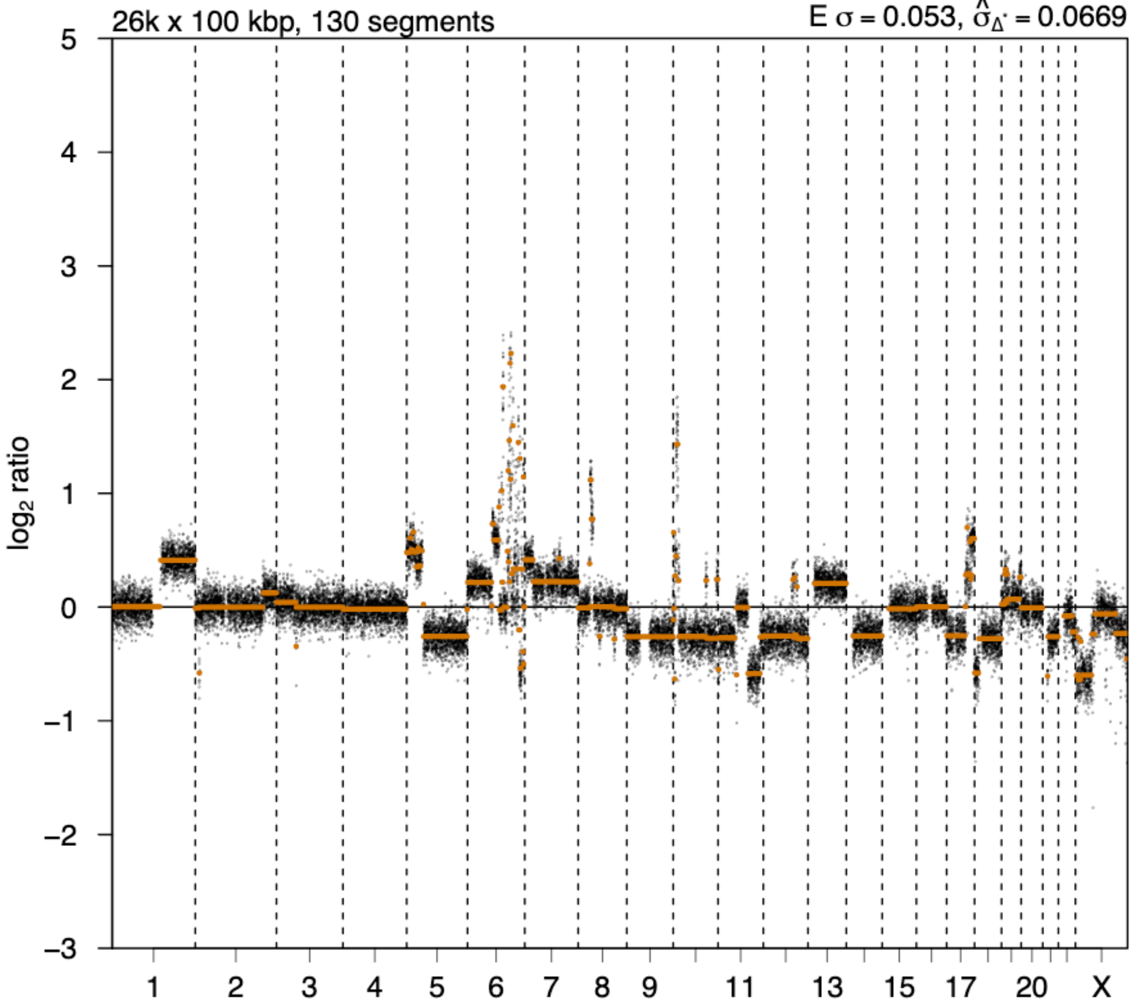

# P018

| Syn/Meta | Time from 1st surgery to 2nd event (Months) | Side 2nd event | Histology 2nd event | Surgery | AdjuvanT Treatment Pri (RT/ HT) | ER Pri | ER 2nd event | Her2 Pri | Her2 2nd event | Grade Pri | Grade 2nd event | Quadrant 2nd event | Margins | Screening | Clonality P value Copy N | Clonality P value Panel seq | Clonality P value WES | Final verdict |
|---|---|---|---|---|---|---|---|---|---|---|---|---|---|---|---|---|---|---|
| metachronous | 24 | Ipsilateral | DCIS only | lumpectomy | None | NA | NA | NA | NA | 3 | 3 | NA | NA | screen-detected | 0.003663004 | NA | NA | Related |

Primary event

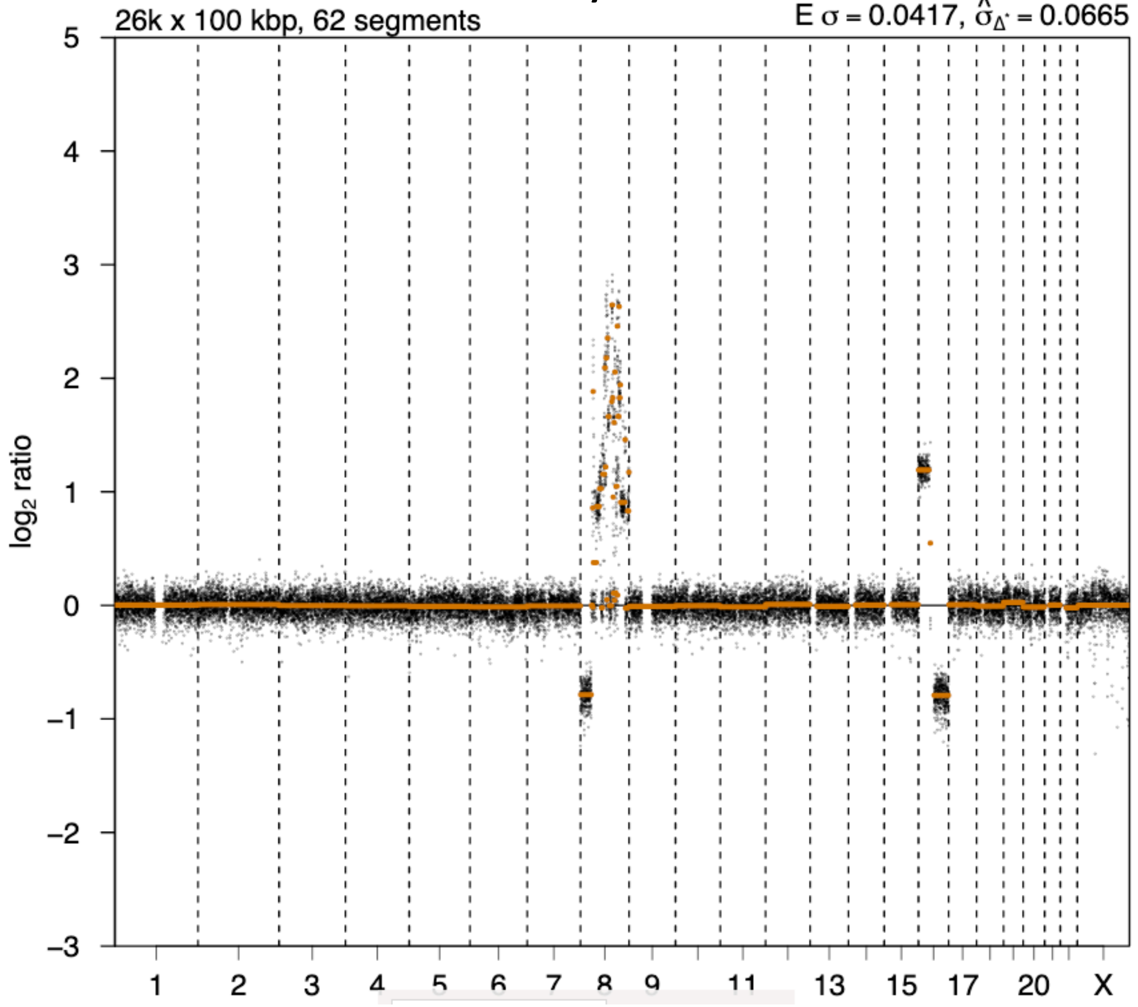


2nd event

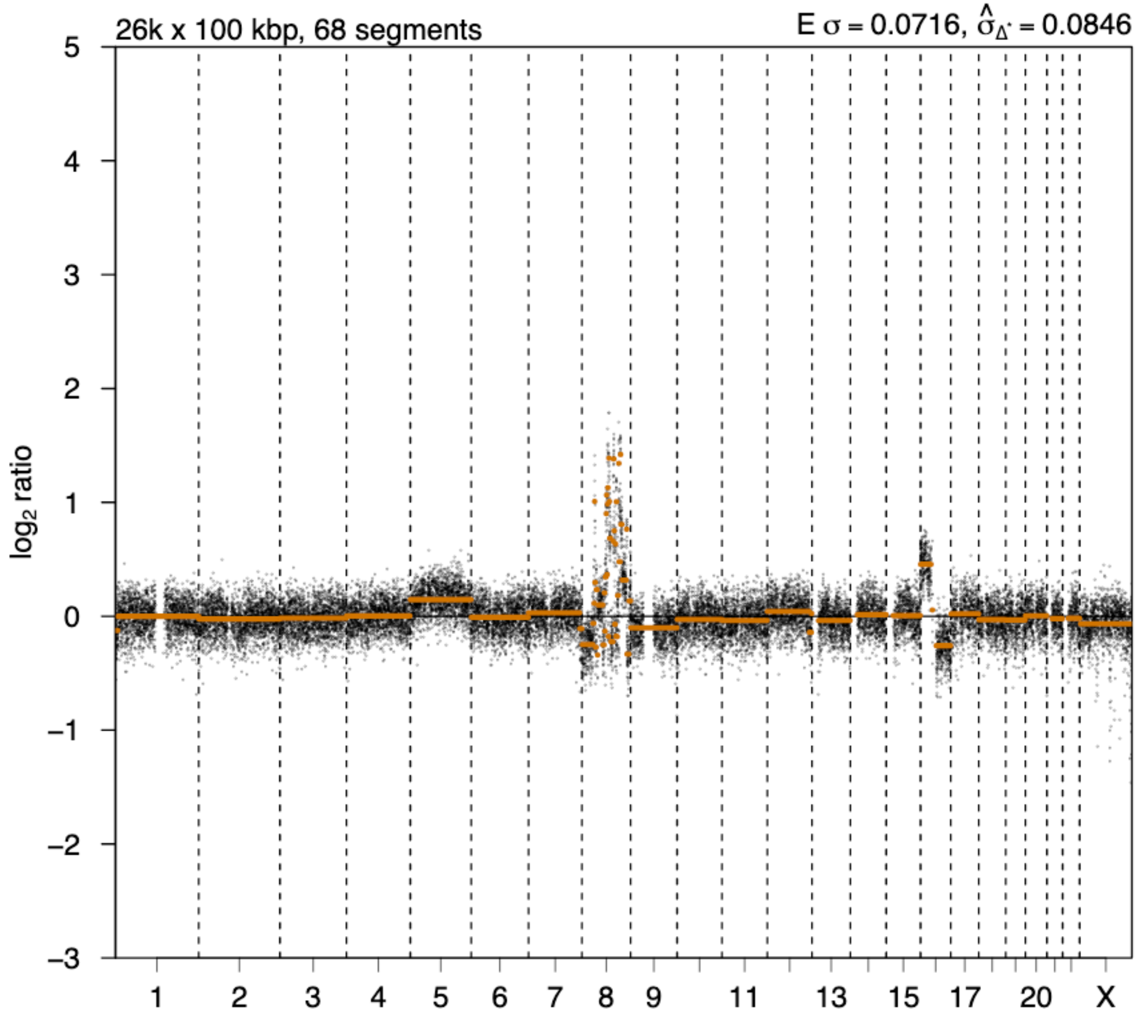

# P019

| Syn/Meta | Time from 1st surgery to 2nd event (Months) | Side 2nd event | Histology 2nd event | Surgery | AdjuvanT Treatment Pri (RT/ HT) | ER Pri | ER 2nd event | Her2 Pri | Her2 2nd event | Grade Pri | Grade 2nd event | Quadrant 2nd event | Margins | Screening | Clonality P value Copy N | Clonality P value Panel seq | Clonality P value WES | Final verdict |
|---|---|---|---|---|---|---|---|---|---|---|---|---|---|---|---|---|---|---|
| metachronous | 18 | Ipsilateral | DCIS only | lumpectomy | None | - | - | NA | NA | 3 | 3 | NA | Involved | screen-detected | 0.003663 004 | NA | NA | Related |

### Primary event

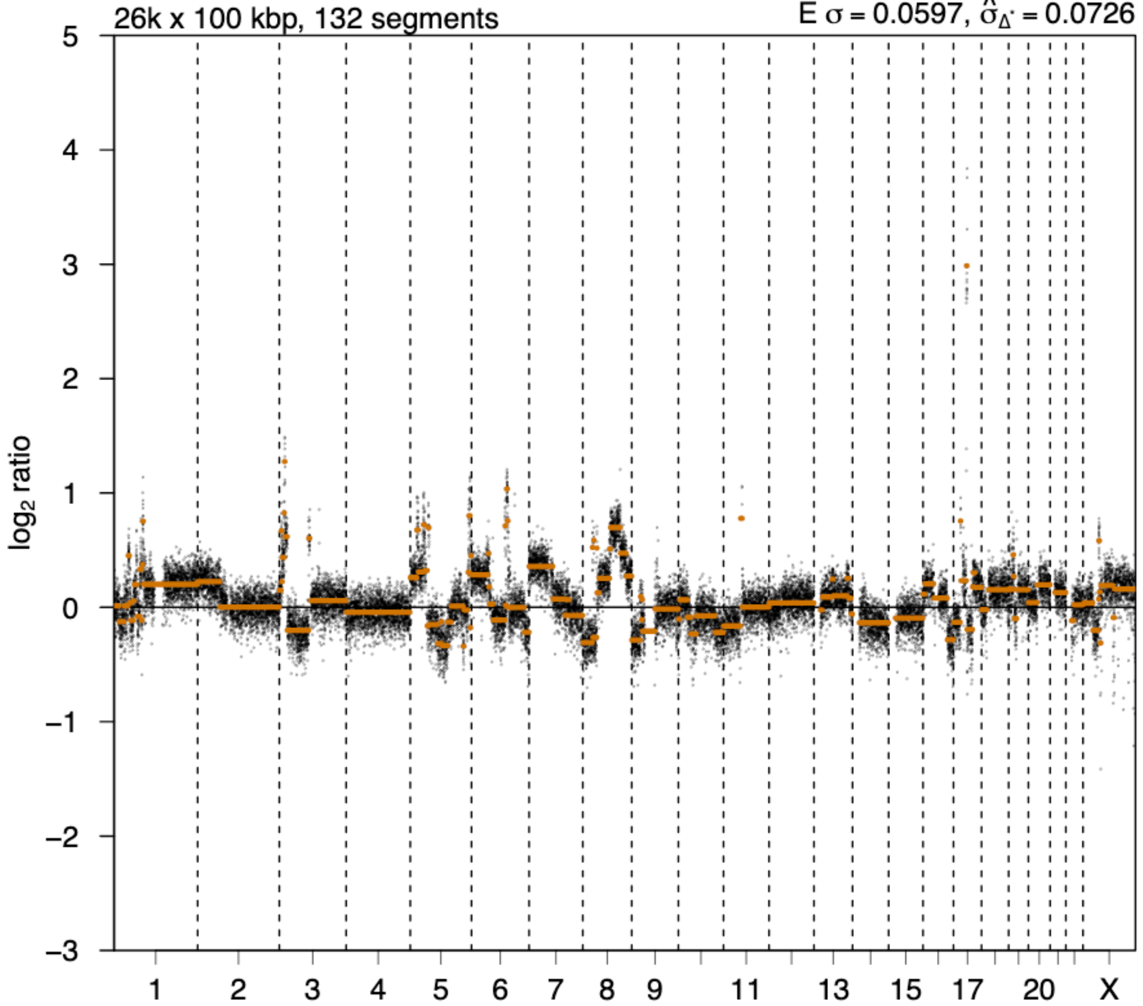


### 2nd event

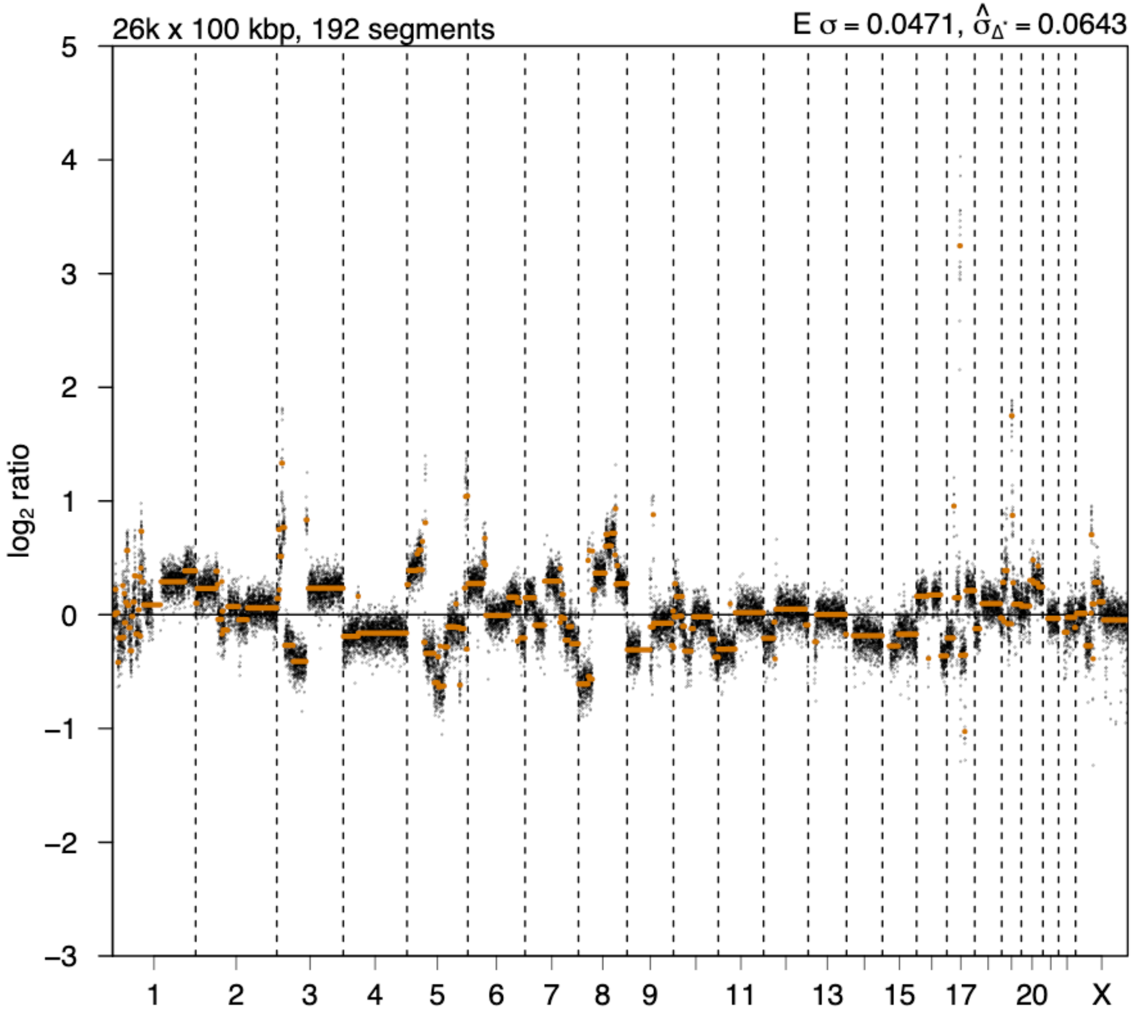

# P020

| Syn/Meta | Time from 1st surgery to 2nd event (Months) | Side 2nd event | Histology 2nd event | Surgery | AdjuvanT Treatment Pri (RT/ HT) | ER Pri | ER 2nd event | Her2 Pri | Her2 2nd event | Grade Pri | Grade 2nd event | Quadrant 2nd event | Margins | Screening | Clonality P value Copy N | Clonality P value Panel seq | Clonality P value WES | Final verdict |
|---|---|---|---|---|---|---|---|---|---|---|---|---|---|---|---|---|---|---|
| metachronous | 17 | Ipsilateral | DCIS only | lumpectomy | None | - | NA | NA | NA | 3 | 3 | NA | Clear | screen-detected | 0.003663004 | NA | NA | Related |

### Primary event

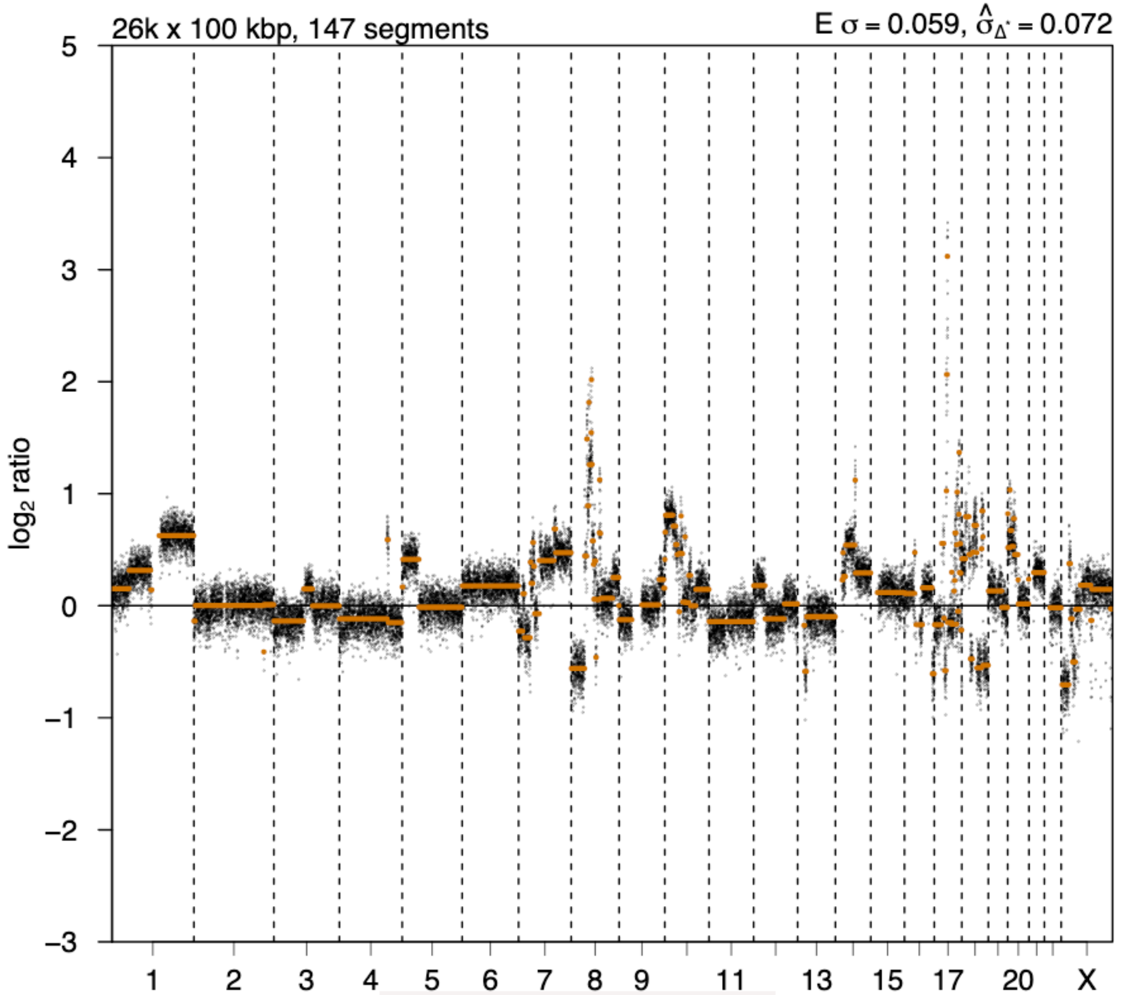


### 2nd event

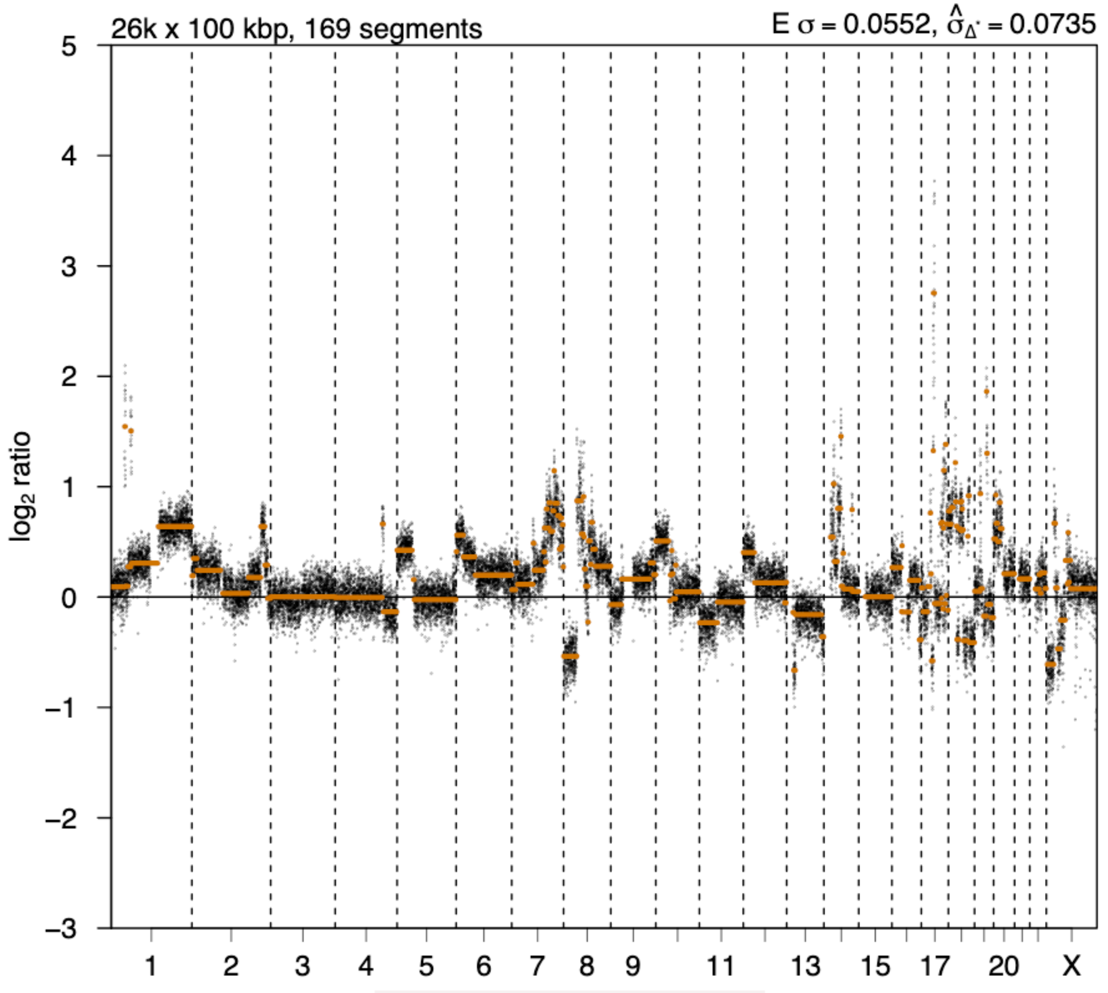

# P021

| Syn/Meta | Time from 1st surgery to 2nd event (Months) | Side 2nd event | Histology 2nd event | Surgery | AdjuvanT Treatment Pri (RT/ HT) | ER Pri | ER 2nd event | Her2 Pri | Her2 2nd event | Grade Pri | Grade 2nd event | Quadrant 2nd event | Margins | Screening | Clonality P value Copy N | Clonality P value Panel seq | Clonality P value WES | Final verdict |
|---|---|---|---|---|---|---|---|---|---|---|---|---|---|---|---|---|---|---|
| metachronous | 18 | Ipsilateral | DCIS only | lumpectomy | None | - | - | NA | NA | 3 | 3 | NA | Clear | screen-detected | 0.003663004 | NA | NA | Related |

### Primary event

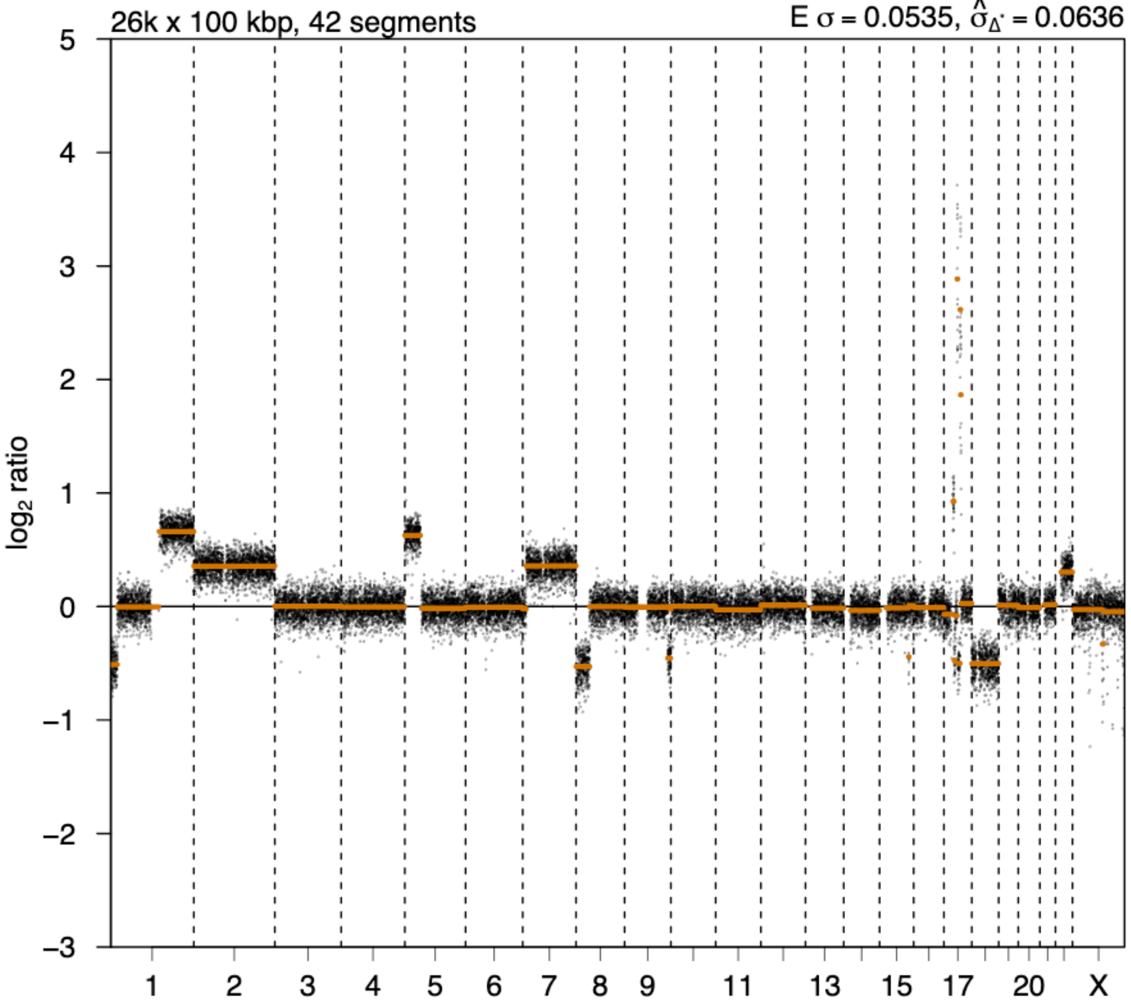


### 2nd event

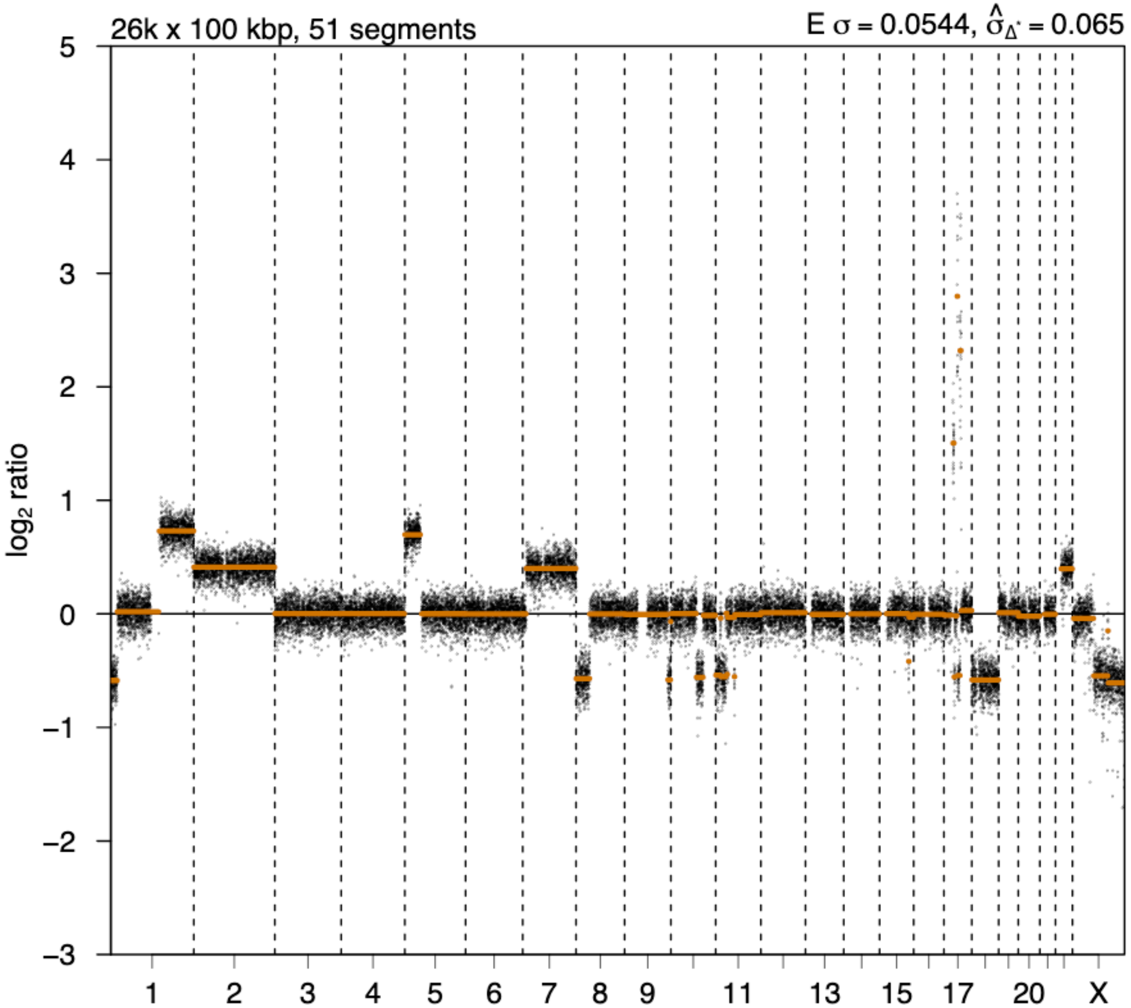

# P023

| Syn/Meta | Time from 1st surgery to 2nd event (Months) | Side 2nd event | Histology 2nd event | Surgery | AdjuvanT Treatment Pri (RT/ HT) | ER Pri | ER 2nd event | Her2 Pri | Her2 2nd event | Grade Pri | Grade 2nd event | Quadrant 2nd event | Margins | Screening | Clonality P value Copy N | Clonality P value Panel seq | Clonality P value WES | Final verdict |
|---|---|---|---|---|---|---|---|---|---|---|---|---|---|---|---|---|---|---|
| metachronous | 25 | Ipsilateral | DCIS only | lumpectomy | RT | NA | NA | - | NA | 3 | 3 | NA | Clear | screen-detected | 0.003663004 | NA | NA | Related |

### Primary event

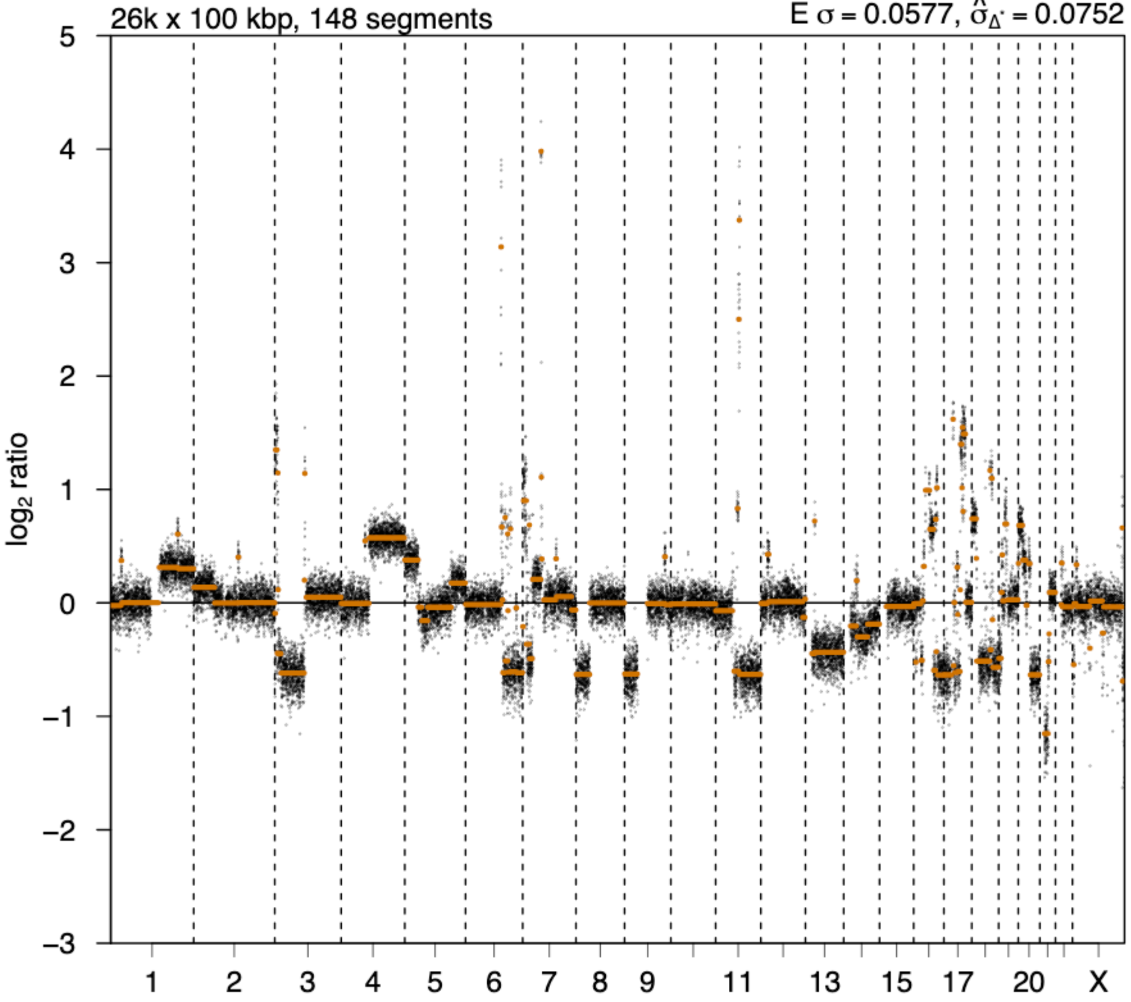


### 2nd event

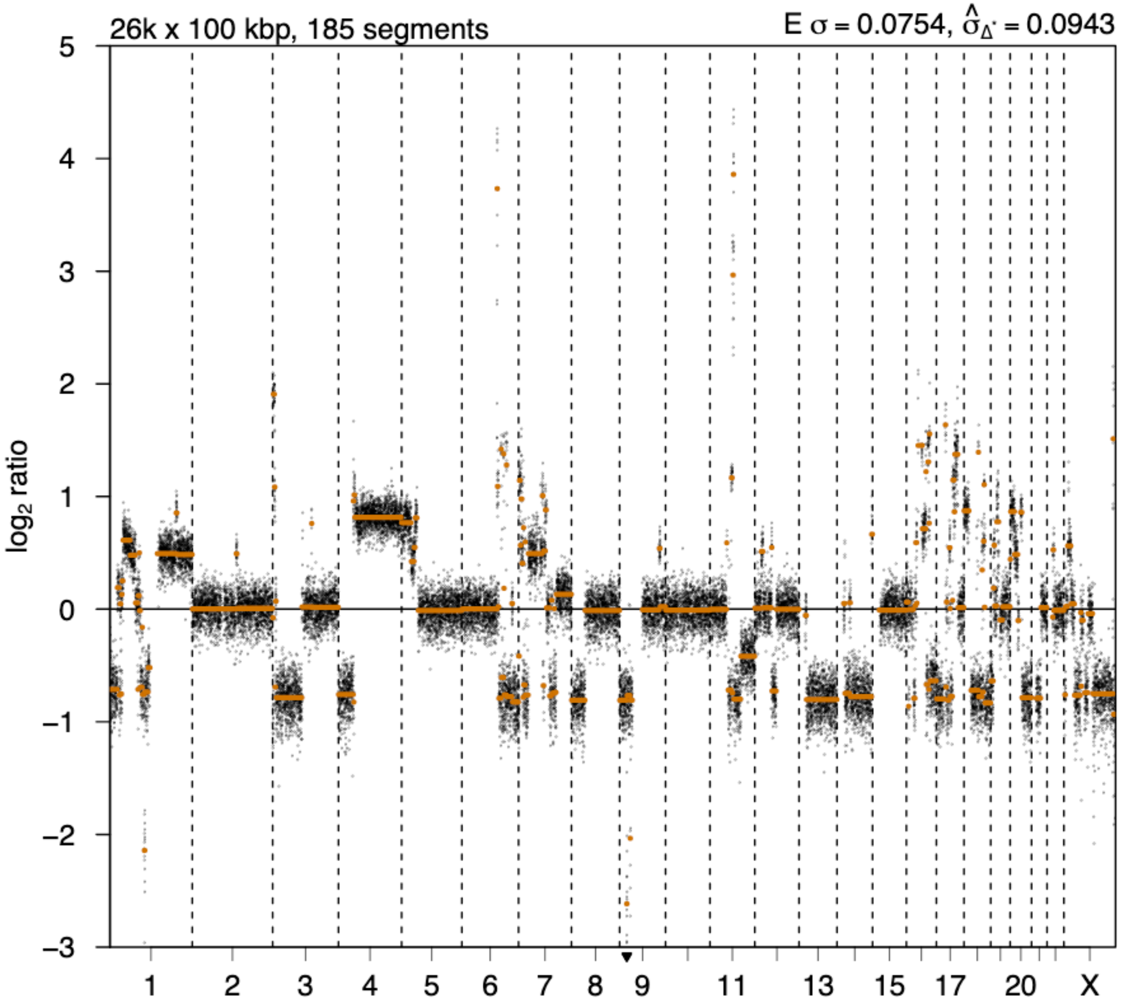

# P024

| Syn/Meta | Time from 1st surgery to 2nd event (Months) | Side 2nd event | Histology 2nd event | Surgery | AdjuvanT Treatment Pri (RT/ HT) | ER Pri | ER 2nd event | Her2 Pri | Her2 2nd event | Grade Pri | Grade 2nd event | Quadrant 2nd event | Margins | Screening | Clonality P value Copy N | Clonality P value Panel seq | Clonality P value WES | Final verdict |
|---|---|---|---|---|---|---|---|---|---|---|---|---|---|---|---|---|---|---|
| metachronous | 27 | Ipsilateral | DCIS only | lumpectomy | None | + | NA | - | NA | 2 | 3 | at or adjacent to primary | Clear | screen-detected | 0.000466 | NA | NA | Related |

### Primary event

LogR

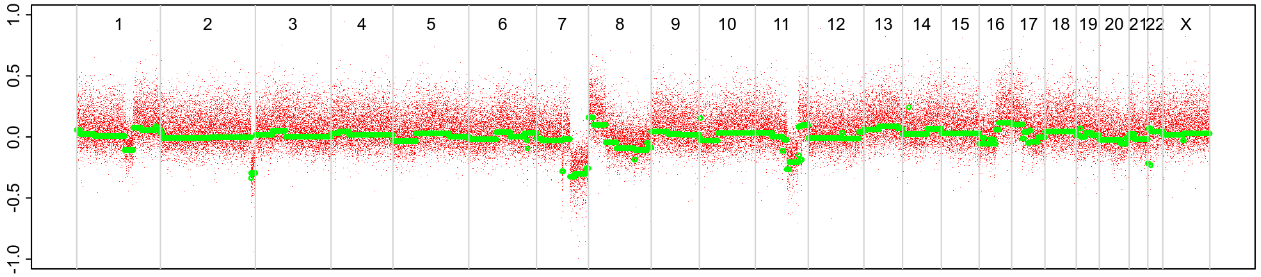


BAF

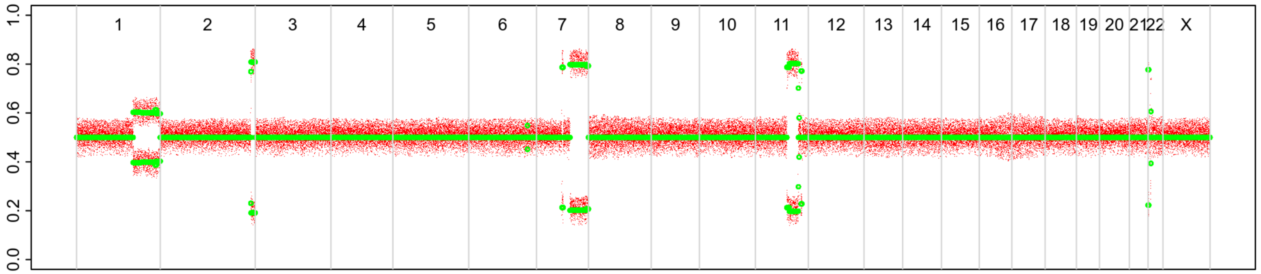


### 2nd event

LogR

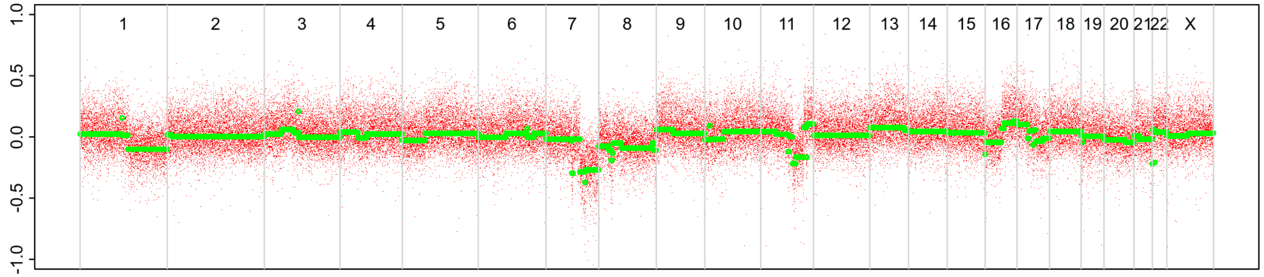


BAF

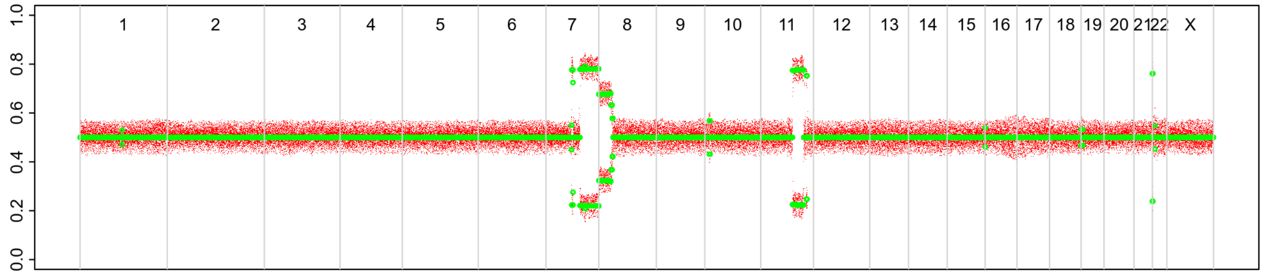

# P026

| Syn/Meta | Time from 1st surgery to 2nd event (Months) | Side 2nd event | Histology 2nd event | Surgery | AdjuvanT Treatment Pri (RT/ HT) | ER Pri | ER 2nd event | Her2 Pri | Her2 2nd event | Grade Pri | Grade 2nd event | Quadrant 2nd event | Margins | Screening | Clonality P value Copy N | Clonality P value Panel seq | Clonality P value WES | Final verdict |
|---|---|---|---|---|---|---|---|---|---|---|---|---|---|---|---|---|---|---|
| metachronous | 26 | Ipsilateral | DCIS only | lumpectomy | RT | + | + | + | + | 3 | 3 | at or adjacent to primary | Clear | screen-detected | 0.000466 | NA | NA | Related |

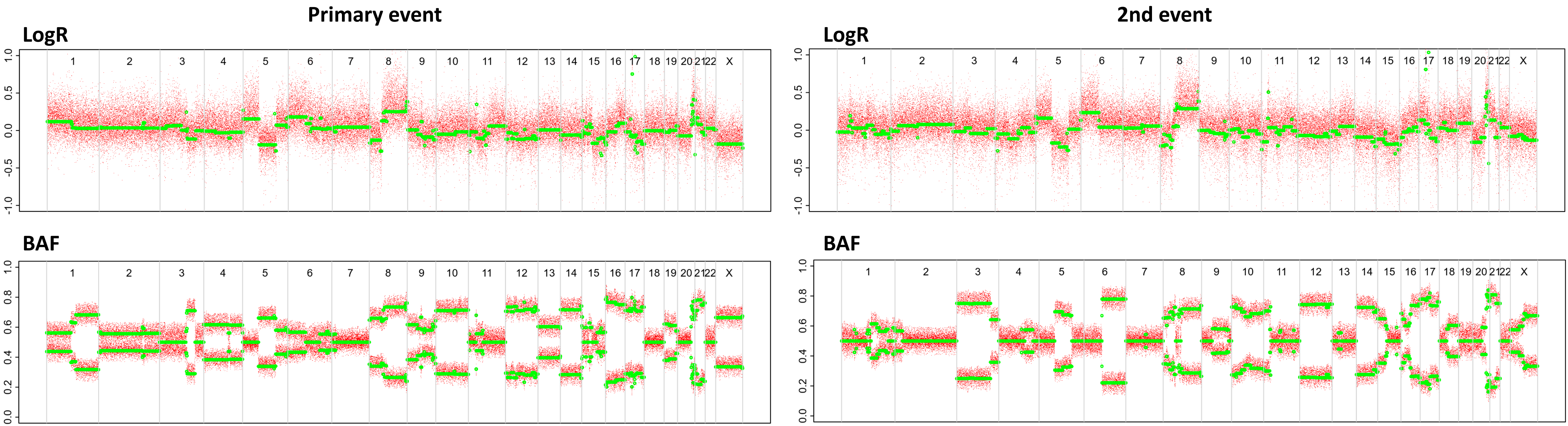

# P027

| Syn/Meta | Time from 1st surgery to 2nd event (Months) | Side 2nd event | Histology 2nd event | Surgery | AdjuvanT Treatment Pri (RT/ HT) | ER Pri | ER 2nd event | Her2 Pri | Her2 2nd event | Grade Pri | Grade 2nd event | Quadrant 2nd event | Margins | Screening | Clonality P value Copy N | Clonality P value Panel seq | Clonality P value WES | Final verdict |
|---|---|---|---|---|---|---|---|---|---|---|---|---|---|---|---|---|---|---|
| metachronous | 15 | Ipsilateral | DCIS only | lumpectomy | RT | - | NA | + | + | 3 | 3 | at or adjacent to primary | Clear | screen-detected | 0.000466 | 0 | NA | Related |

### Primary event

LogR

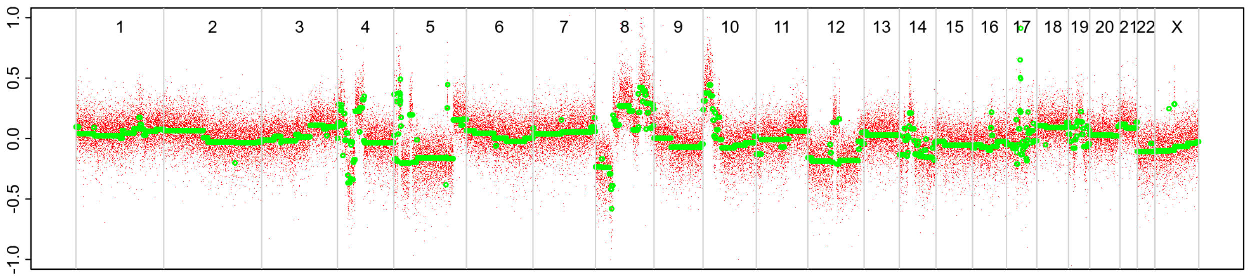


BAF

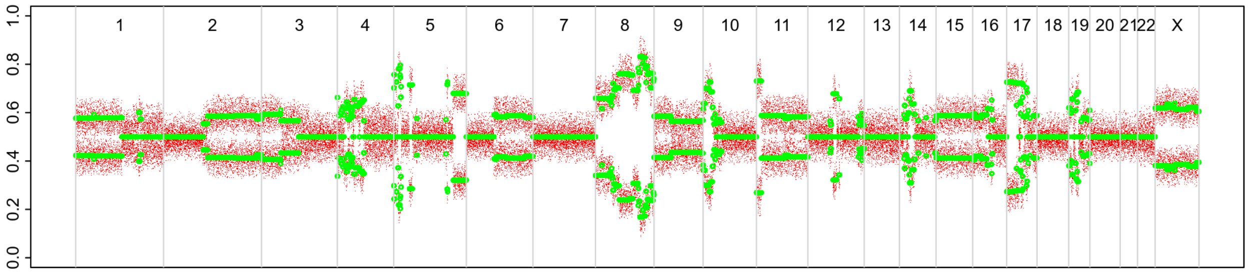


### 2nd event

LogR

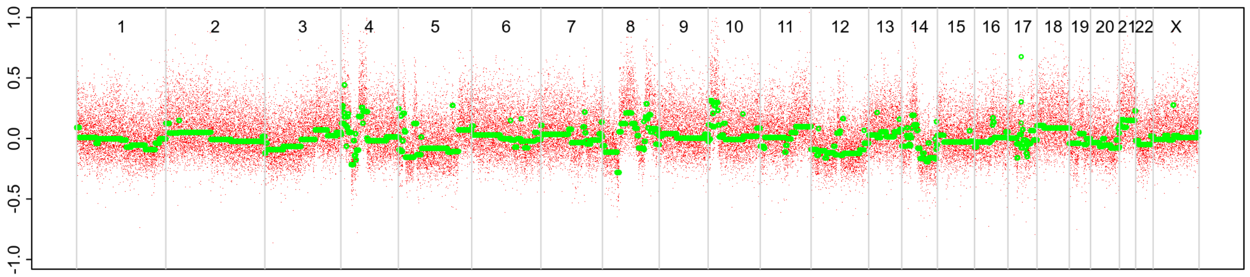


BAF

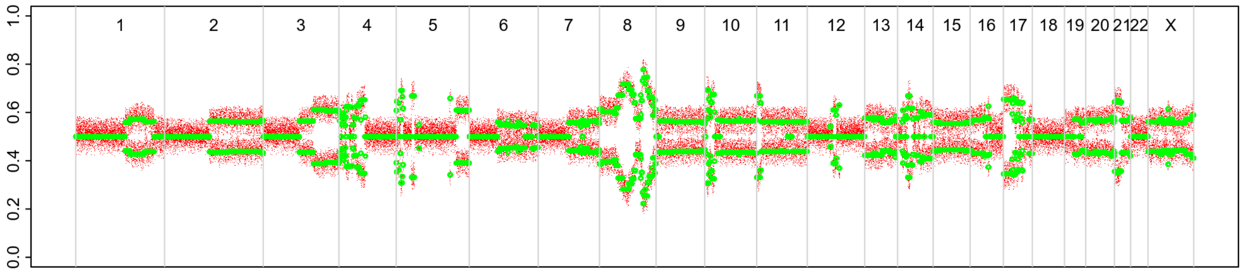

# P028

| Syn/Meta | Time from 1st surgery to 2nd event (Months) | Side 2nd event | Histology 2nd event | Surgery | AdjuvanT Treatment Pri (RT/ HT) | ER Pri | ER 2nd event | Her2 Pri | Her2 2nd event | Grade Pri | Grade 2nd event | Quadrant 2nd event | Margins | Screening | Clonality P value Copy N | Clonality P value Panel seq | Clonality P value WES | Final verdict |
|---|---|---|---|---|---|---|---|---|---|---|---|---|---|---|---|---|---|---|
| metachronous | 16 | Ipsilateral | DCIS only | lumpectomy | RT | - | NA | + | NA | 3 | 3 | at or adjacent to primary | Clear | screen-detected | 0.000466 | NA | NA | Related |

### Primary event

**LogR**

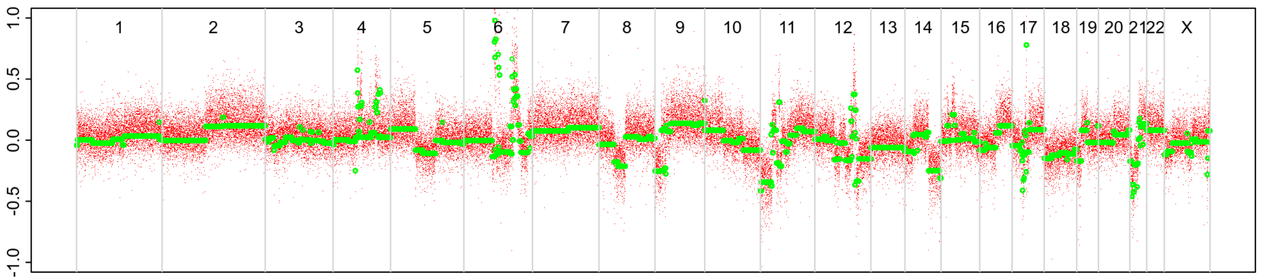


**BAF**

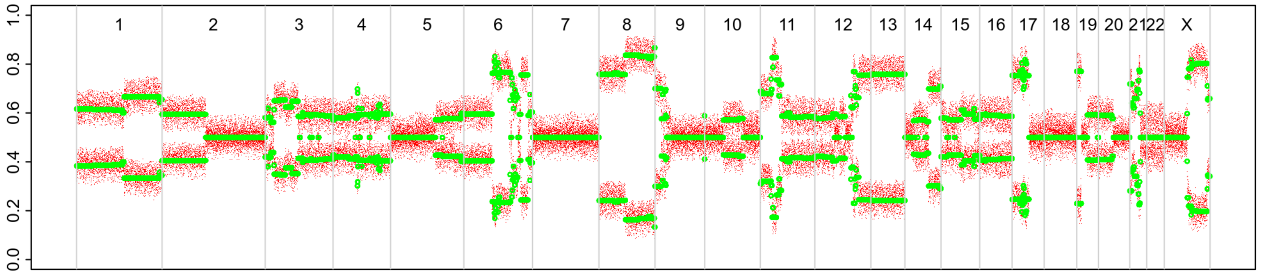


### 2nd event

**LogR**

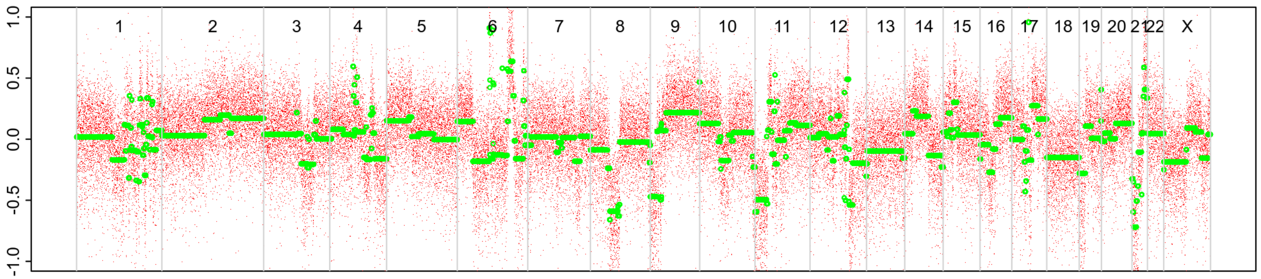


**BAF**

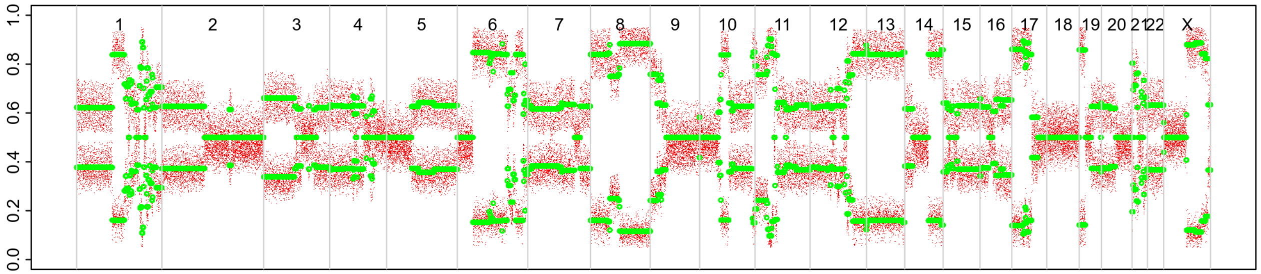

# P029

| Syn/Meta | Time from 1st surgery to 2nd event (Months) | Side 2nd event | Histology 2nd event | Surgery | AdjuvanT Treatment Pri (RT/ HT) | ER Pri | ER 2nd event | Her2 Pri | Her2 2nd event | Grade Pri | Grade 2nd event | Quadrant 2nd event | Margins | Screening | Clonality P value Copy N | Clonality P value Panel seq | Clonality P value WES | Final verdict |
|---|---|---|---|---|---|---|---|---|---|---|---|---|---|---|---|---|---|---|
| metachronous | 26 | Ipsilateral | DCIS only | lumpectomy | None | - | NA | NA | NA | 3 | 3 | at or adjacent to primary | Clear | screen-detected | 0.000466 | 0 | NA | Related |

### Primary event

LogR

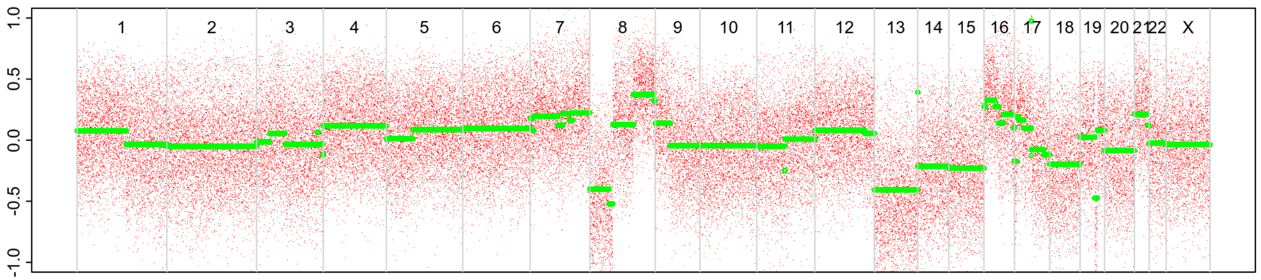


BAF

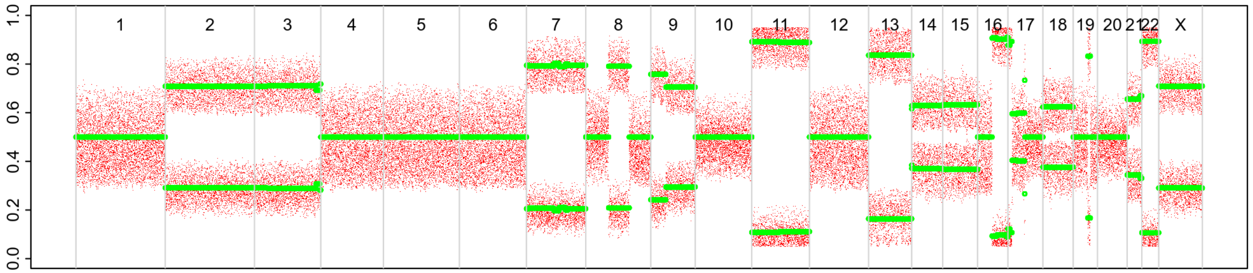


### 2nd event

LogR

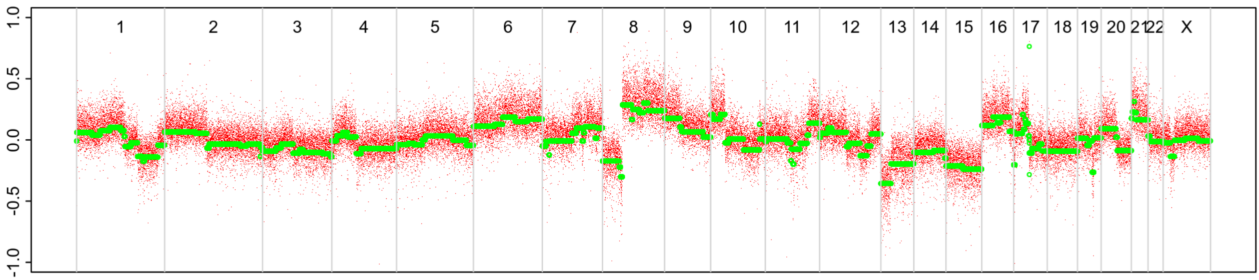


BAF

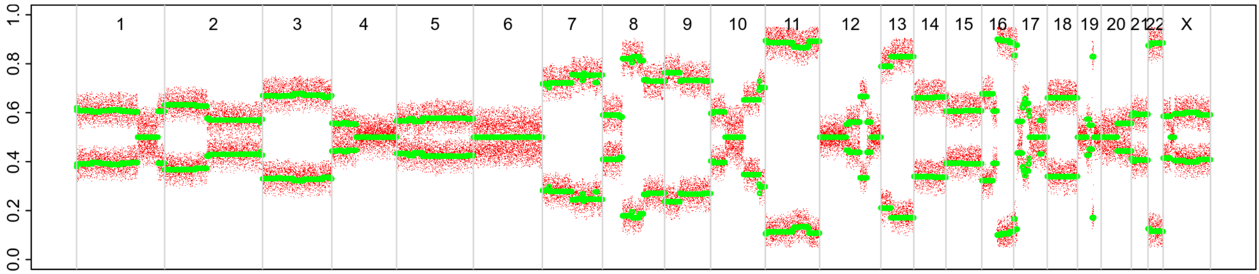

# P058

| Syn/Meta | Time from 1st surgery to 2nd event (Months) | Side 2nd event | Histology 2nd event | Surgery | AdjuvanT Treatment Pri (RT/ HT) | ER Pri | ER 2nd event | Her2 Pri | Her2 2nd event | Grade Pri | Grade 2nd event | Quadrant 2nd event | Margins | Screening | Clonality P value Copy N | Clonality P value Panel seq | Clonality P value WES | Final verdict |
|---|---|---|---|---|---|---|---|---|---|---|---|---|---|---|---|---|---|---|
| metachronous | 62 | Ipsilateral | IDC no DCIS | lumpectomy | RTHT | + | NA | - | NA | 3 | 3 | at or adjacent to primary | Clear | screen-detected | 0.000466 | 0 | NA | Related |

### Primary event

LogR

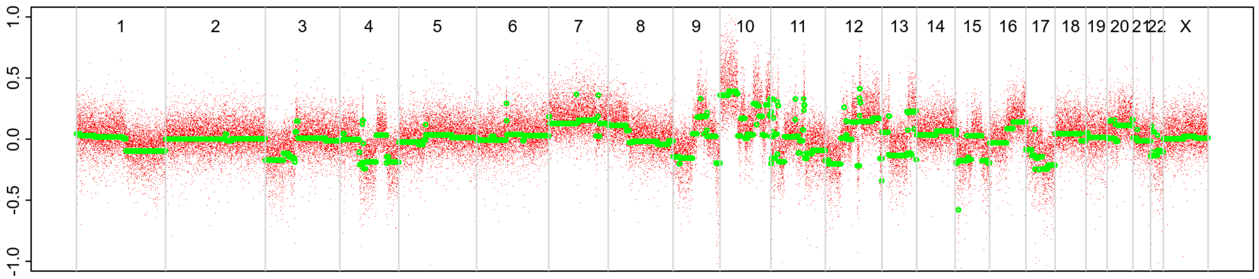


BAF

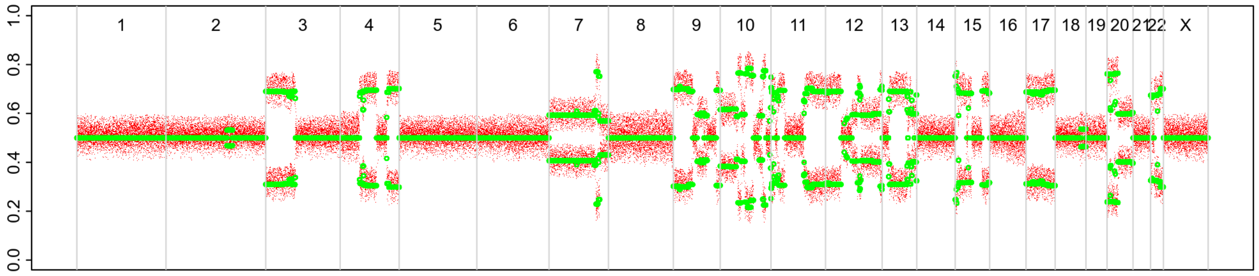


### 2nd event

LogR

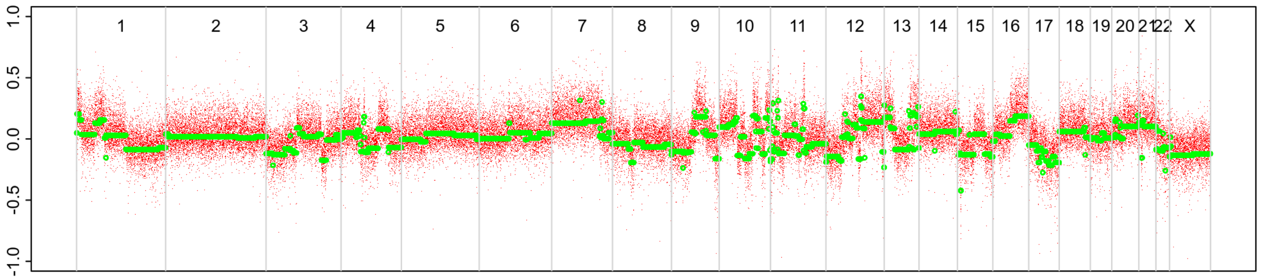


BAF

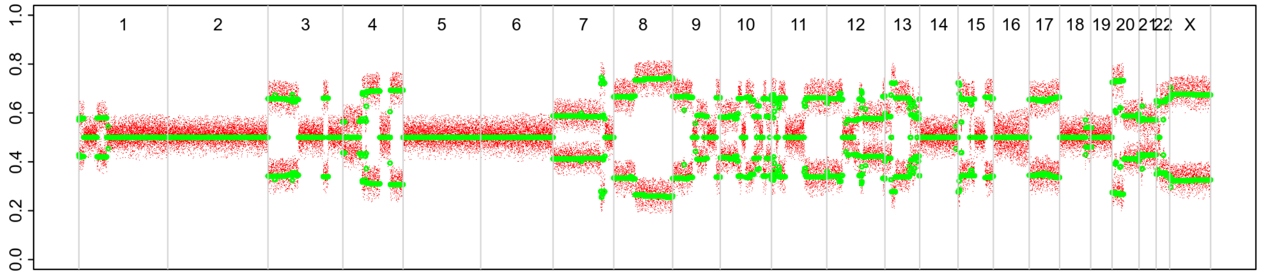

# P064

| Syn/Meta | Time from 1st surgery to 2nd event (Months) | Side 2nd event | Histology 2nd event | Surgery | AdjuvanT Treatment Pri (RT/ HT) | ER Pri | ER 2nd event | Her2 Pri | Her2 2nd event | Grade Pri | Grade 2nd event | Quadrant 2nd event | Margins | Screening | Clonality P value Copy N | Clonality P value Panel seq | Clonality P value WES | Final verdict |
|---|---|---|---|---|---|---|---|---|---|---|---|---|---|---|---|---|---|---|
| metachronous | 54 | Ipsilateral | IDC with DCIS | lumpectomy | None | + | + | - | NA | 3 | 2 | NA | Clear | screen-detected | 0.000466 | 0 | NA | Related |

### Primary event

LogR

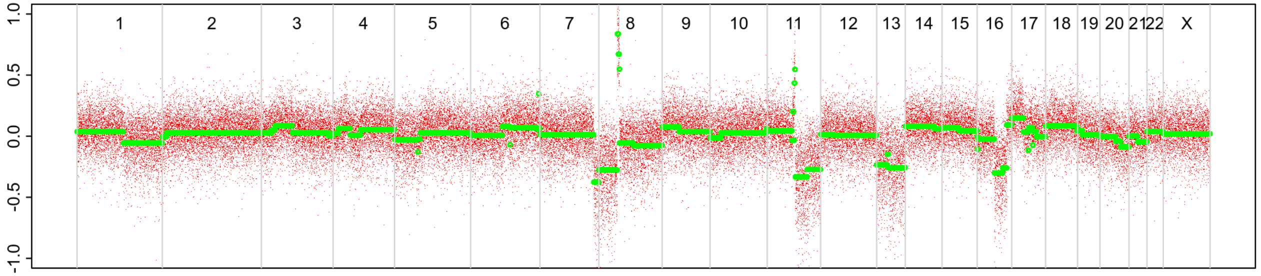


BAF

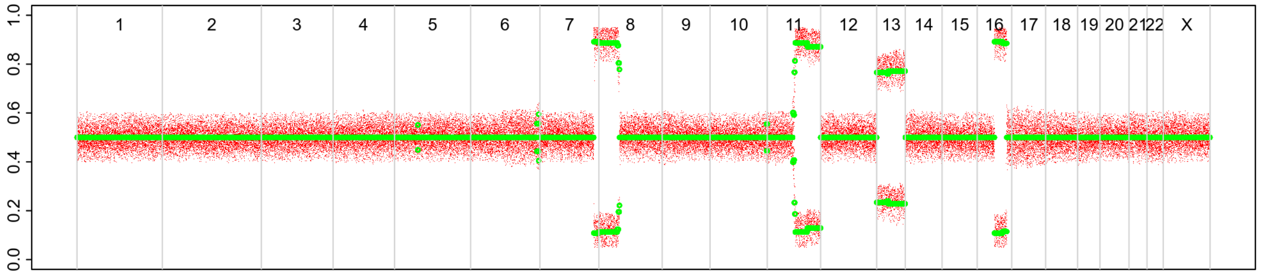


### 2nd event

LogR

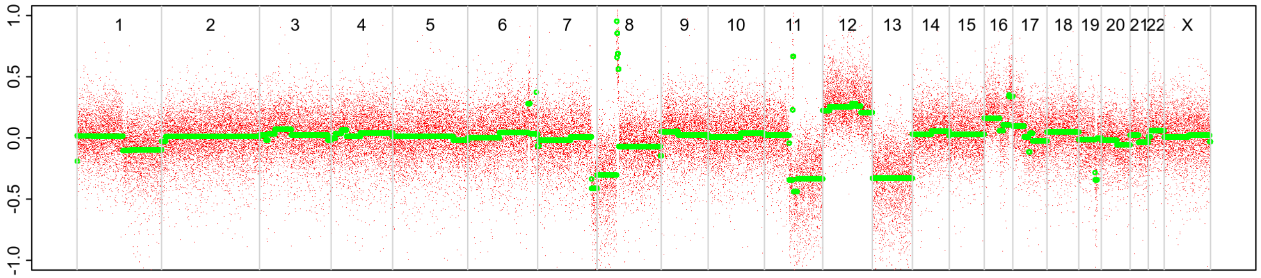


BAF

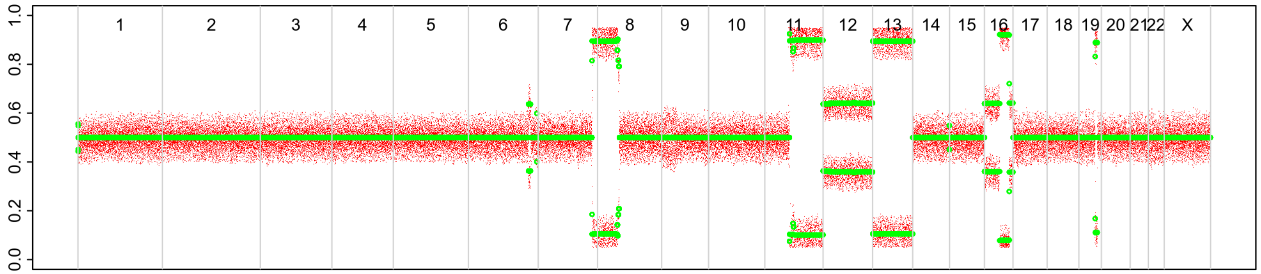

# P070

| Syn/Meta | Time from 1st surgery to 2nd event (Months) | Side 2nd event | Histology 2nd event | Surgery | AdjuvanT Treatment Pri (RT/ HT) | ER Pri | ER 2nd event | Her2 Pri | Her2 2nd event | Grade Pri | Grade 2nd event | Quadrant 2nd event | Margins | Screening | Clonality P value Copy N | Clonality P value Panel seq | Clonality P value WES | Final verdict |
|---|---|---|---|---|---|---|---|---|---|---|---|---|---|---|---|---|---|---|
| metachronous | 35 | Ipsilateral | IDC no DCIS | lumpectomy | None | + | + | + | - | 3 | 2 | at or adjacent to primary | Clear | screen-detected | 0.000466 | 0 | NA | Related |

### Primary event

LogR

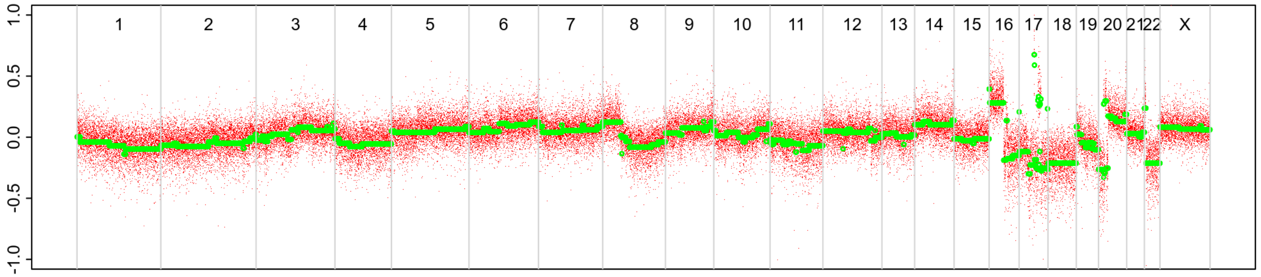


BAF

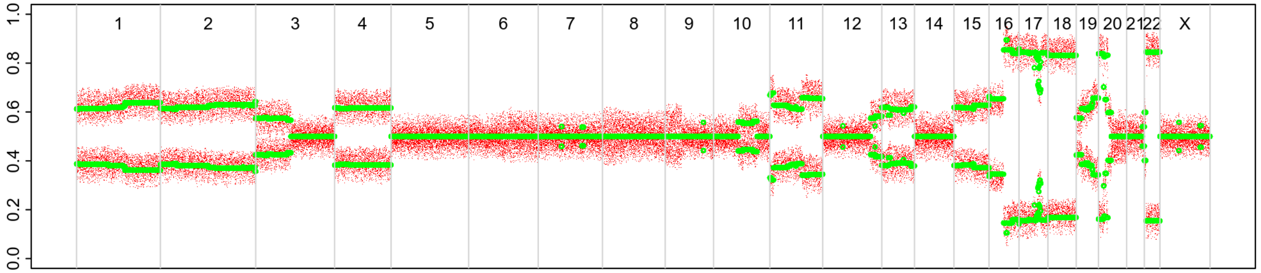


### 2nd event

LogR

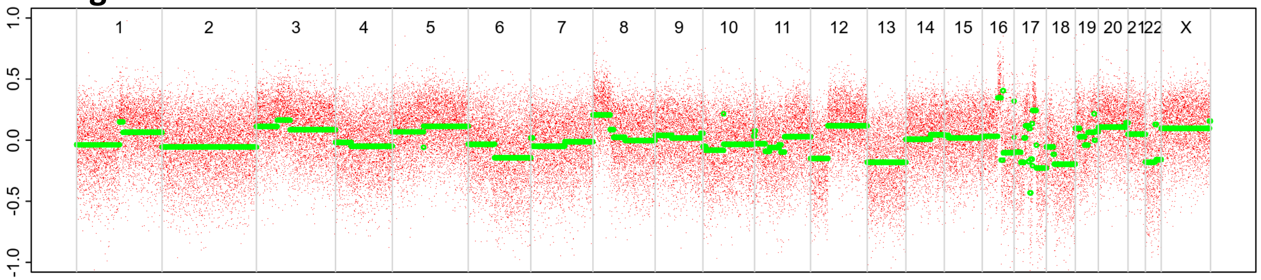


BAF

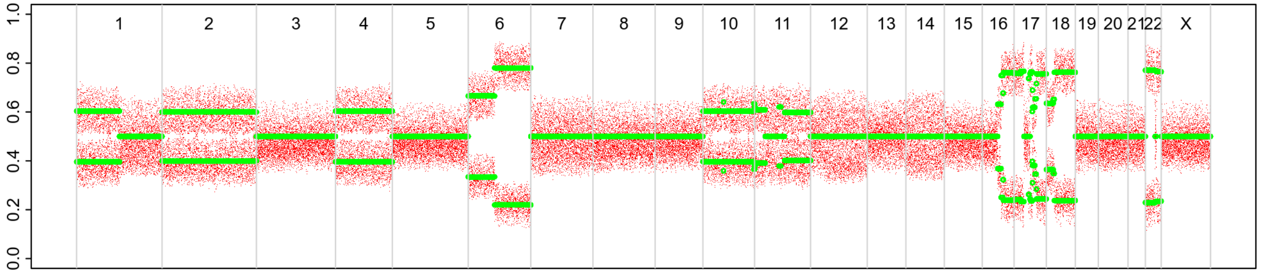

# P071

| Syn/Meta | Time from 1st surgery to 2nd event (Months) | Side 2nd event | Histology 2nd event | Surgery | AdjuvanT Treatment Pri (RT/ HT) | ER Pri | ER 2nd event | Her2 Pri | Her2 2nd event | Grade Pri | Grade 2nd event | Quadrant 2nd event | Margins | Screening | Clonality P value Copy N | Clonality P value Panel seq | Clonality P value WES | Final verdict |
|---|---|---|---|---|---|---|---|---|---|---|---|---|---|---|---|---|---|---|
| metachronous | 52 | Ipsilateral | IDC with DCIS* | lumpectomy | RT | + | - | + | NA | 3 | 3 | at or adjacent to primary | Clear | screen-detected | 0.000932 | NA | NA | Related |

| Clonality P value for ipsilateral synchronous 2nd event Copy Number |
|---|
| 0.004662 |

### Primary event

LogR

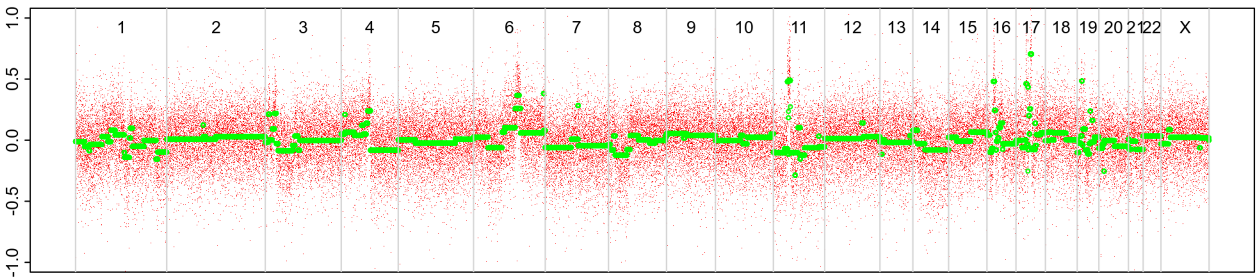


BAF

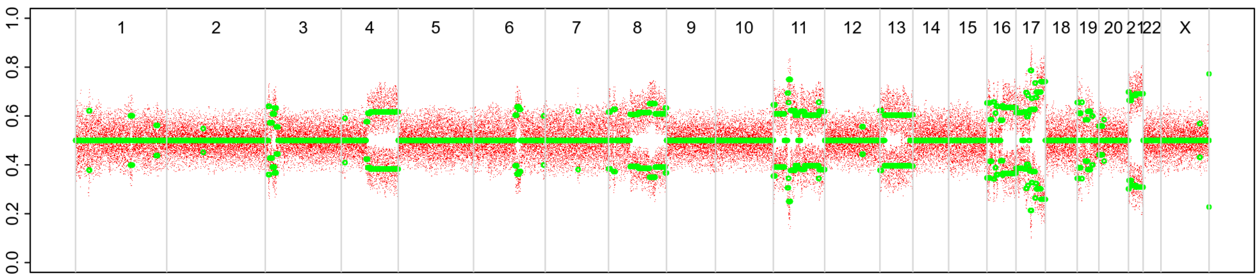


### 2nd event (INV)

LogR

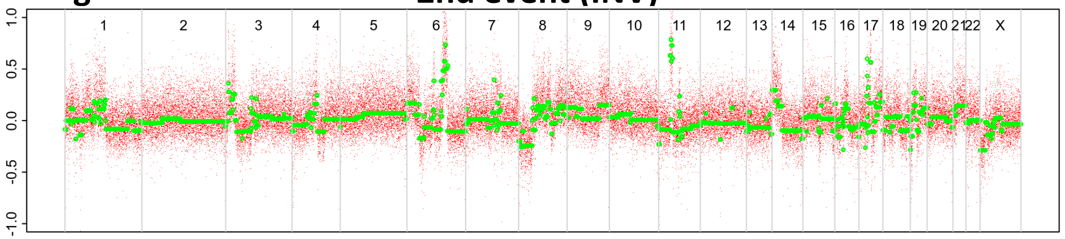


BAF

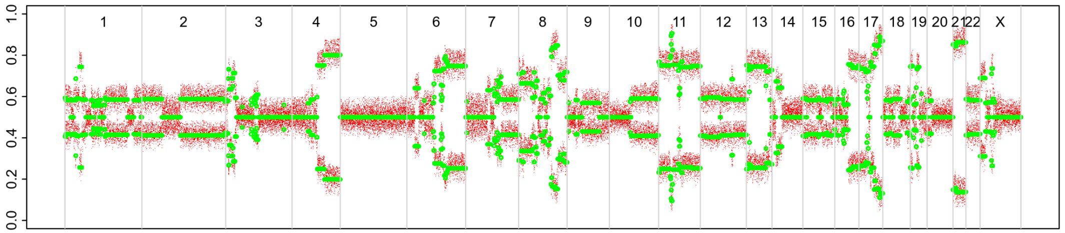


### 2nd event (DCIS)

LogR

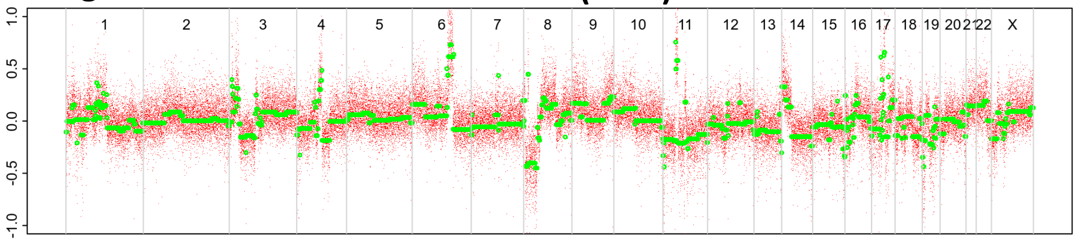


BAF

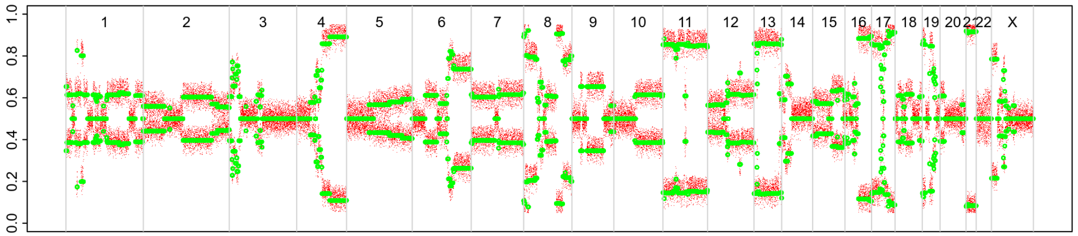

# P074

| Syn/Meta | Time from 1st surgery to 2nd event (Months) | Side 2nd event | Histology 2nd event | Surgery | AdjuvanT Treatment Pri (RT/ HT) | ER Pri | ER 2nd event | Her2 Pri | Her2 2nd event | Grade Pri | Grade 2nd event | Quadrant 2nd event | Margins | Screening | Clonality P value Copy N | Clonality P value Panel seq | Clonality P value WES | Final verdict |
|---|---|---|---|---|---|---|---|---|---|---|---|---|---|---|---|---|---|---|
| metachronous | 53 | Ipsilateral | IDC no DCIS | lumpectomy | None | - | NA | + | + | 3 | 3 | at or adjacent to primary | Clear | screen-detected | 0.000466 | 0.0010090 82 | NA | Related |

### Primary event

LogR

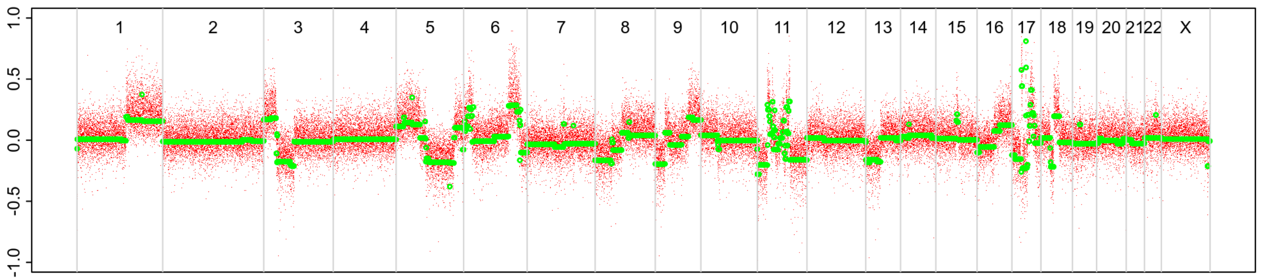


BAF

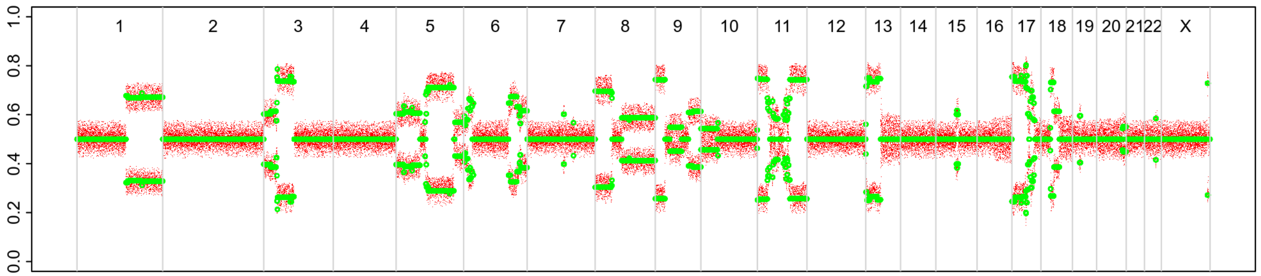


### 2nd event

LogR

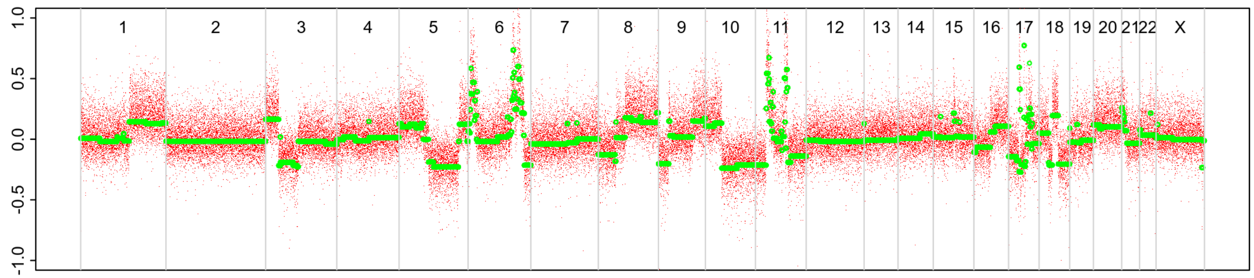


BAF

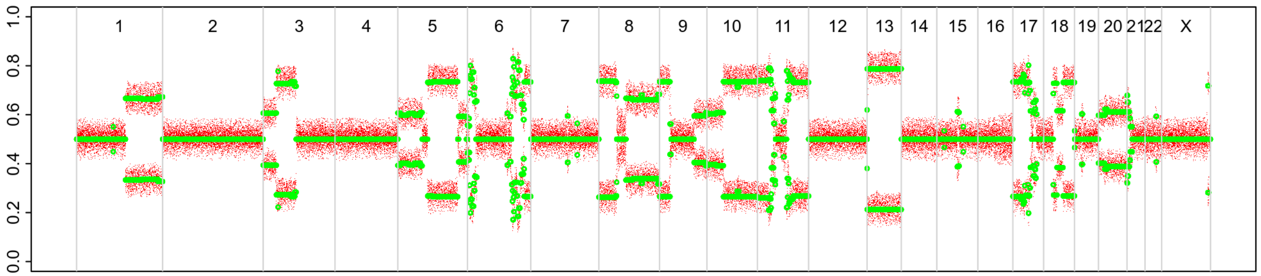

# P075

| Syn/Meta | Time from 1st surgery to 2nd event (Months) | Side 2nd event | Histology 2nd event | Surgery | AdjuvanT Treatment Pri (RT/ HT) | ER Pri | ER 2nd event | Her2 Pri | Her2 2nd event | Grade Pri | Grade 2nd event | Quadrant 2nd event | Margins | Screening | Clonality P value Copy N | Clonality P value Panel seq | Clonality P value WES | Final verdict |
|---|---|---|---|---|---|---|---|---|---|---|---|---|---|---|---|---|---|---|
| metachronous | 31 | Ipsilateral | IDC with DCIS* | lumpectomy | None | + | NA | NA | NA | 3 | 2 | at or adjacent to primary | Clear | screen-detected | 0.0004662 | NA | NA | Related |

| Clonality P value for ipsilateral synchronous 2nd event Copy Number |
|---|
| 0.004662 |

### Primary event

LogR

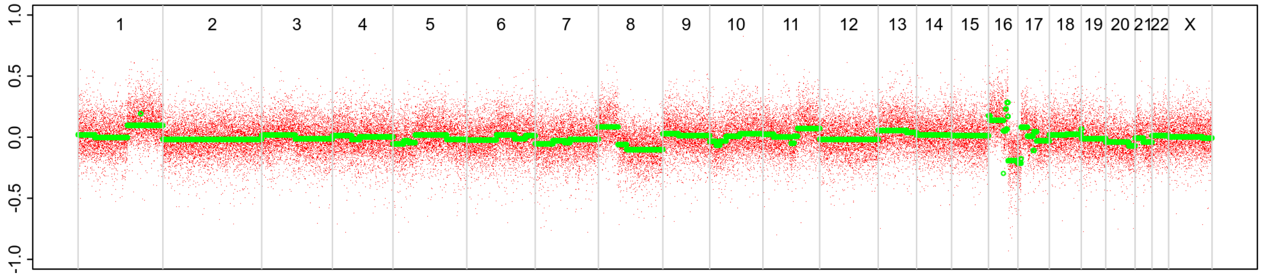


BAF

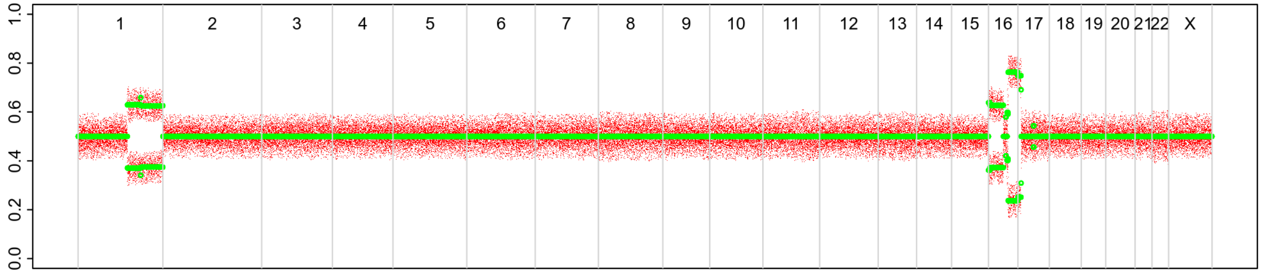


### 2nd event (INV)

LogR

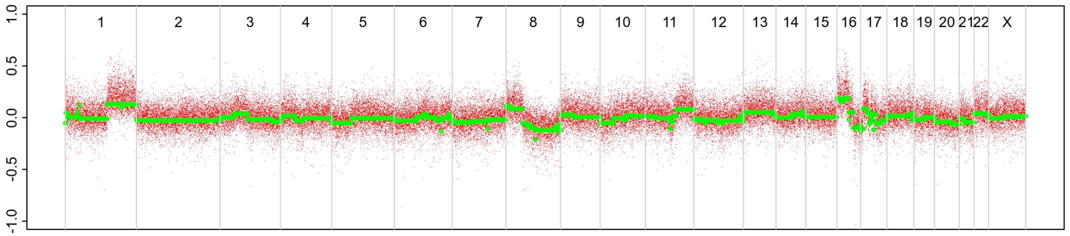


BAF

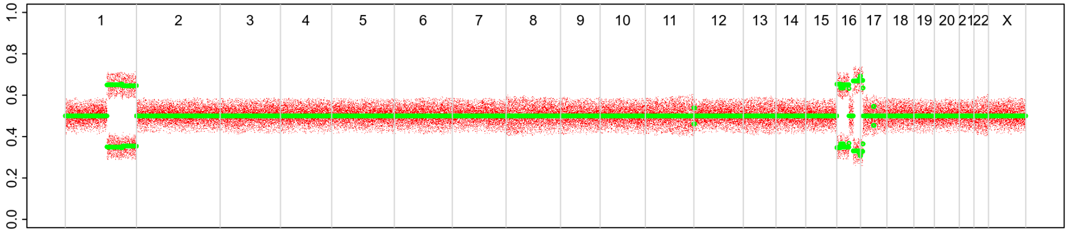


### 2nd event (DCIS)

LogR

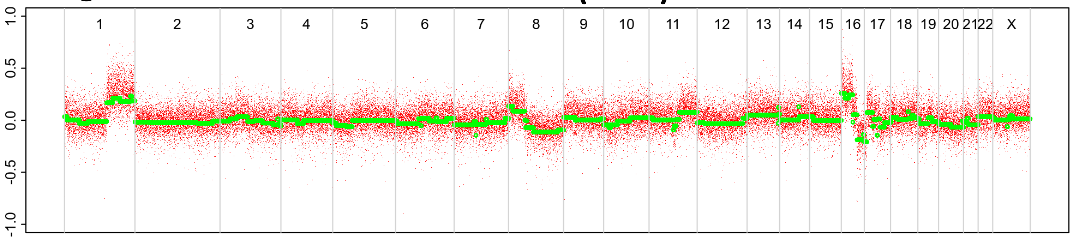


BAF

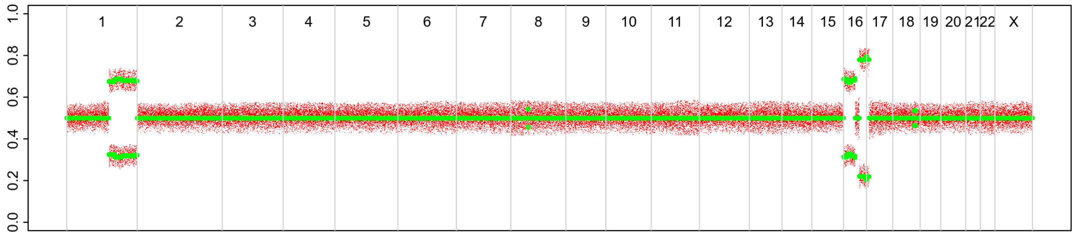

# P076

| Syn/Meta | Time from 1st surgery to 2nd event (Months) | Side 2nd event | Histology 2nd event | Surgery | AdjuvanT Treatment Pri (RT/ HT) | ER Pri | ER 2nd event | Her2 Pri | Her2 2nd event | Grade Pri | Grade 2nd event | Quadrant 2nd event | Margins | Screening | Clonality P value Copy N | Clonality P value Panel seq | Clonality P value WES | Final verdict |
|---|---|---|---|---|---|---|---|---|---|---|---|---|---|---|---|---|---|---|
| metachronous | 49 | Ipsilateral | IDC with DCIS | lumpectomy | None | + | + | - | NA | 3 | 2 | at or adjacent to primary | Clear | screen-detected | 0.000466 2 | 0 | NA | Related |

### Primary event

LogR

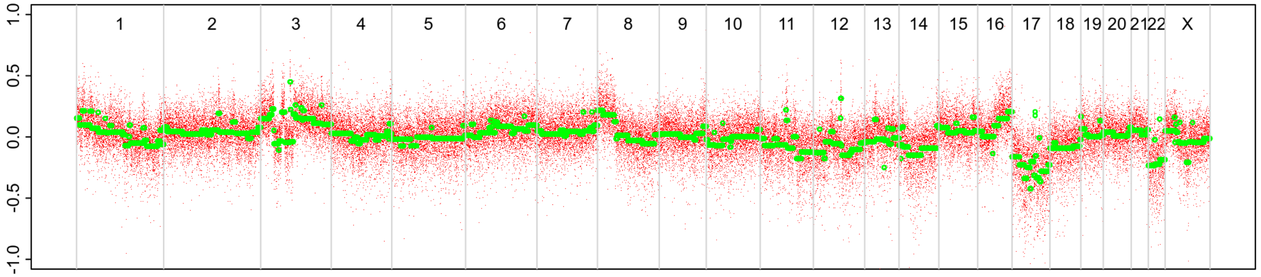


BAF

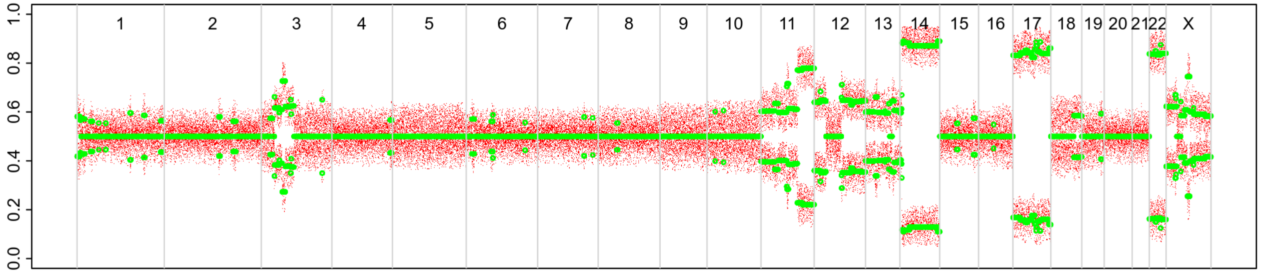


### 2nd event

LogR

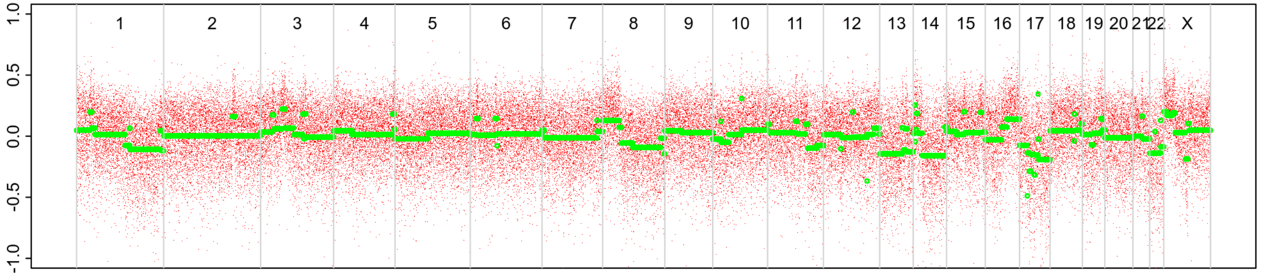


BAF

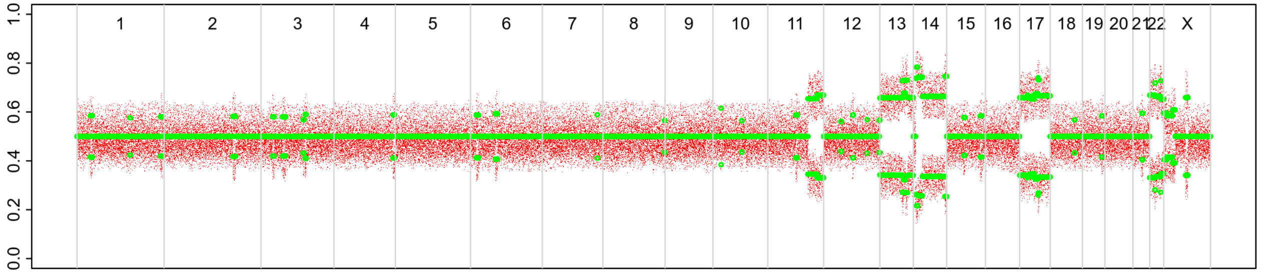

# P077

| Syn/Meta | Time from 1st surgery to 2nd event (Months) | Side 2nd event | Histology 2nd event | Surgery | AdjuvanT Treatment Pri (RT/ HT) | ER Pri | ER 2nd event | Her2 Pri | Her2 2nd event | Grade Pri | Grade 2nd event | Quadrant 2nd event | Margins | Screening | Clonality P value Copy N | Clonality P value Panel seq | Clonality P value WES | Final verdict |
|---|---|---|---|---|---|---|---|---|---|---|---|---|---|---|---|---|---|---|
| metachronous | 54 | Ipsilateral | IDC with DCIS* | lumpectomy | RT | + | + | - | NA | 3 | 3 | at or adjacent to primary | Clear | screen-detected | 0.000466 2 | NA | NA | Related |

| Clonality P value for ipsilateral synchronous 2nd event Copy Number |
|---|
| 0.004662 |

### 2nd event (INV)

LogR

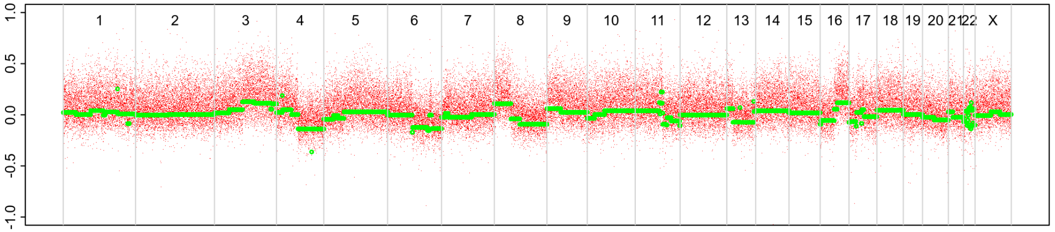

BAF

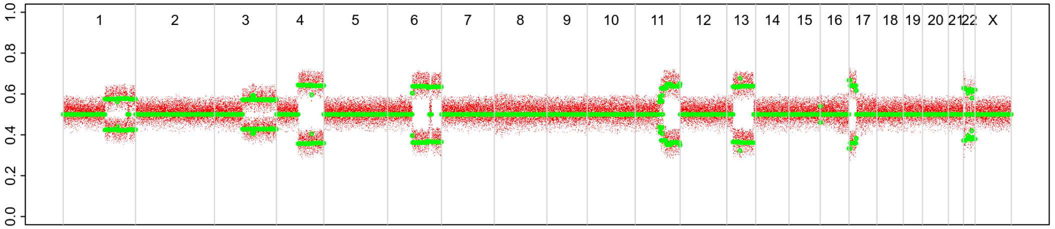

### Primary event

LogR

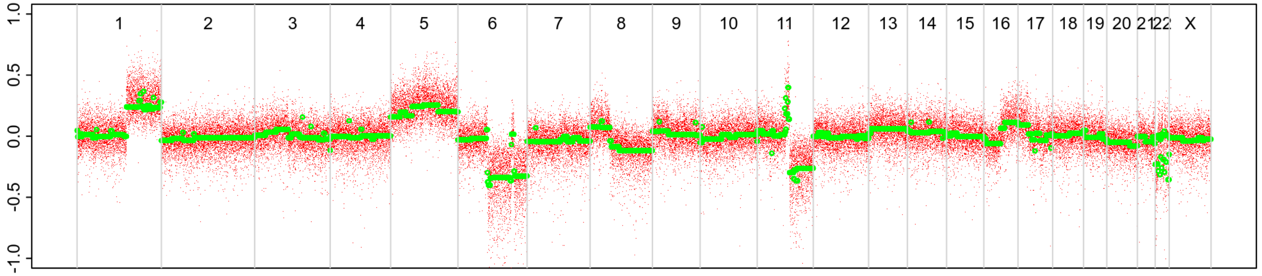

BAF

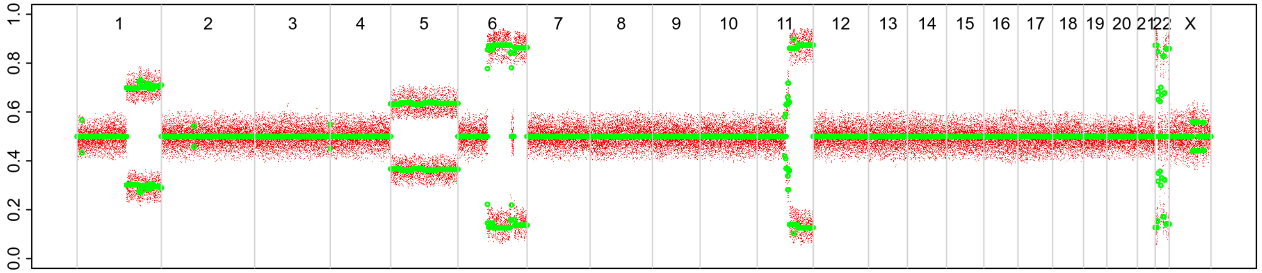

### 2nd event (DCIS)

LogR

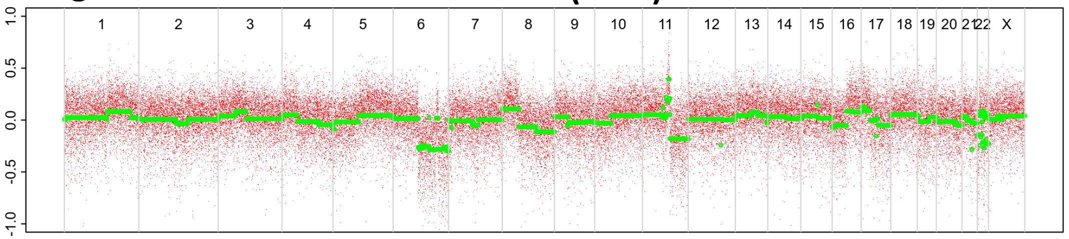

BAF

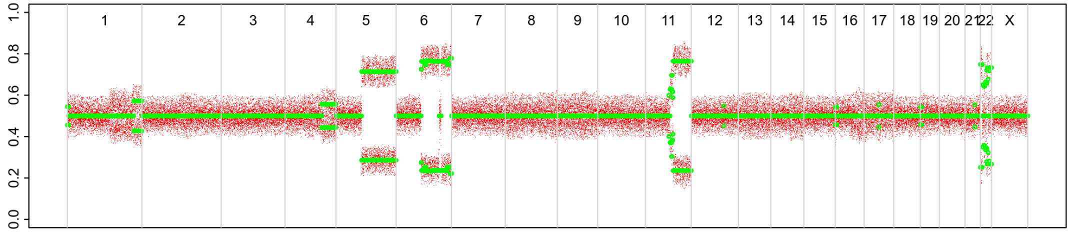

# P078

| Syn/Meta | Time from 1st surgery to 2nd event (Months) | Side 2nd event | Histology 2nd event | Surgery | AdjuvanT Treatment Pri (RT/ HT) | ER Pri | ER 2nd event | Her2 Pri | Her2 2nd event | Grade Pri | Grade 2nd event | Quadrant 2nd event | Margins | Screening | Clonality P value Copy N | Clonality P value Panel seq | Clonality P value WES | Final verdict |
|---|---|---|---|---|---|---|---|---|---|---|---|---|---|---|---|---|---|---|
| metachronous | 51 | Ipsilateral | IDC no DCIS | lumpectomy | CTRTHT | + | NA | - | NA | 3 | 1 | at or adjacent to primary | Clear | screen-detected | 0.000466 2 | NA | NA | Related |

### Primary event

**LogR**

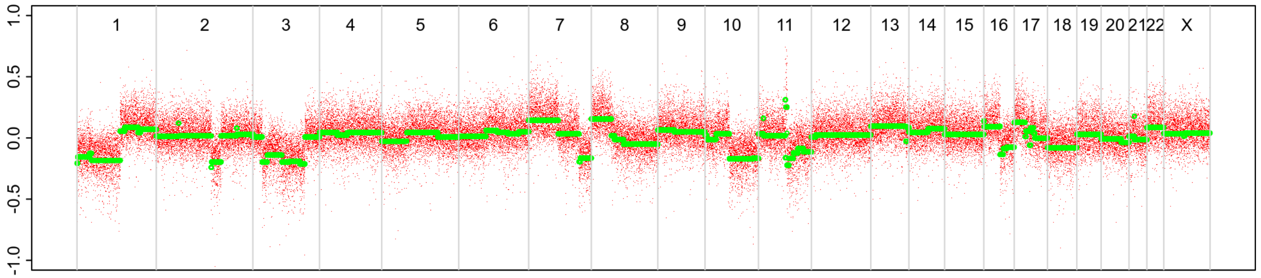


**BAF**

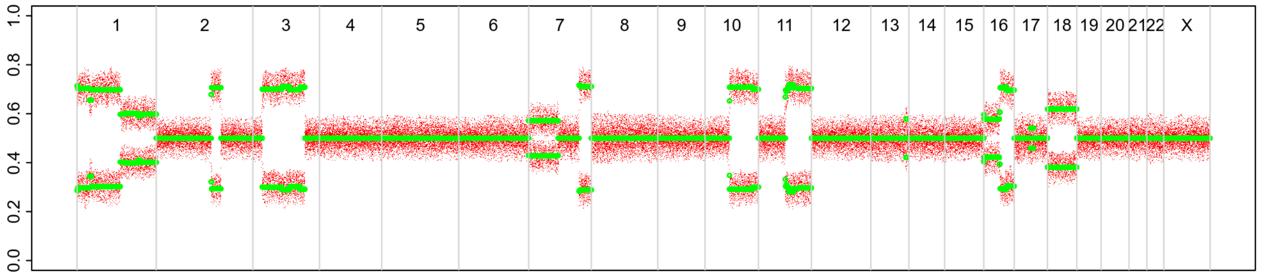


### 2nd event

**LogR**

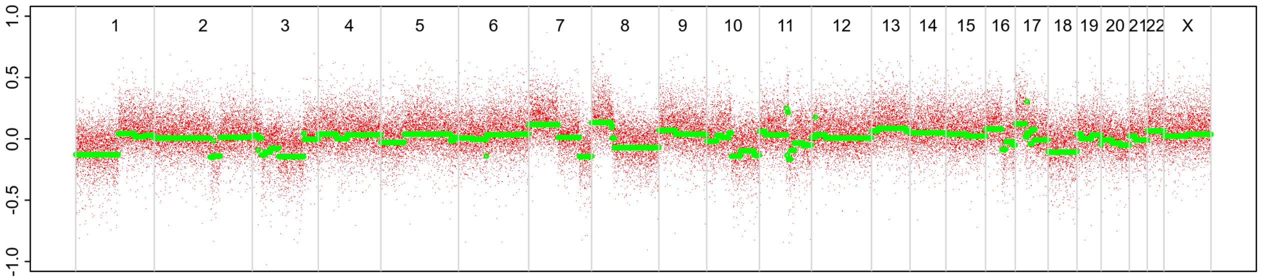


**BAF**

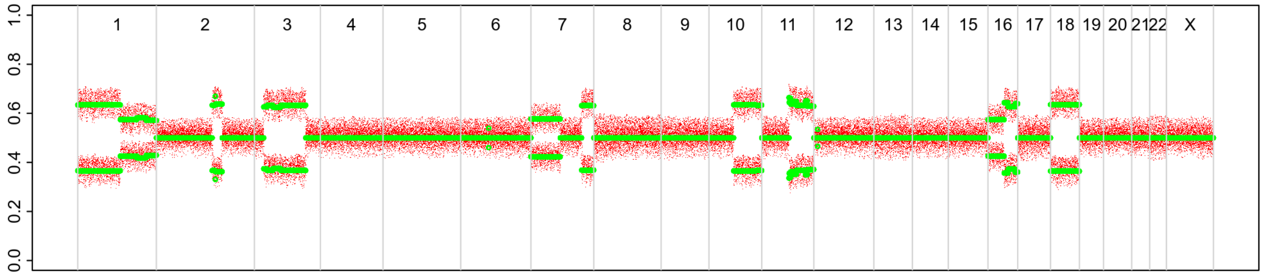

# P079

| Syn/Meta | Time from 1st surgery to 2nd event (Months) | Side 2nd event | Histology 2nd event | Surgery | AdjuvanT Treatment Pri (RT/ HT) | ER Pri | ER 2nd event | Her2 Pri | Her2 2nd event | Grade Pri | Grade 2nd event | Quadrant 2nd event | Margins | Screening | Clonality P value Copy N | Clonality P value Panel seq | Clonality P value WES | Final verdict |
|---|---|---|---|---|---|---|---|---|---|---|---|---|---|---|---|---|---|---|
| metachronous | 38 | Ipsilateral | IDC with DCIS* | lumpectomy | None | + | NA | - | - | 3 | 3 | at or adjacent to primary | Clear | screen-detected | 0.001864802 | 0 | NA | Related |

| Clonality P value for ipsilateral synchronous 2nd event Copy Number |
|---|
| 0.004662 |

### Primary event

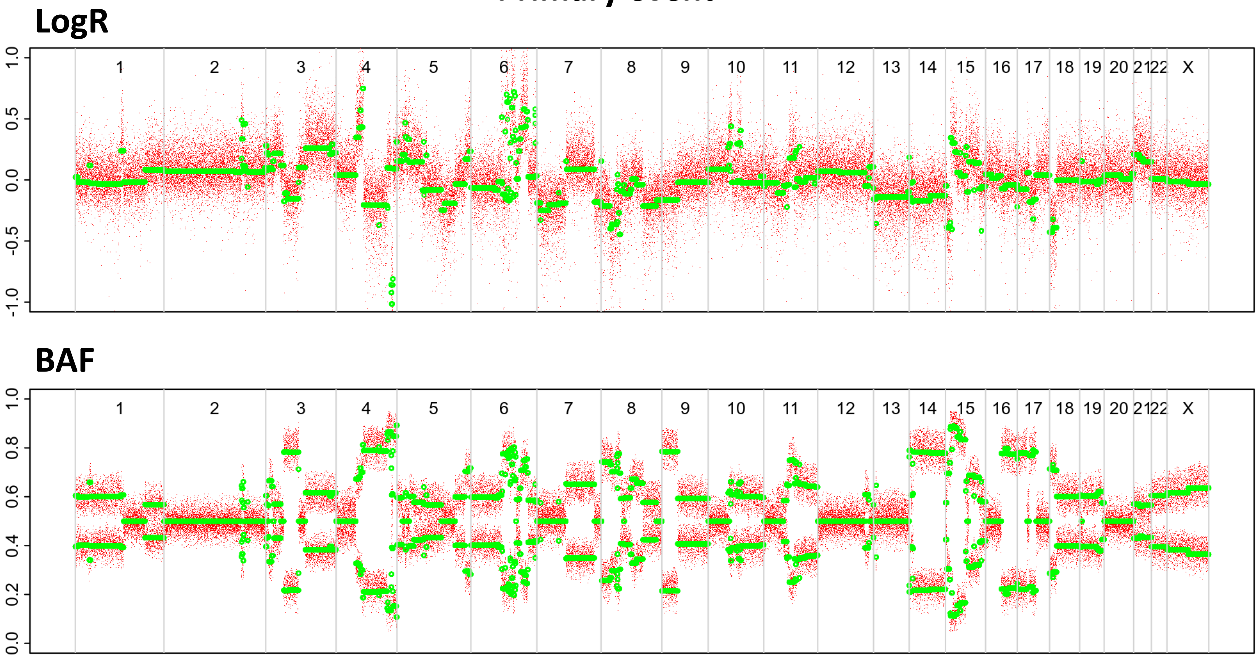

### 2nd event (INV)

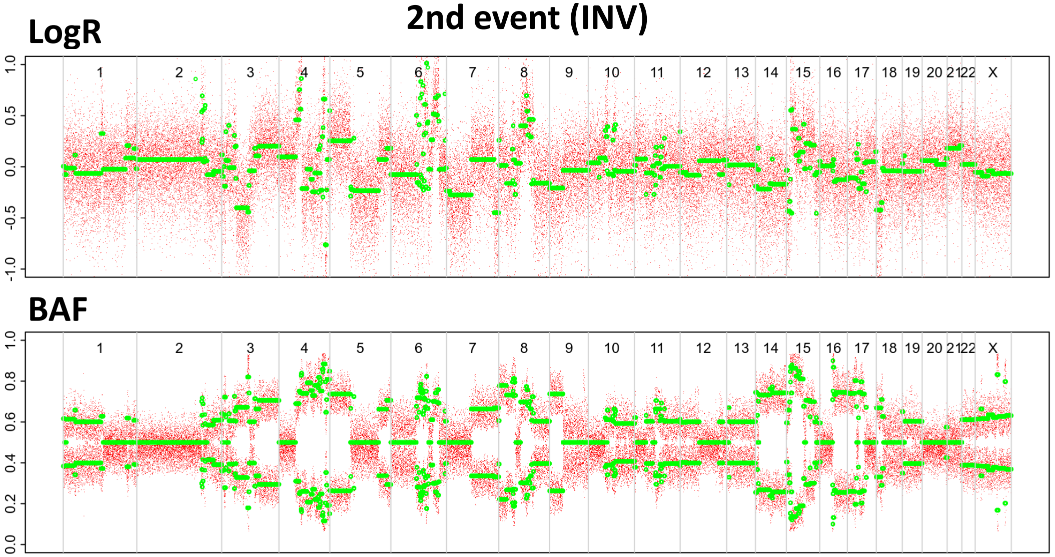

### 2nd event (DCIS)

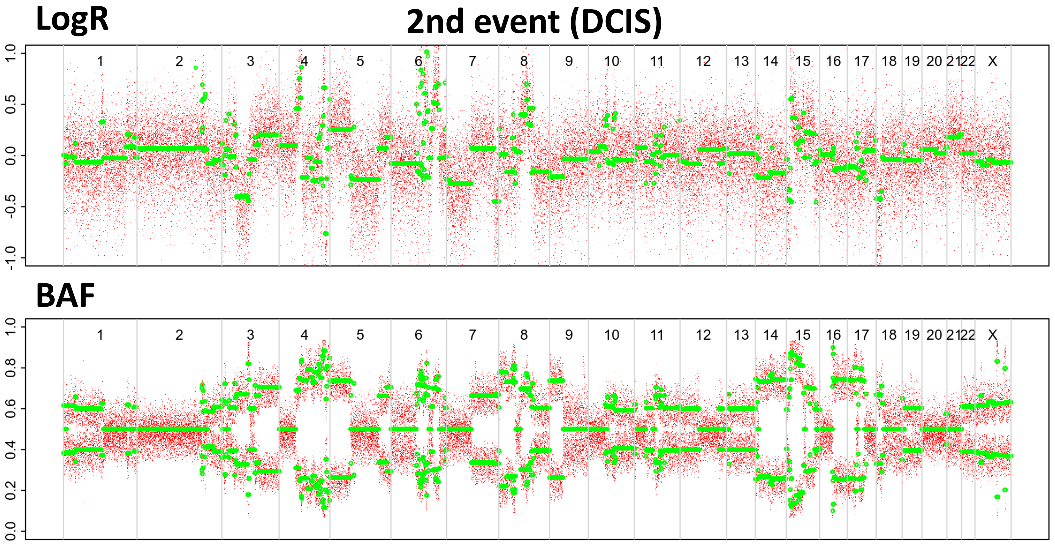

# P080

| Syn/Meta | Time from 1st surgery to 2nd event (Months) | Side 2nd event | Histology 2nd event | Surgery | AdjuvanT Treatment Pri (RT/ HT) | ER Pri | ER 2nd event | Her2 Pri | Her2 2nd event | Grade Pri | Grade 2nd event | Quadrant 2nd event | Margins | Screening | Clonality P value Copy N | Clonality P value Panel seq | Clonality P value WES | Final verdict |
|---|---|---|---|---|---|---|---|---|---|---|---|---|---|---|---|---|---|---|
| metachronous | 35 | Ipsilateral | IDC with DCIS | lumpectomy | RT | + | NA | - | NA | 3 | 3 | at or adjacent to primary | Clear | screen-detected | 0.043822844 | 0.004036327 | NA | Related |

### Primary event

LogR

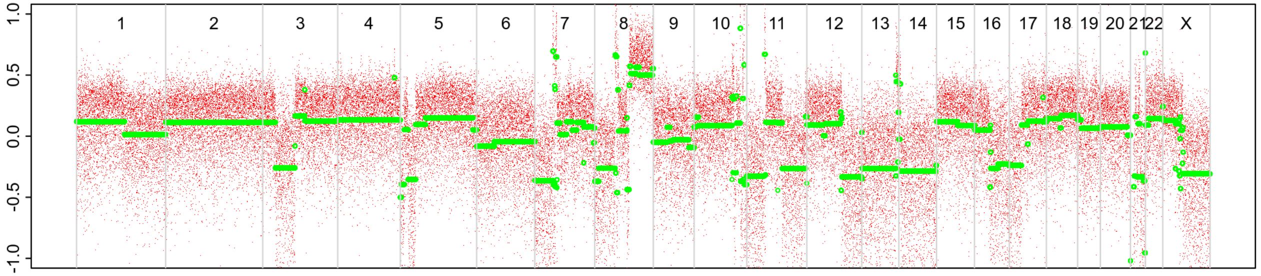


BAF

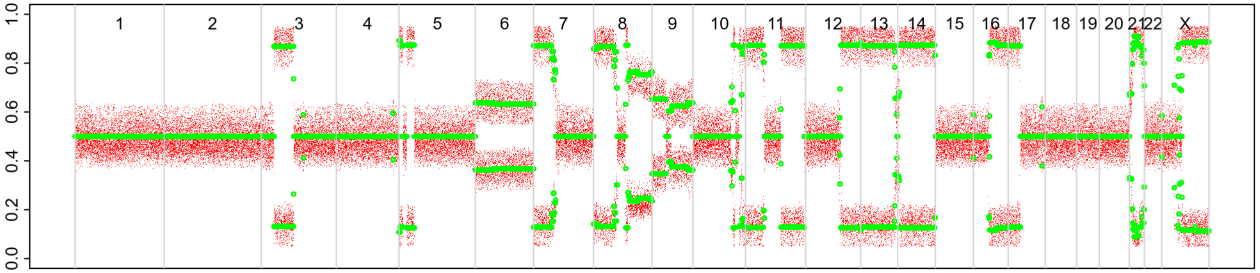


### 2nd event

LogR

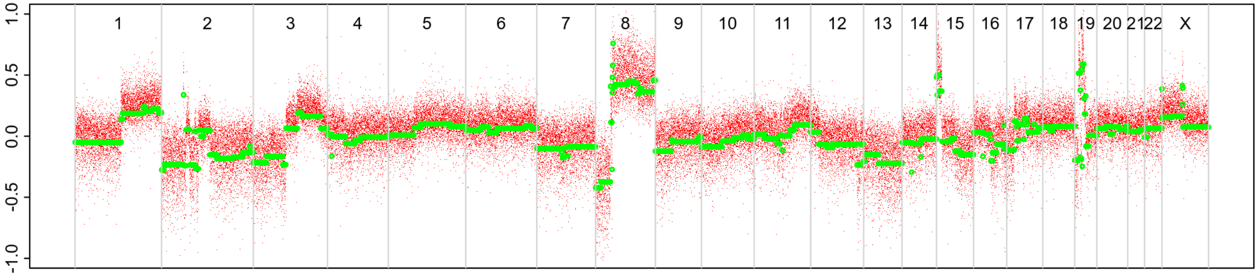


BAF

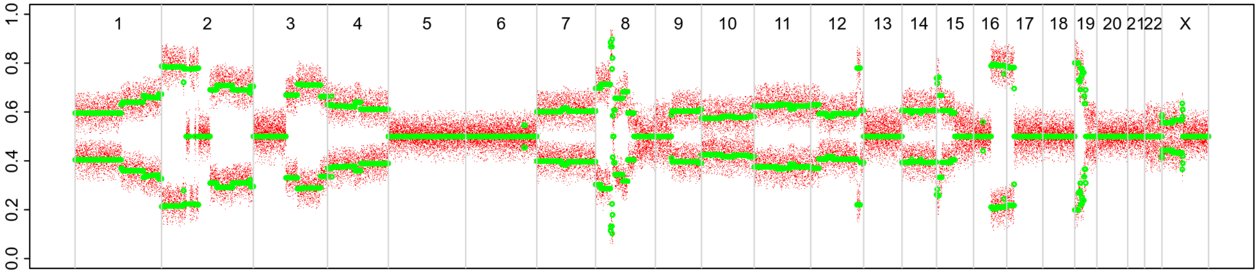

# P092

| Syn/Meta | Time from 1st surgery to 2nd event (Months) | Side 2nd event | Histology 2nd event | Surgery | AdjuvanT Treatment Pri (RT/ HT) | ER Pri | ER 2nd event | Her2 Pri | Her2 2nd event | Grade Pri | Grade 2nd event | Quadrant 2nd event | Margins | Screening | Clonality P value Copy N | Clonality P value Panel seq | Clonality P value WES | Final verdict |
|---|---|---|---|---|---|---|---|---|---|---|---|---|---|---|---|---|---|---|
| metachronous | 22 | Ipsilateral | IDC with DCIS* | lumpectomy | RT | + | + | NA | NA | 2 | 2 | at or adjacent to primary | Clear | screen-detected | 0.000466 2 | NA | 0.00075244 5 | Related |

| Clonality P value for ipsilateral synchronous 2nd event Copy Number |
|---|
| 0.004662 |

### 2nd event (INV)

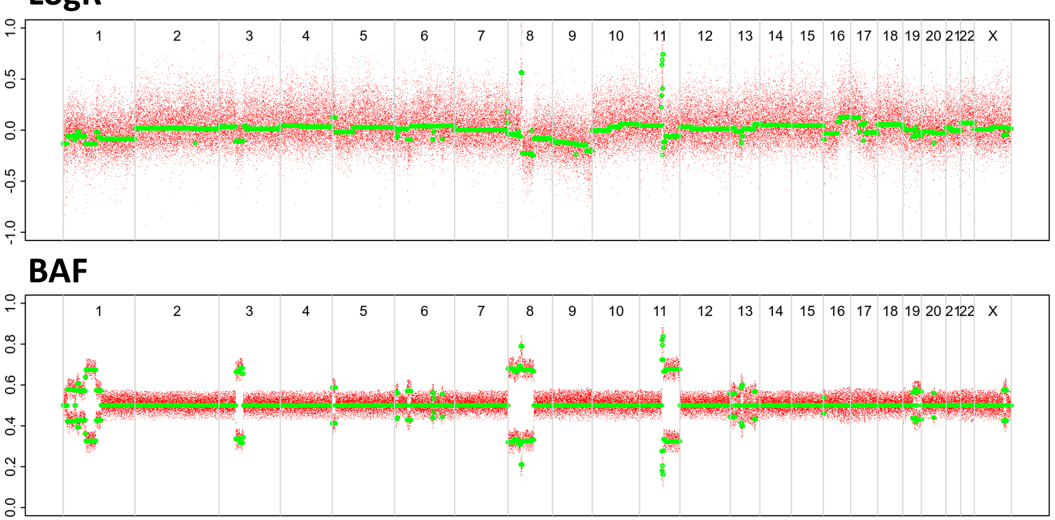


### Primary event

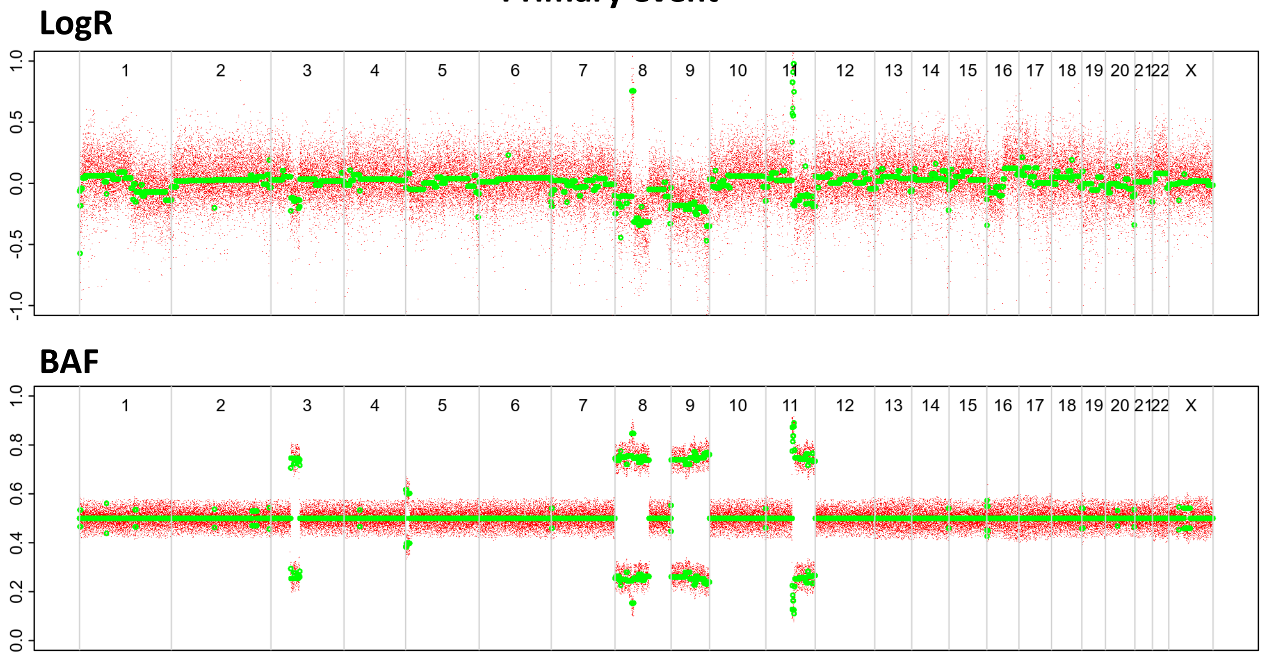


### 2nd event (DCIS)

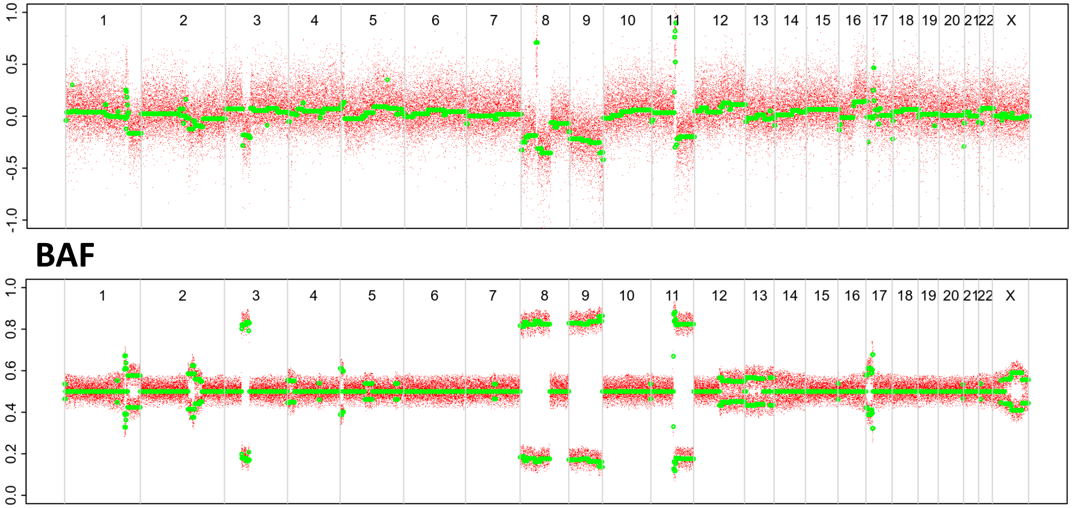

# P099

| Syn/Meta | Time from 1st surgery to 2nd event (Months) | Side 2nd event | Histology 2nd event | Surgery | AdjuvanT Treatment Pri (RT/ HT) | ER Pri | ER 2nd event | Her2 Pri | Her2 2nd event | Grade Pri | Grade 2nd event | Quadrant 2nd event | Margins | Screening | Clonality P value Copy N | Clonality P value Panel seq | Clonality P value WES | Final verdict |
|---|---|---|---|---|---|---|---|---|---|---|---|---|---|---|---|---|---|---|
| metachronous | 22 | Ipsilateral | IDC no DCIS | lumpectomy | None | + | + | - | - | 3 | 3 | at or adjacent to primary | Clear | screen-detected | 0.000466 2 | 0 | NA | Related |

### Primary event

LogR

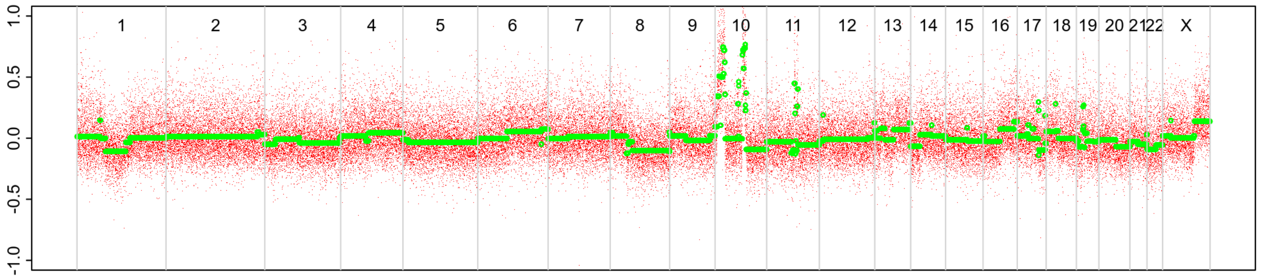


BAF

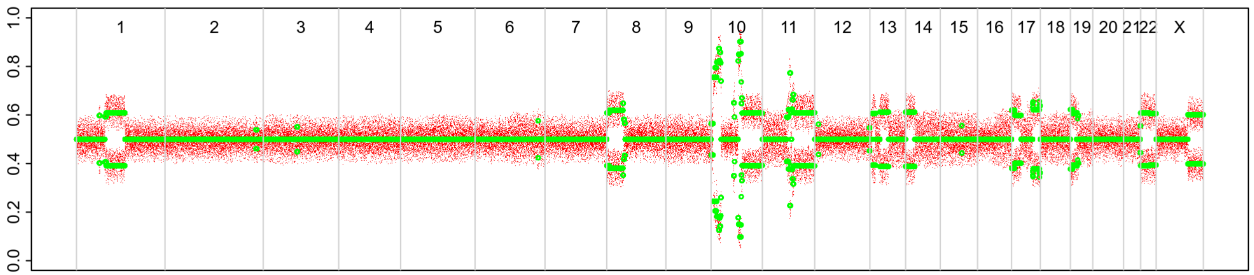


### 2nd event

LogR

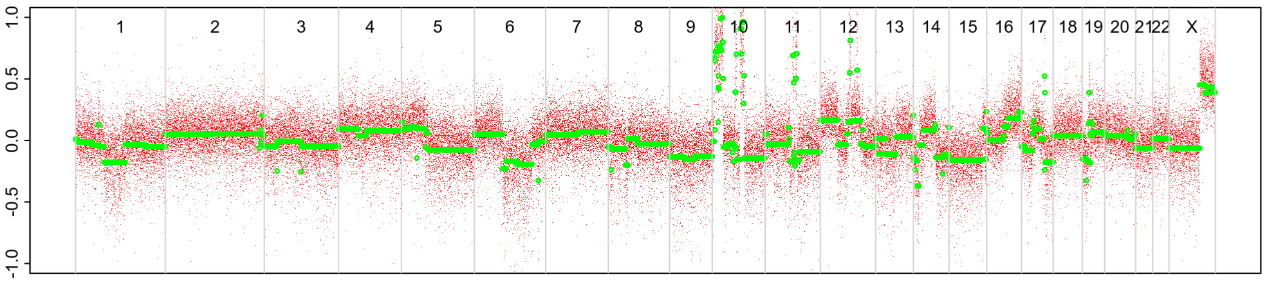


BAF

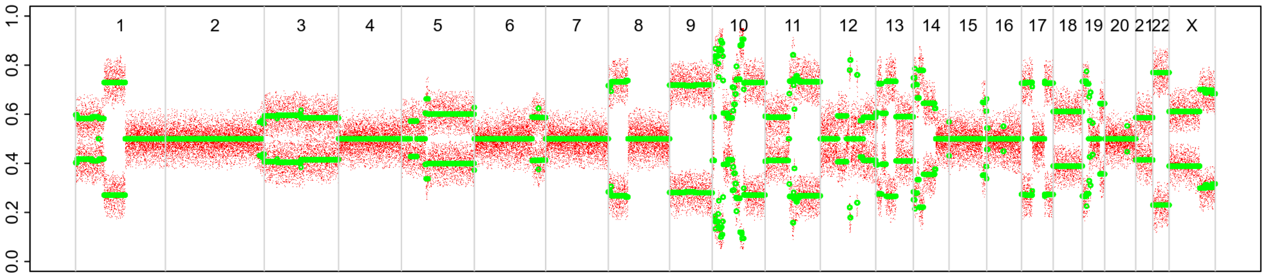

# P100

| Syn/Meta | Time from 1st surgery to 2nd event (Months) | Side 2nd event | Histology 2nd event | Surgery | AdjuvanT Treatment Pri (RT/ HT) | ER Pri | ER 2nd event | Her2 Pri | Her2 2nd event | Grade Pri | Grade 2nd event | Quadrant 2nd event | Margins | Screening | Clonality P value Copy N | Clonality P value Panel seq | Clonality P value WES | Final verdict |
|---|---|---|---|---|---|---|---|---|---|---|---|---|---|---|---|---|---|---|
| metachronous | 59 | Ipsilateral | DCIS only | lumpectomy | None | + | + | + | NA | 3 | 3 | NA | Clear | screen-detected | 0.014652015 | NA | NA | Equivocal |

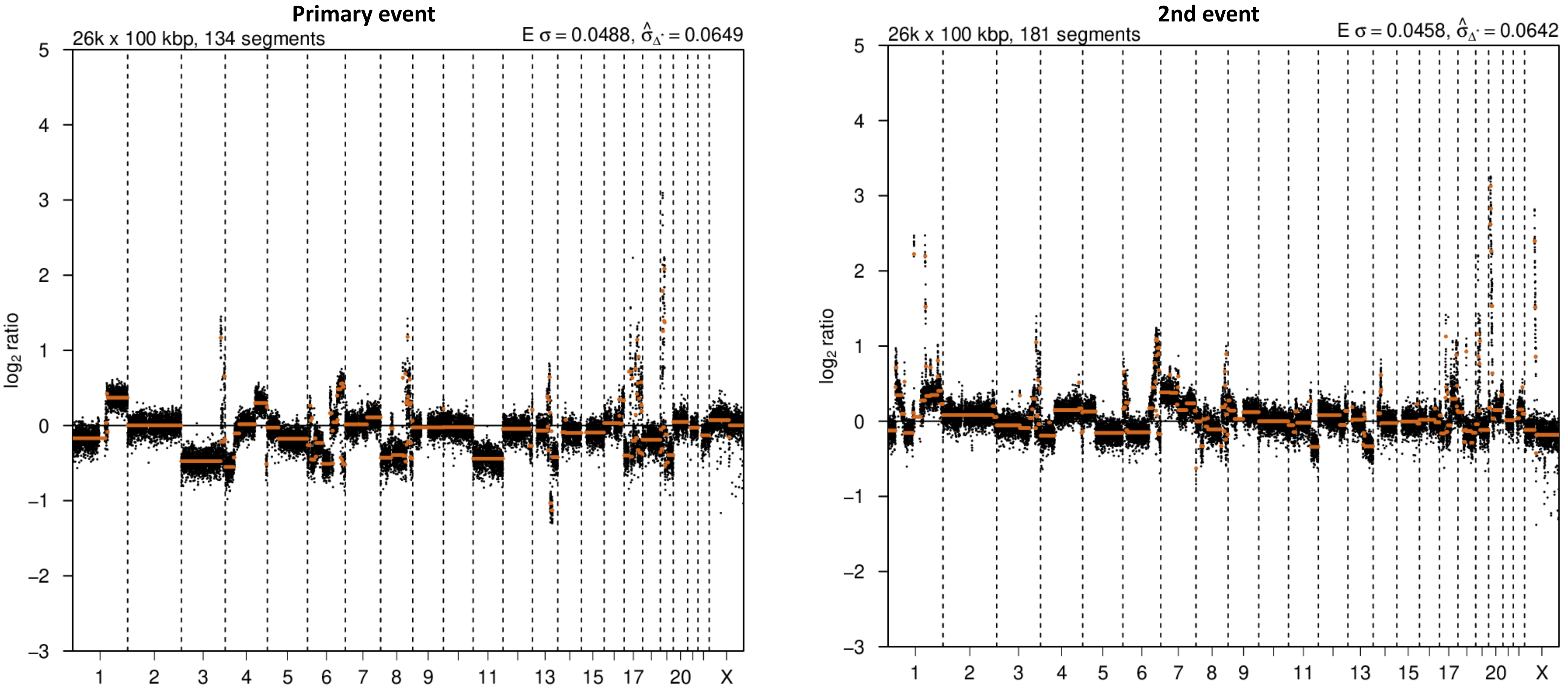

# P101

| Syn/Meta | Time from 1st surgery to 2nd event (Months) | Side 2nd event | Histology 2nd event | Surgery | AdjuvanT Treatment Pri (RT/ HT) | ER Pri | ER 2nd event | Her2 Pri | Her2 2nd event | Grade Pri | Grade 2nd event | Quadrant 2nd event | Margins | Screening | Clonality P value Copy N | Clonality P value Panel seq | Clonality P value WES | Final verdict |
|---|---|---|---|---|---|---|---|---|---|---|---|---|---|---|---|---|---|---|
| metachronous | 41 | Ipsilateral | DCIS only | lumpectomy | None | NA | NA | NA | NA | 3 | 3 | NA | Clear | screen-detected | 0.025641026 | NA | NA | Equivocal |

### Primary event

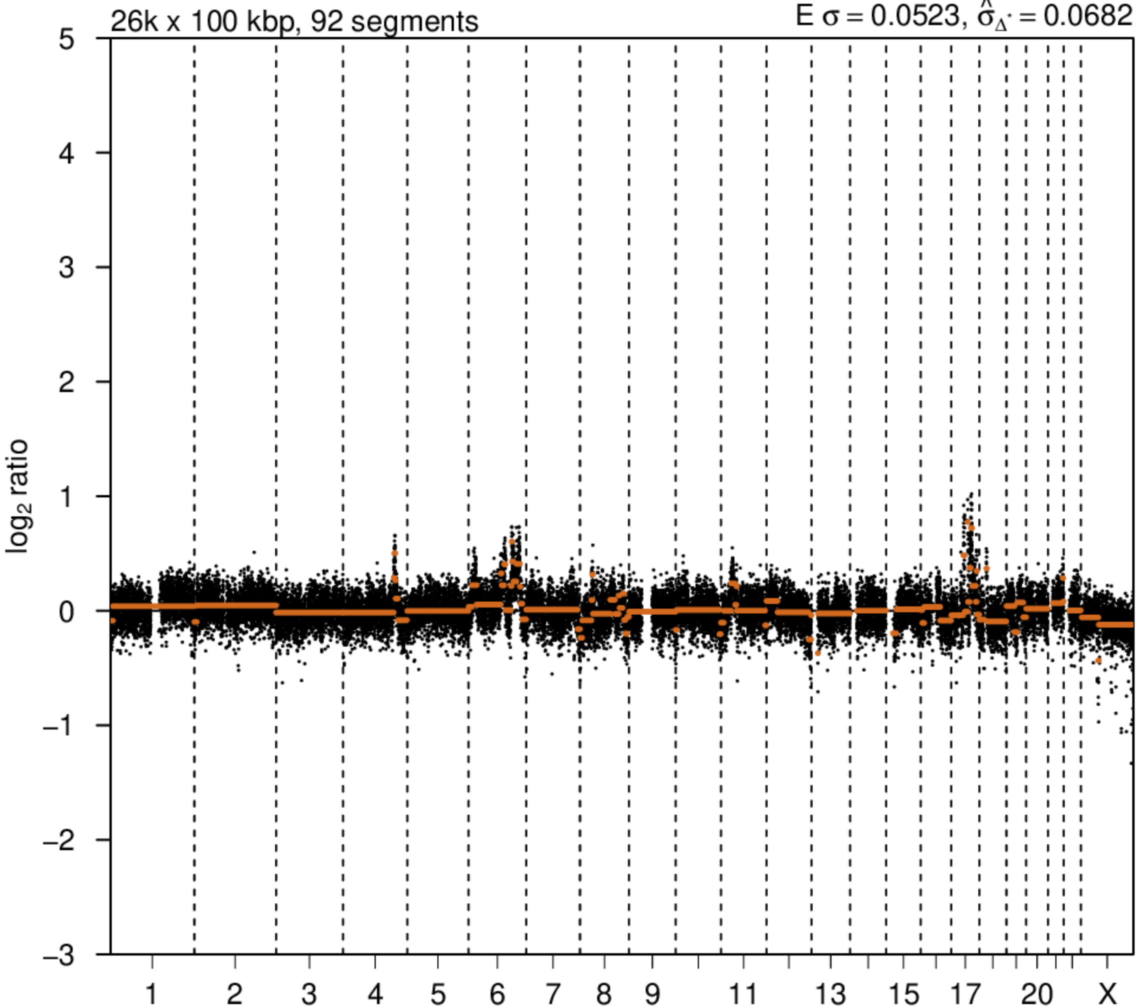


### 2nd event

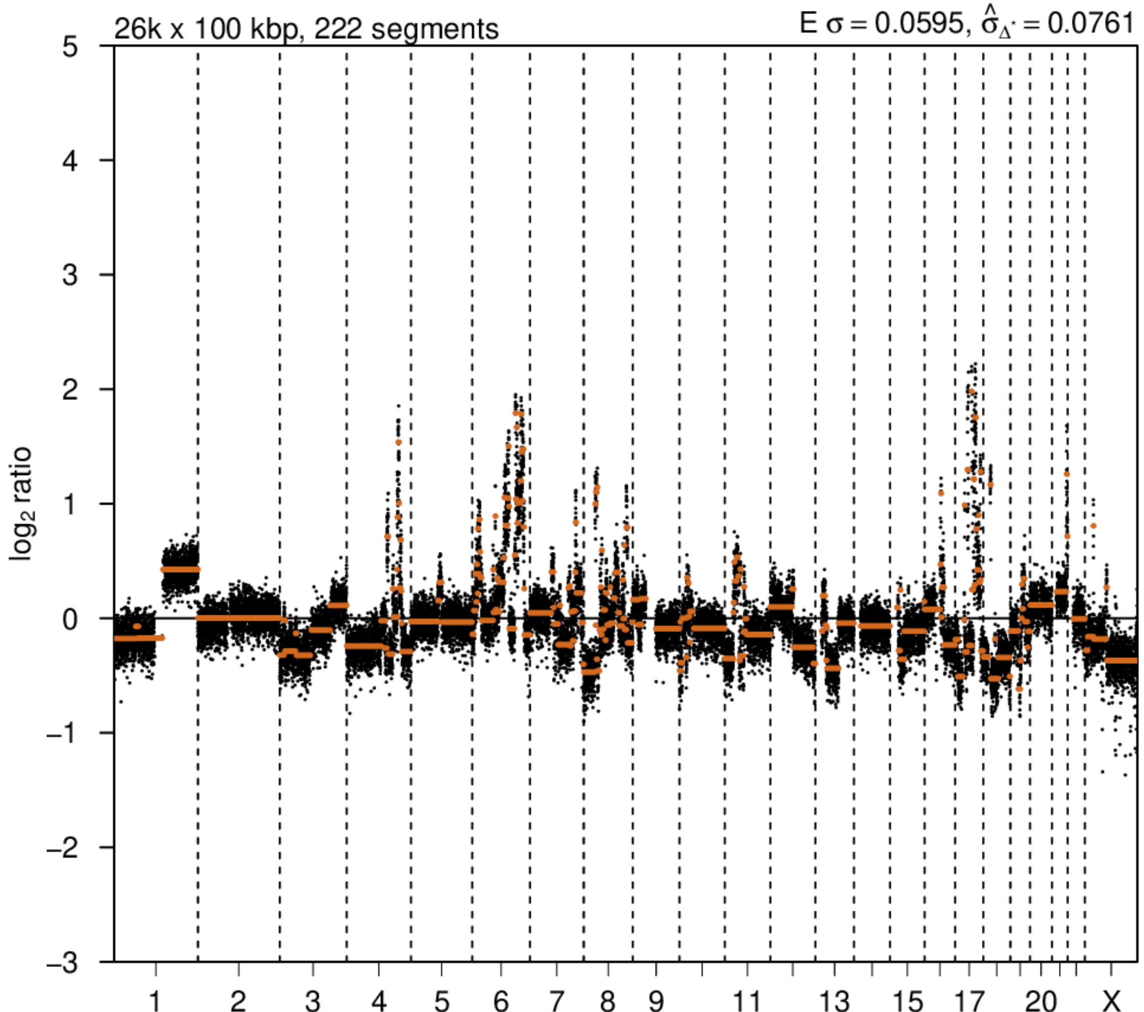

# P108

| Syn/Meta | Time from 1st surgery to 2nd event (Months) | Side 2nd event | Histology 2nd event | Surgery | AdjuvanT Treatment Pri (RT/ HT) | ER Pri | ER 2nd event | Her2 Pri | Her2 2nd event | Grade Pri | Grade 2nd event | Quadrant 2nd event | Margins | Screening | Clonality P value Copy N | Clonality P value Panel seq | Clonality P value WES | Final verdict |
|---|---|---|---|---|---|---|---|---|---|---|---|---|---|---|---|---|---|---|
| metachronous | 74 | Ipsilateral | IDC with DCIS* | lumpectomy | RT | + | + | - | - | 3 | 3 | at or adjacent to primary | Clear | screen-detected | 0.009324009 | NA | NA | Related |

| Clonality P value for ipsilateral synchronous 2nd event Copy Number |
|---|
| 0.004662 |

### Primary event

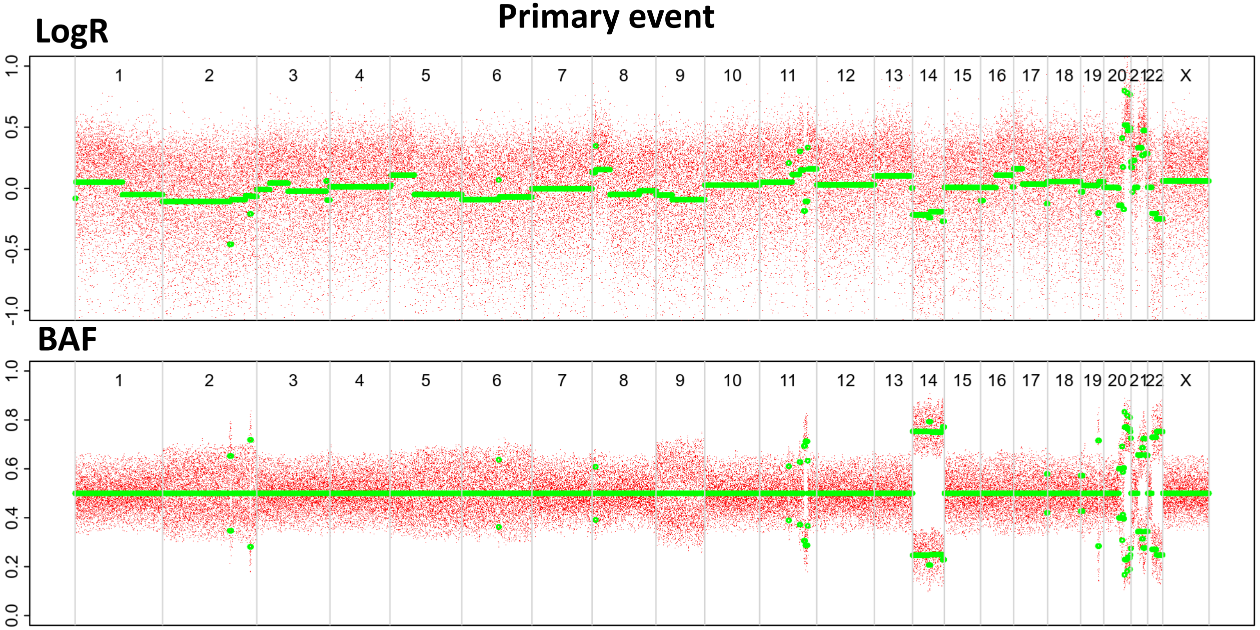


### 2nd event (INV)

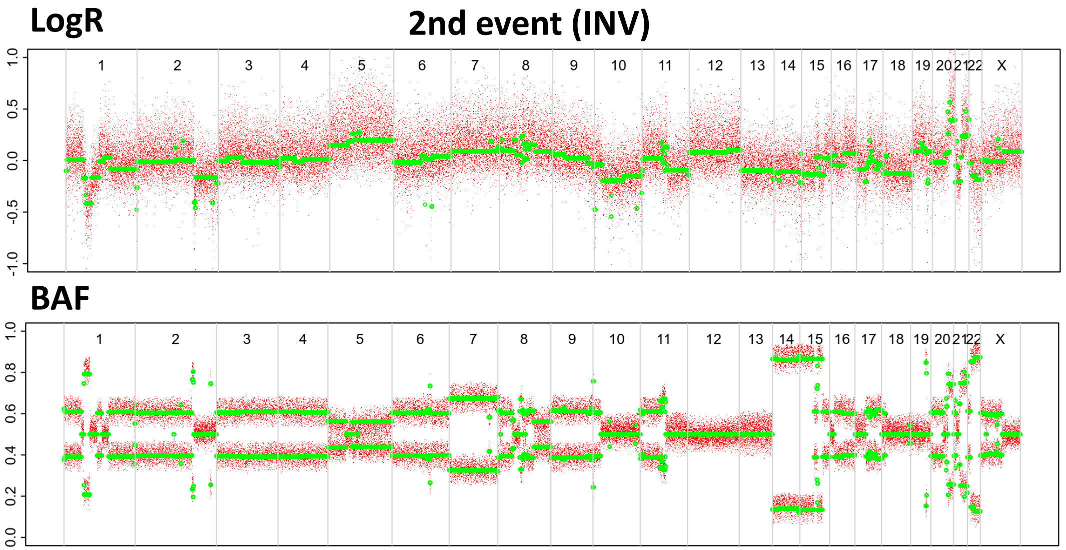


### 2nd event (DCIS)

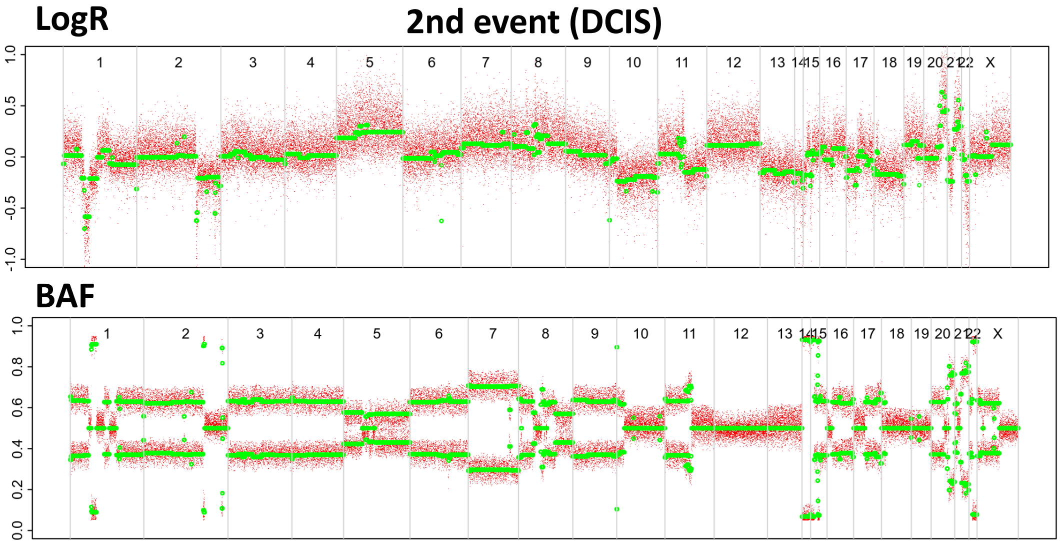

# P112

| Syn/Meta | Time from 1st surgery to 2nd event (Months) | Side 2nd event | Histology 2nd event | Surgery | AdjuvanT Treatment Pri (RT/ HT) | ER Pri | ER 2nd event | Her2 Pri | Her2 2nd event | Grade Pri | Grade 2nd event | Quadrant 2nd event | Margins | Screening | Clonality P value Copy N | Clonality P value Panel seq | Clonality P value WES | Final verdict |
|---|---|---|---|---|---|---|---|---|---|---|---|---|---|---|---|---|---|---|
| metachronous | 24 | Ipsilateral | DCIS only | lumpectomy | None | NA | NA | - | NA | 3 | 3 | NA | Clear | screen-detected | 0.608058608 | NA | NA | Unrelated |

### Primary event

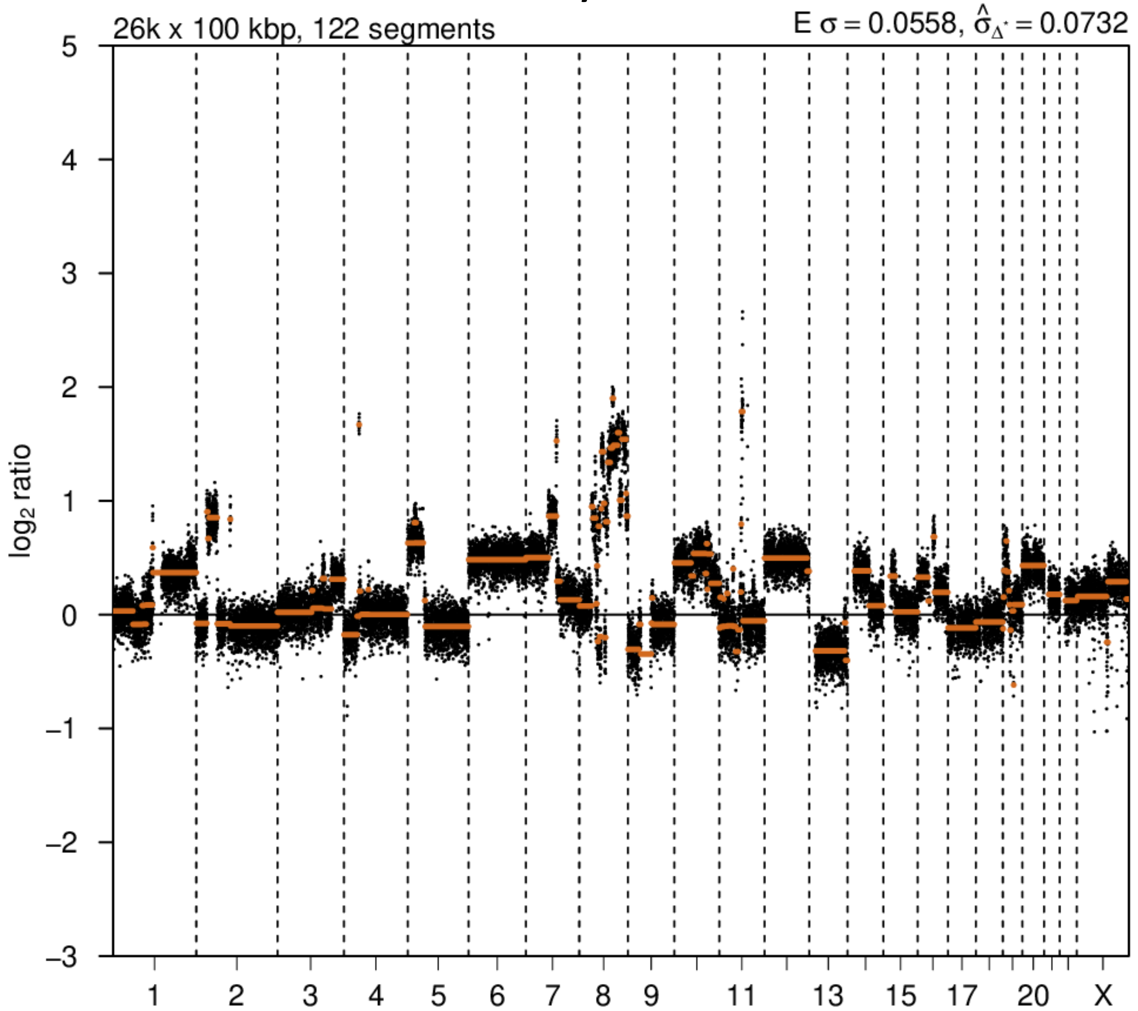


### 2nd event

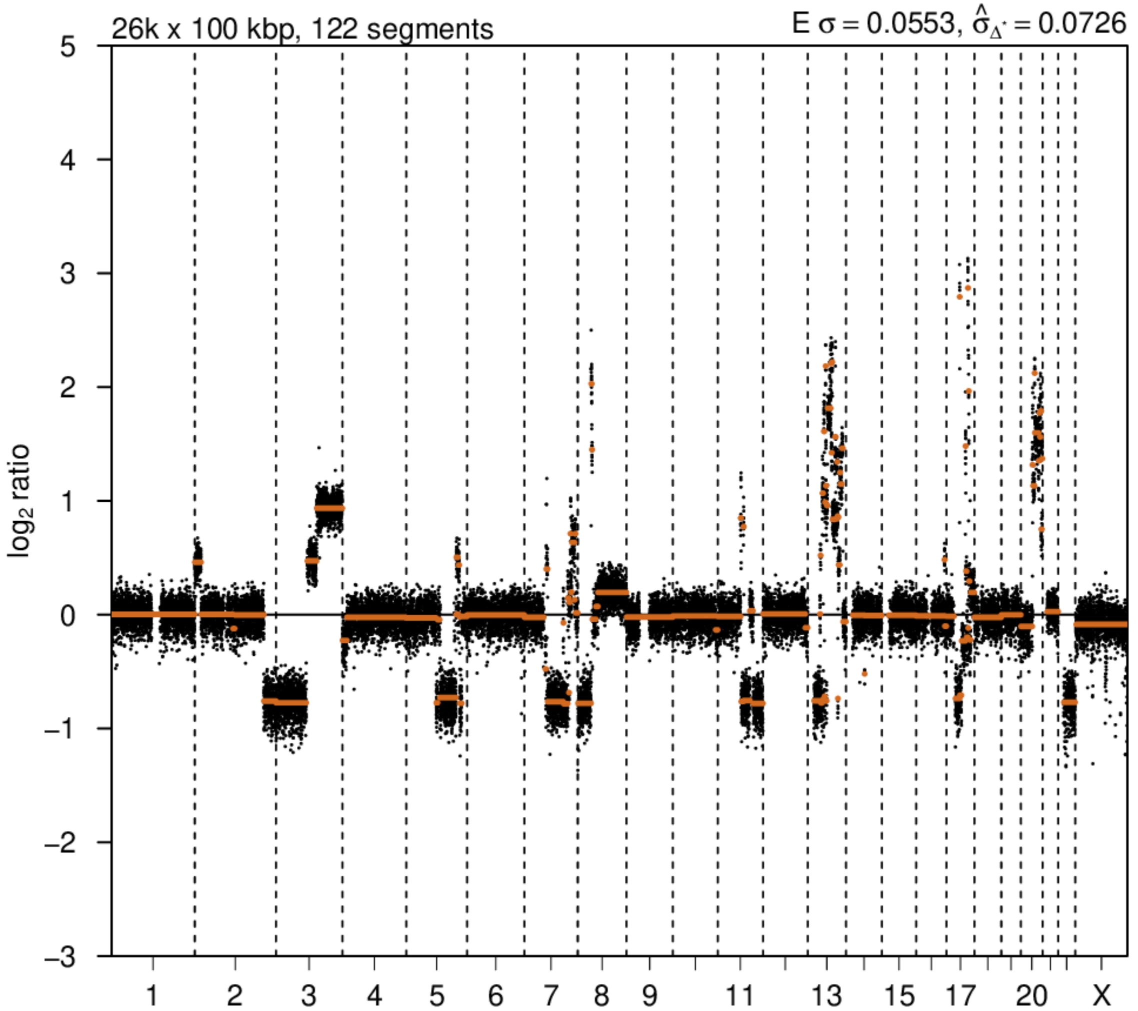

# P122

| Syn/Meta | Time from 1st surgery to 2nd event (Months) | Side 2nd event | Histology 2nd event | Surgery | AdjuvanT Treatment Pri (RT/ HT) | ER Pri | ER 2nd event | Her2 Pri | Her2 2nd event | Grade Pri | Grade 2nd event | Quadrant 2nd event | Margins | Screening | Clonality P value Copy N | Clonality P value Panel seq | Clonality P value WES | Final verdict |
|---|---|---|---|---|---|---|---|---|---|---|---|---|---|---|---|---|---|---|
| metachronous | 88 | Ipsilateral | IDC with DCIS | lumpectomy | RT | - | + | + | + | 3 | 3 | at or adjacent to primary | Clear | screen-detected | 0.158508159 | NA | NA | Unrelated |

| Clonality P value for ipsilateral synchronous 2nd event Copy Number |
|---|
| 0.004662 |

### 2nd event (INV)

LogR

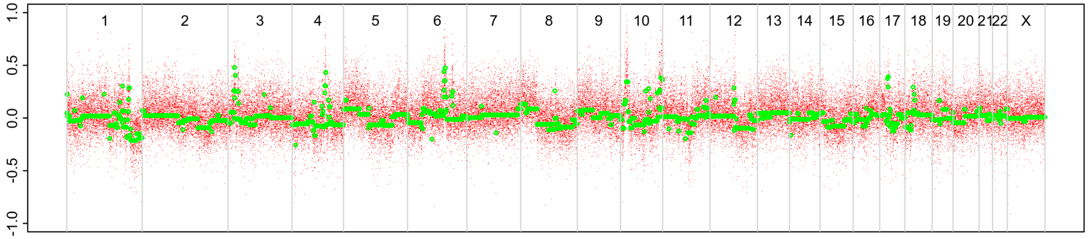


BAF

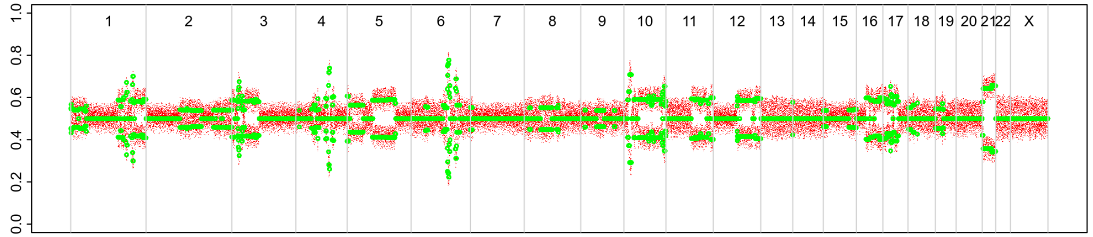


### Primary event

LogR

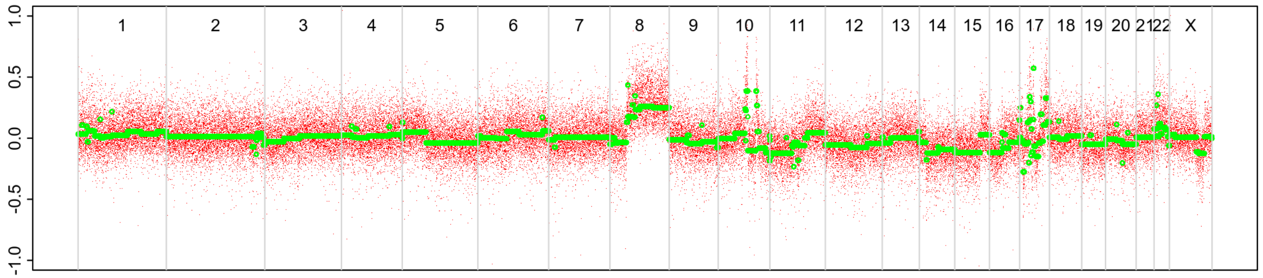


BAF

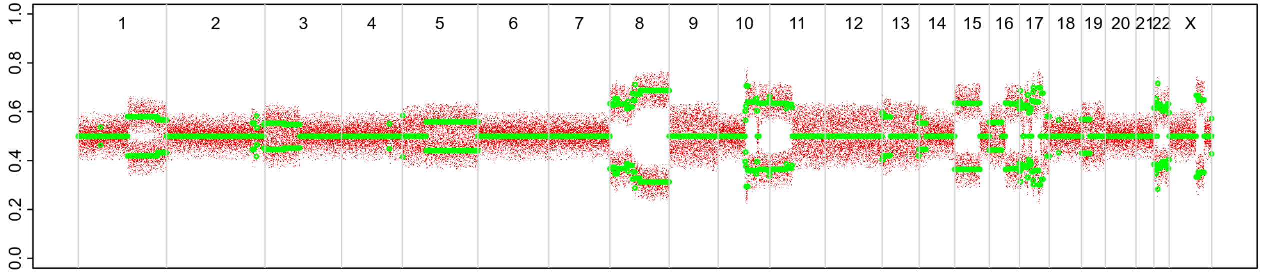


### 2nd event (DCIS)

LogR

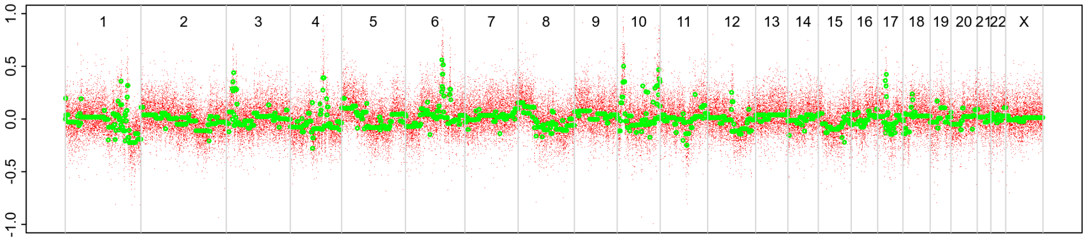


BAF

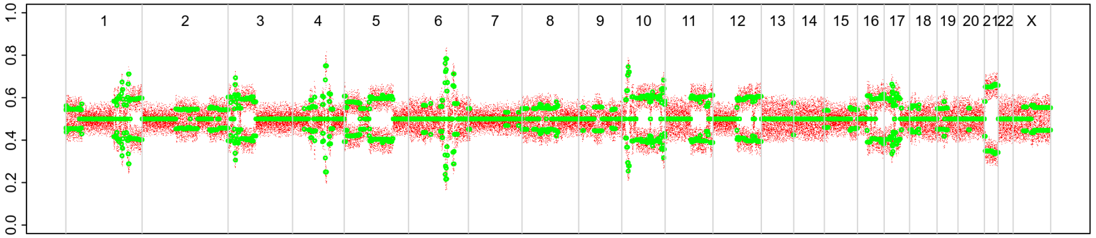

# P128

| Syn/Meta | Time from 1st surgery to 2nd event (Months) | Side 2nd event | Histology 2nd event | Surgery | AdjuvanT Treatment Pri (RT/ HT) | ER Pri | ER 2nd event | Her2 Pri | Her2 2nd event | Grade Pri | Grade 2nd event | Quadrant 2nd event | Margins | Screening | Clonality P value Copy N | Clonality P value Panel seq | Clonality P value WES | Final verdict |
|---|---|---|---|---|---|---|---|---|---|---|---|---|---|---|---|---|---|---|
| metachronous | 24 | Ipsilateral | IDC with DCIS* | lumpectomy | None | + | NA | NA | NA | 3 | 2 | distant from primary | Clear | screen-detected | 0.592540793 | 1 | NA | Unrelated |

| Clonality P value for ipsilateral synchronous 2nd event Copy Number |
|---|
| 0.004662 |

### Primary event

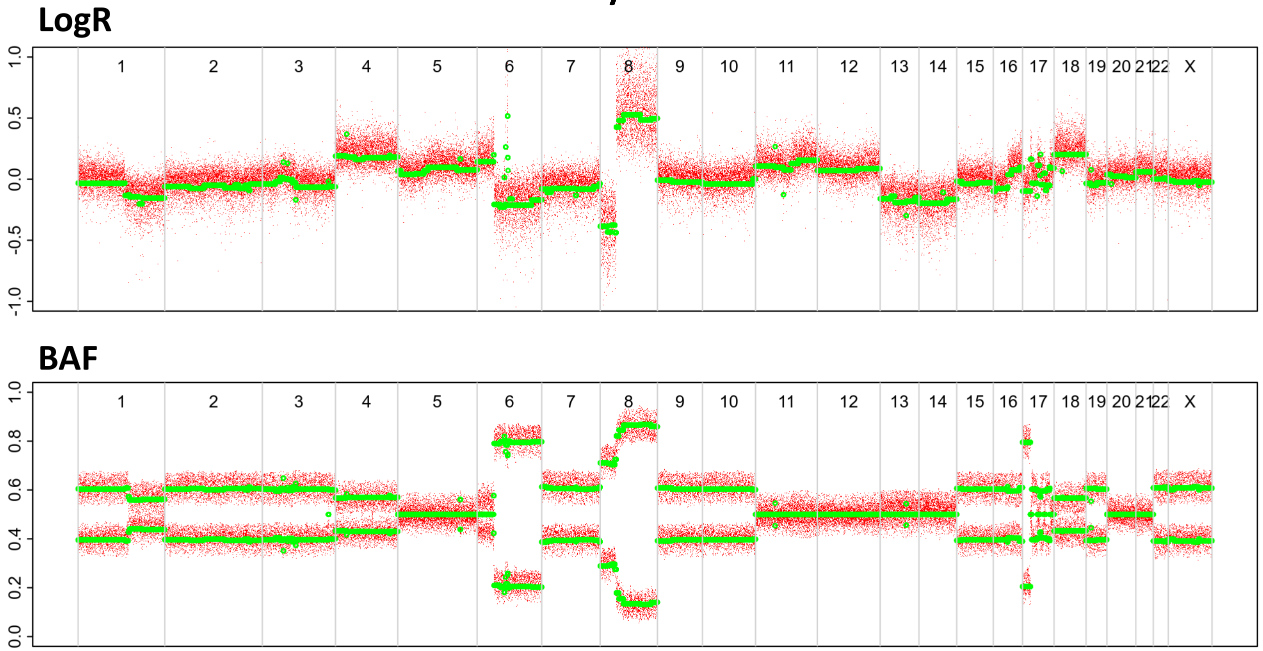


### 2nd event (INV)

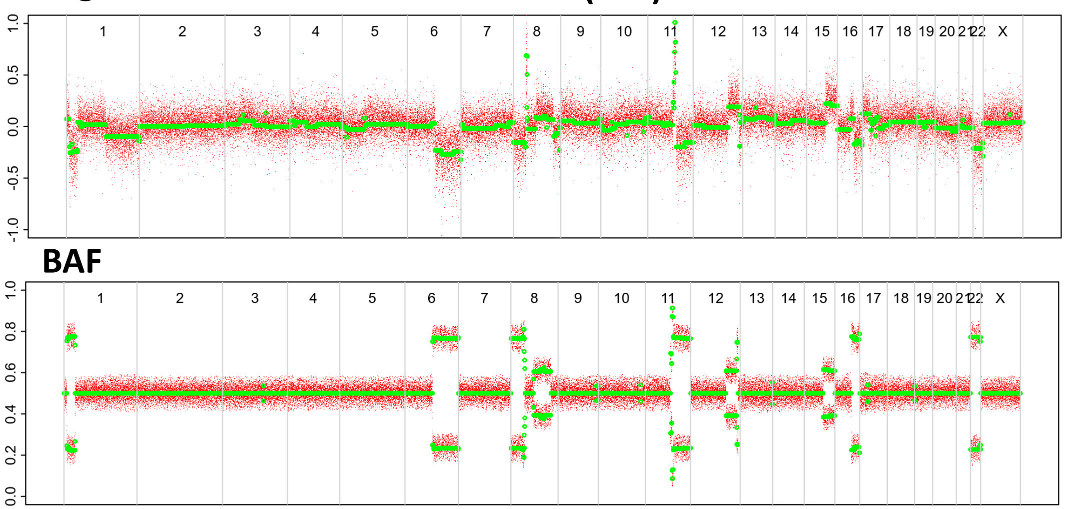


### 2nd event (DCIS)

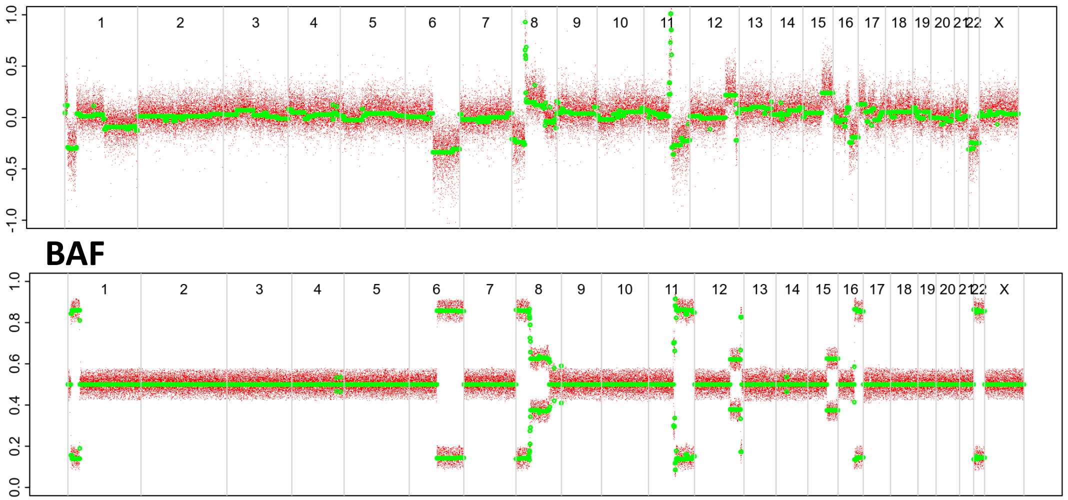

# P130

| Syn/Meta | Time from 1st surgery to 2nd event (Months) | Side 2nd event | Histology 2nd event | Surgery | AdjuvanT Treatment Pri (RT/ HT) | ER Pri | ER 2nd event | Her2 Pri | Her2 2nd event | Grade Pri | Grade 2nd event | Quadrant 2nd event | Margins | Screening | Clonality P value Copy N | Clonality P value Panel seq | Clonality P value WES | Final verdict |
|---|---|---|---|---|---|---|---|---|---|---|---|---|---|---|---|---|---|---|
| metachronous | 34 | Contralateral | IDC no DCIS | lumpectomy | None | + | NA | NA | NA | 2 | 2 | NA | Clear | screen-detected | 1 | NA | 0.002851033 | Related |

### Primary event

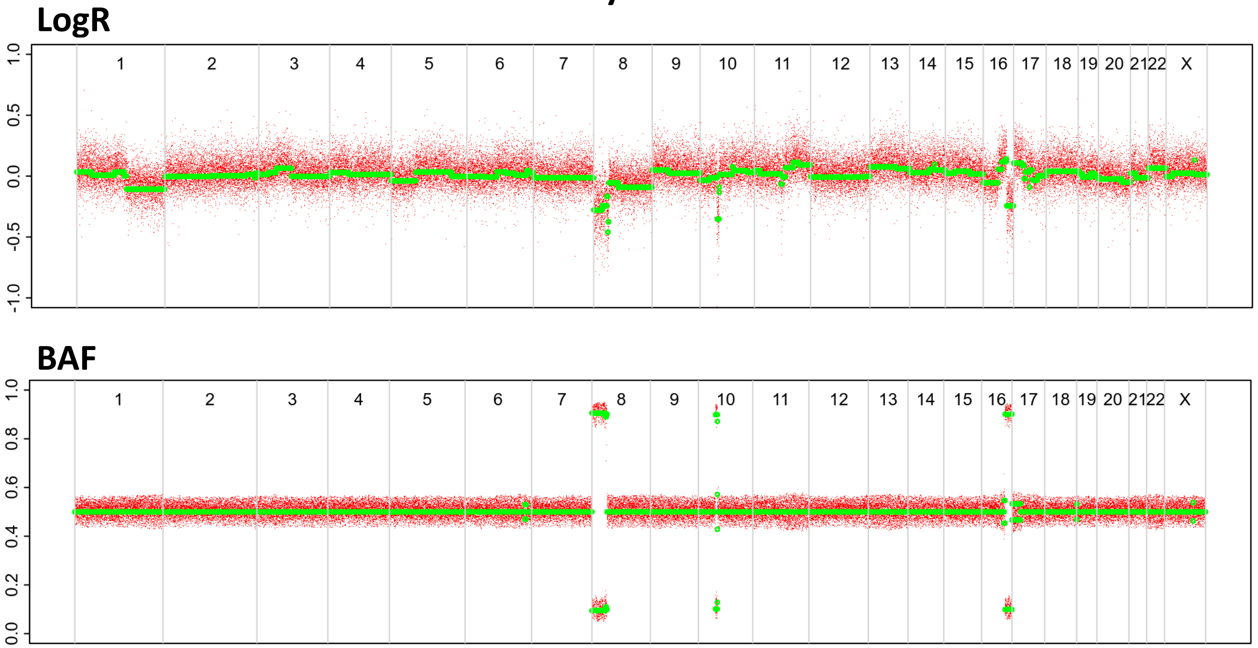


### 2nd event

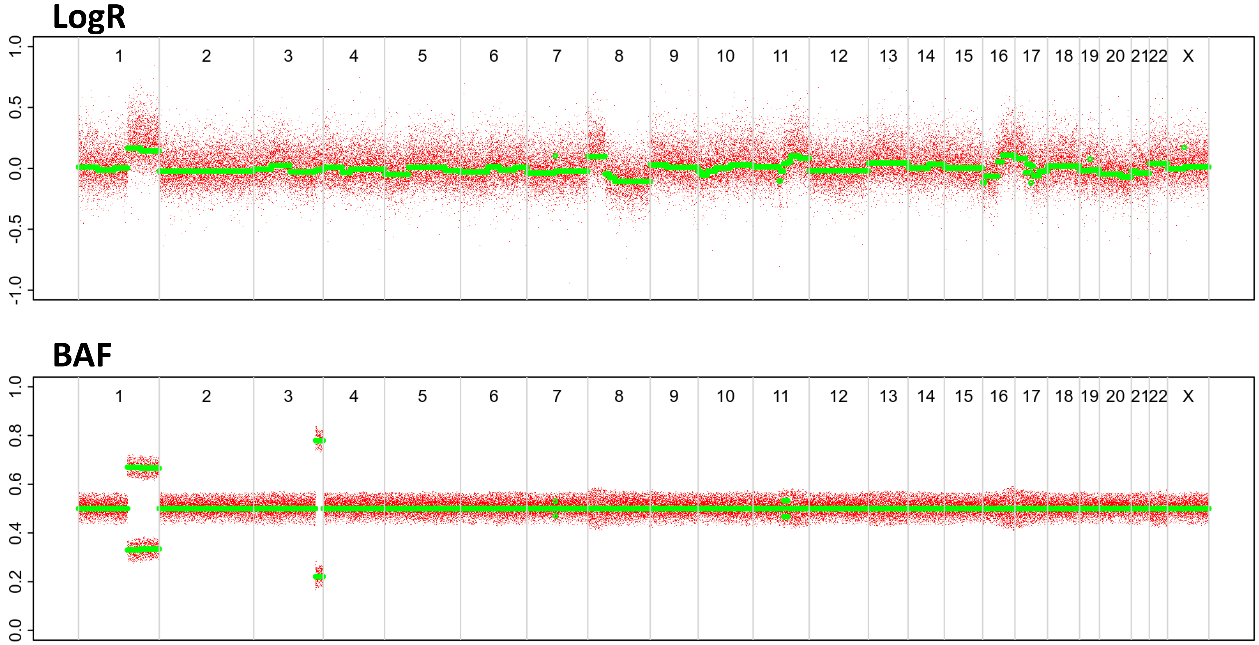

# P131

| Syn/Meta | Time from 1st surgery to 2nd event (Months) | Side 2nd event | Histology 2nd event | Surgery | AdjuvanT Treatment Pri (RT/ HT) | ER Pri | ER 2nd event | Her2 Pri | Her2 2nd event | Grade Pri | Grade 2nd event | Quadrant 2nd event | Margins | Screening | Clonality P value Copy N | Clonality P value Panel seq | Clonality P value WES | Final verdict |
|---|---|---|---|---|---|---|---|---|---|---|---|---|---|---|---|---|---|---|
| metachronous | 15 | Contralateral | IDC no DCIS | lumpectomy | RT | - | - | + | + | 3 | 3 | NA | <2mm | screen-detected | 0.056410256 | 1 | NA | Unrelated |

### Primary event

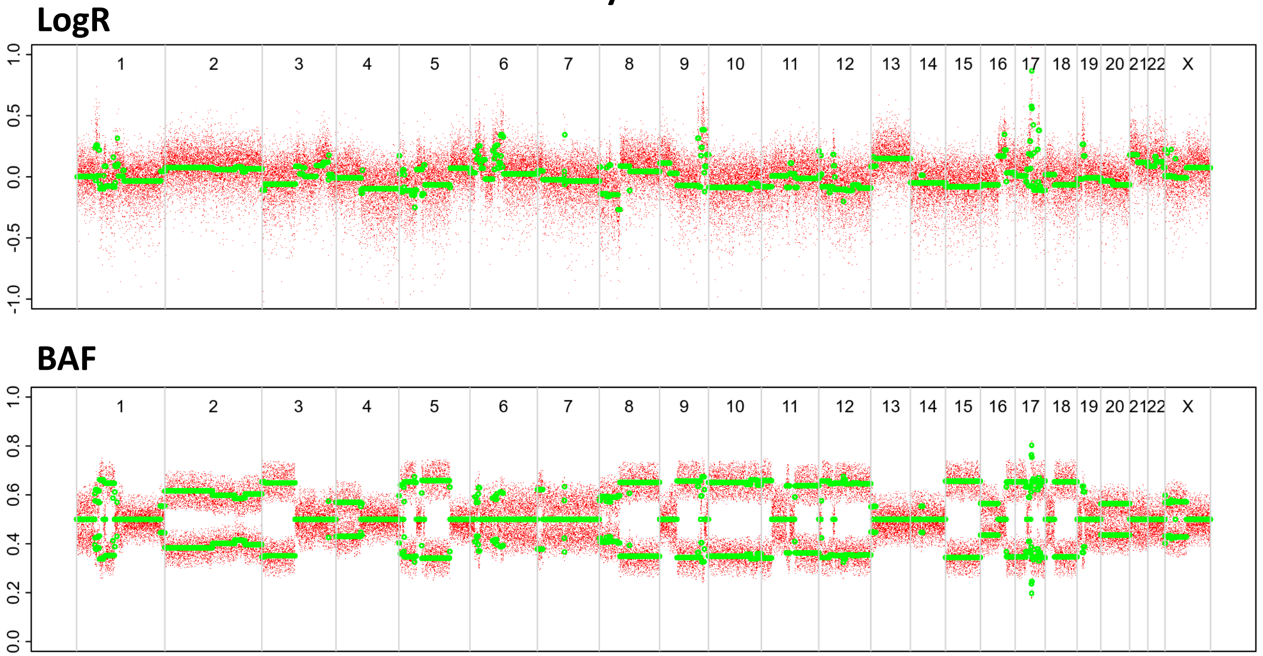


### 2nd event

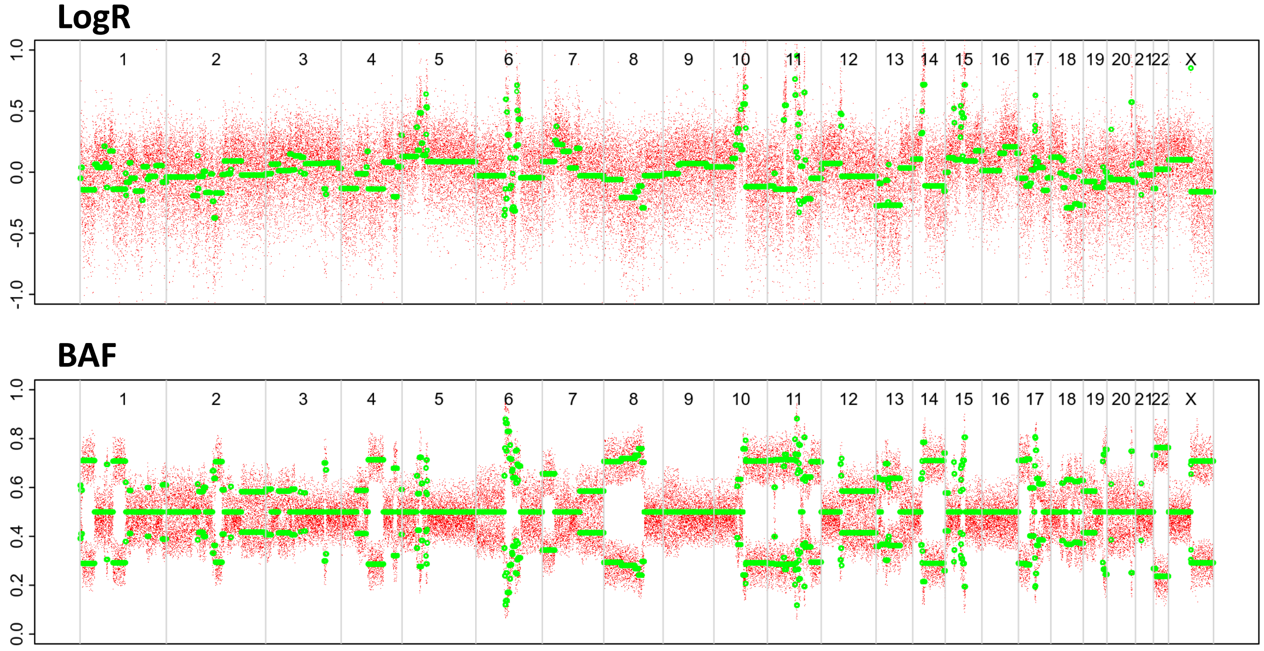

# P132

| Syn/Meta | Time from 1st surgery to 2nd event (Months) | Side 2nd event | Histology 2nd event | Surgery | AdjuvanT Treatment Pri (RT/ HT) | ER Pri | ER 2nd event | Her2 Pri | Her2 2nd event | Grade Pri | Grade 2nd event | Quadrant 2nd event | Margins | Screening | Clonality P value Copy N | Clonality P value Panel seq | Clonality P value WES | Final verdict |
|---|---|---|---|---|---|---|---|---|---|---|---|---|---|---|---|---|---|---|
| metachronous | 29 | Contralateral | IDC no DCIS | lumpectomy | RTHT | + | NA | NA | NA | 2 | 2 | NA | Clear | screen-detected | 0.115151515 | 1 | NA | Unrelated |

### Primary event

LogR

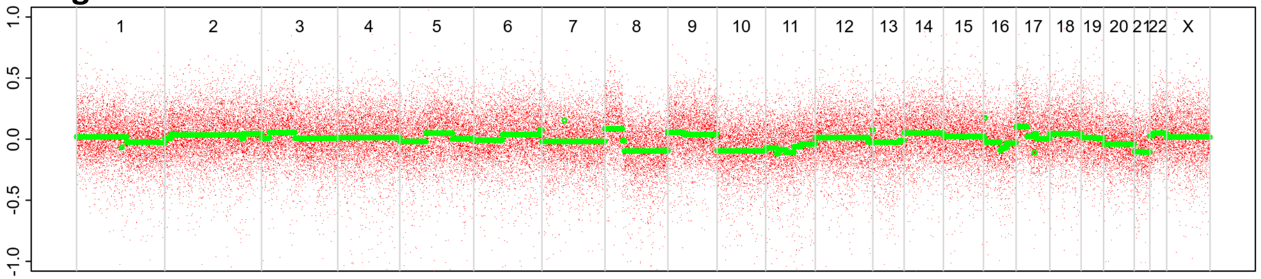


BAF

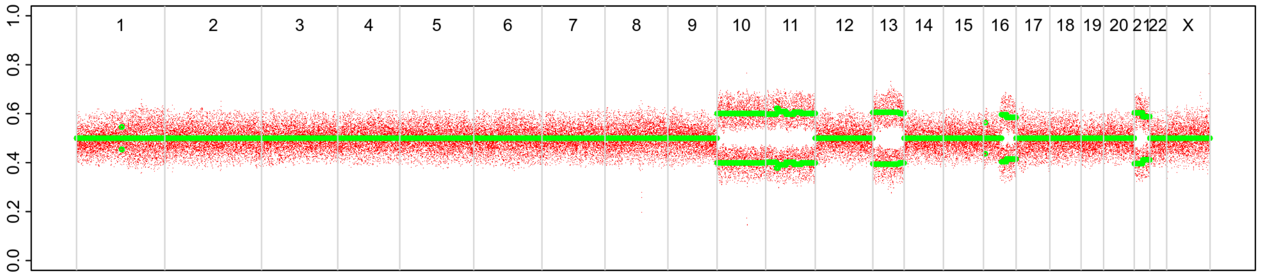


### 2nd event

LogR

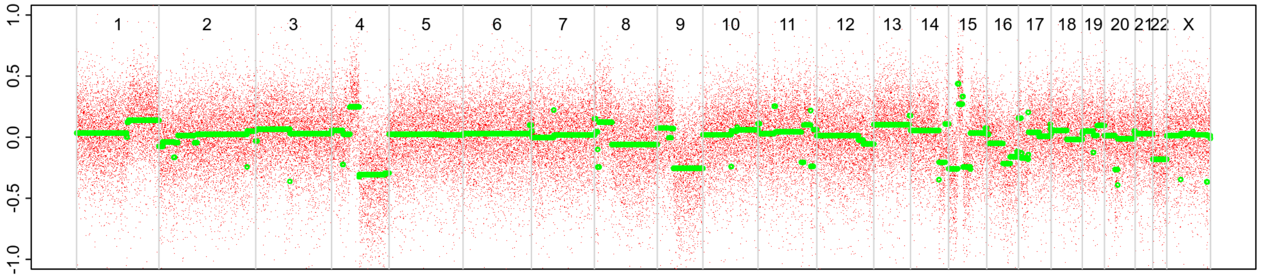


BAF

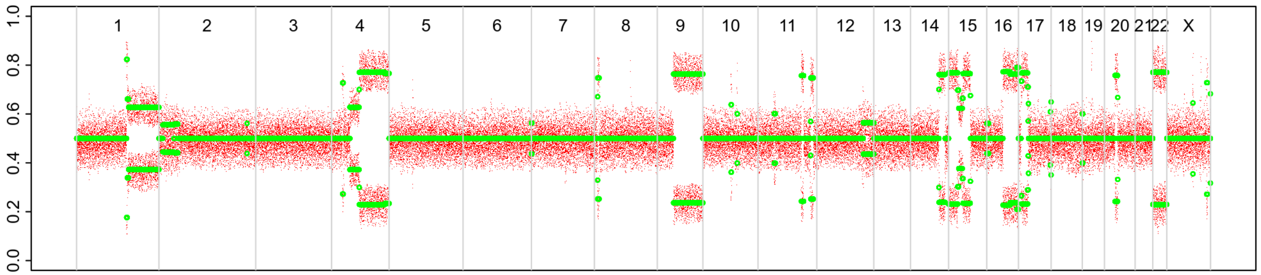

# P133

| Syn/Meta | Time from 1st surgery to 2nd event (Months) | Side 2nd event | Histology 2nd event | Surgery | AdjuvanT Treatment Pri (RT/ HT) | ER Pri | ER 2nd event | Her2 Pri | Her2 2nd event | Grade Pri | Grade 2nd event | Quadrant 2nd event | Margins | Screening | Clonality P value Copy N | Clonality P value Panel seq | Clonality P value WES | Final verdict |
|---|---|---|---|---|---|---|---|---|---|---|---|---|---|---|---|---|---|---|
| metachronous | 63 | Contralateral | IDC no DCIS | lumpectomy | HT | + | + | NA | NA | 3 | 2 | NA | Clear | screen-detected | 0.134731935 | NA | NA | Unrelated |

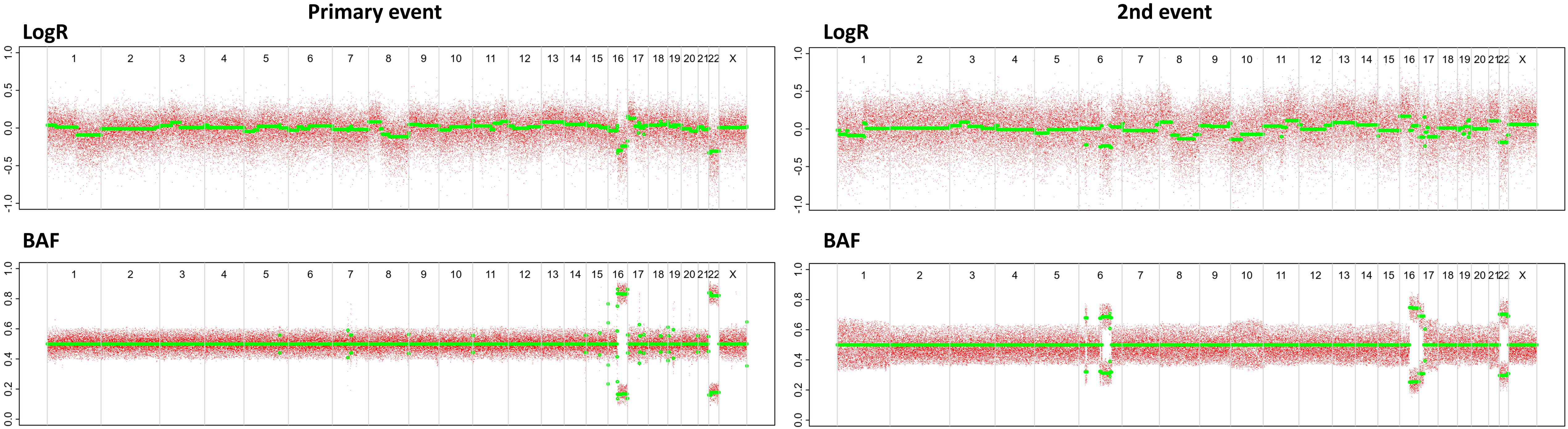

# P134

| Syn/Meta | Time from 1st surgery to 2nd event (Months) | Side 2nd event | Histology 2nd event | Surgery | AdjuvanT Treatment Pri (RT/ HT) | ER Pri | ER 2nd event | Her2 Pri | Her2 2nd event | Grade Pri | Grade 2nd event | Quadrant 2nd event | Margins | Screening | Clonality P value Copy N | Clonality P value Panel seq | Clonality P value WES | Final verdict |
|---|---|---|---|---|---|---|---|---|---|---|---|---|---|---|---|---|---|---|
| metachronous | 61 | Contralateral | IDC no DCIS | lumpectomy | RT | - | + | NA | NA | 3 | 2 | NA | Clear | screen-detected | 0.216317016 | NA | NA | Unrelated |

### Primary event

LogR

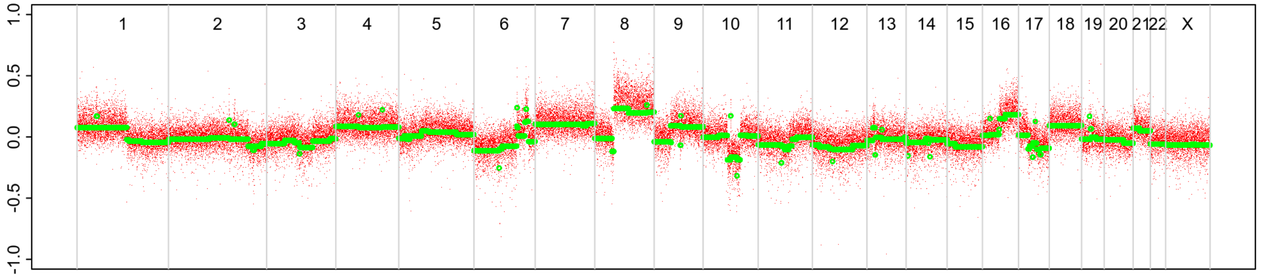


BAF

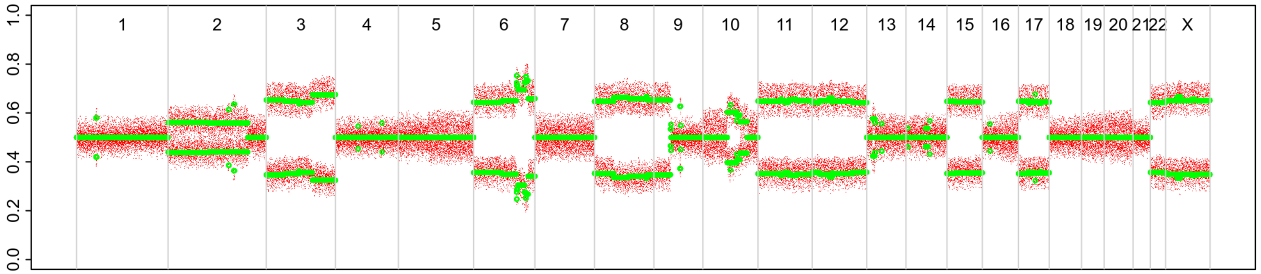


### 2nd event

LogR

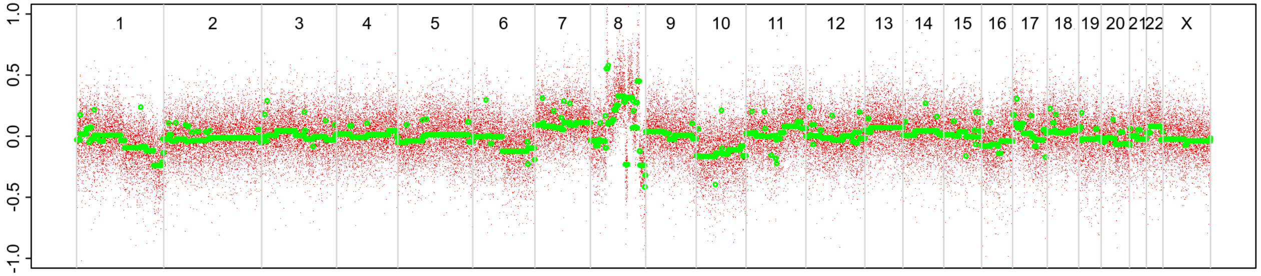


BAF

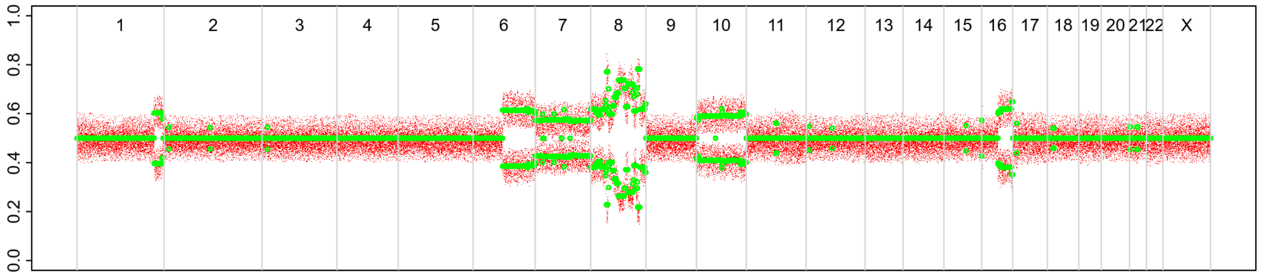

# P135

| Syn/Meta | Time from 1st surgery to 2nd event (Months) | Side 2nd event | Histology 2nd event | Surgery | AdjuvanT Treatment Pri (RT/ HT) | ER Pri | ER 2nd event | Her2 Pri | Her2 2nd event | Grade Pri | Grade 2nd event | Quadrant 2nd event | Margins | Screening | Clonality P value Copy N | Clonality P value Panel seq | Clonality P value WES | Final verdict |
|---|---|---|---|---|---|---|---|---|---|---|---|---|---|---|---|---|---|---|
| metachronous | 34 | Contralateral | IDC no DCIS | lumpectomy | RTHT | + | NA | - | NA | 3 | 2 | NA | Clear | screen-detected | 0.244755245 | NA | NA | Unrelated |

### Primary event

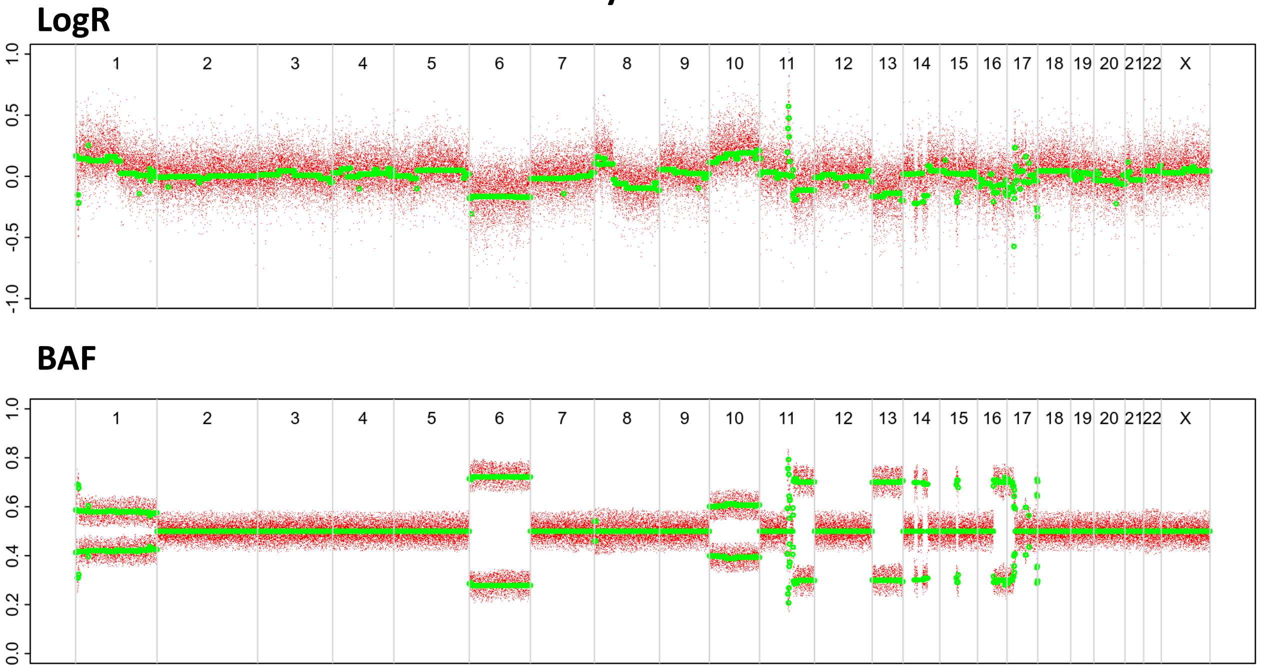


### 2nd event

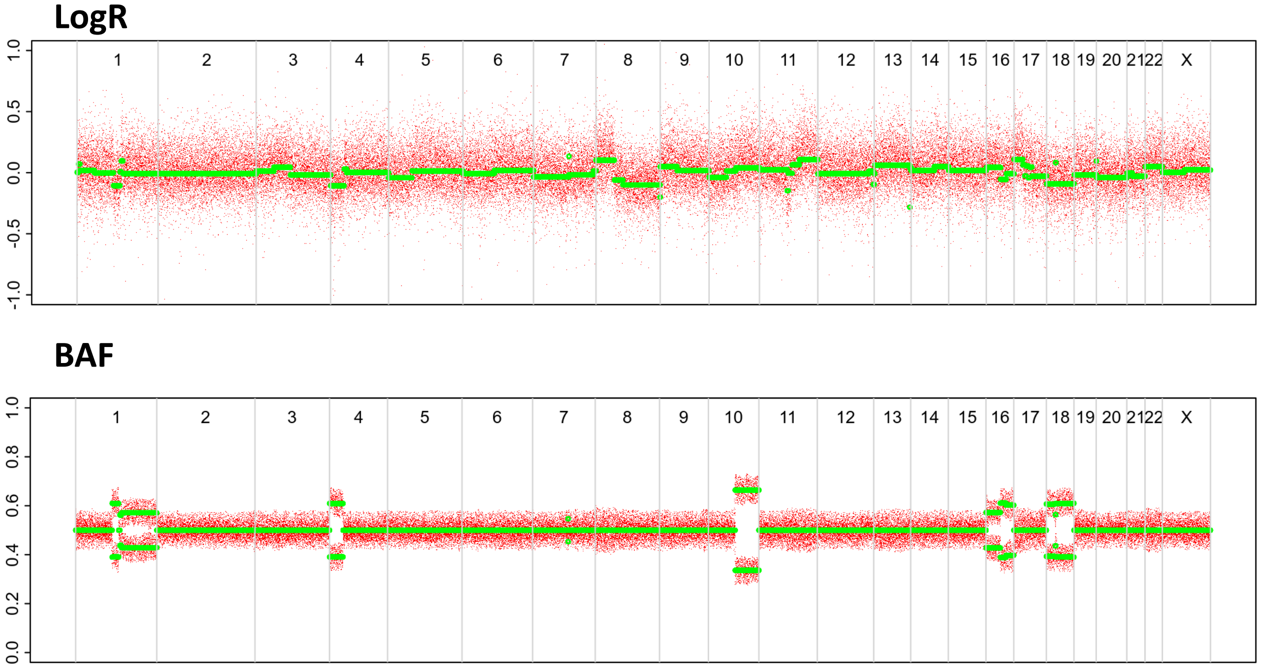

# P136

| Syn/Meta | Time from 1st surgery to 2nd event (Months) | Side 2nd event | Histology 2nd event | Surgery | AdjuvanT Treatment Pri (RT/ HT) | ER Pri | ER 2nd event | Her2 Pri | Her2 2nd event | Grade Pri | Grade 2nd event | Quadrant 2nd event | Margins | Screening | Clonality P value Copy N | Clonality P value Panel seq | Clonality P value WES | Final verdict |
|---|---|---|---|---|---|---|---|---|---|---|---|---|---|---|---|---|---|---|
| metachronous | 20 | Contralateral | IDC no DCIS | lumpectomy | RT | - | + | + | NA | 3 | 2 | NA | Clear | screen-detected | 0.305361305 | 0.16 | NA | Unrelated |

### Primary event

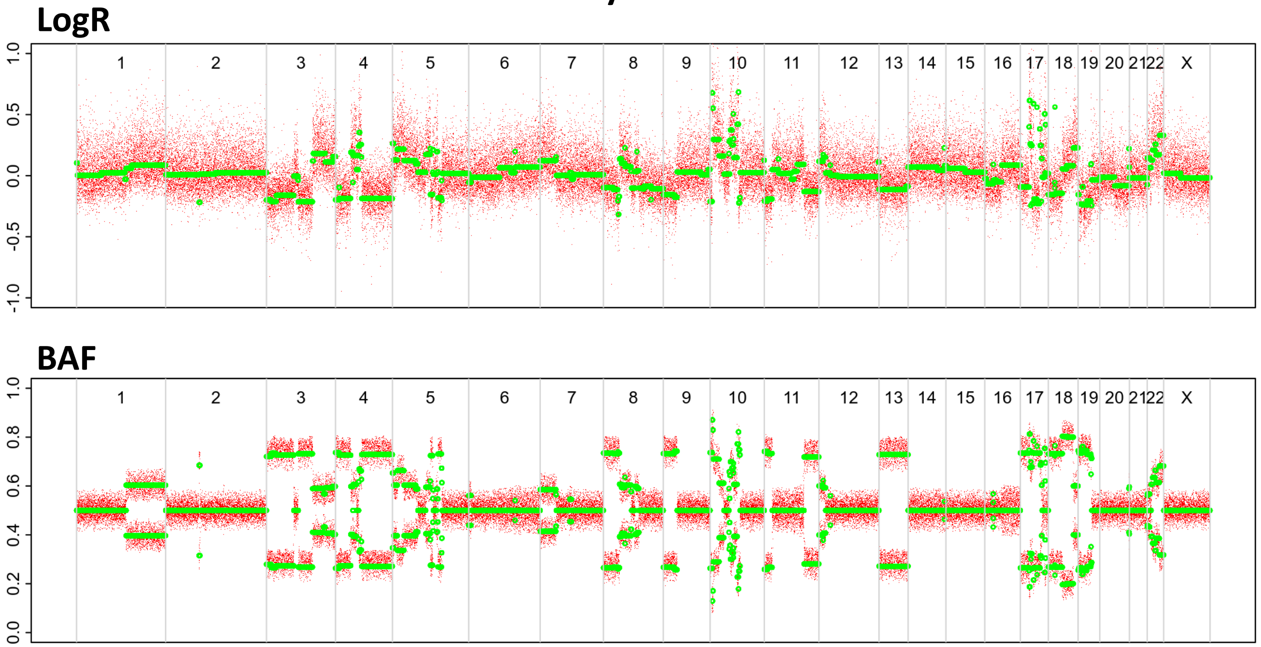


### 2nd event

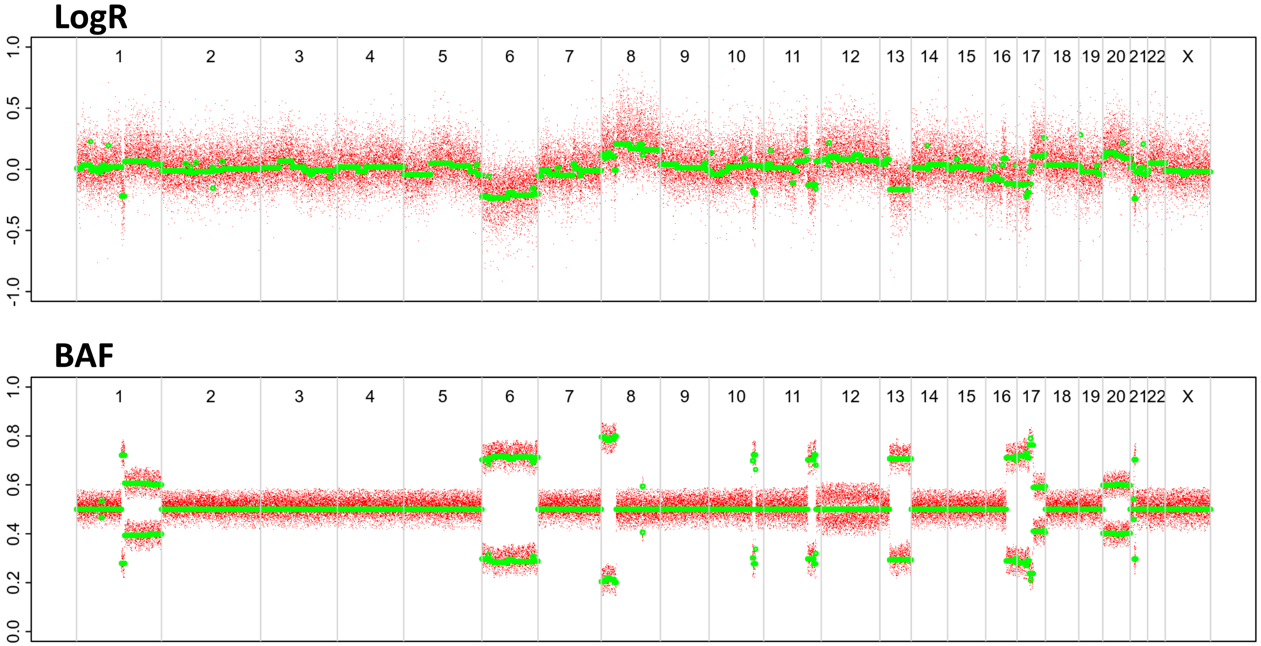

# P137

| Syn/Meta | Time from 1st surgery to 2nd event (Months) | Side 2nd event | Histology 2nd event | Surgery | AdjuvanT Treatment Pri (RT/ HT) | ER Pri | ER 2nd event | Her2 Pri | Her2 2nd event | Grade Pri | Grade 2nd event | Quadrant 2nd event | Margins | Screening | Clonality P value Copy N | Clonality P value Panel seq | Clonality P value WES | Final verdict |
|---|---|---|---|---|---|---|---|---|---|---|---|---|---|---|---|---|---|---|
| metachronous | 111 | Contralateral | IDC no DCIS | lumpectomy | None | + | NA | NA | NA | 3 | 3 | NA | Clear | screen-detected | 0.373892774 | 1 | NA | Unrelated |

### Primary event

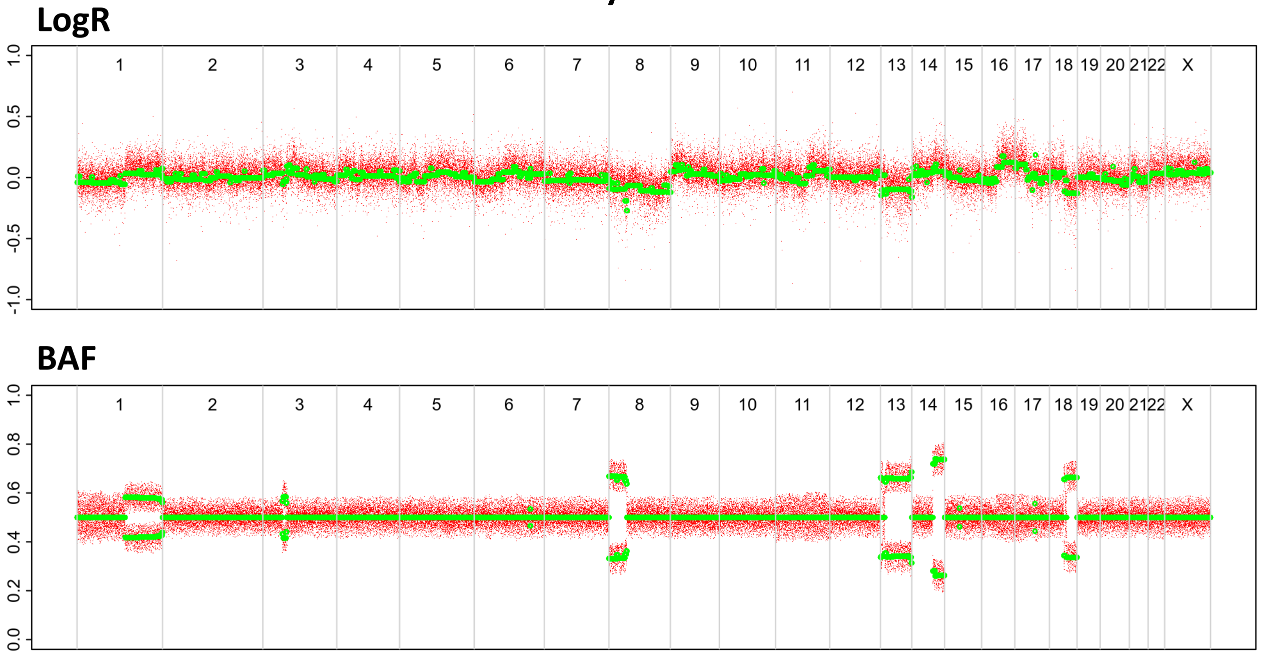


### 2nd event

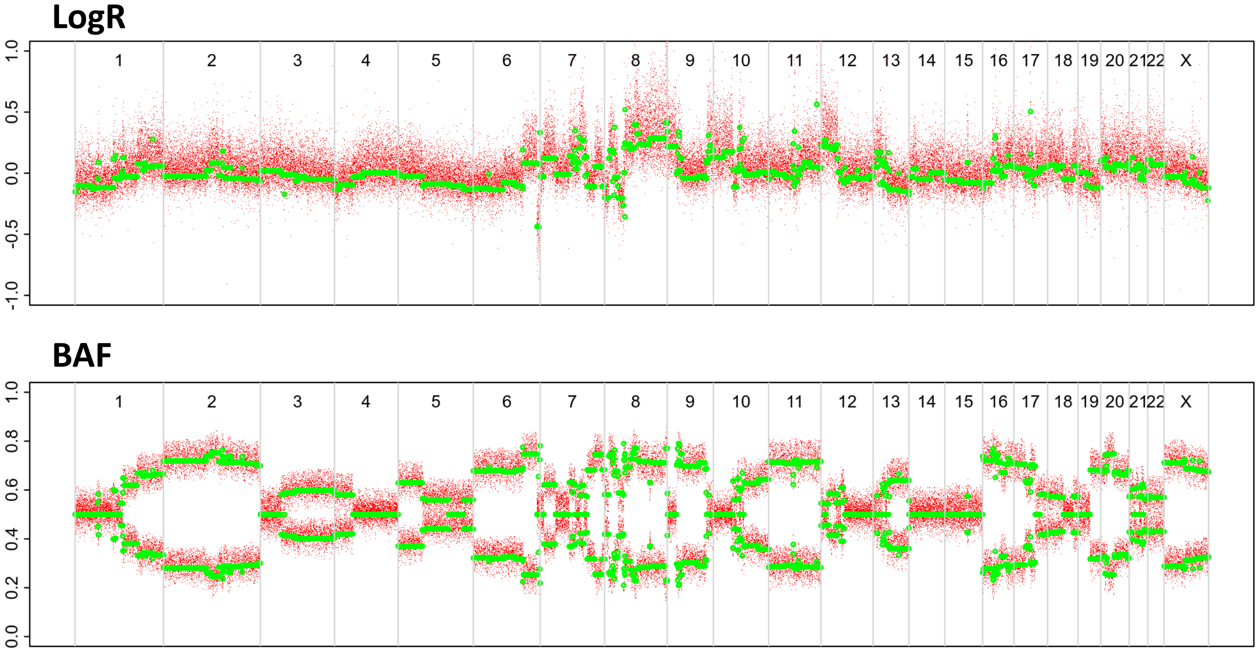

# P138

| Syn/Meta | Time from 1st surgery to 2nd event (Months) | Side 2nd event | Histology 2nd event | Surgery | AdjuvanT Treatment Pri (RT/ HT) | ER Pri | ER 2nd event | Her2 Pri | Her2 2nd event | Grade Pri | Grade 2nd event | Quadrant 2nd event | Margins | Screening | Clonality P value Copy N | Clonality P value Panel seq | Clonality P value WES | Final verdict |
|---|---|---|---|---|---|---|---|---|---|---|---|---|---|---|---|---|---|---|
| metachronous | 27 | Contralateral | IDC with DCIS | lumpectomy | None | + | NA | - | - | 2 | 3 | NA | Clear | screen-detected | 0.436363636 | 1 | NA | Unrelated |

### Primary event

LogR

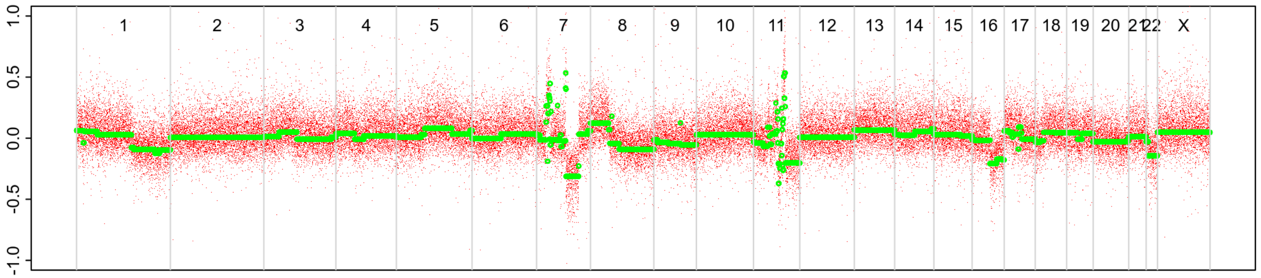


BAF

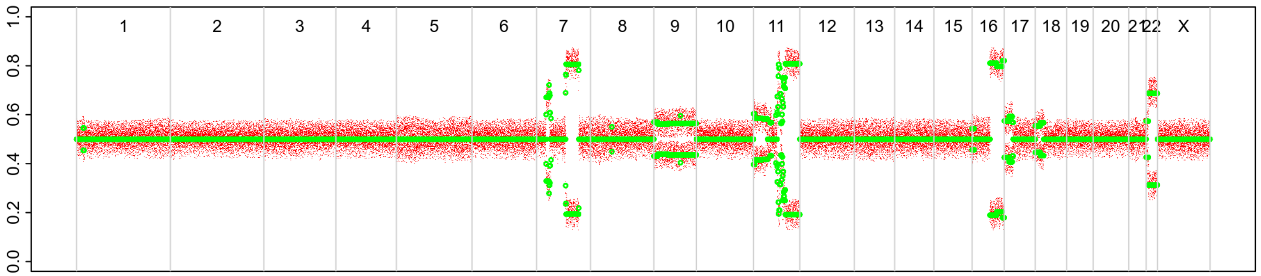


### 2nd event

LogR

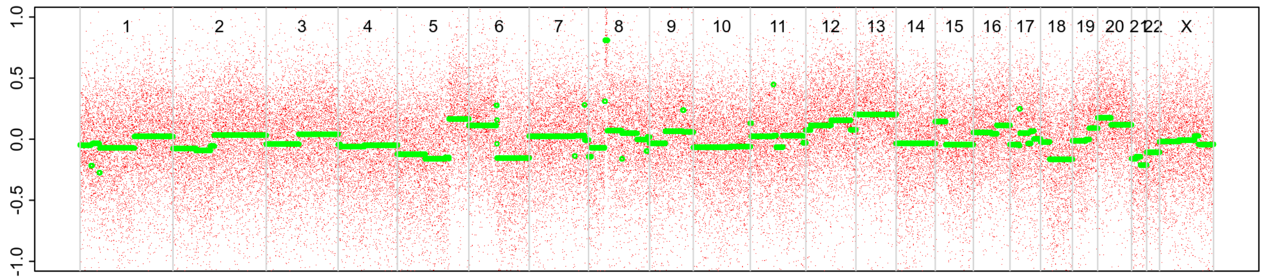


BAF

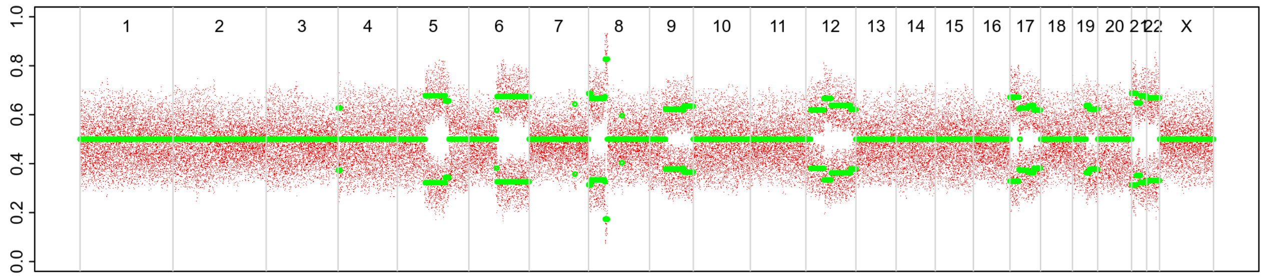

# P139

| Syn/Meta | Time from 1st surgery to 2nd event (Months) | Side 2nd event | Histology 2nd event | Surgery | AdjuvanT Treatment Pri (RT/ HT) | ER Pri | ER 2nd event | Her2 Pri | Her2 2nd event | Grade Pri | Grade 2nd event | Quadrant 2nd event | Margins | Screening | Clonality P value Copy N | Clonality P value Panel seq | Clonality P value WES | Final verdict |
|---|---|---|---|---|---|---|---|---|---|---|---|---|---|---|---|---|---|---|
| metachronous | 56 | Contralateral | IDC with DCIS | lumpectomy | RT | + | + | - | - | 3 | 3 | NA | Clear | screen-detected | 0.468997669 | 0.14 | NA | Unrelated |

### Primary event

**LogR**

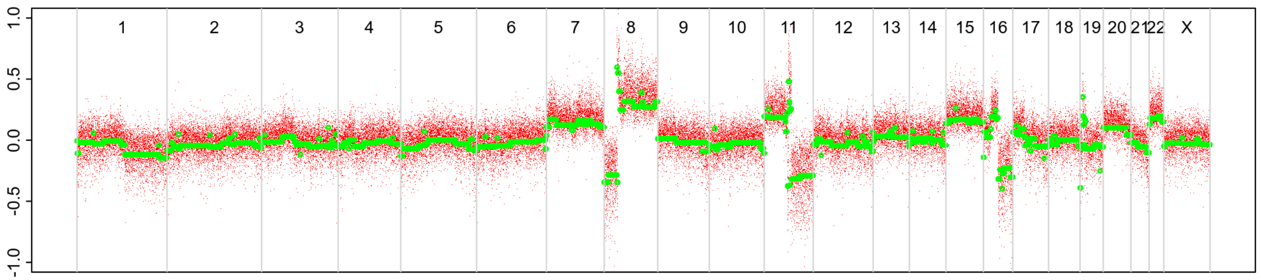


**BAF**

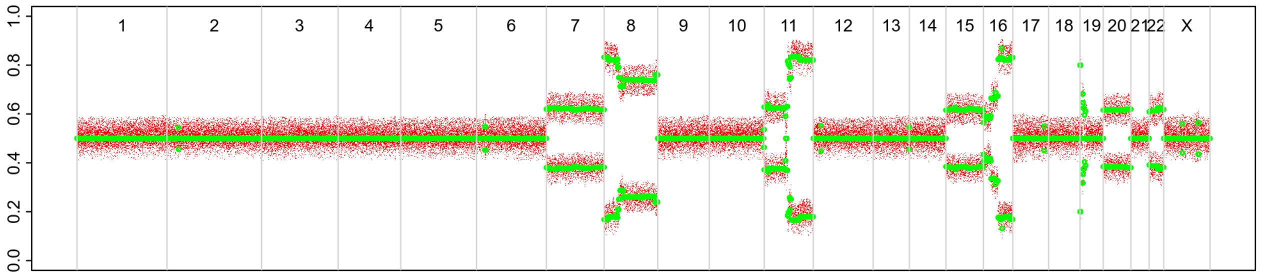


### 2nd event

**LogR**

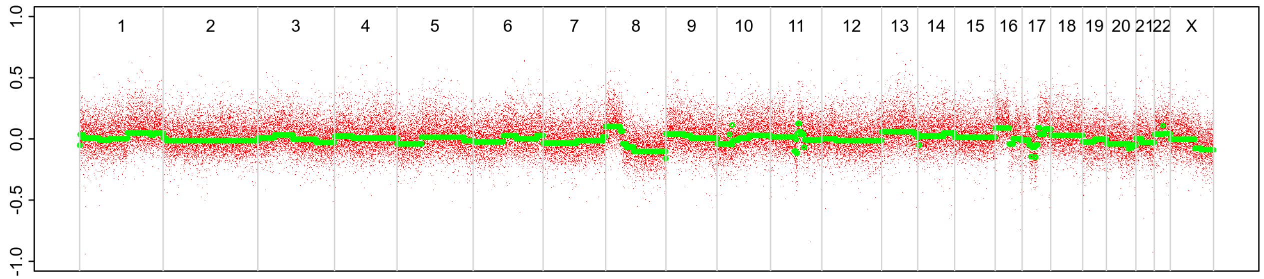


**BAF**

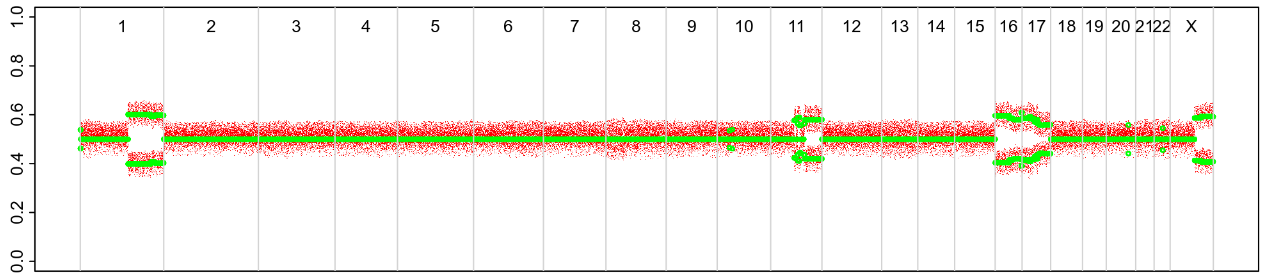

# P140

| Syn/Meta | Time from 1st surgery to 2nd event (Months) | Side 2nd event | Histology 2nd event | Surgery | AdjuvanT Treatment Pri (RT/ HT) | ER Pri | ER 2nd event | Her2 Pri | Her2 2nd event | Grade Pri | Grade 2nd event | Quadrant 2nd event | Margins | Screening | Clonality P value Copy N | Clonality P value Panel seq | Clonality P value WES | Final verdict |
|---|---|---|---|---|---|---|---|---|---|---|---|---|---|---|---|---|---|---|
| metachronous | 12 | Contralateral | IDC no DCIS | lumpectomy | HT | + | + | NA | NA | 3 | 2 | NA | Clear | screen-detected | 0.712354312 | NA | NA | Unrelated |

### Primary event

LogR

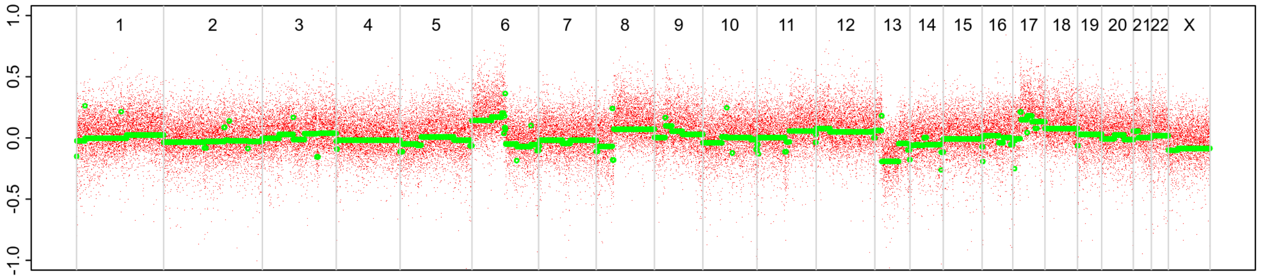


BAF

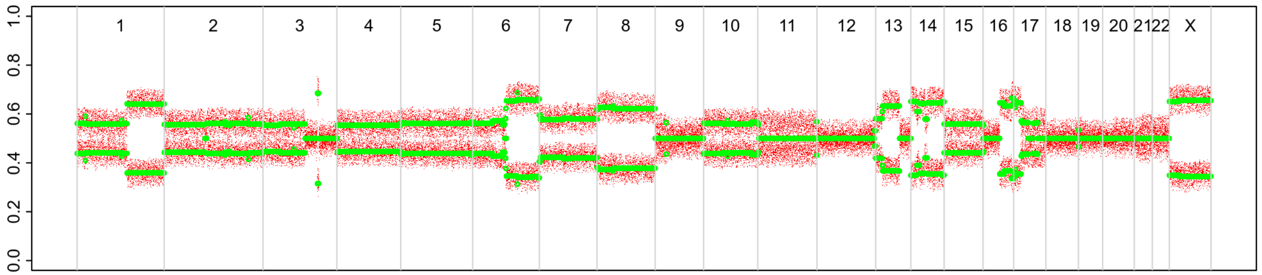


### 2nd event

LogR

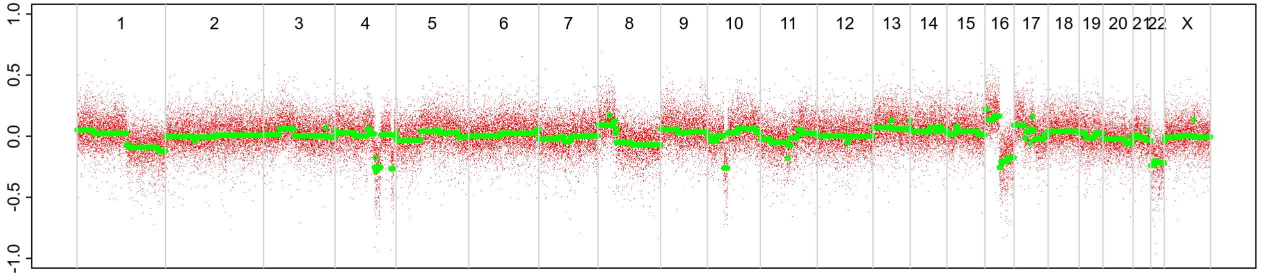


BAF

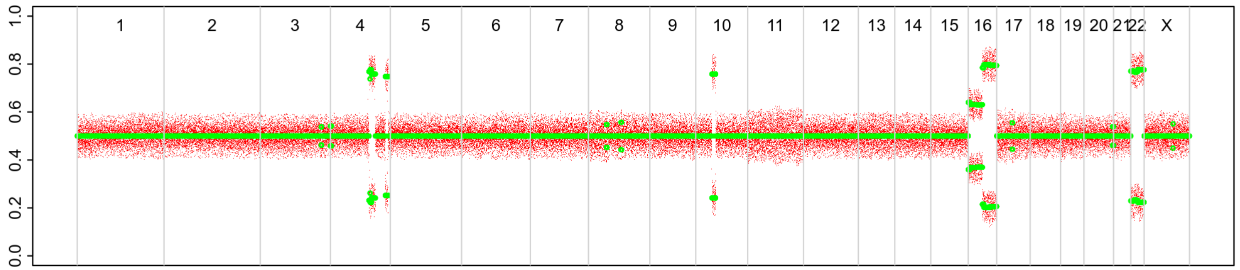

# P141

| Syn/Meta | Time from 1st surgery to 2nd event (Months) | Side 2nd event | Histology 2nd event | Surgery | AdjuvanT Treatment Pri (RT/ HT) | ER Pri | ER 2nd event | Her2 Pri | Her2 2nd event | Grade Pri | Grade 2nd event | Quadrant 2nd event | Margins | Screening | Clonality P value Copy N | Clonality P value Panel seq | Clonality P value WES | Final verdict |
|---|---|---|---|---|---|---|---|---|---|---|---|---|---|---|---|---|---|---|
| metachronous | 79 | Contralateral | IDC with DCIS | lumpectomy | RT | - | NA | NA | NA | 3 | 3 | NA | Clear | screen-detected | 0.722610723 | NA | NA | Unrelated |

### Primary event

LogR

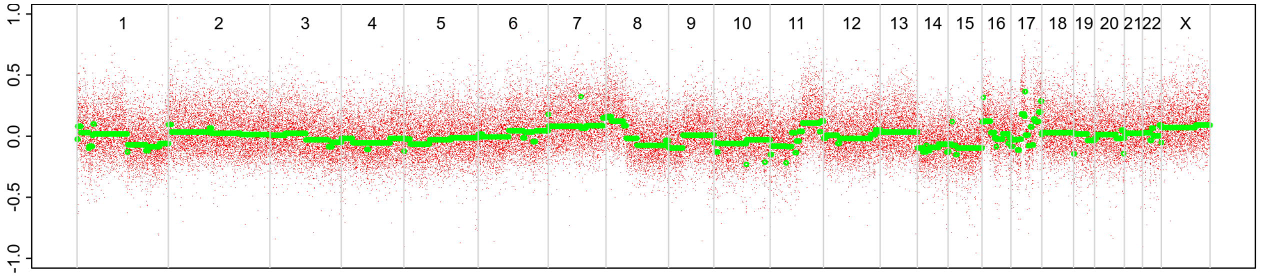


BAF

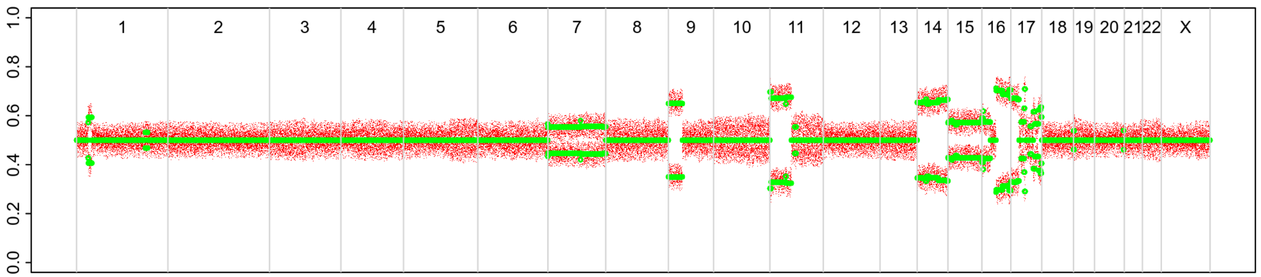


### 2nd event

LogR

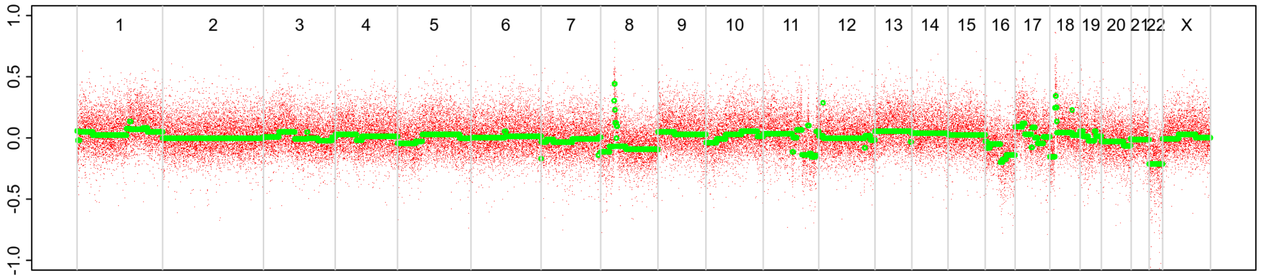


BAF

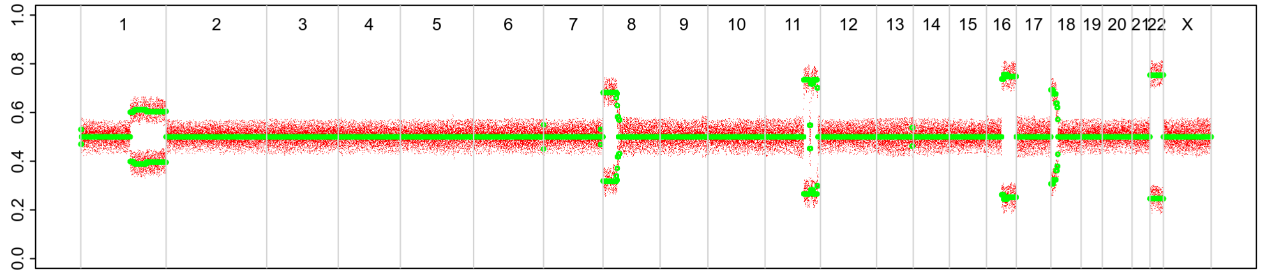

# P142

| Syn/Meta | Time from 1st surgery to 2nd event (Months) | Side 2nd event | Histology 2nd event | Surgery | AdjuvanT Treatment Pri (RT/ HT) | ER Pri | ER 2nd event | Her2 Pri | Her2 2nd event | Grade Pri | Grade 2nd event | Quadrant 2nd event | Margins | Screening | Clonality P value Copy N | Clonality P value Panel seq | Clonality P value WES | Final verdict |
|---|---|---|---|---|---|---|---|---|---|---|---|---|---|---|---|---|---|---|
| metachronous | 26 | Contralateral | IDC no DCIS | lumpectomy | None | - | + | - | NA | 2 | 1 | NA | Clear | screen-detected | 1 | 0.16 | NA | Unrelated |

Primary event

2nd event

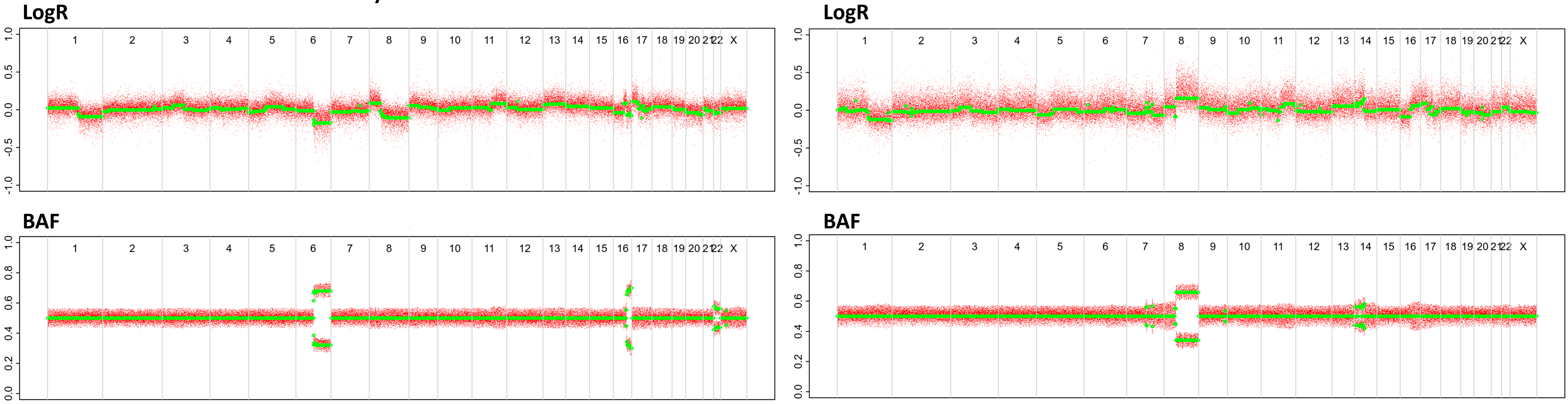
