## Supplementary file 1B for "Genomic profiling defines variable clonal relatedness between invasive breast cancer and primary ductal carcinoma *in situ*"

# P031

| Syn/Meta | Time from 1st surgery to 2nd event (Months) | Side 2nd event | Histology 2nd event | Surgery | AdjuvanT Treatment Pri (RT/ HT) | ER Pri | ER 2nd event | Her2 Pri | Her2 2nd event | Grade Pri | Grade 2nd event | Quadrant 2nd event | Margins | Screening | Clonality P value Copy N | Clonality P value Panel seq | Clonality P value WES | Final verdict |
|---|---|---|---|---|---|---|---|---|---|---|---|---|---|---|---|---|---|---|
| metachronous | 167 | Ipsilateral | Invasive | lumpectomy | None | + | + | - | - | 2 | 2 | NA | involved | screen-detected | 0.000649 14 | NA | NA | Related |

### Primary event

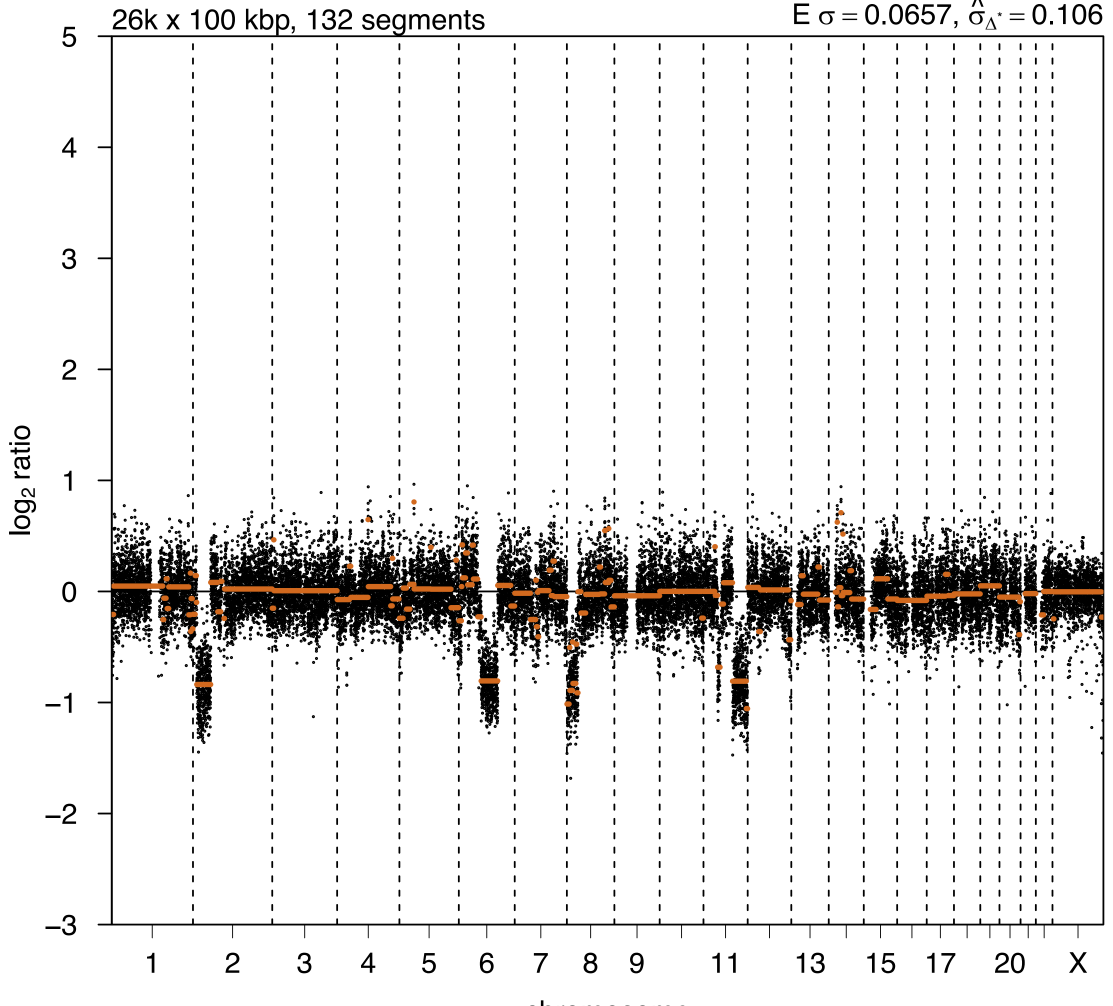


### 2nd event

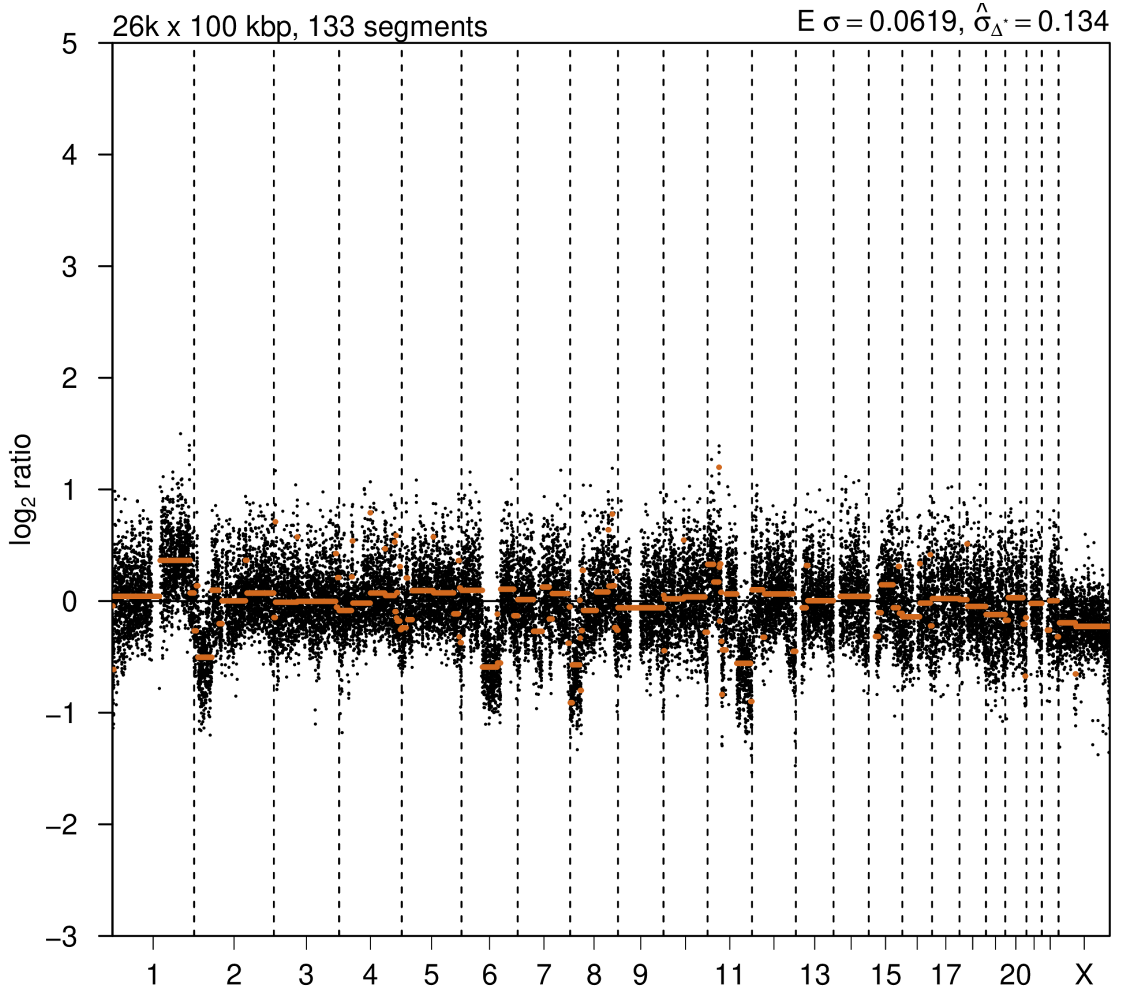

# P032

| Syn/Meta | Time from 1st surgery to 2nd event (Months) | Side 2nd event | Histology 2nd event | Surgery | AdjuvanT Treatment Pri (RT/ HT) | ER Pri | ER 2nd event | Her2 Pri | Her2 2nd event | Grade Pri | Grade 2nd event | Quadrant 2nd event | Margins | Screening | Clonality P value Copy N | Clonality P value Panel seq | Clonality P value WES | Final verdict |
|---|---|---|---|---|---|---|---|---|---|---|---|---|---|---|---|---|---|---|
| metachronous | 130 | Ipsilateral | IDC with DCIS | lumpectomy | None | + | + | - | - | 2 | 3 | NA | NA | screen-detected | 0.000649 14 | 0.006 | NA | Related |

### Primary event

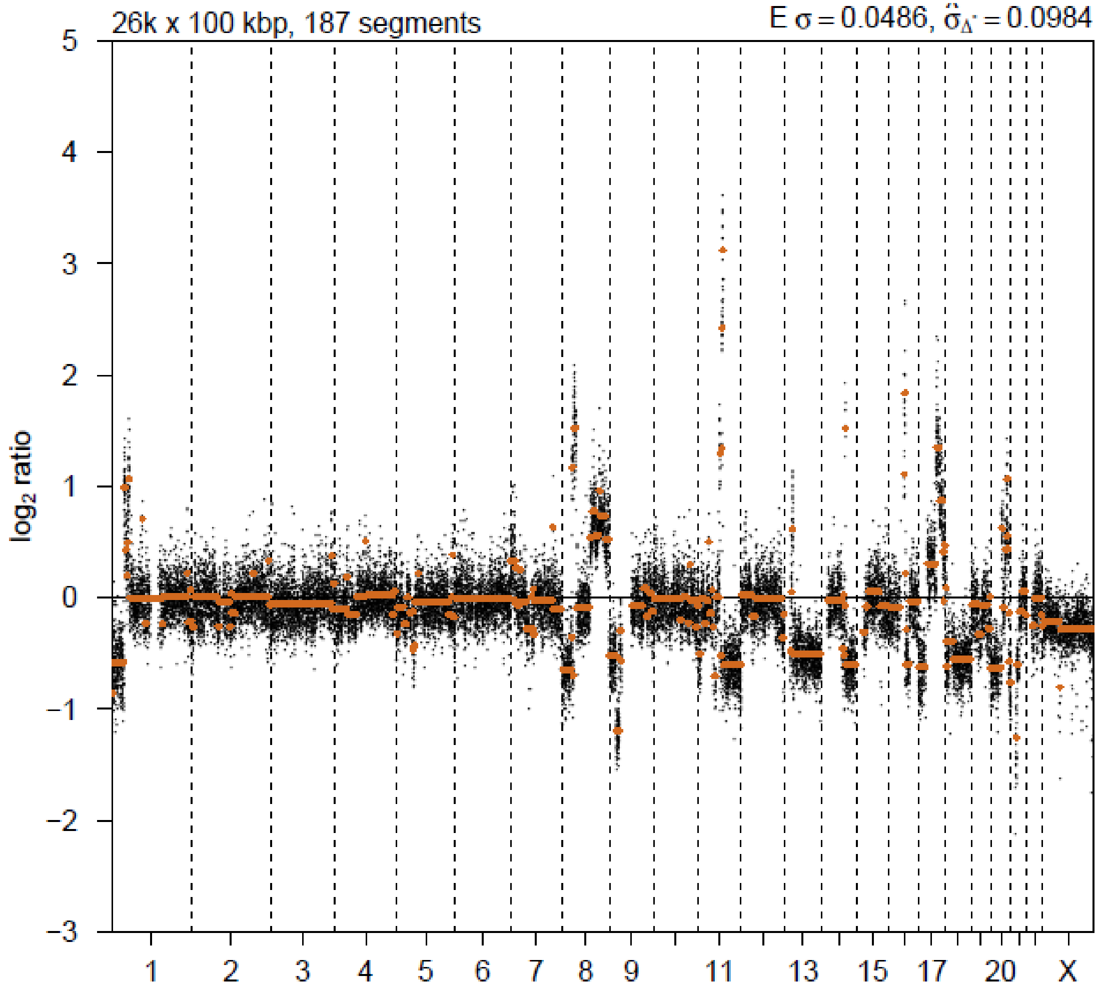


### 2nd event

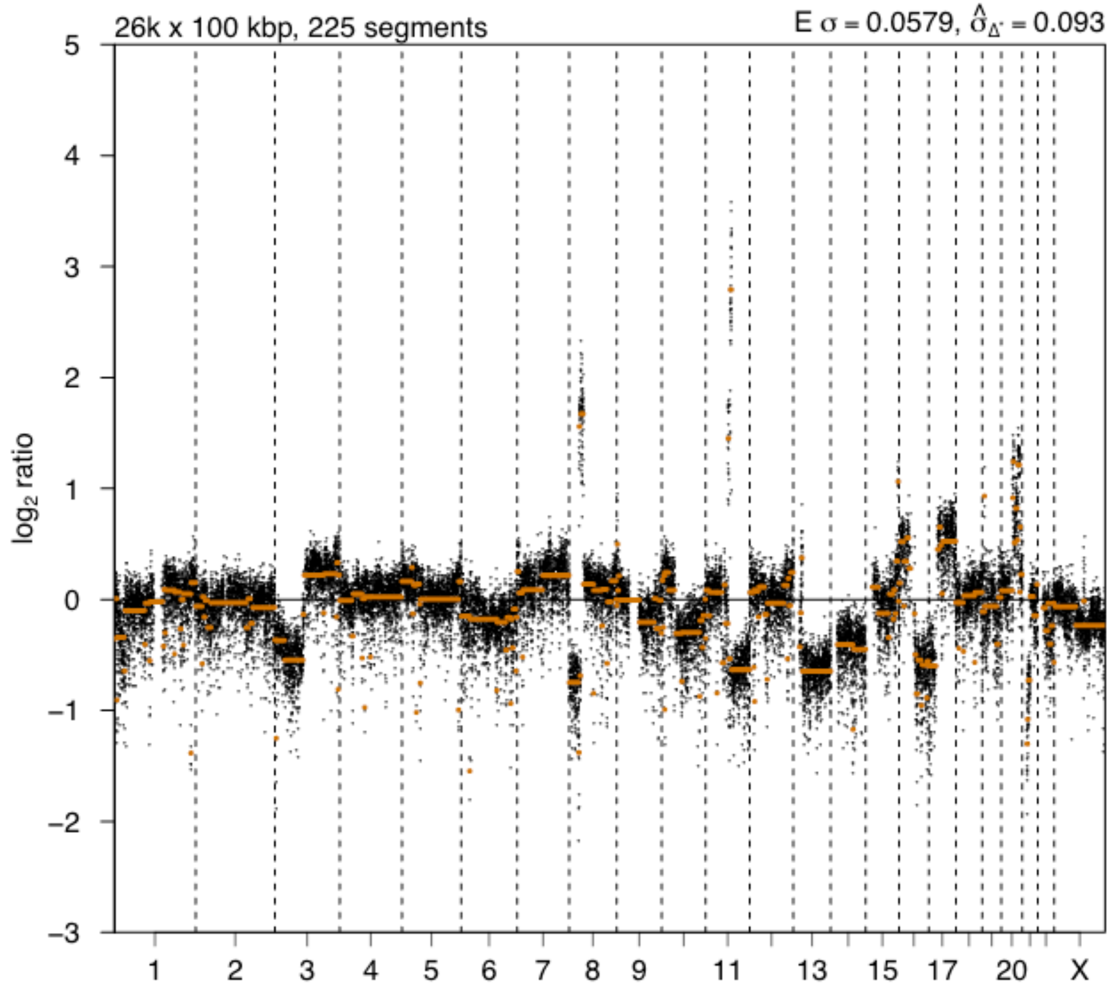

# P033

| Syn/Meta | Time from 1st surgery to 2nd event (Months) | Side 2nd event | Histology 2nd event | Surgery | AdjuvanT Treatment Pri (RT/ HT) | ER Pri | ER 2nd event | Her2 Pri | Her2 2nd event | Grade Pri | Grade 2nd event | Quadrant 2nd event | Margins | Screening | Clonality P value Copy N | Clonality P value Panel seq | Clonality P value WES | Final verdict |
|---|---|---|---|---|---|---|---|---|---|---|---|---|---|---|---|---|---|---|
| metachronous | 125 | Ipsilateral | IDC with DCIS | lumpectomy | None | + | + | - | - | 2 | 2 | NA | NA | screen-detected | 0.003894839 | 0.003 | NA | Related |

### Primary event

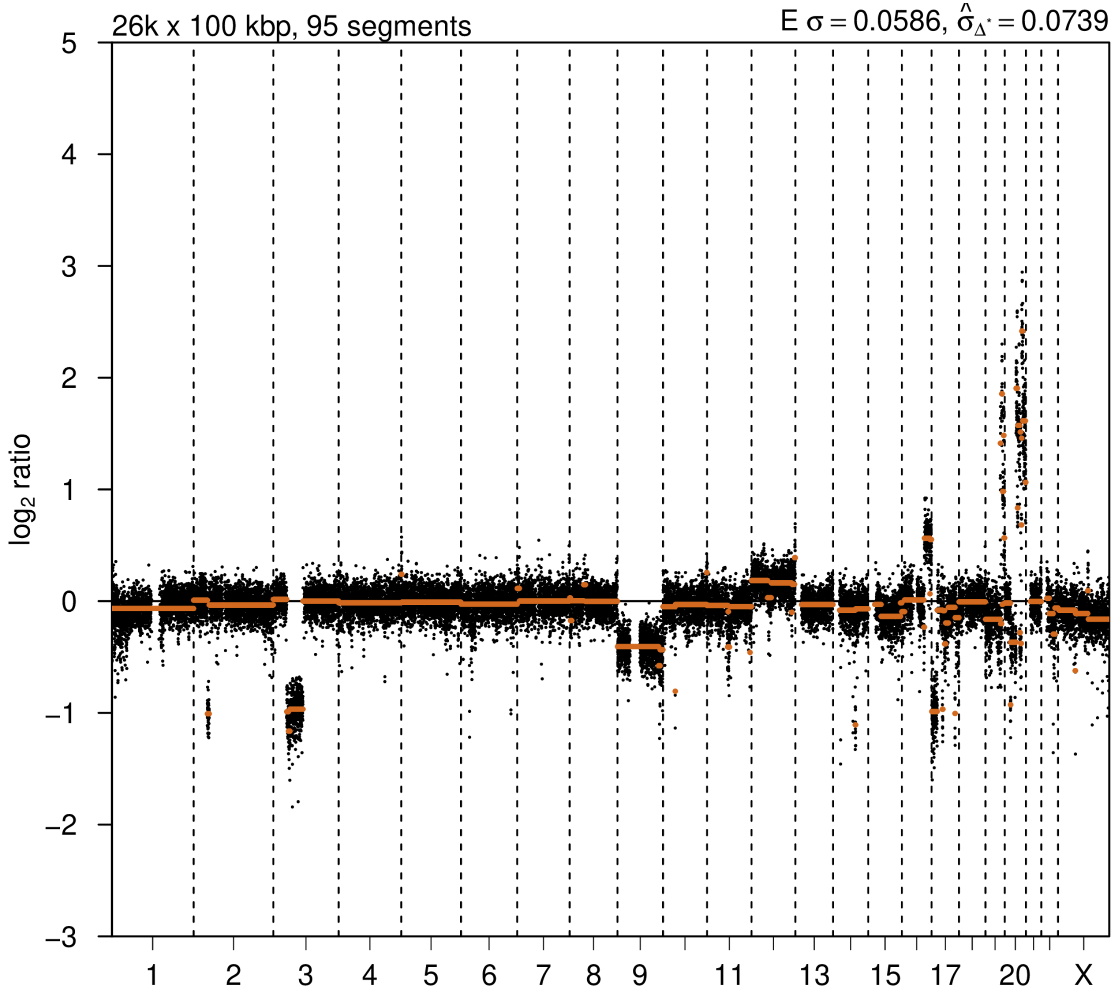


### 2nd event

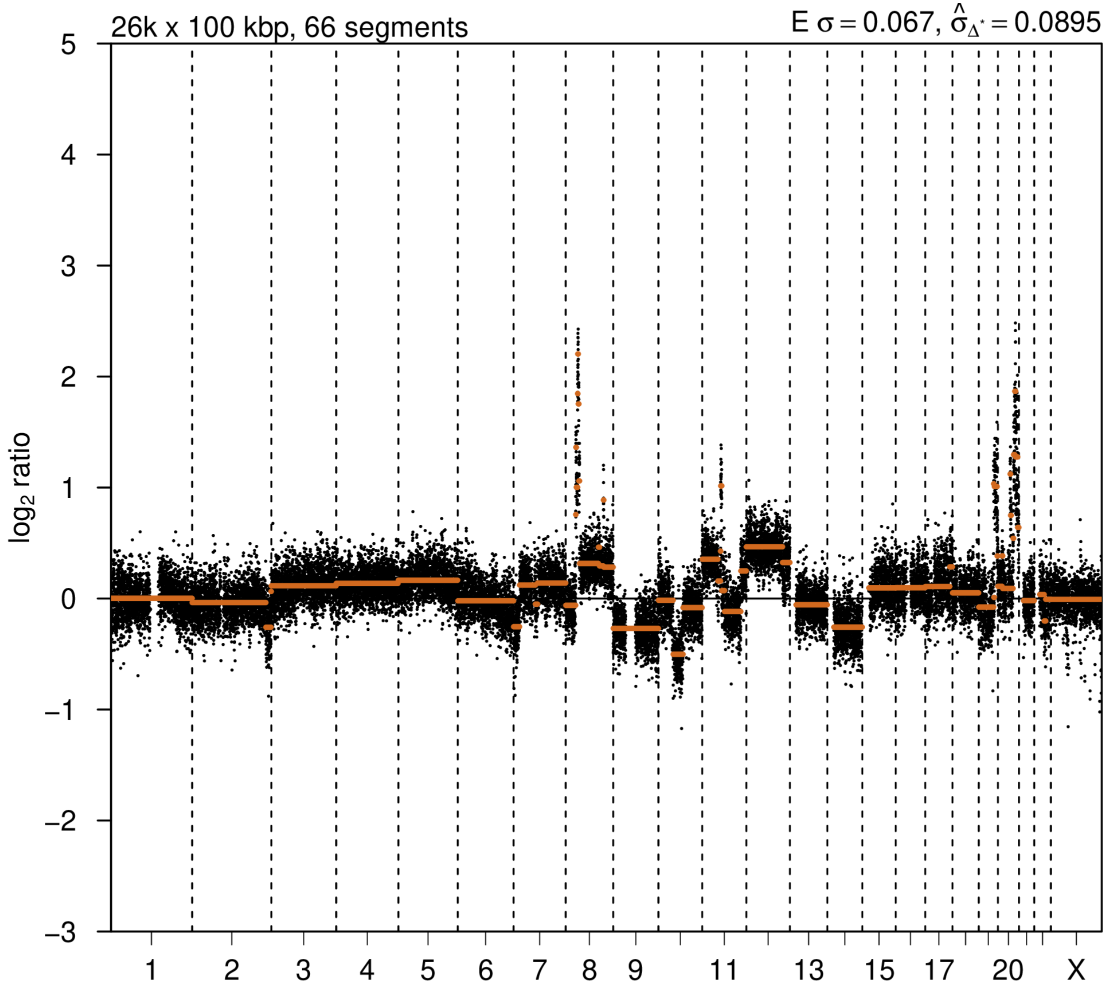

# P035

| Syn/Meta | Time from 1st surgery to 2nd event (Months) | Side 2nd event | Histology 2nd event | Surgery | AdjuvanT Treatment Pri (RT/ HT) | ER Pri | ER 2nd event | Her2 Pri | Her2 2nd event | Grade Pri | Grade 2nd event | Quadrant 2nd event | Margins | Screening | Clonality P value Copy N | Clonality P value Panel seq | Clonality P value WES | Final verdict |
|---|---|---|---|---|---|---|---|---|---|---|---|---|---|---|---|---|---|---|
| metachronous | 188 | Ipsilateral | Invasive | lumpectomy | None | NA | - | NA | + | 3 | 3 | NA | NA | symptomatic | 0.027588445 | NA | 0.000752445 | Related |

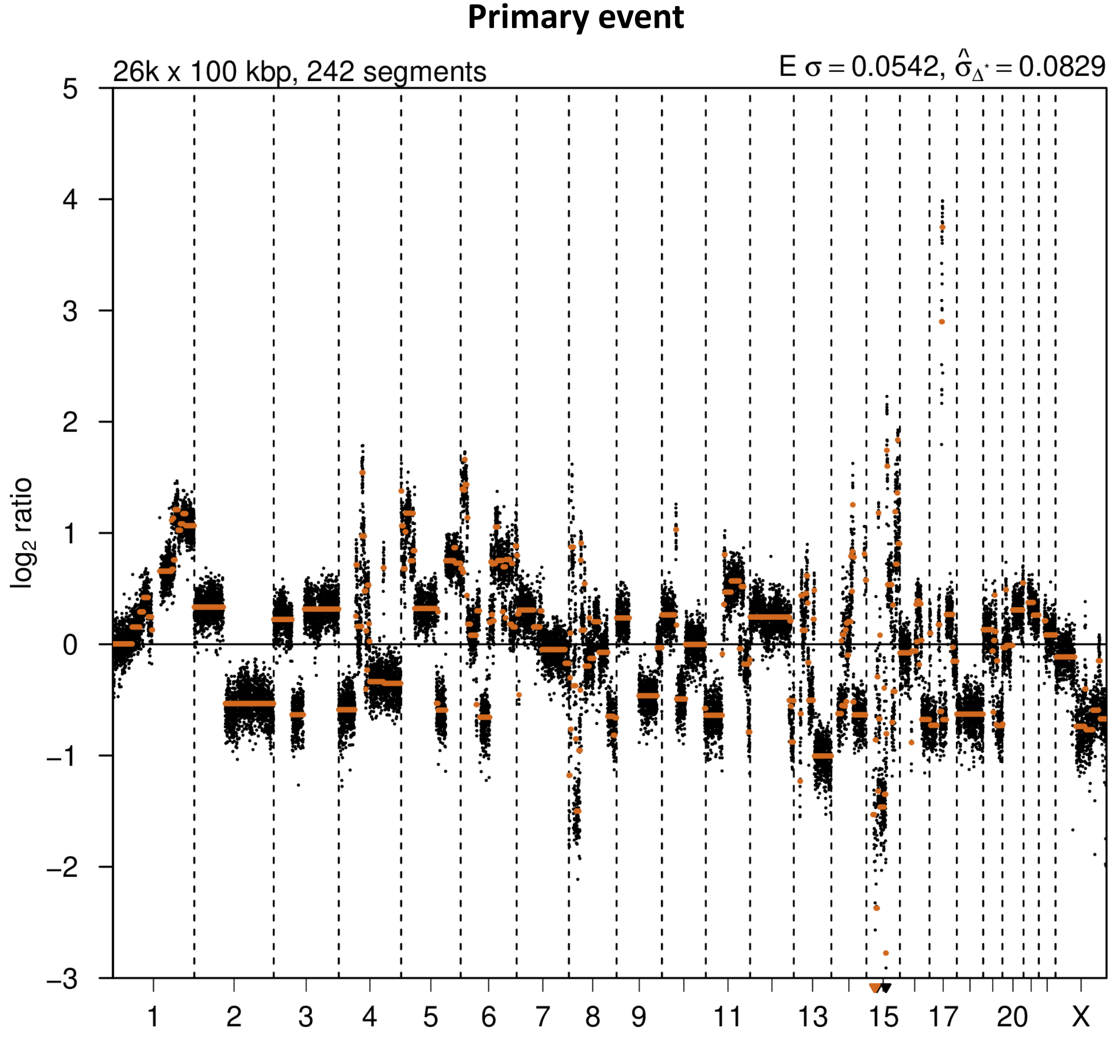


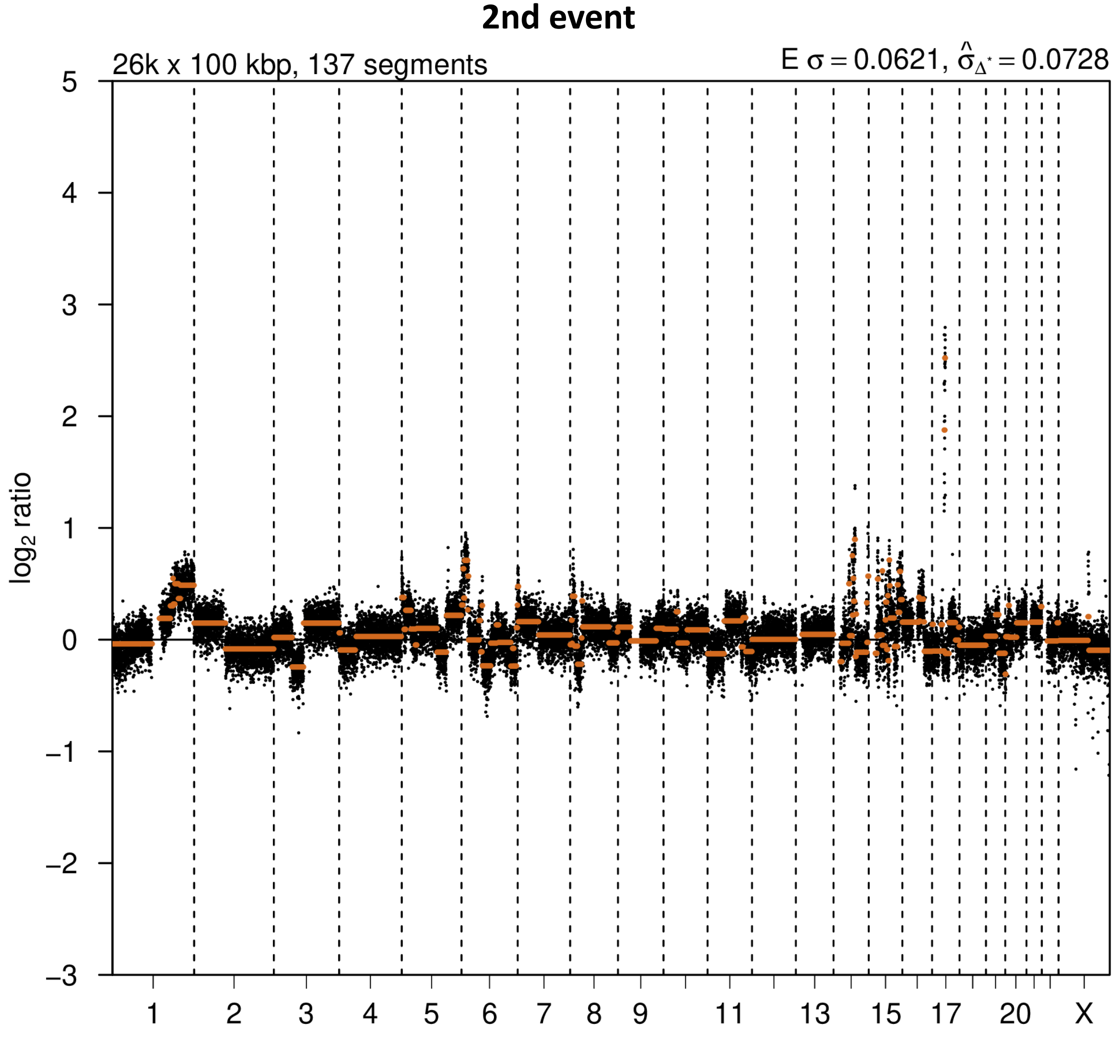

# P036

| Syn/Meta | Time from 1st surgery to 2nd event (Months) | Side 2nd event | Histology 2nd event | Surgery | AdjuvanT Treatment Pri (RT/ HT) | ER Pri | ER 2nd event | Her2 Pri | Her2 2nd event | Grade Pri | Grade 2nd event | Quadrant 2nd event | Margins | Screening | Clonality P value Copy N | Clonality P value Panel seq | Clonality P value WES | Final verdict |
|---|---|---|---|---|---|---|---|---|---|---|---|---|---|---|---|---|---|---|
| metachronous | 125 | Ipsilateral | IDC with DCIS | lumpectomy | None | + | + | - | - | 2 | 2 | at or adjacent to primary | Clear | screen-detected | 0.000324 57 | 0.003 | NA | Related |

### Primary event

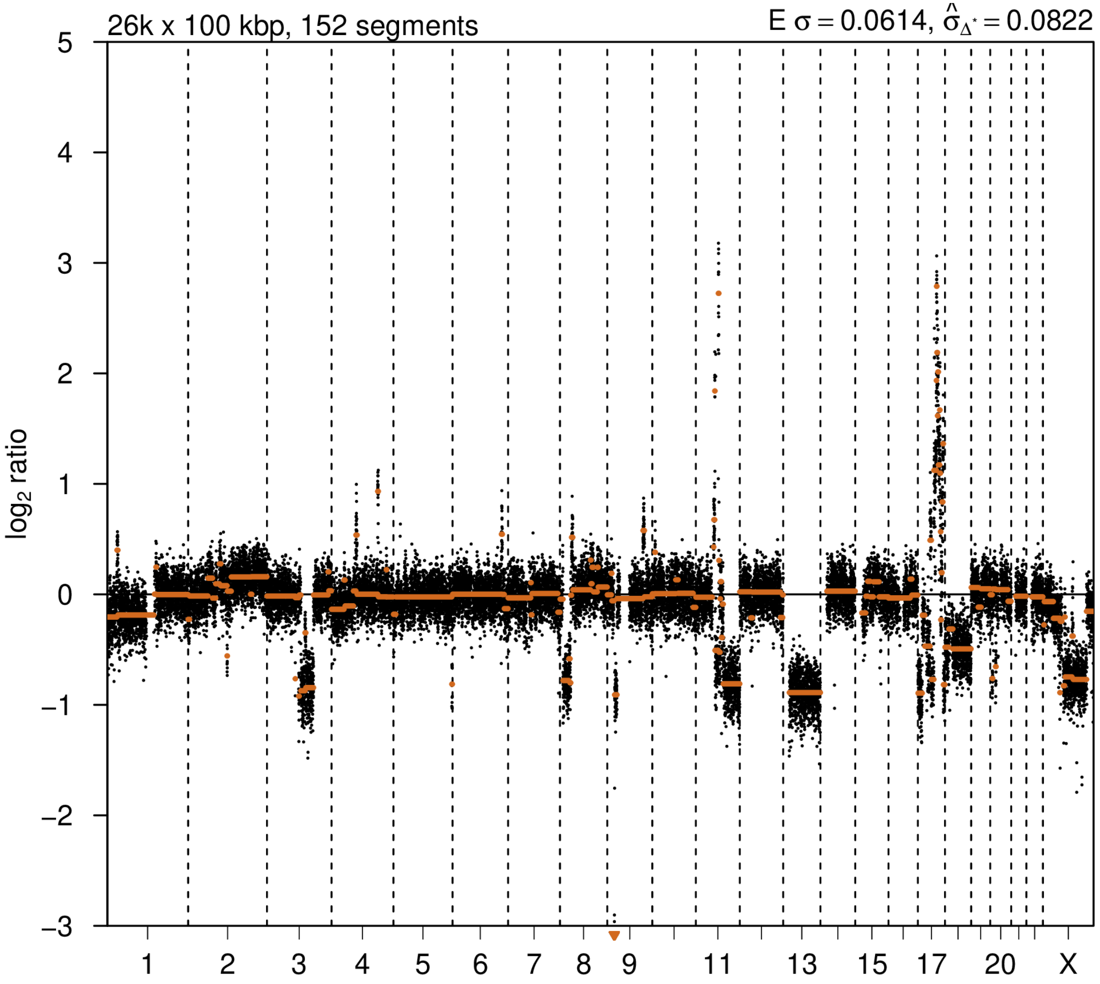


### 2nd event

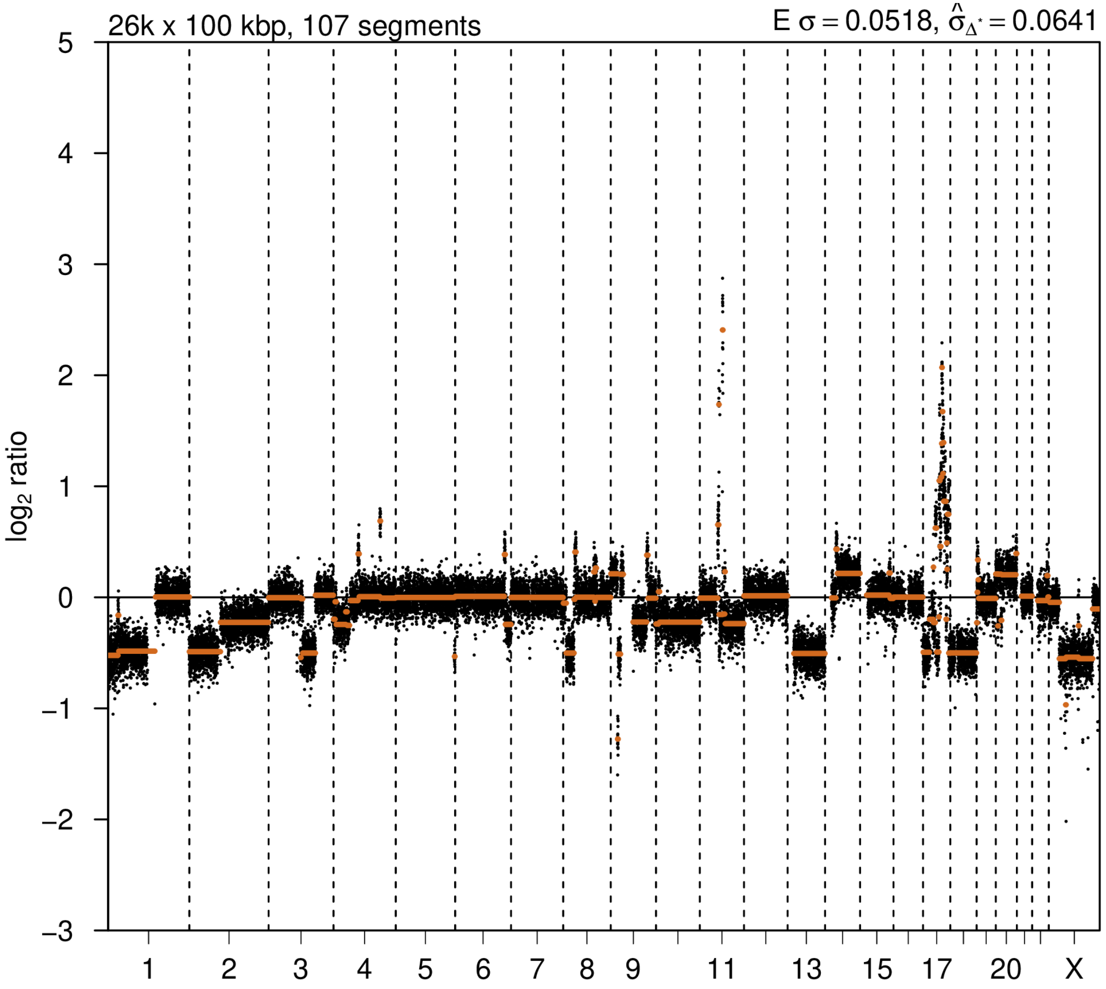

# P038

| Syn/Meta | Time from 1st surgery to 2nd event (Months) | Side 2nd event | Histology 2nd event | Surgery | AdjuvanT Treatment Pri (RT/ HT) | ER Pri | ER 2nd event | Her2 Pri | Her2 2nd event | Grade Pri | Grade 2nd event | Quadrant 2nd event | Margins | Screening | Clonality P value Copy N | Clonality P value Panel seq | Clonality P value WES | Final verdict |
|---|---|---|---|---|---|---|---|---|---|---|---|---|---|---|---|---|---|---|
| metachronous | 100 | Ipsilateral | IDC no DCIS | lumpectomy | None | + | + | - | - | 2 | 3 | distant from primary | involved | symptomatic | 0.00032457 | NA | NA | Related |

### Primary event

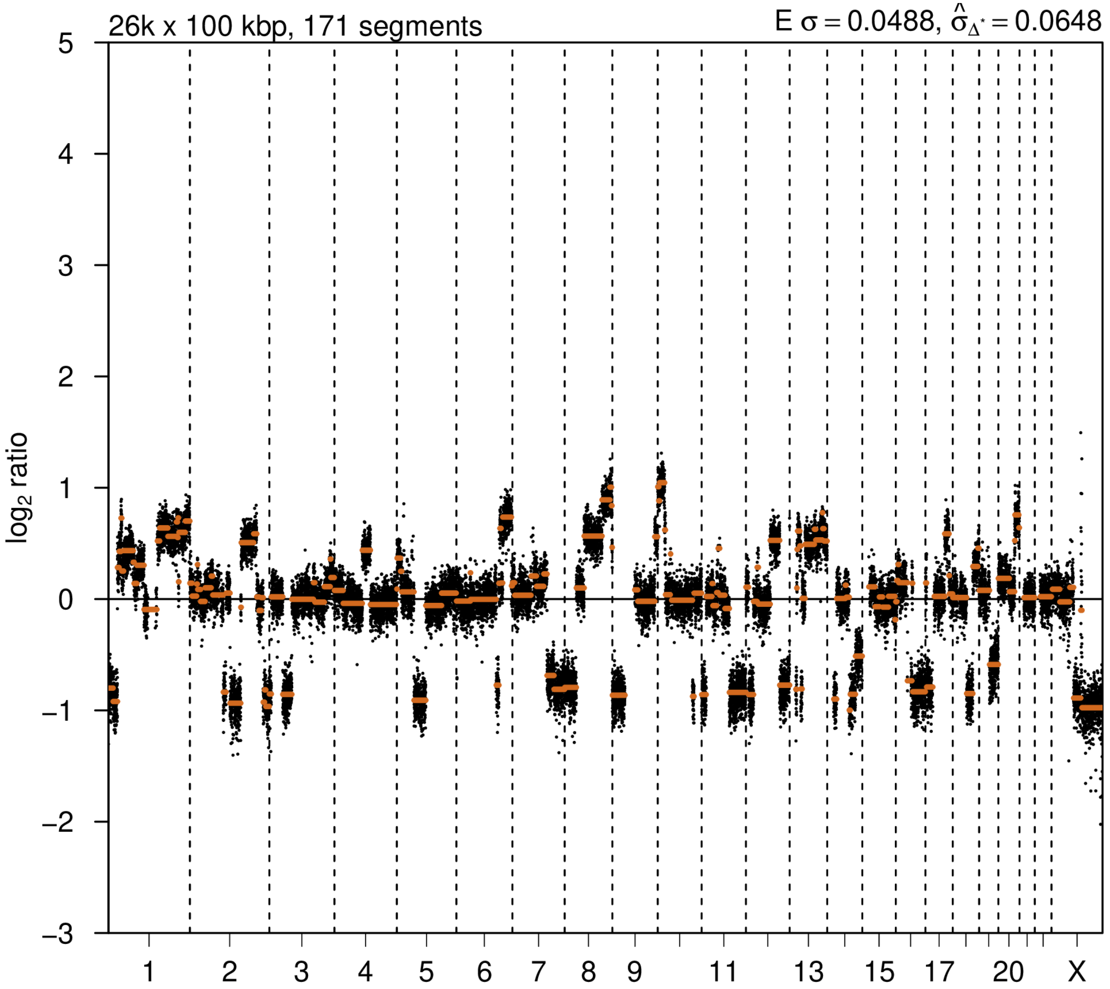


### 2nd event

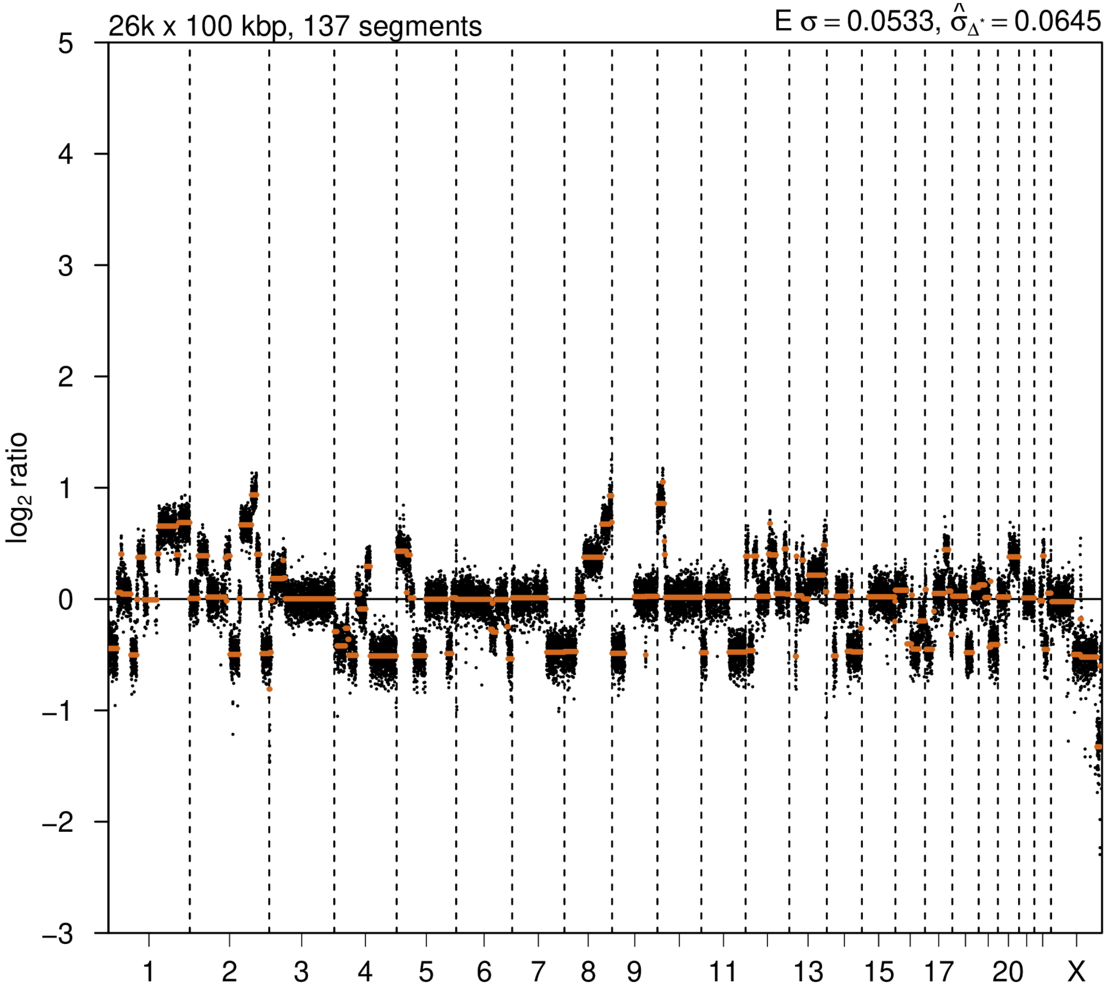

# P039

| Syn/Meta | Time from 1st surgery to 2nd event (Months) | Side 2nd event | Histology 2nd event | Surgery | AdjuvanT Treatment Pri (RT/ HT) | ER Pri | ER 2nd event | Her2 Pri | Her2 2nd event | Grade Pri | Grade 2nd event | Quadrant 2nd event | Margins | Screening | Clonality P value Copy N | Clonality P value Panel seq | Clonality P value WES | Final verdict |
|---|---|---|---|---|---|---|---|---|---|---|---|---|---|---|---|---|---|---|
| metachronous | 66 | Ipsilateral | IDC no DCIS | lumpectomy | None | - | - | + | + | 3 | 3 | distant from primary | Clear | symptomatic | 0.00032457 | 0.004 | NA | Related |

### Primary event

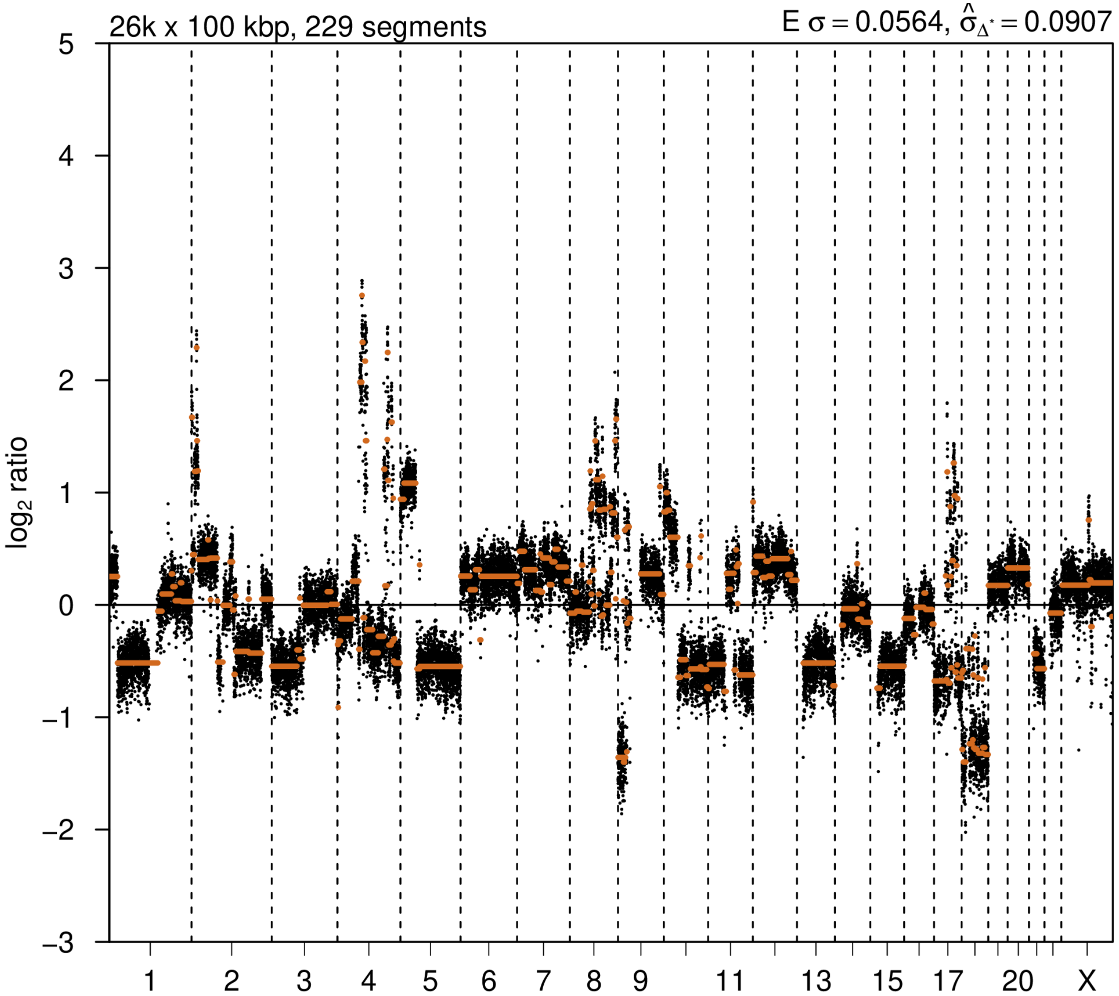


### 2nd event

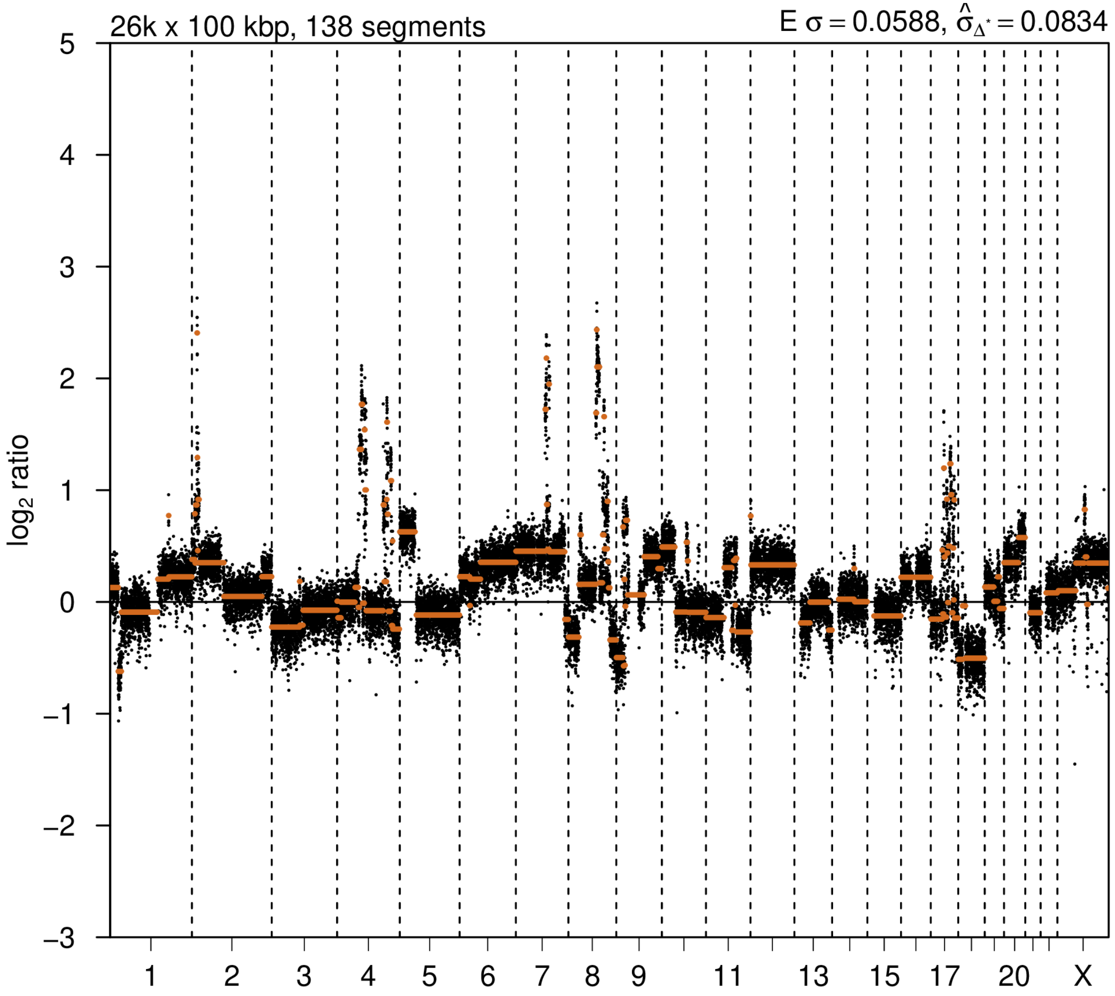

# P040

| Syn/Meta | Time from 1st surgery to 2nd event (Months) | Side 2nd event | Histology 2nd event | Surgery | AdjuvanT Treatment Pri (RT/ HT) | ER Pri | ER 2nd event | Her2 Pri | Her2 2nd event | Grade Pri | Grade 2nd event | Quadrant 2nd event | Margins | Screening | Clonality P value Copy N | Clonality P value Panel seq | Clonality P value WES | Final verdict |
|---|---|---|---|---|---|---|---|---|---|---|---|---|---|---|---|---|---|---|
| metachronous | 84 | Ipsilateral | IDC with DCIS | lumpectomy | None | + | + | + | + | 1 | 2 | NA | Clear | symptomatic | 0.00032457 | NA | 0.000752445 | Related |

### Primary event

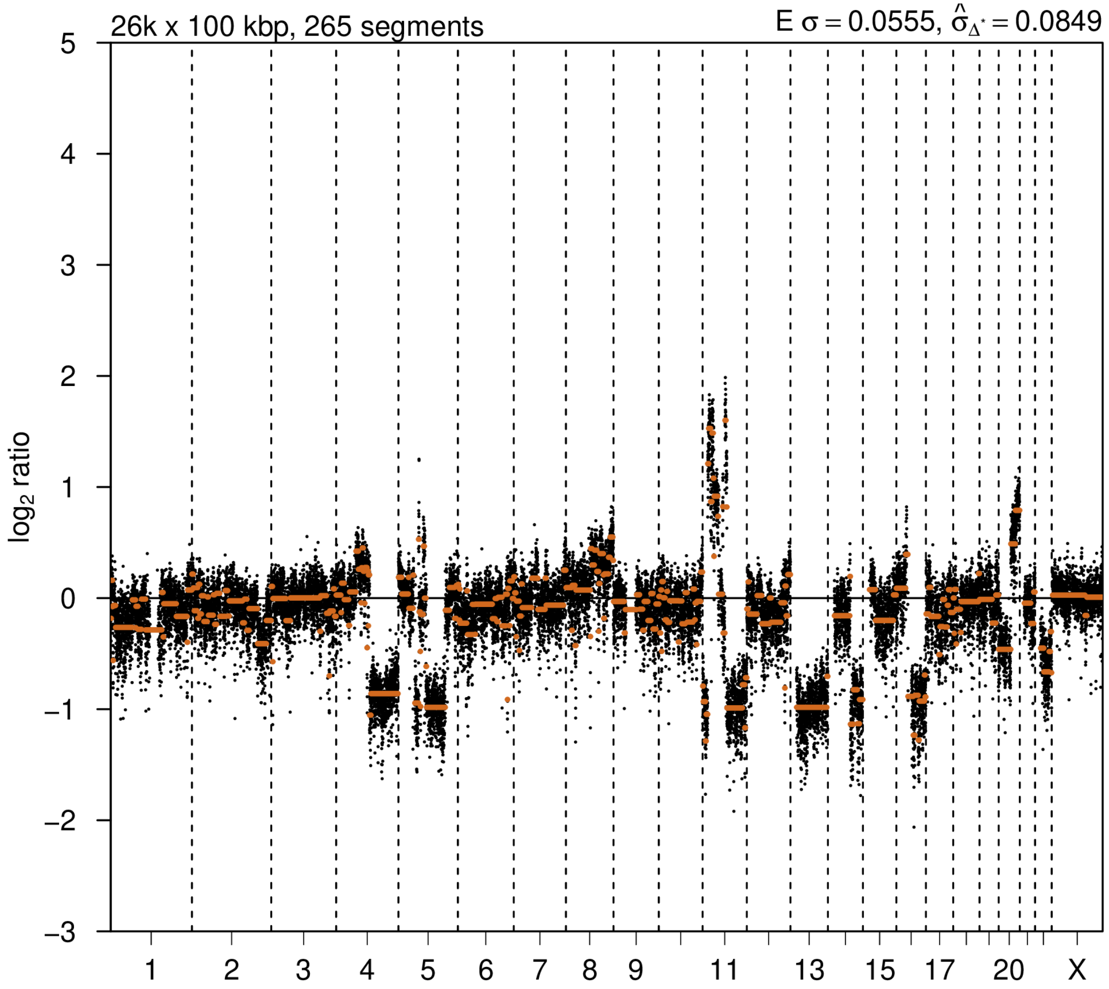


### 2nd event

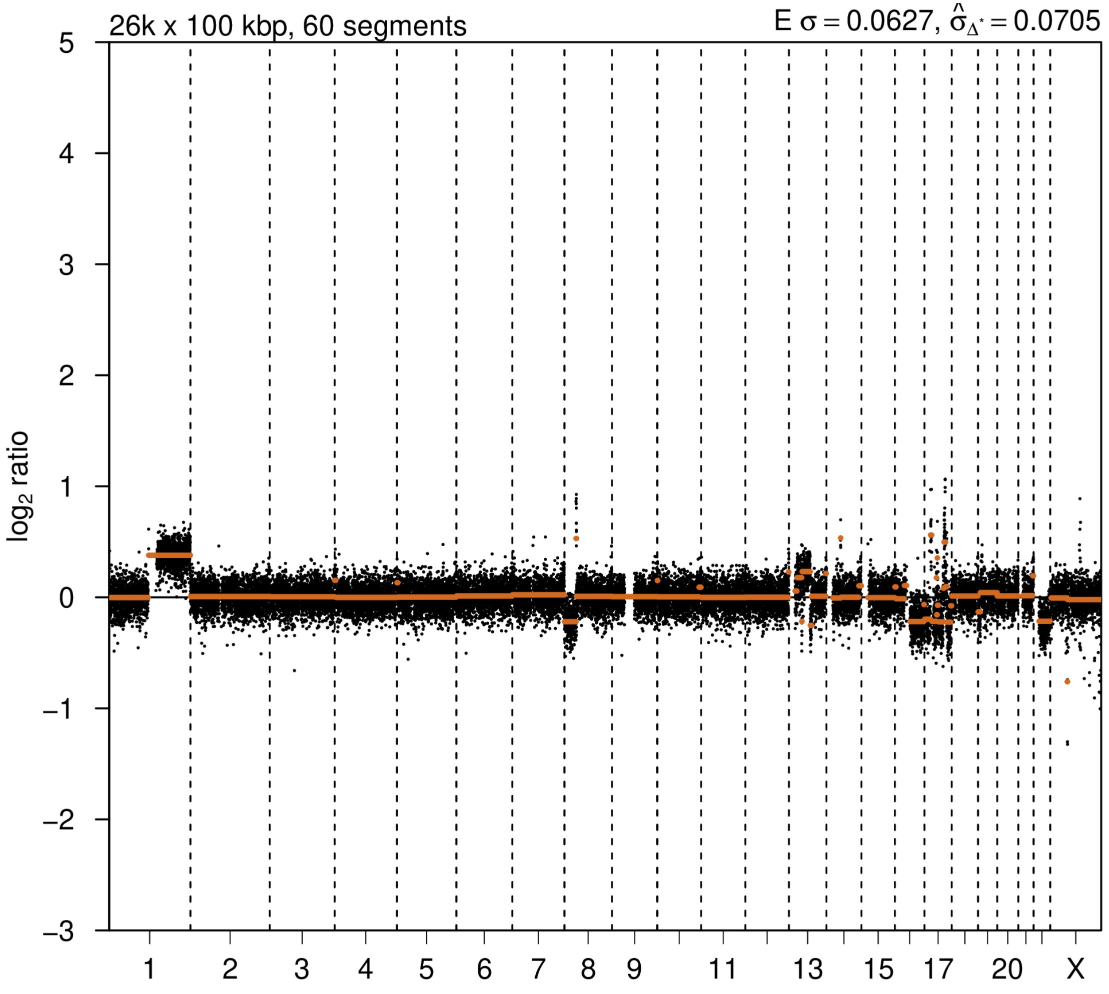

# P041

| Syn/Meta | Time from 1st surgery to 2nd event (Months) | Side 2nd event | Histology 2nd event | Surgery | AdjuvanT Treatment Pri (RT/ HT) | ER Pri | ER 2nd event | Her2 Pri | Her2 2nd event | Grade Pri | Grade 2nd event | Quadrant 2nd event | Margins | Screening | Clonality P value Copy N | Clonality P value Panel seq | Clonality P value WES | Final verdict |
|---|---|---|---|---|---|---|---|---|---|---|---|---|---|---|---|---|---|---|
| metachronous | 97 | Ipsilateral | IDC with DCIS | lumpectomy | None | + | + | + | + | 2 | 2 | NA | Clear | NA | 0.000324 57 | Single mutation - shared | NA | Related |

### Primary event

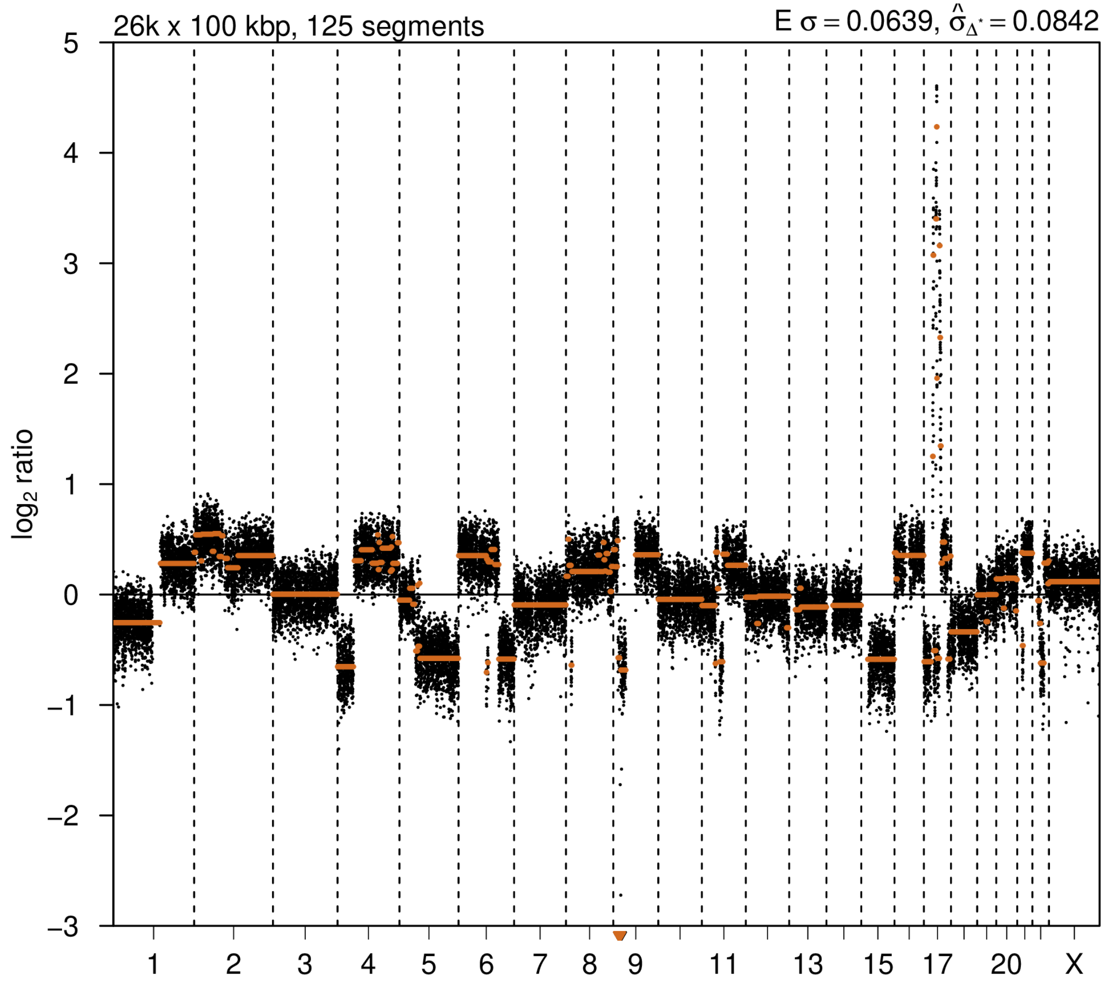


### 2nd event

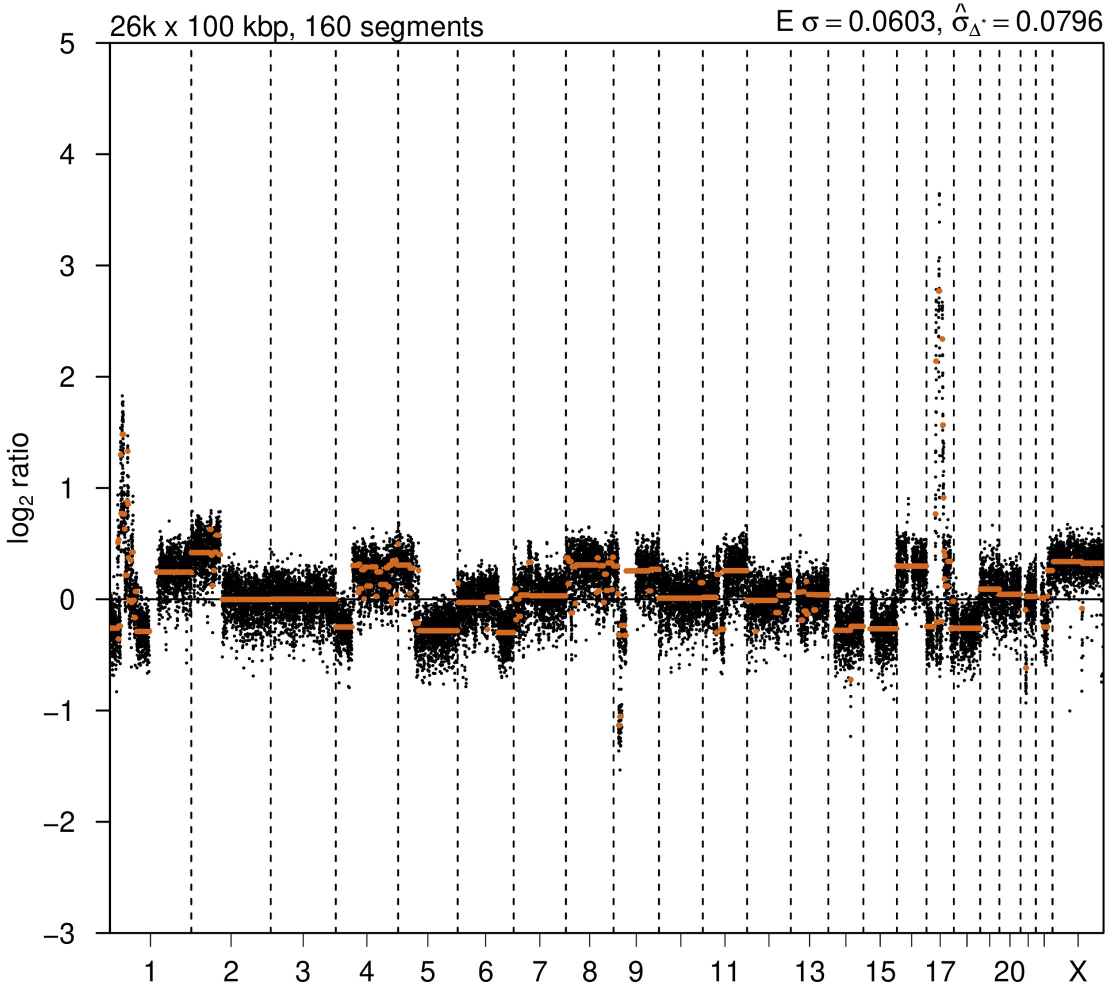

# P042

| Syn/Meta | Time from 1st surgery to 2nd event (Months) | Side 2nd event | Histology 2nd event | Surgery | AdjuvanT Treatment Pri (RT/ HT) | ER Pri | ER 2nd event | Her2 Pri | Her2 2nd event | Grade Pri | Grade 2nd event | Quadrant 2nd event | Margins | Screening | Clonality P value Copy N | Clonality P value Panel seq | Clonality P value WES | Final verdict |
|---|---|---|---|---|---|---|---|---|---|---|---|---|---|---|---|---|---|---|
| metachronous | 72 | Ipsilateral | DCIS only | lumpectomy | None | NA | NA | NA | NA | 3 | 3 | Unknown | Clear | screen-detected | 0.003663 | NA | NA | Related |

### Primary event

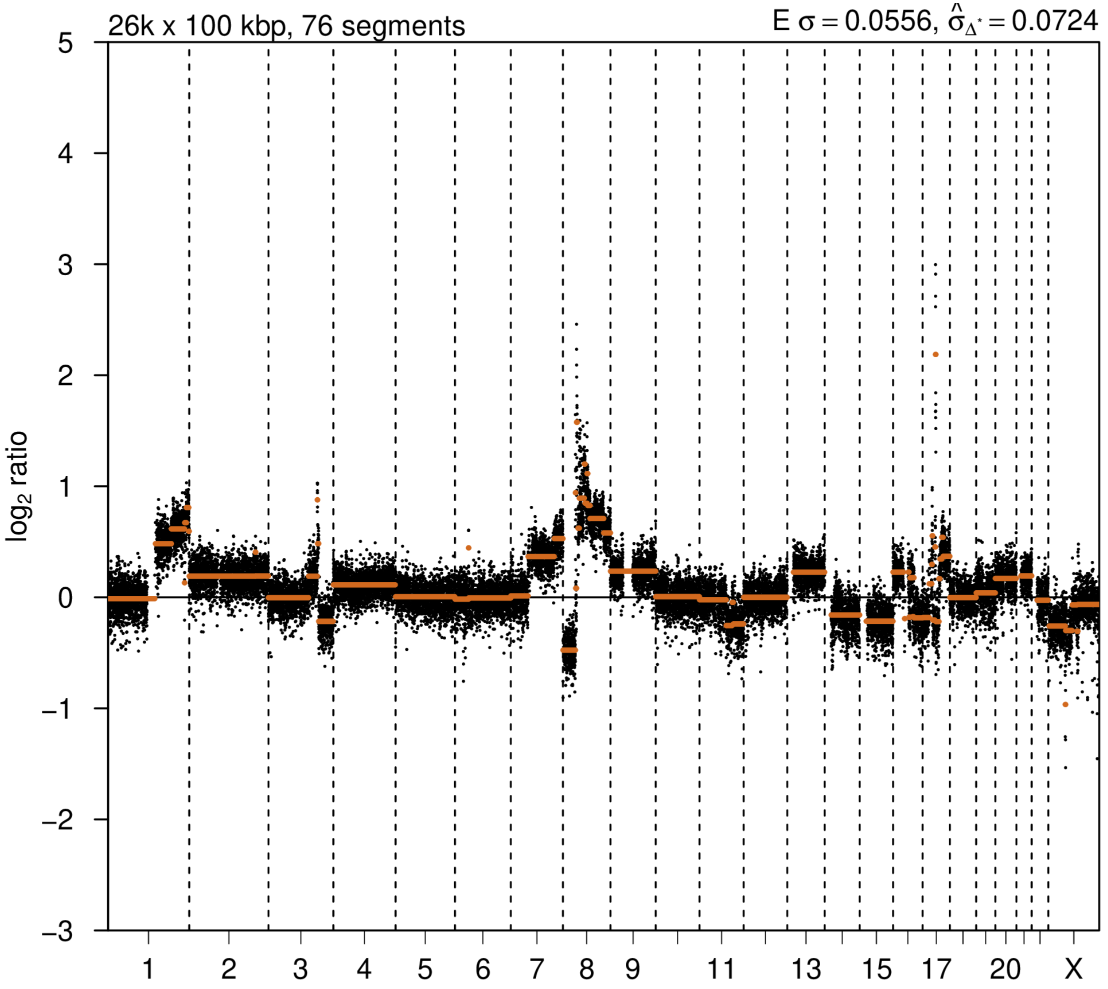


### 2nd event

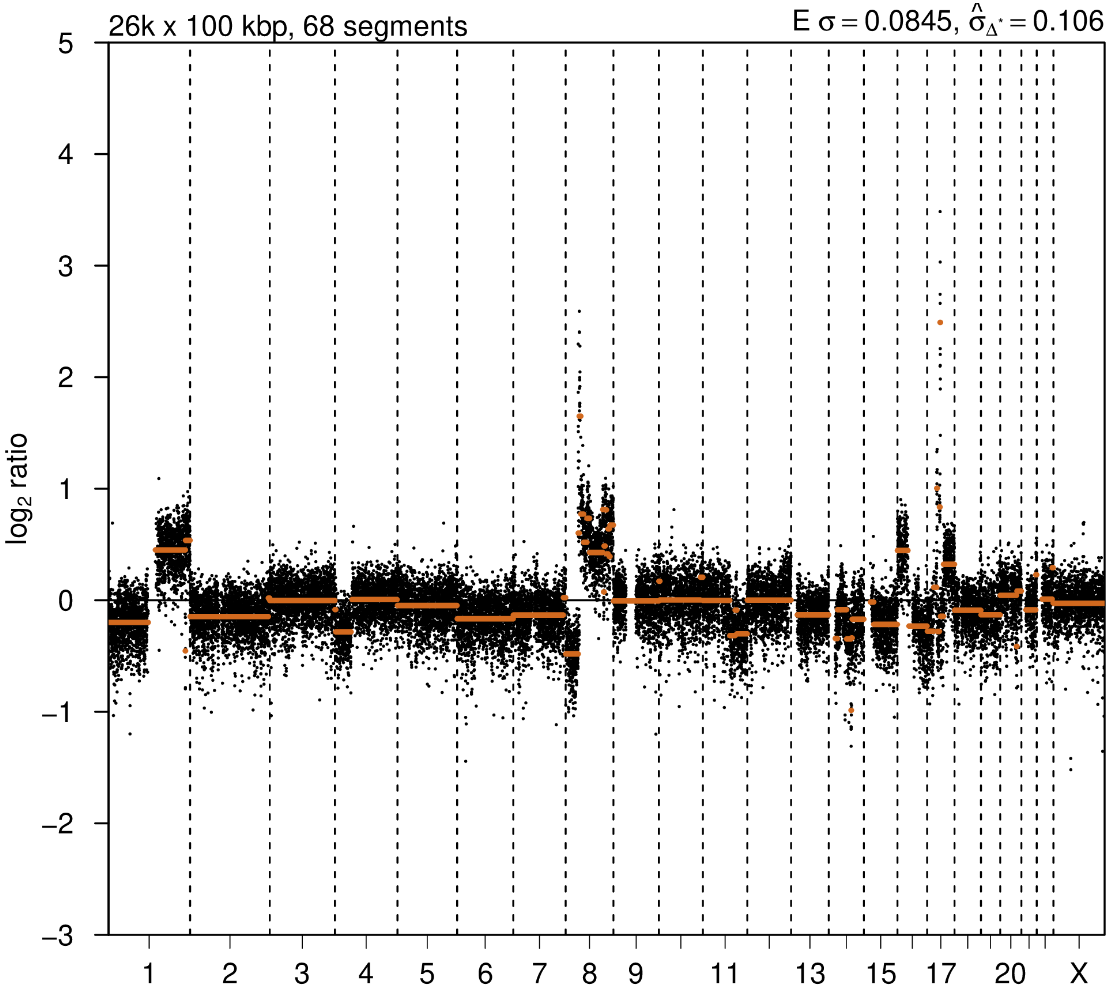

# P043

| Syn/Meta | Time from 1st surgery to 2nd event (Months) | Side 2nd event | Histology 2nd event | Surgery | AdjuvanT Treatment Pri (RT/ HT) | ER Pri | ER 2nd event | Her2 Pri | Her2 2nd event | Grade Pri | Grade 2nd event | Quadrant 2nd event | Margins | Screening | Clonality P value Copy N | Clonality P value Panel seq | Clonality P value WES | Final verdict |
|---|---|---|---|---|---|---|---|---|---|---|---|---|---|---|---|---|---|---|
| metachronous | 74 | Ipsilateral | IDC with DCIS | lumpectomy | None | - | + | + | + | 2 | 3 | NA | Clear | symptomatic | 0.00162285 | 0.003 | 0.000752445 | Related |

### Primary event

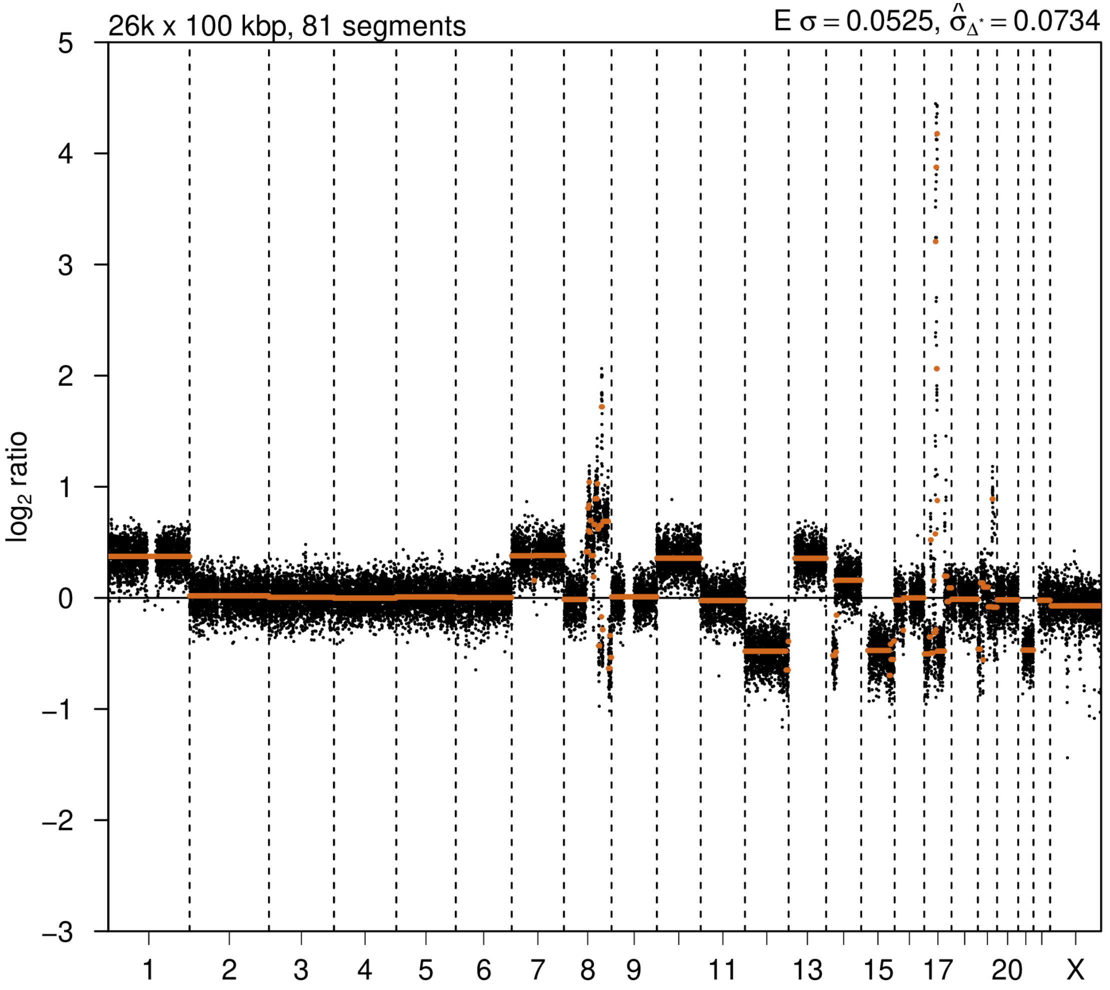


### 2nd event

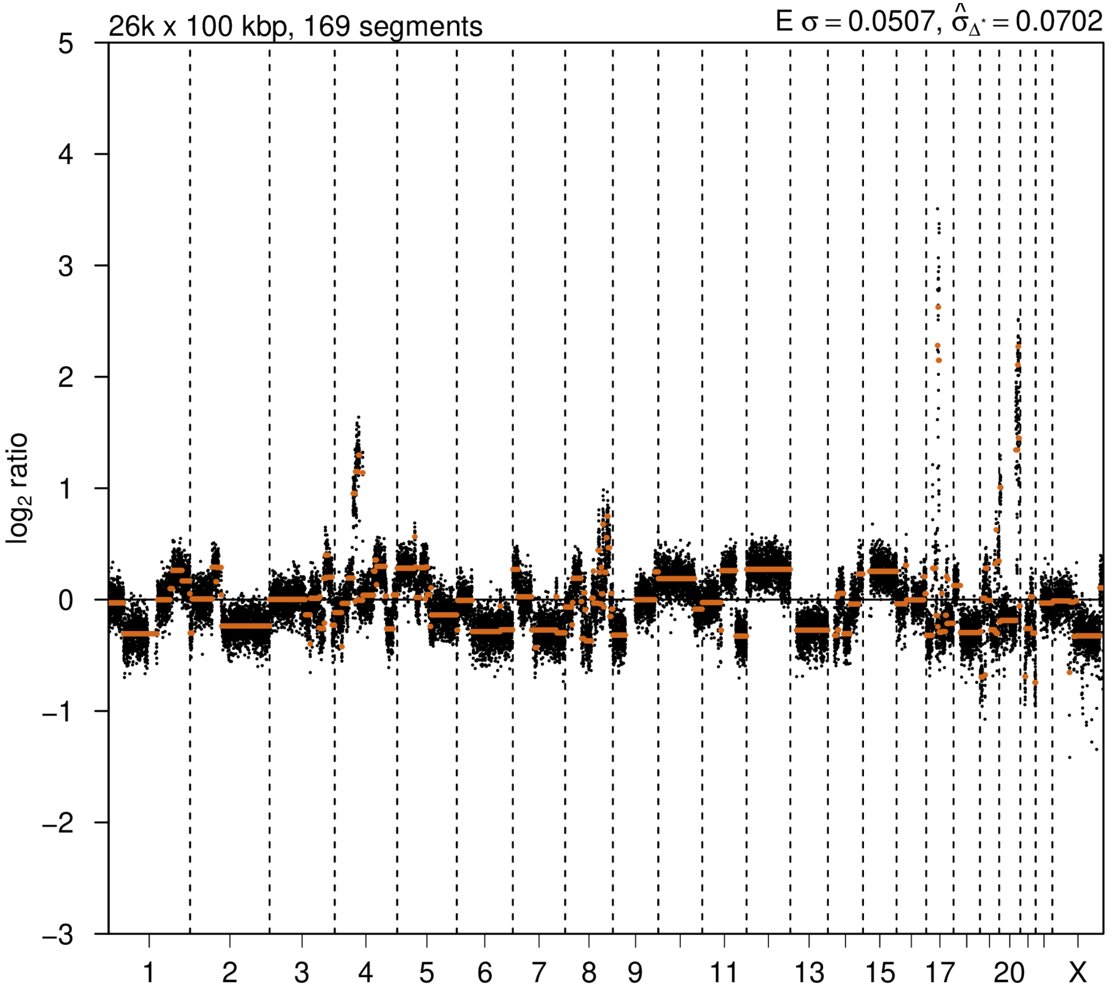

# P044

| Syn/Meta | Time from 1st surgery to 2nd event (Months) | Side 2nd event | Histology 2nd event | Surgery | AdjuvanT Treatment Pri (RT/ HT) | ER Pri | ER 2nd event | Her2 Pri | Her2 2nd event | Grade Pri | Grade 2nd event | Quadrant 2nd event | Margins | Screening | Clonality P value Copy N | Clonality P value Panel seq | Clonality P value WES | Final verdict |
|---|---|---|---|---|---|---|---|---|---|---|---|---|---|---|---|---|---|---|
| metachronous | 109 | Ipsilateral | IDC no DCIS | lumpectomy | None | + | + | - | - | 2 | 3 | NA | Clear | NA | 0.000324 57 | NA | NA | Related |

### Primary event

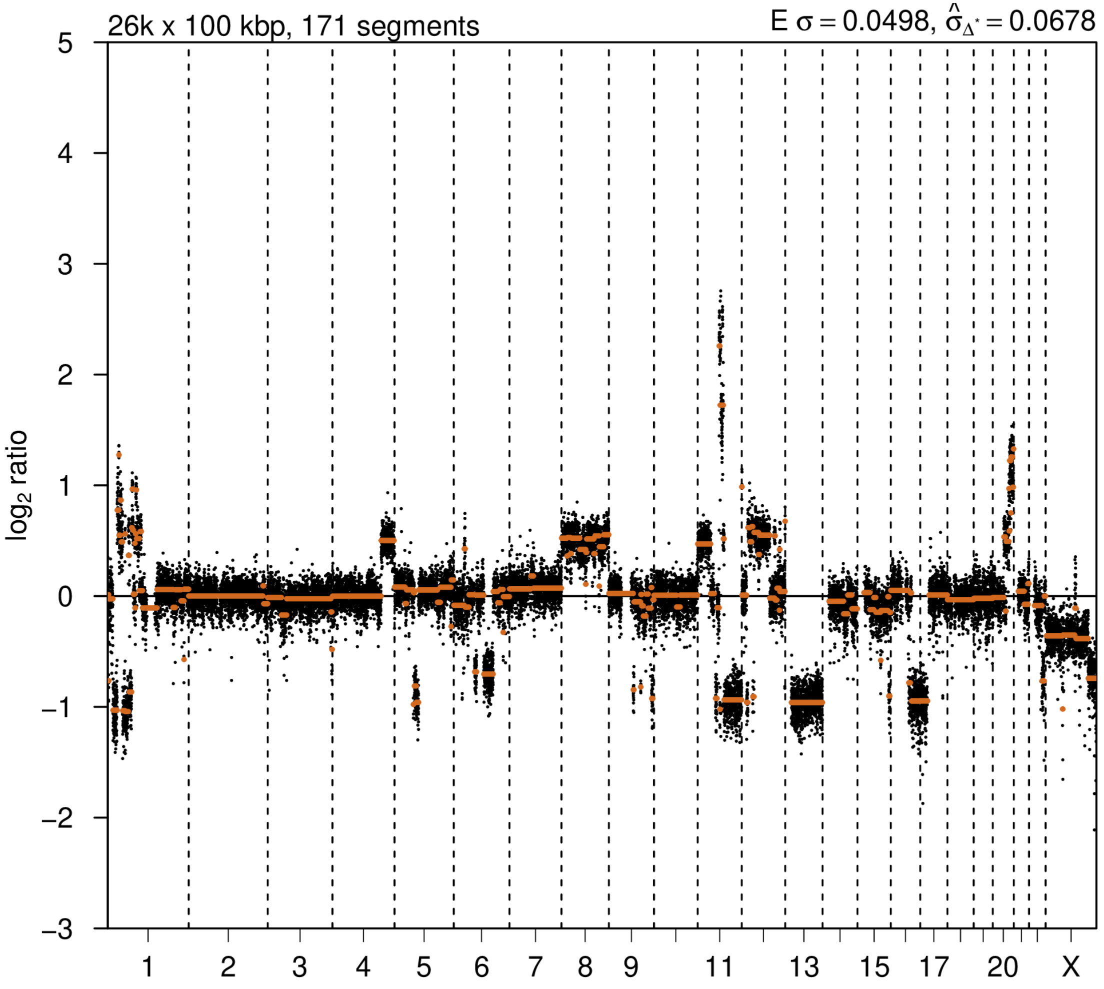


### 2nd event

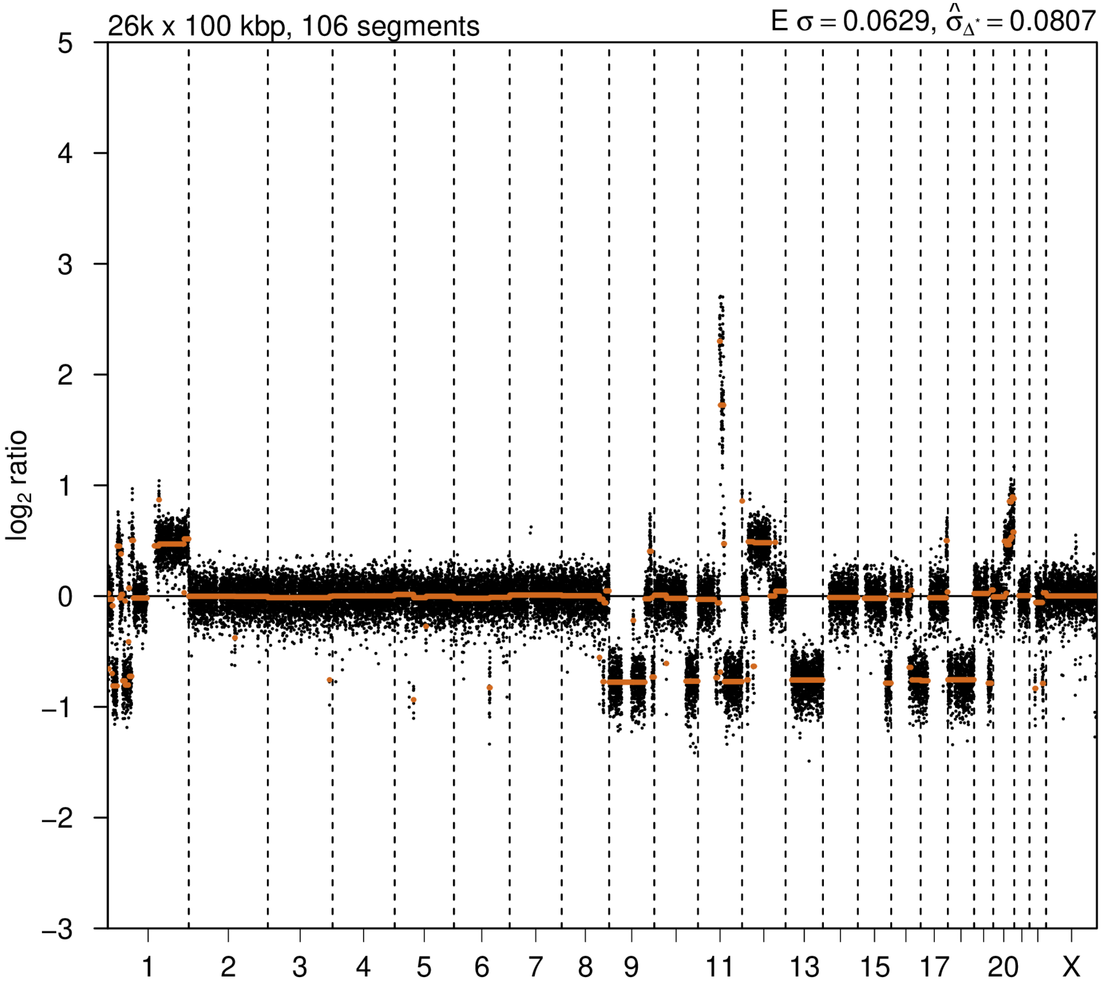

# P045

| Syn/Meta | Time from 1st surgery to 2nd event (Months) | Side 2nd event | Histology 2nd event | Surgery | AdjuvanT Treatment Pri (RT/ HT) | ER Pri | ER 2nd event | Her2 Pri | Her2 2nd event | Grade Pri | Grade 2nd event | Quadrant 2nd event | Margins | Screening | Clonality P value Copy N | Clonality P value Panel seq | Clonality P value WES | Final verdict |
|---|---|---|---|---|---|---|---|---|---|---|---|---|---|---|---|---|---|---|
| metachronous | 60 | Ipsilateral | IDC no DCIS | lumpectomy | None | + | + | - | - | 2 | 2 | NA | involved | symptomatic | 0.00032457 | NA | NA | Related |

### Primary event

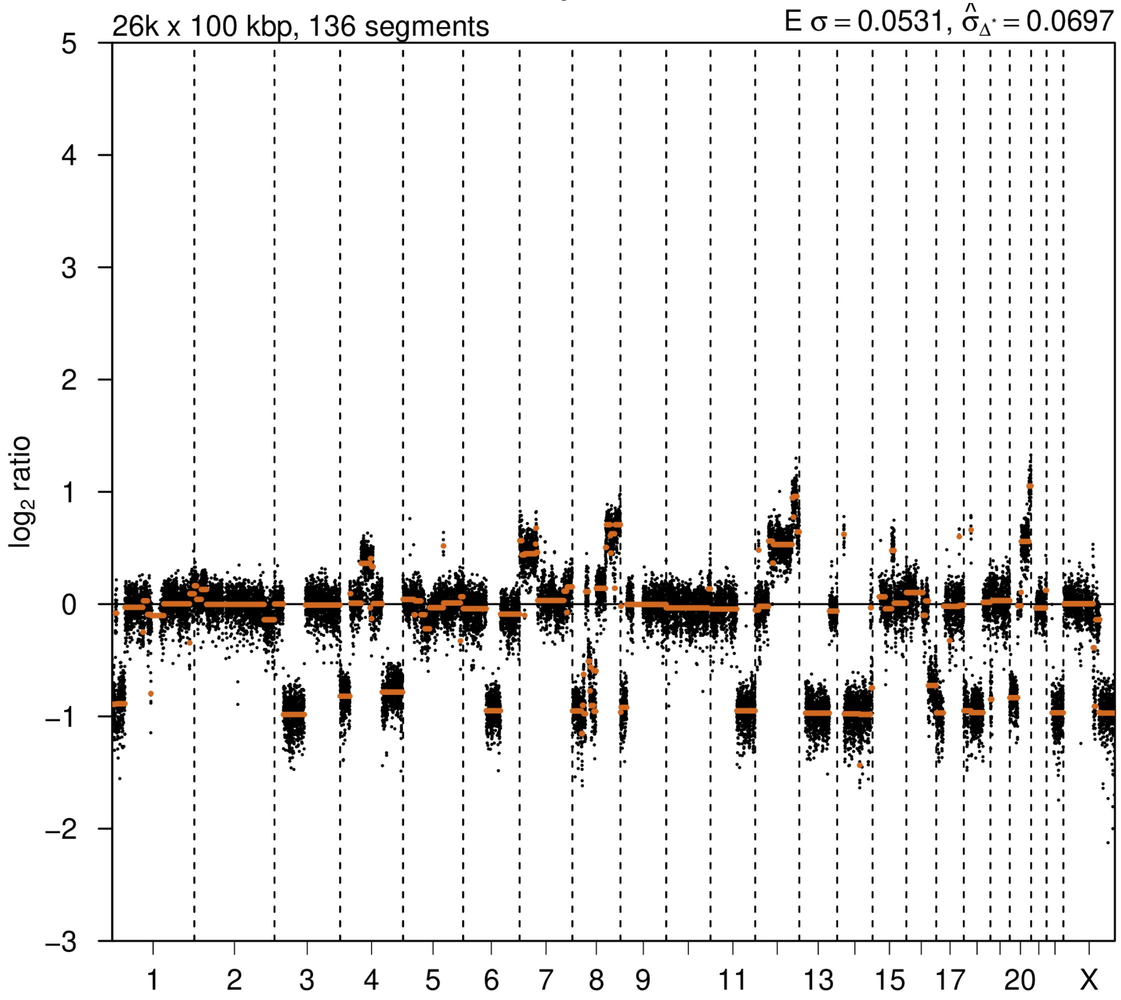


### 2nd event

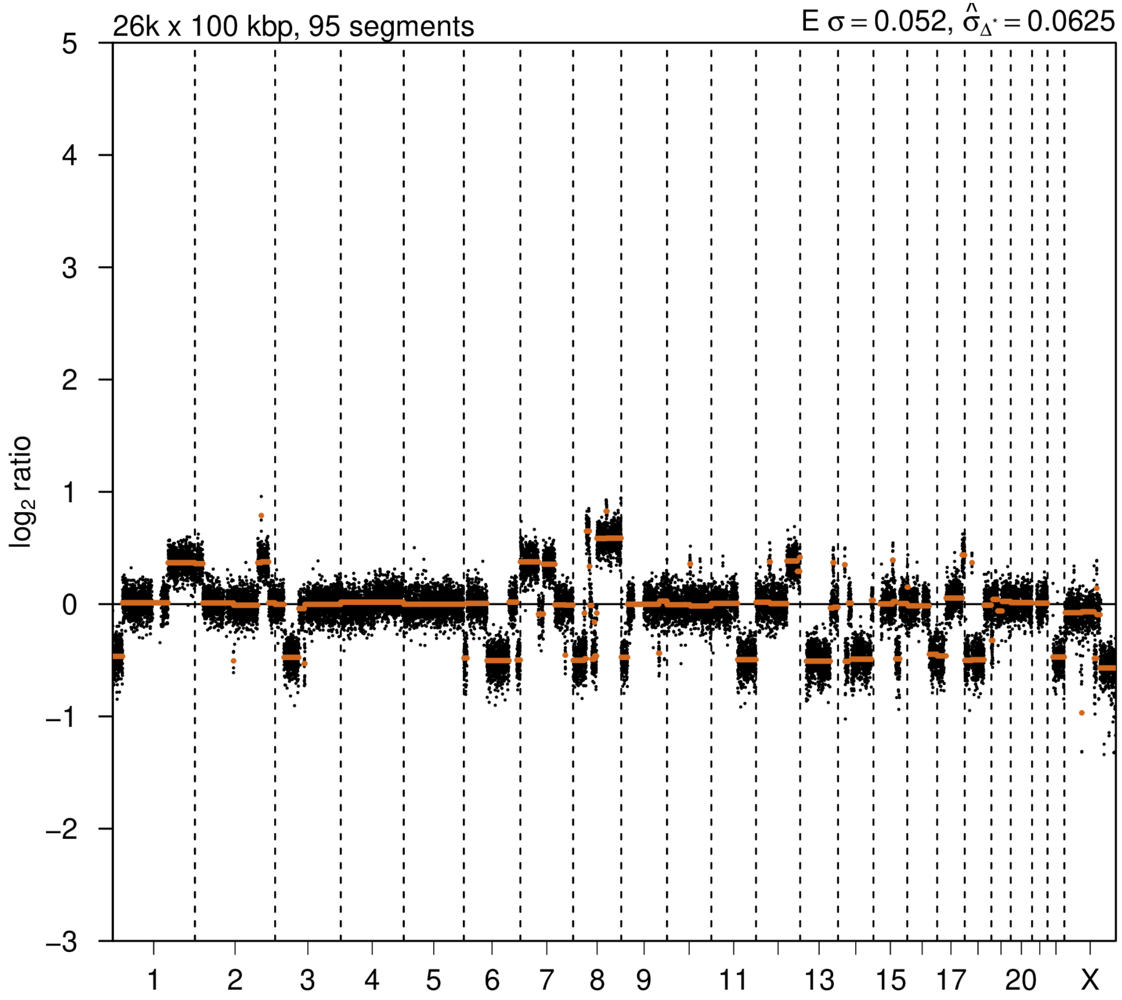

# P046

| Syn/Meta | Time from 1st surgery to 2nd event (Months) | Side 2nd event | Histology 2nd event | Surgery | AdjuvanT Treatment Pri (RT/ HT) | ER Pri | ER 2nd event | Her2 Pri | Her2 2nd event | Grade Pri | Grade 2nd event | Quadrant 2nd event | Margins | Screening | Clonality P value Copy N | Clonality P value Panel seq | Clonality P value WES | Final verdict |
|---|---|---|---|---|---|---|---|---|---|---|---|---|---|---|---|---|---|---|
| metachronous | 95 | Ipsilateral | IDC with DCIS | lumpectomy | None | + | + | - | - | 2 | 2 | NA | Clear | screen-detected | 0.780266147 | 0.006 | NA | Related |

### Primary event

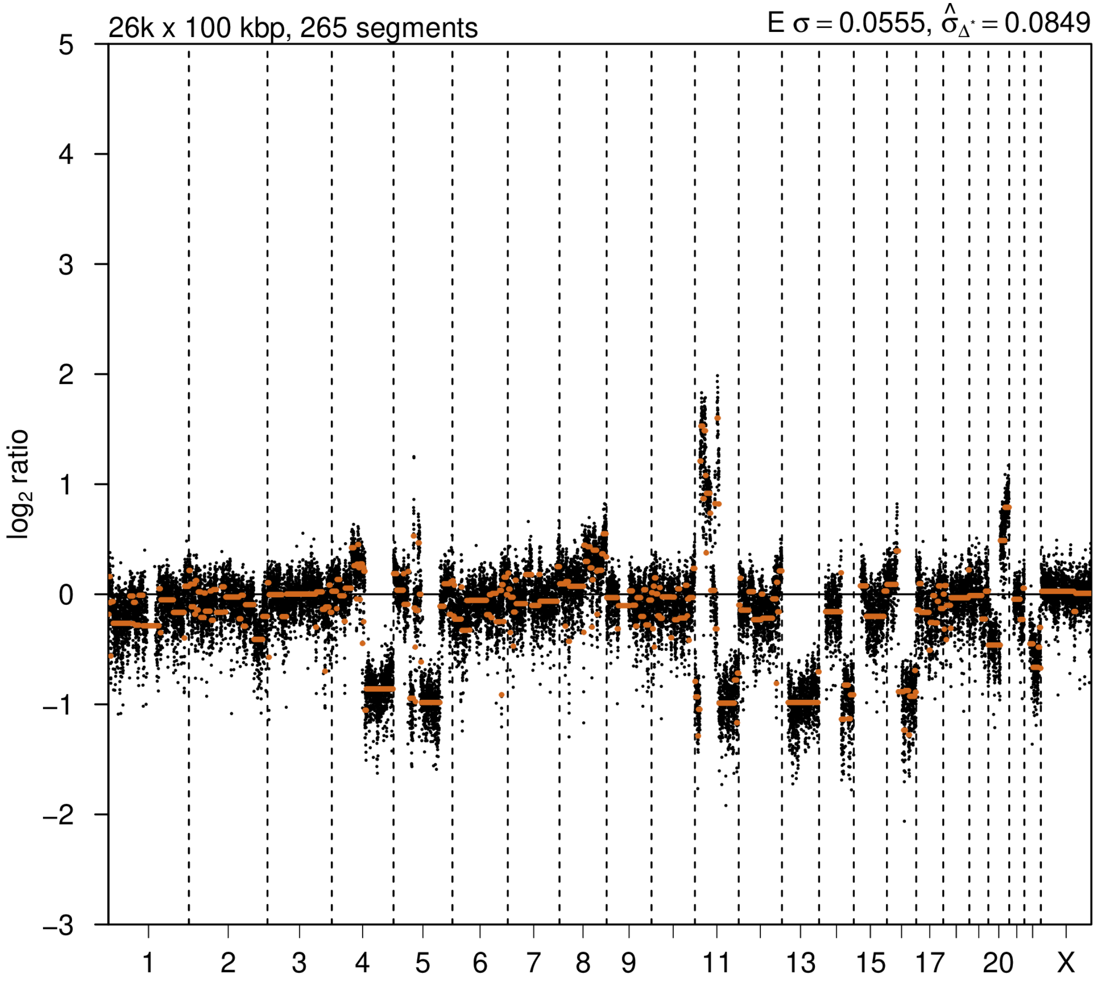


### 2nd event

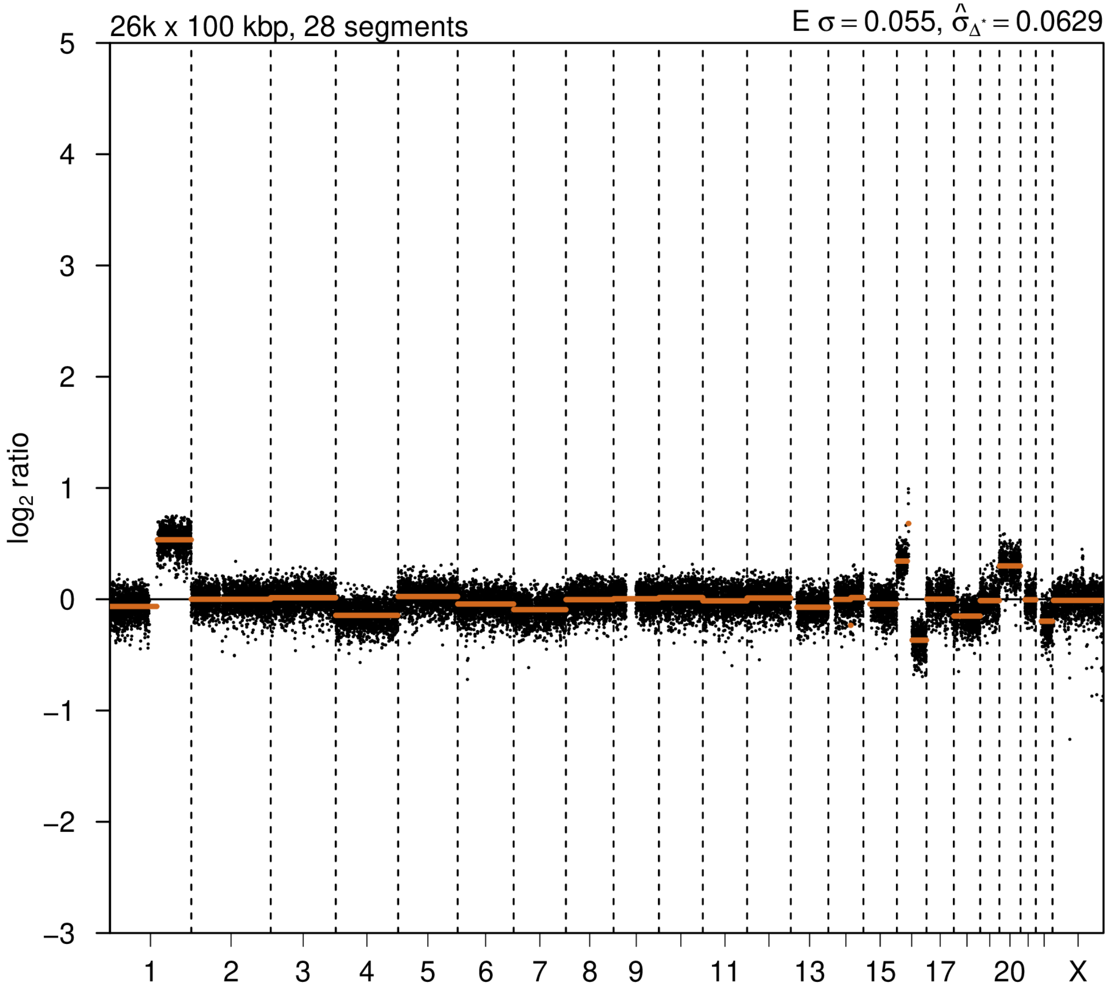

# P050

| Syn/Meta | Time from 1st surgery to 2nd event (Months) | Side 2nd event | Histology 2nd event | Surgery | AdjuvanT Treatment Pri (RT/ HT) | ER Pri | ER 2nd event | Her2 Pri | Her2 2nd event | Grade Pri | Grade 2nd event | Quadrant 2nd event | Margins | Screening | Clonality P value Copy N | Clonality P value Panel seq | Clonality P value WES | Final verdict |
|---|---|---|---|---|---|---|---|---|---|---|---|---|---|---|---|---|---|---|
| metachronous | 88 | Ipsilateral | Invasive | lumpectomy | None | + | + | + | + | 2 | 3 | at or adjacent to primary | Clear | screen-detected | 0.000649 14 | Single mutation - shared | NA | Related |

### Primary event

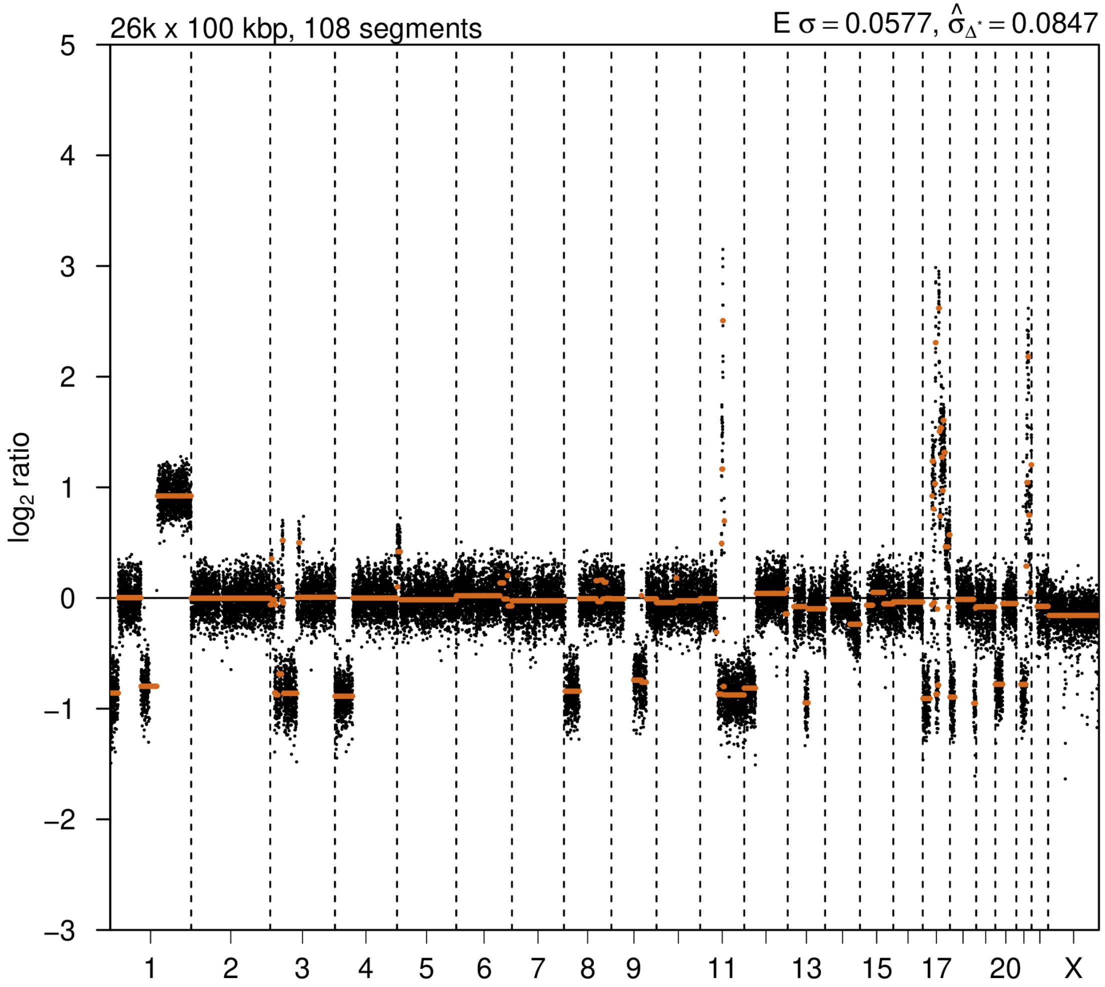


### 2nd event

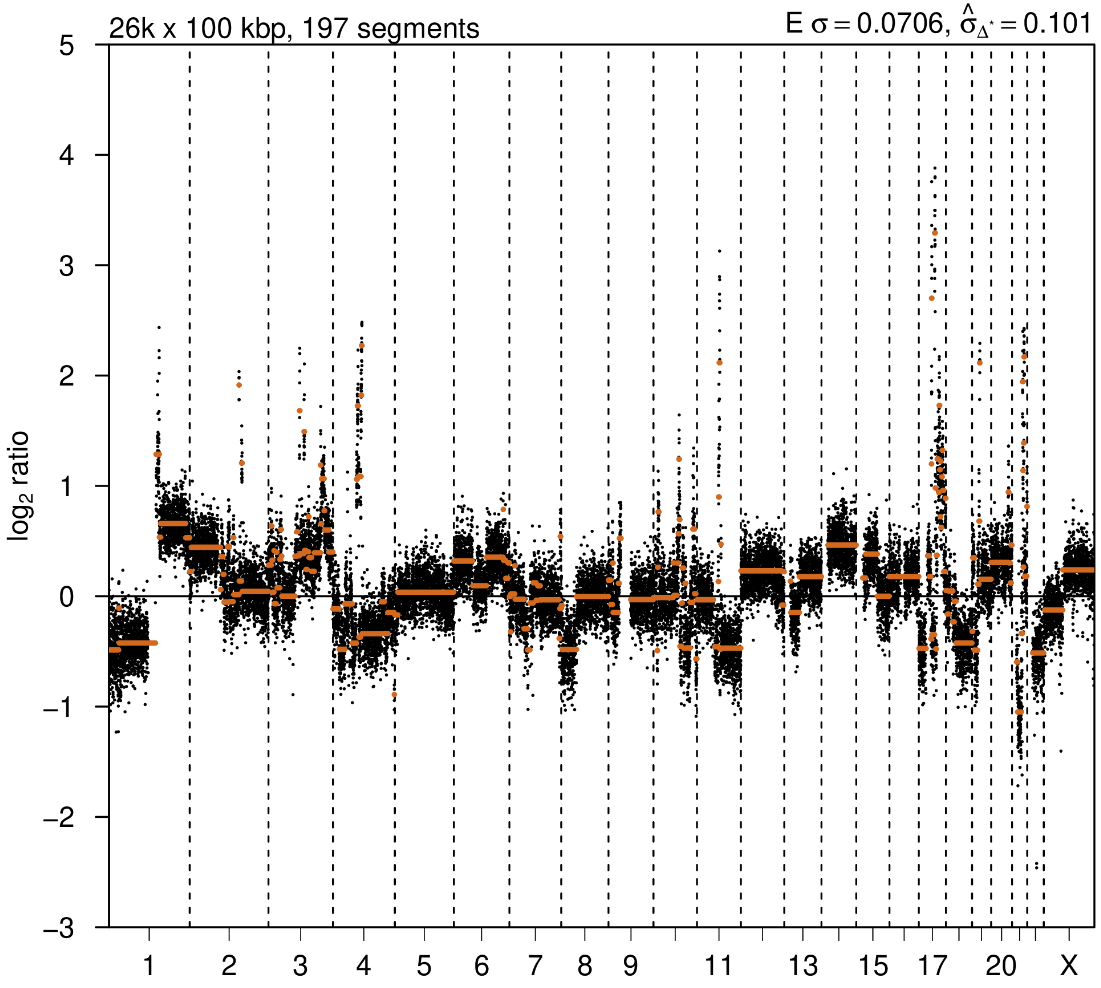

# P051

| Syn/Meta | Time from 1st surgery to 2nd event (Months) | Side 2nd event | Histology 2nd event | Surgery | AdjuvanT Treatment Pri (RT/ HT) | ER Pri | ER 2nd event | Her2 Pri | Her2 2nd event | Grade Pri | Grade 2nd event | Quadrant 2nd event | Margins | Screening | Clonality P value Copy N | Clonality P value Panel seq | Clonality P value WES | Final verdict |
|---|---|---|---|---|---|---|---|---|---|---|---|---|---|---|---|---|---|---|
| metachronous | 62 | Ipsilateral | IDC with DCIS | lumpectomy | None | + | + | + | + | 2 | 3 | at or adjacent to primary | Clear | screen-detected | 0.000649 14 | Single mutation - shared | NA | Related |

### Primary event

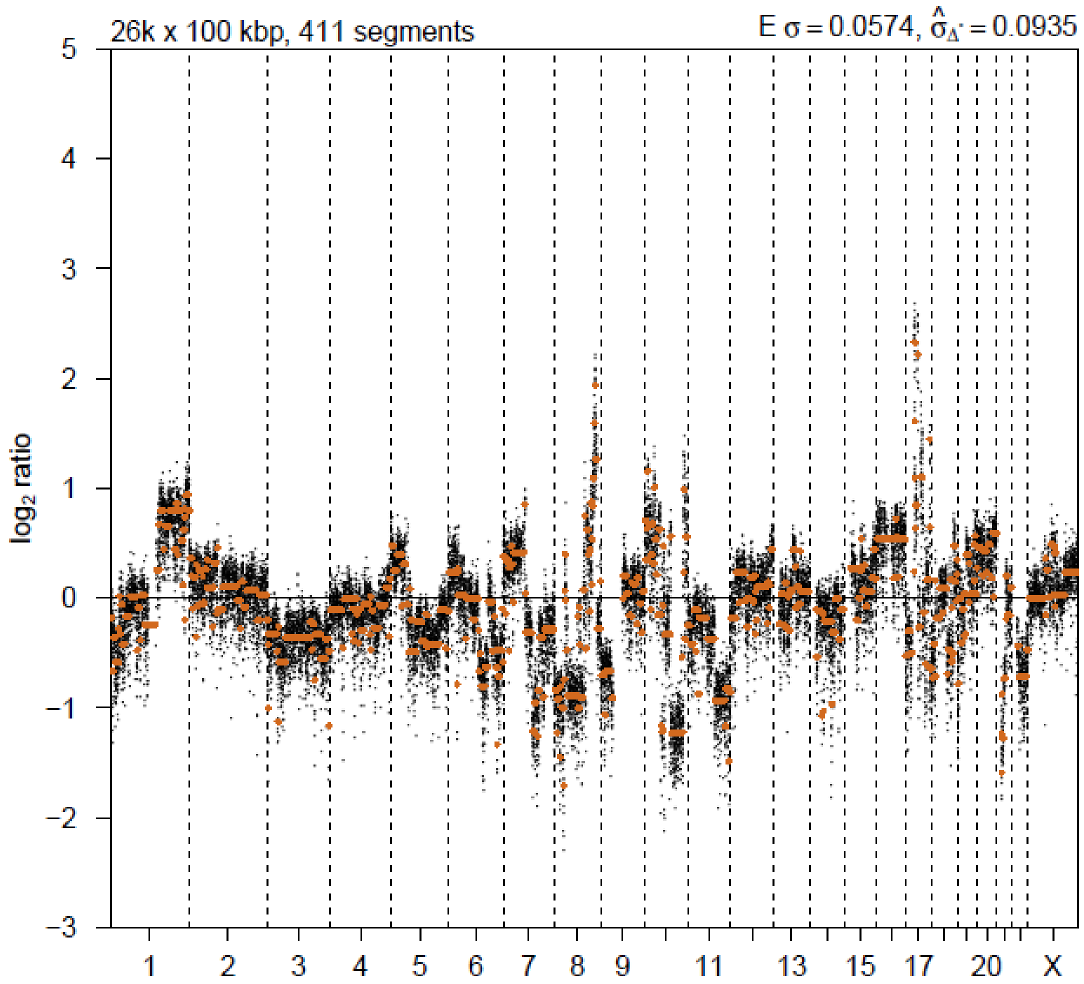


### 2nd event

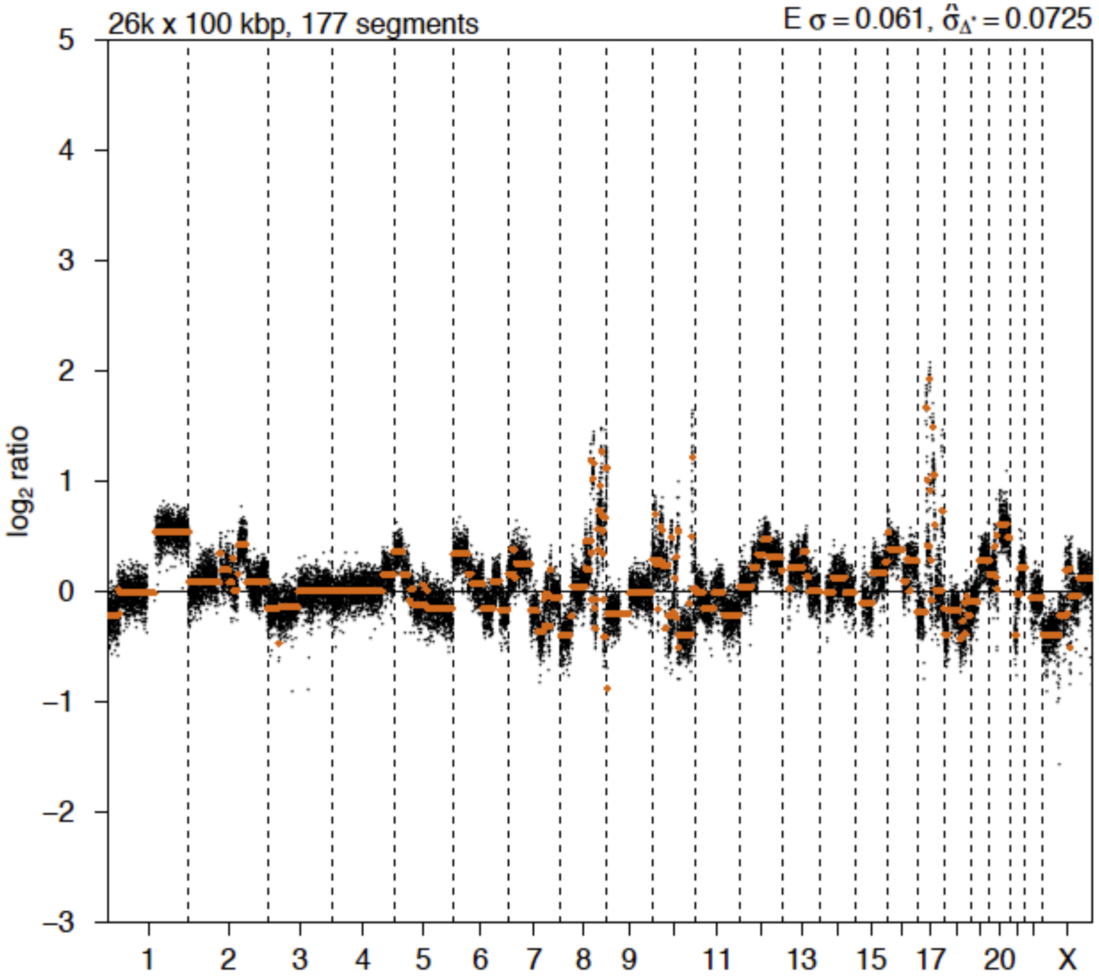

# P052

| Syn/Meta | Time from 1st surgery to 2nd event (Months) | Side 2nd event | Histology 2nd event | Surgery | AdjuvanT Treatment Pri (RT/ HT) | ER Pri | ER 2nd event | Her2 Pri | Her2 2nd event | Grade Pri | Grade 2nd event | Quadrant 2nd event | Margins | Screening | Clonality P value Copy N | Clonality P value Panel seq | Clonality P value WES | Final verdict |
|---|---|---|---|---|---|---|---|---|---|---|---|---|---|---|---|---|---|---|
| metachronous | 86 | Ipsilateral | IDC with DCIS | lumpectomy | None | + | + | - | - | 2 | 2 | at or adjacent to primary | Clear | symptomatic | 0.00032457 | 0.003 | NA | Related |

### Primary event

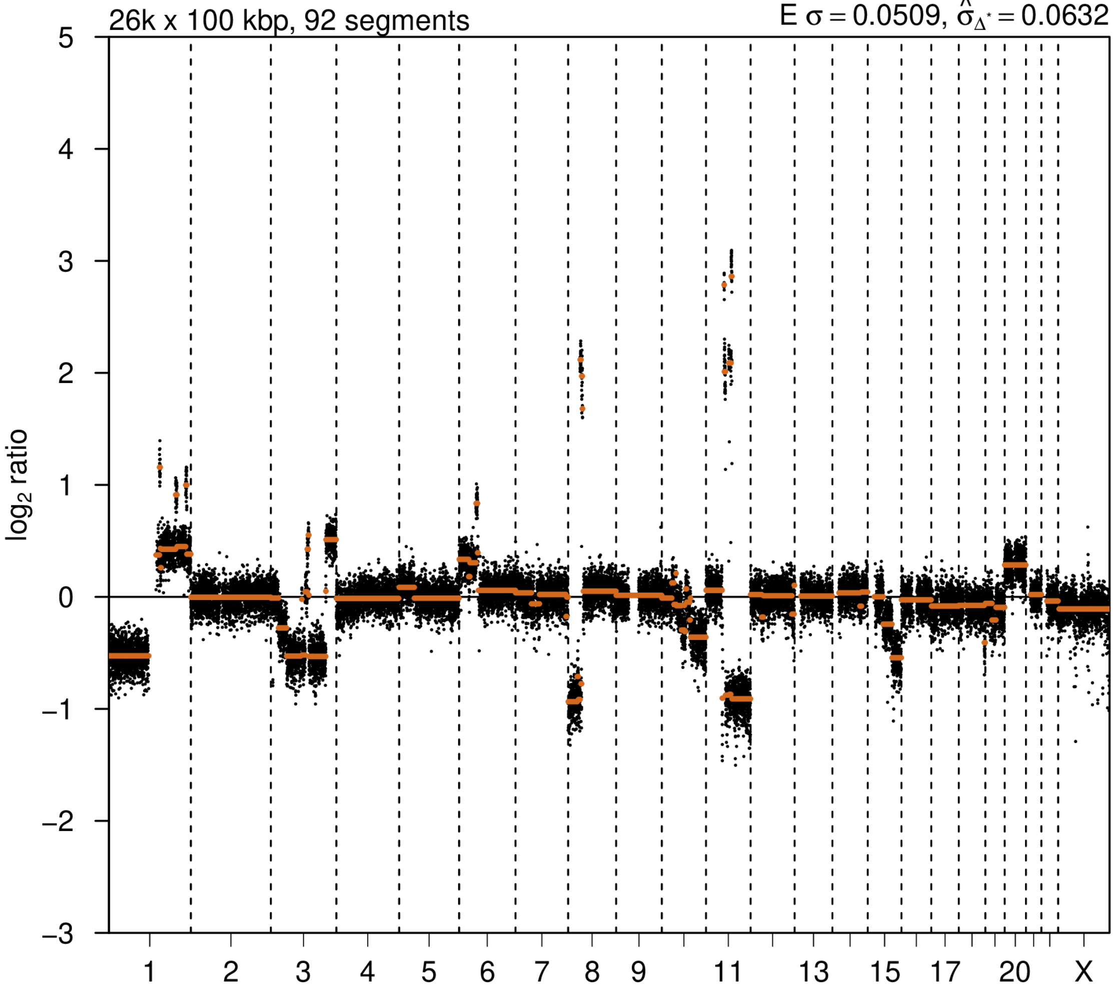


### 2nd event

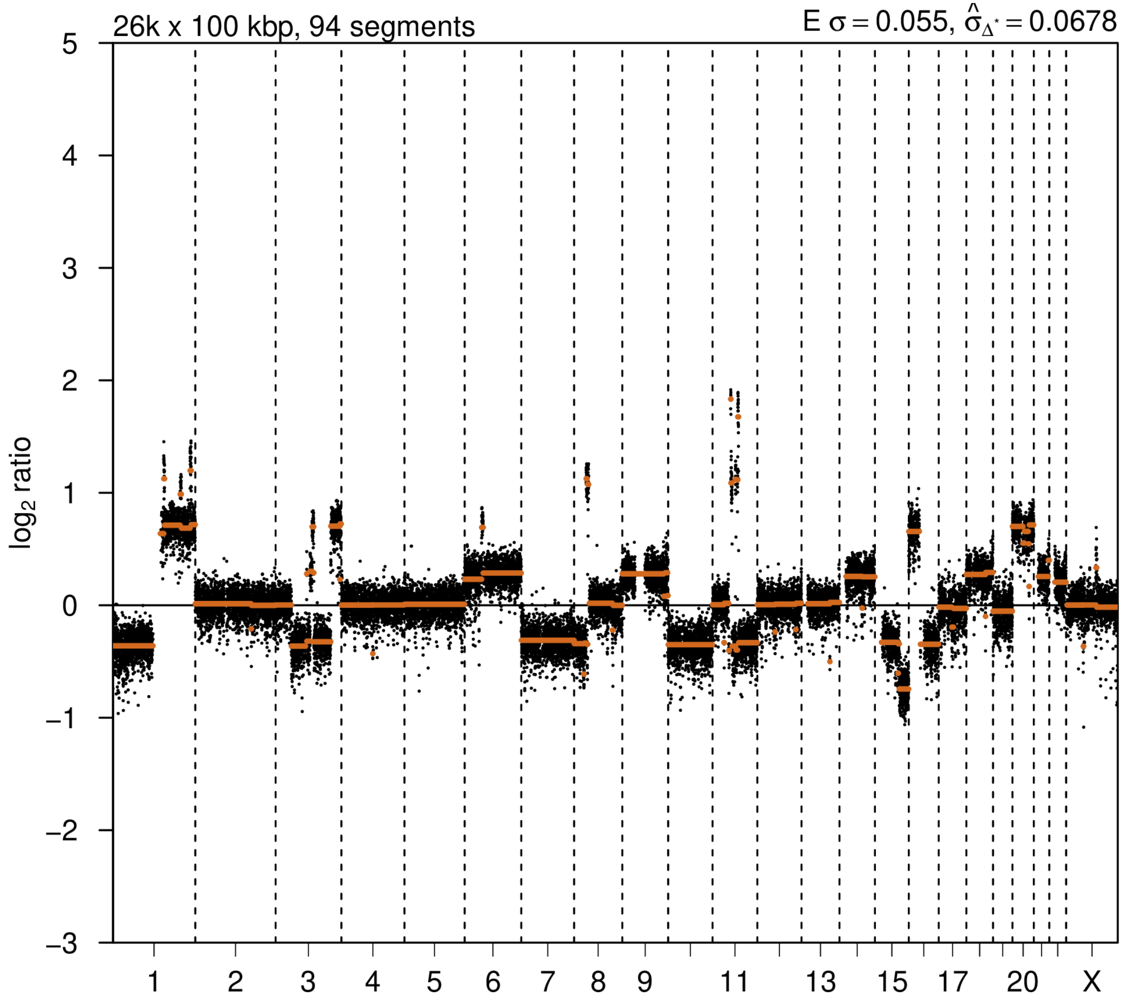

# P053

| Syn/Meta | Time from 1st surgery to 2nd event (Months) | Side 2nd event | Histology 2nd event | Surgery | AdjuvanT Treatment Pri (RT/ HT) | ER Pri | ER 2nd event | Her2 Pri | Her2 2nd event | Grade Pri | Grade 2nd event | Quadrant 2nd event | Margins | Screening | Clonality P value Copy N | Clonality P value Panel seq | Clonality P value WES | Final verdict |
|---|---|---|---|---|---|---|---|---|---|---|---|---|---|---|---|---|---|---|
| metachronous | 72 | Ipsilateral | DCIS only | lumpectomy | None | NA | NA | NA | NA | 3 | 3 | Unknown | Clear | screen-detected | 0.003663 | NA | NA | Related |

### Primary event

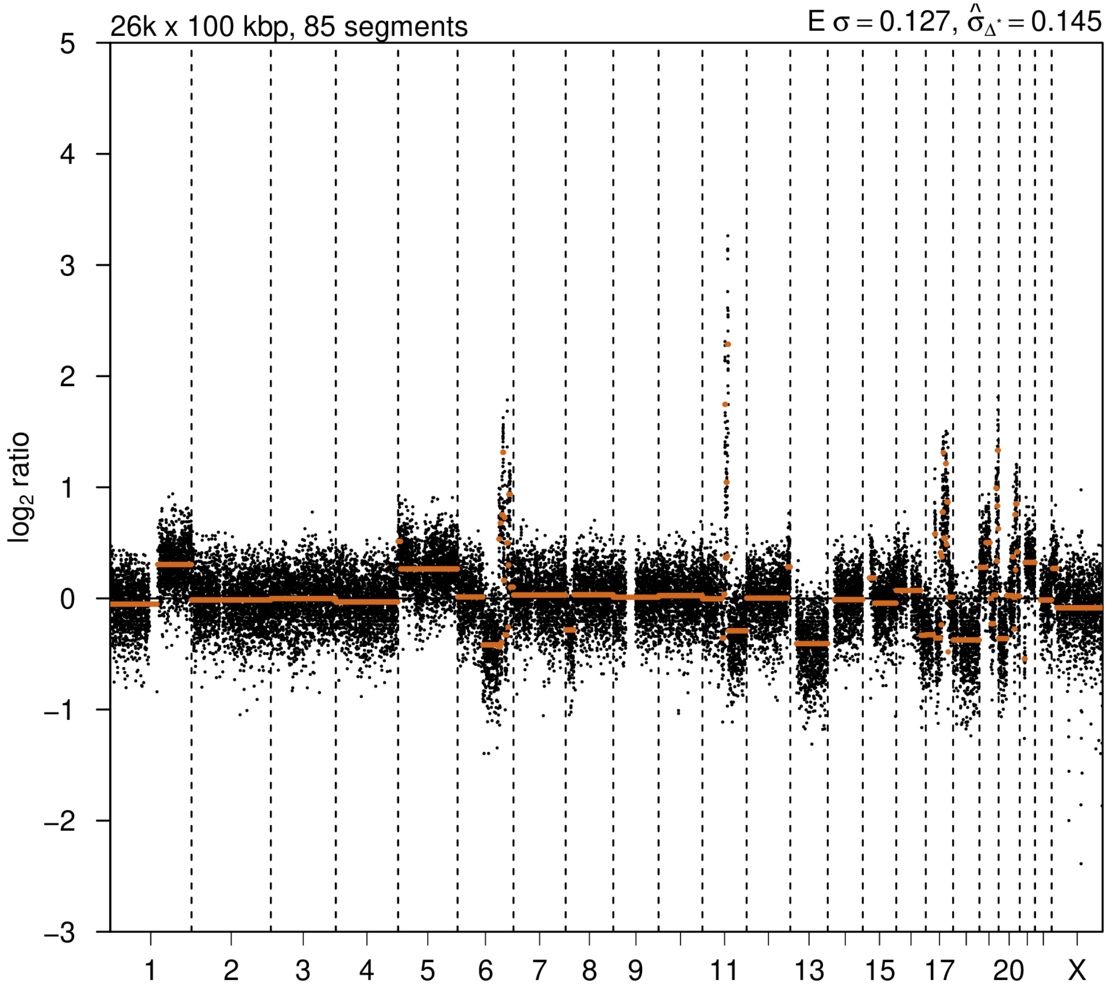


### 2nd event

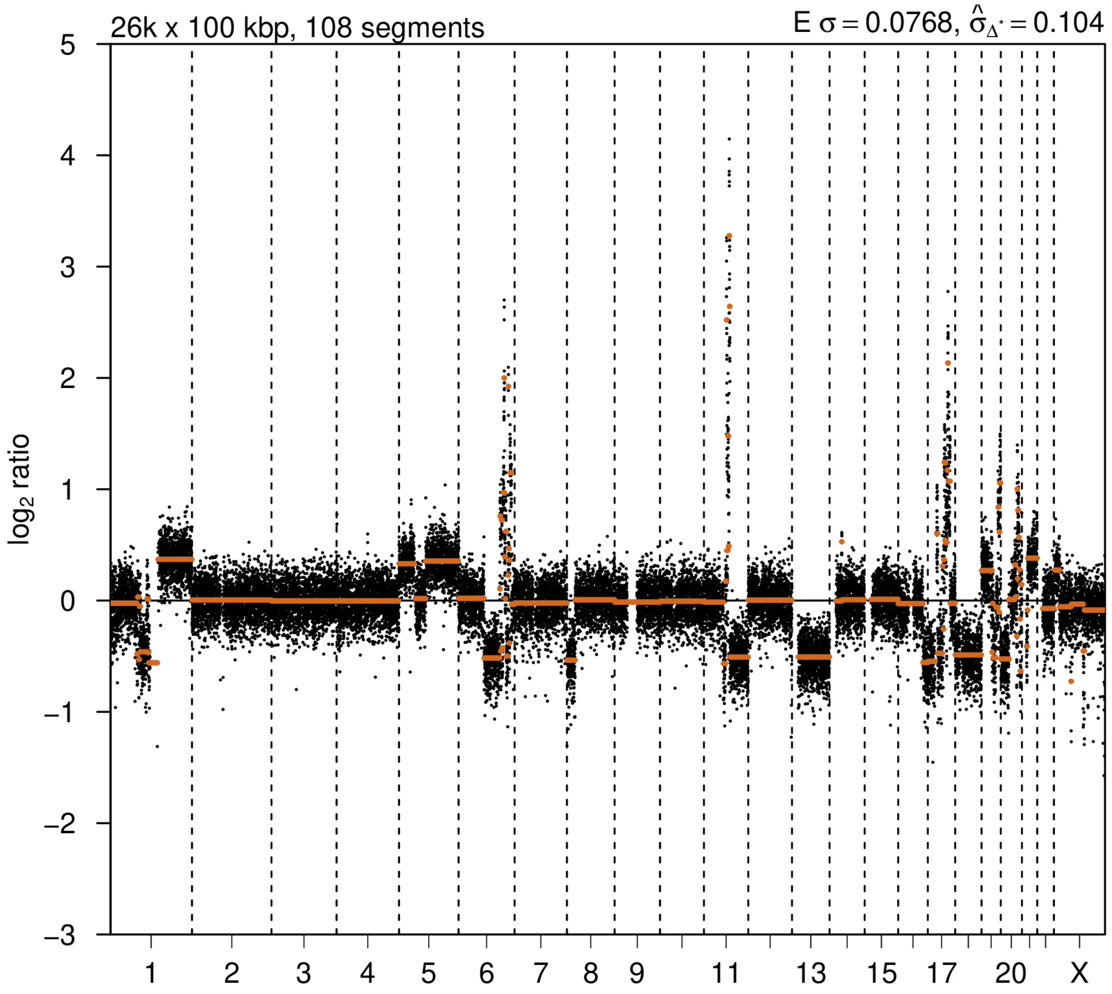

# P054

| Syn/Meta | Time from 1st surgery to 2nd event (Months) | Side 2nd event | Histology 2nd event | Surgery | AdjuvanT Treatment Pri (RT/ HT) | ER Pri | ER 2nd event | Her2 Pri | Her2 2nd event | Grade Pri | Grade 2nd event | Quadrant 2nd event | Margins | Screening | Clonality P value Copy N | Clonality P value Panel seq | Clonality P value WES | Final verdict |
|---|---|---|---|---|---|---|---|---|---|---|---|---|---|---|---|---|---|---|
| metachronous | 63 | Ipsilateral | IDC with DCIS | lumpectomy | None | + | + | - | - | 1 | 3 | at or adjacent to primary | NA | screen-detected | 0.000324 57 | NA | NA | Related |

### Primary event

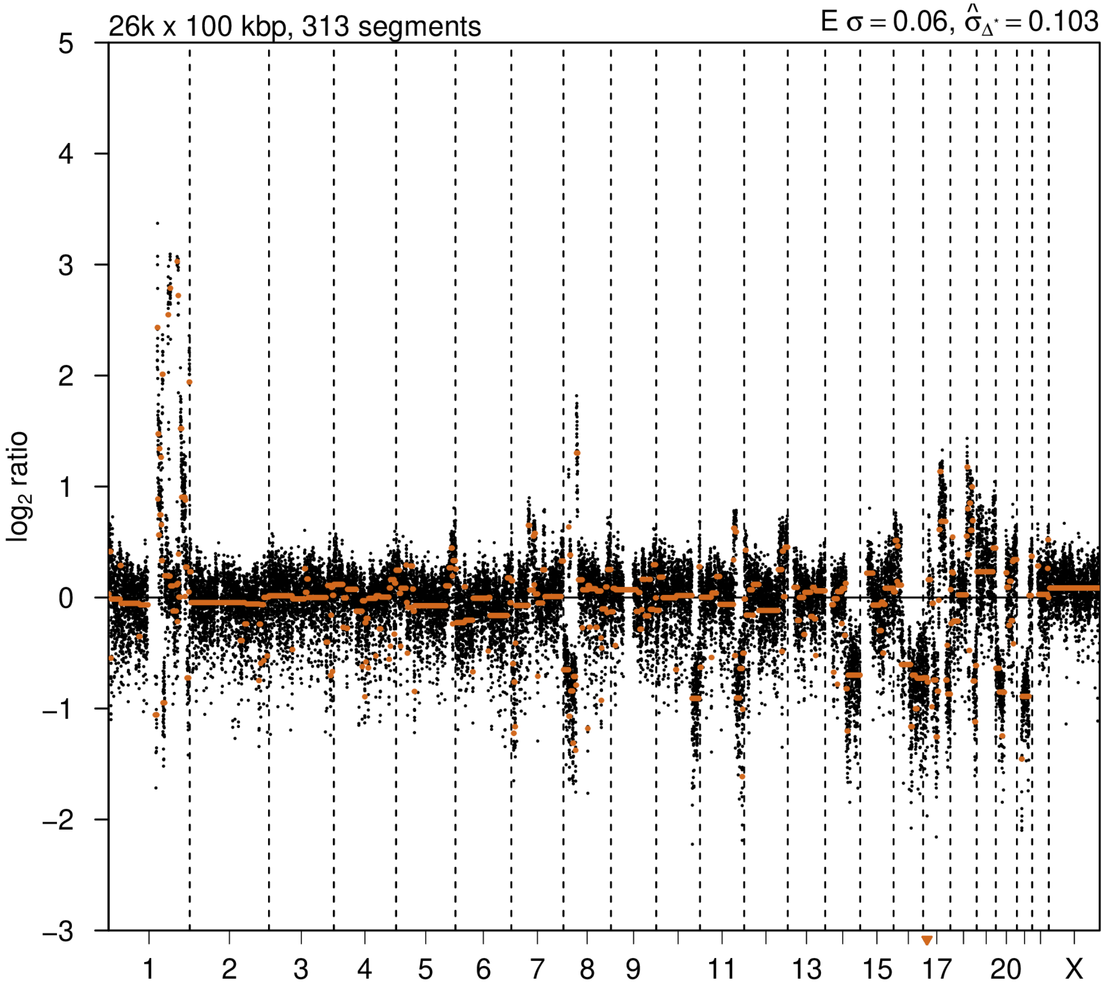


### 2nd event

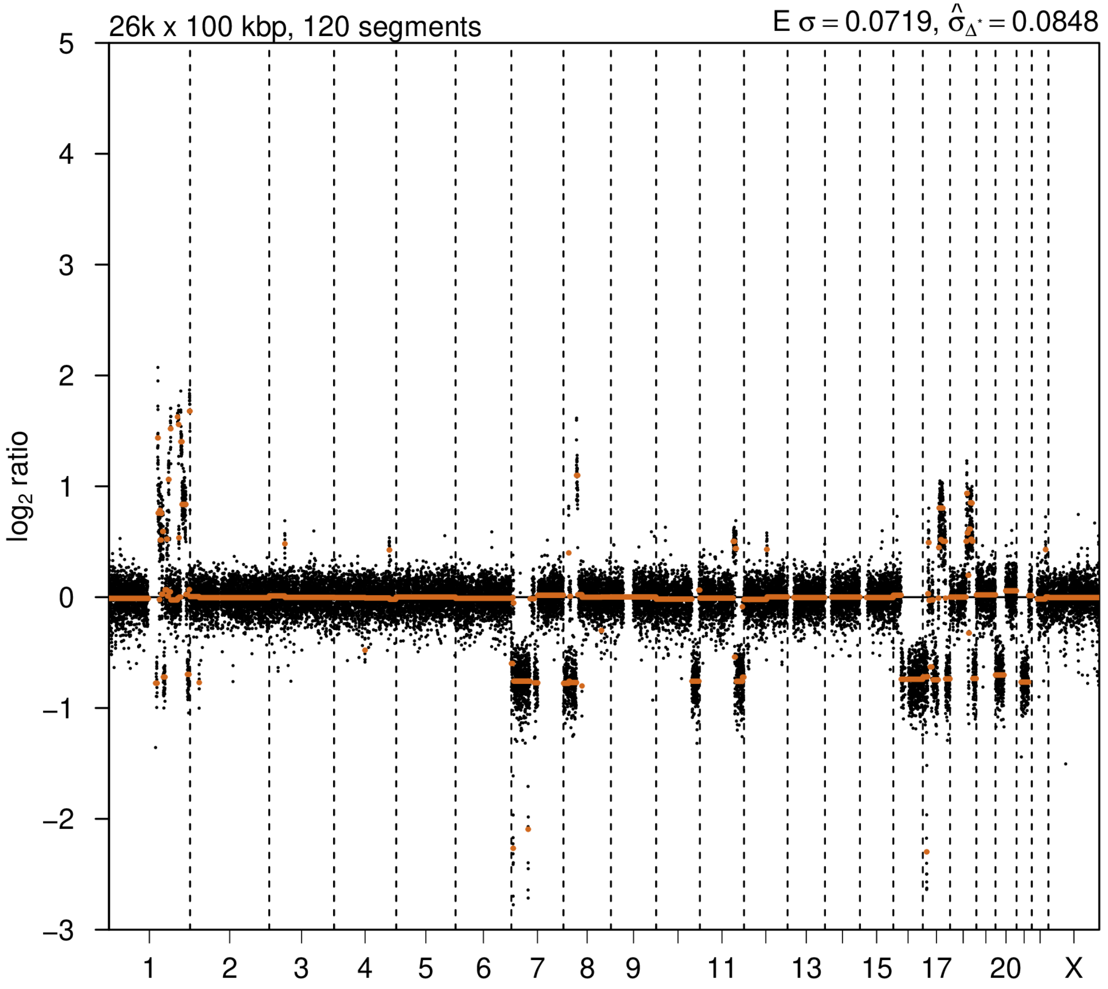

# P055

| Syn/Meta | Time from 1st surgery to 2nd event (Months) | Side 2nd event | Histology 2nd event | Surgery | AdjuvanT Treatment Pri (RT/ HT) | ER Pri | ER 2nd event | Her2 Pri | Her2 2nd event | Grade Pri | Grade 2nd event | Quadrant 2nd event | Margins | Screening | Clonality P value Copy N | Clonality P value Panel seq | Clonality P value WES | Final verdict |
|---|---|---|---|---|---|---|---|---|---|---|---|---|---|---|---|---|---|---|
| metachronous | 65 | Ipsilateral | IDC no DCIS | lumpectomy | None | + | + | - | - | 2 | 2 | at or adjacent to primary | Clear | symptomatic | 0.06134372 | 0.003 | NA | Related |

### Primary event

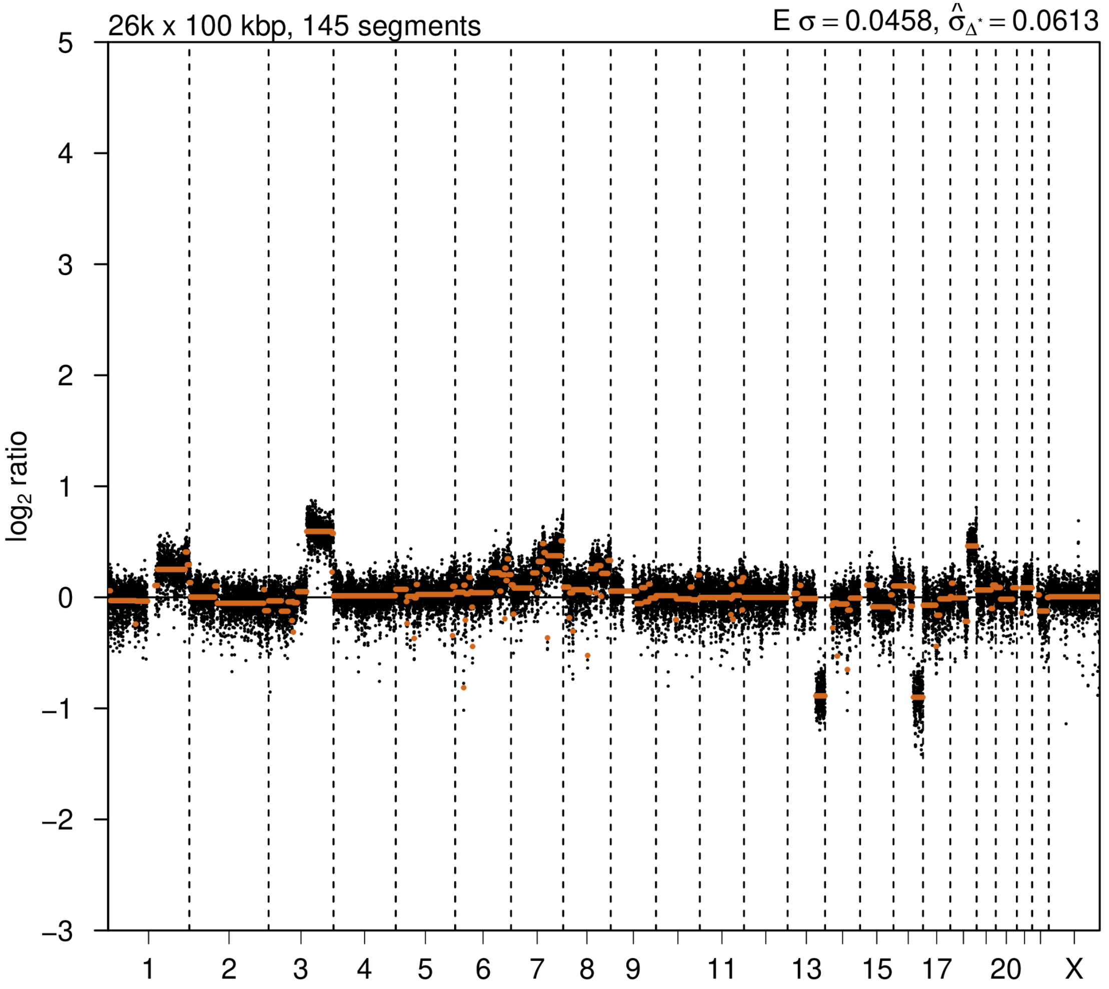


### 2nd event

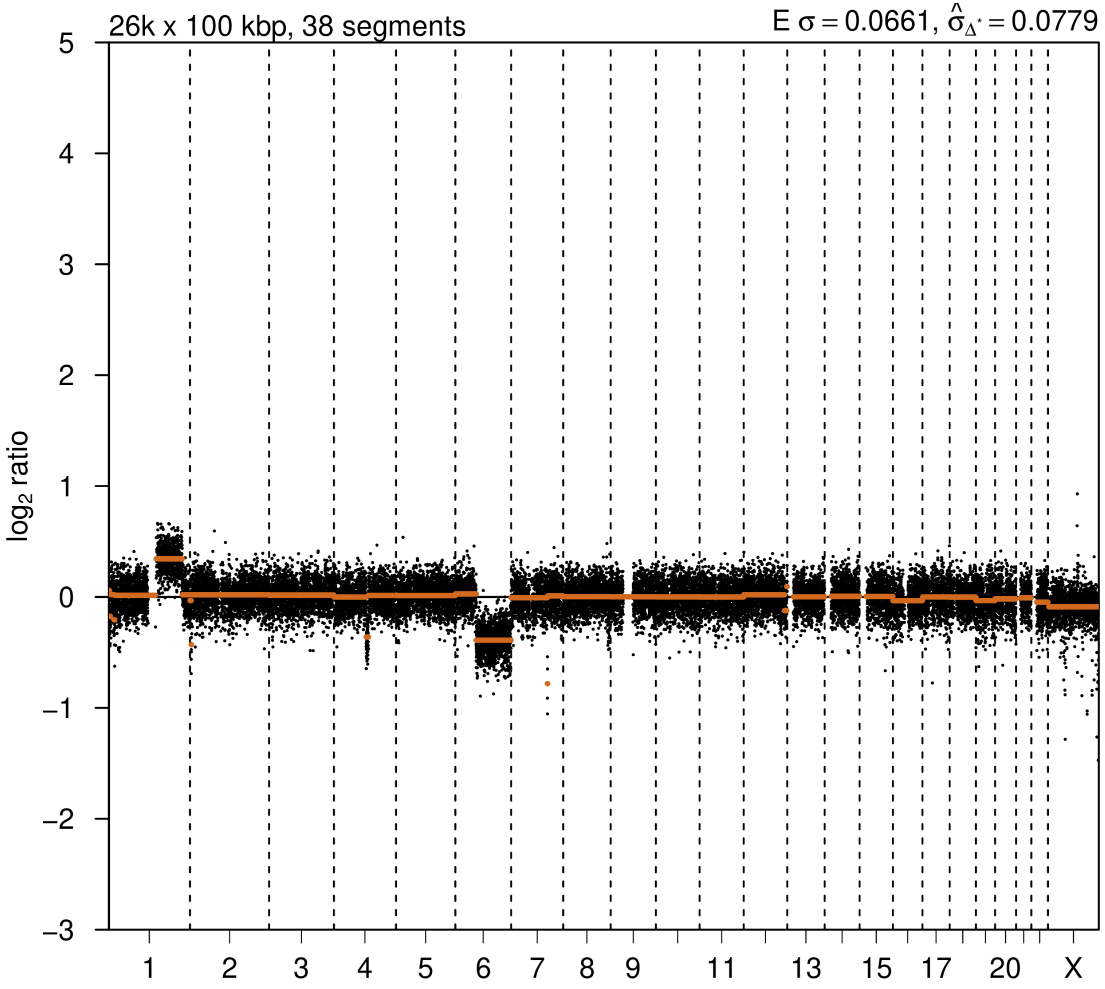

# P056

| Syn/Meta | Time from 1st surgery to 2nd event (Months) | Side 2nd event | Histology 2nd event | Surgery | AdjuvanT Treatment Pri (RT/ HT) | ER Pri | ER 2nd event | Her2 Pri | Her2 2nd event | Grade Pri | Grade 2nd event | Quadrant 2nd event | Margins | Screening | Clonality P value Copy N | Clonality P value Panel seq | Clonality P value WES | Final verdict |
|---|---|---|---|---|---|---|---|---|---|---|---|---|---|---|---|---|---|---|
| metachronous | 77 | Ipsilateral | IDC with DCIS | lumpectomy | None | - | + | - | - | 2 | 3 | at or adjacent to primary | NA | symptomatic | 0.000649 14 | Single mutation - shared | NA | Related |

**Primary event**

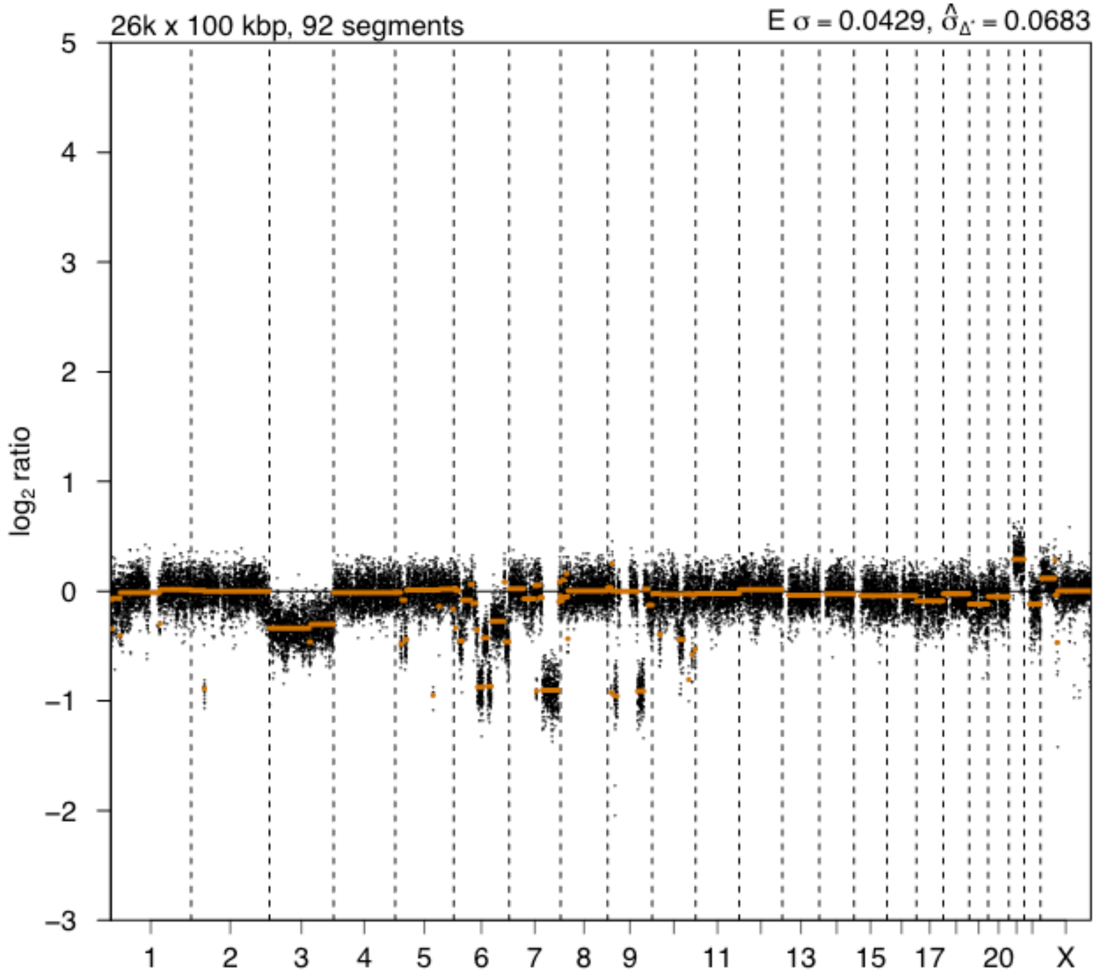


**2nd event**

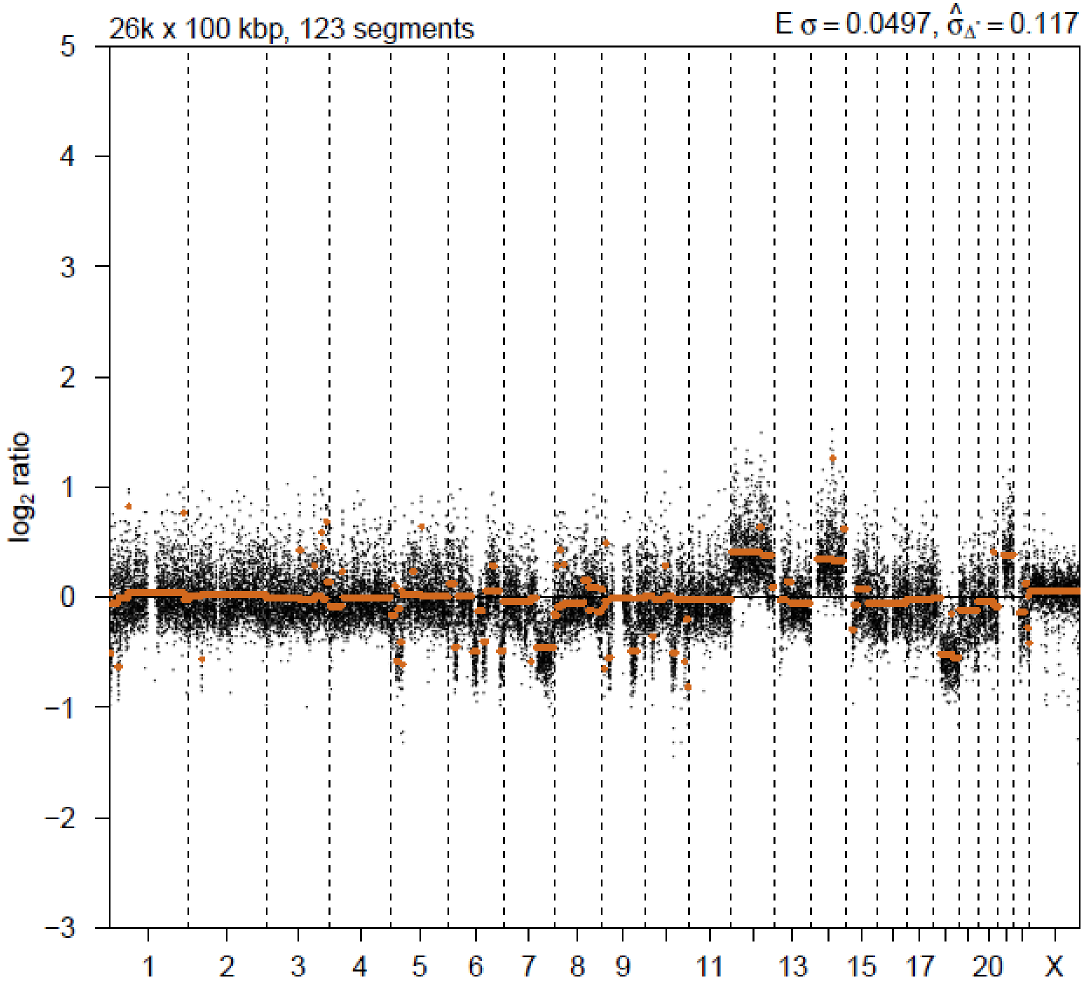

# P057

| Syn/Meta | Time from 1st surgery to 2nd event (Months) | Side 2nd event | Histology 2nd event | Surgery | AdjuvanT Treatment Pri (RT/ HT) | ER Pri | ER 2nd event | Her2 Pri | Her2 2nd event | Grade Pri | Grade 2nd event | Quadrant 2nd event | Margins | Screening | Clonality P value Copy N | Clonality P value Panel seq | Clonality P value WES | Final verdict |
|---|---|---|---|---|---|---|---|---|---|---|---|---|---|---|---|---|---|---|
| metachronous | 76 | Ipsilateral | IDC with DCIS | lumpectomy | None | + | + | + | + | 3 | 3 | at or adjacent to primary | involved | screen-detected | 0.000324 57 | Single mutation - shared | NA | Related |

### Primary event

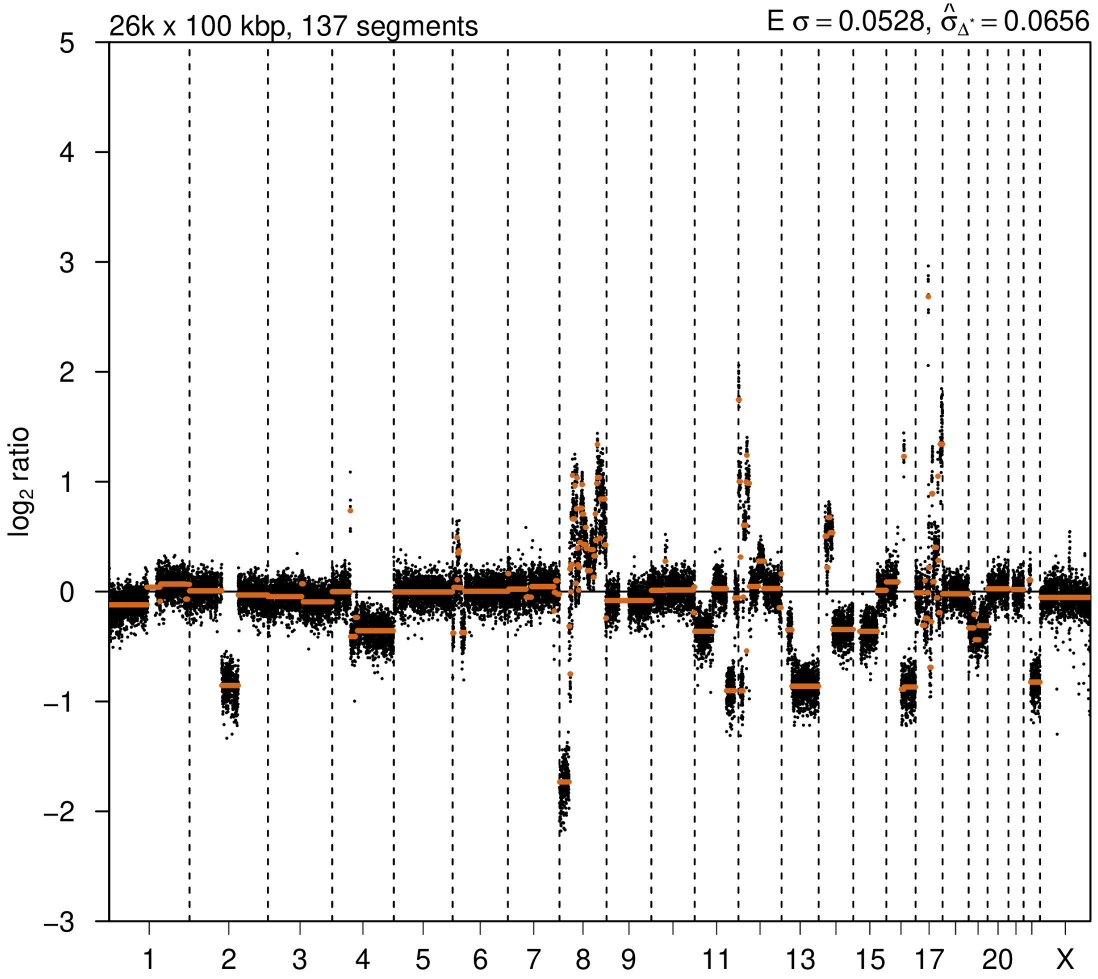


### 2nd event

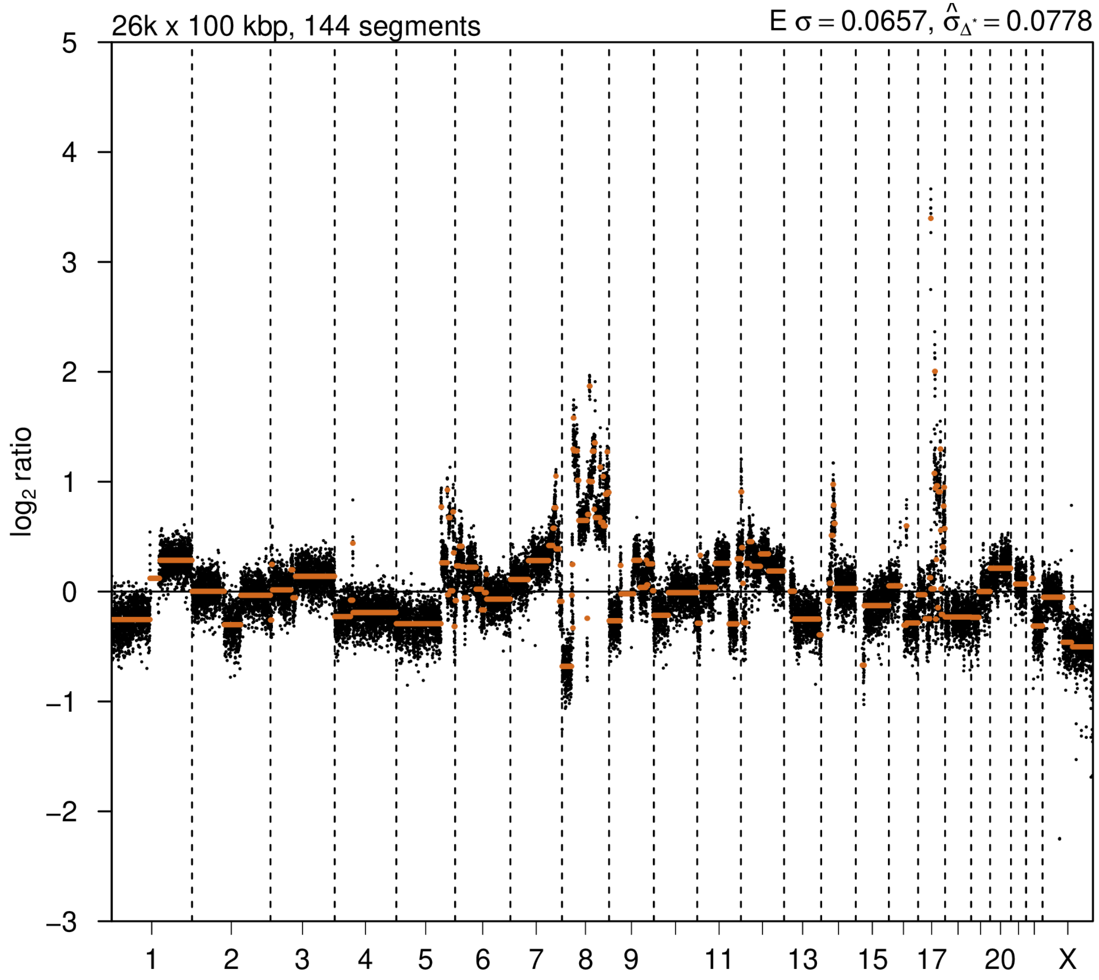

# P059

| Syn/Meta | Time from 1st surgery to 2nd event (Months) | Side 2nd event | Histology 2nd event | Surgery | AdjuvanT Treatment Pri (RT/ HT) | ER Pri | ER 2nd event | Her2 Pri | Her2 2nd event | Grade Pri | Grade 2nd event | Quadrant 2nd event | Margins | Screening | Clonality P value Copy N | Clonality P value Panel seq | Clonality P value WES | Final verdict |
|---|---|---|---|---|---|---|---|---|---|---|---|---|---|---|---|---|---|---|
| metachronous | 83 | Ipsilateral | IDC no DCIS | lumpectomy | None | + | + | - | + | 3 | 3 | at or adjacent to primary | Clear | screen-detected | 0.000649 14 | NA | NA | Related |

### Primary event

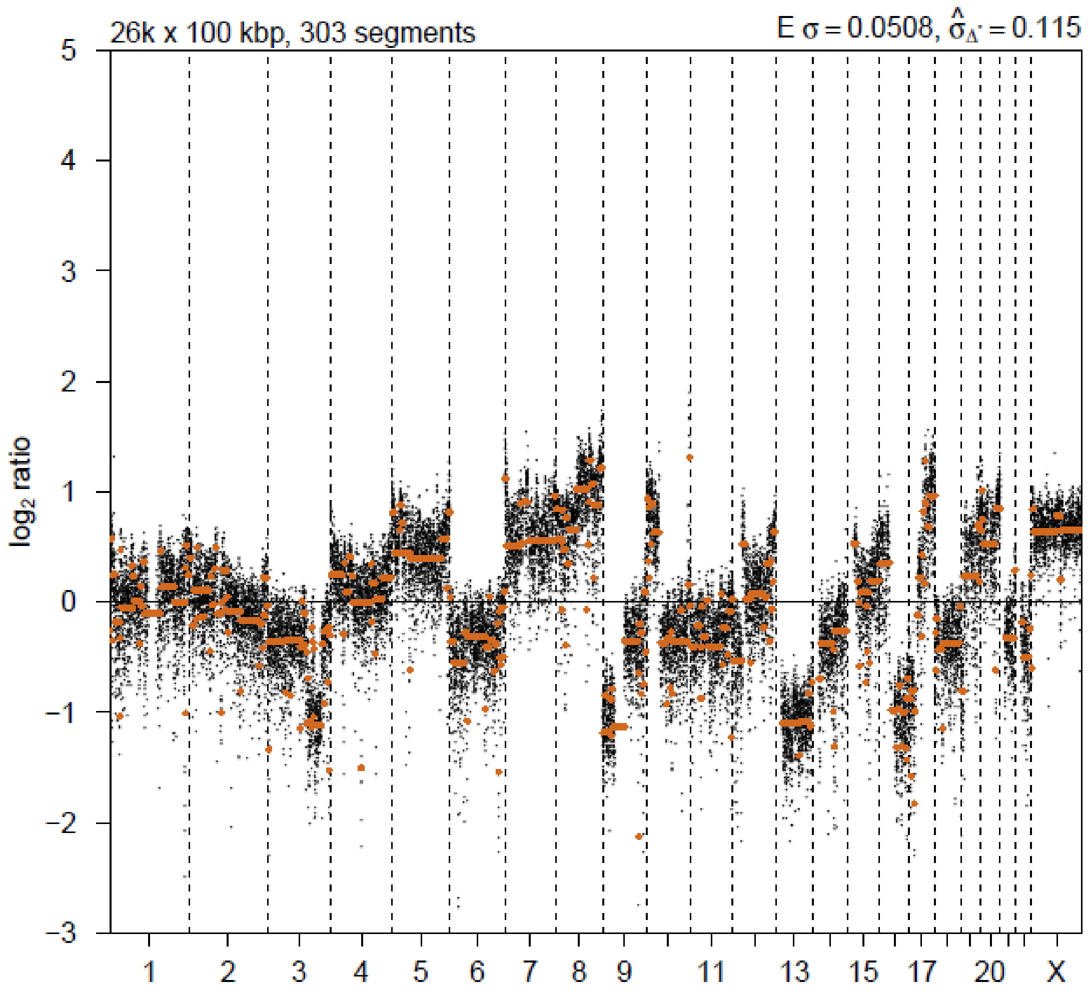


### 2nd event

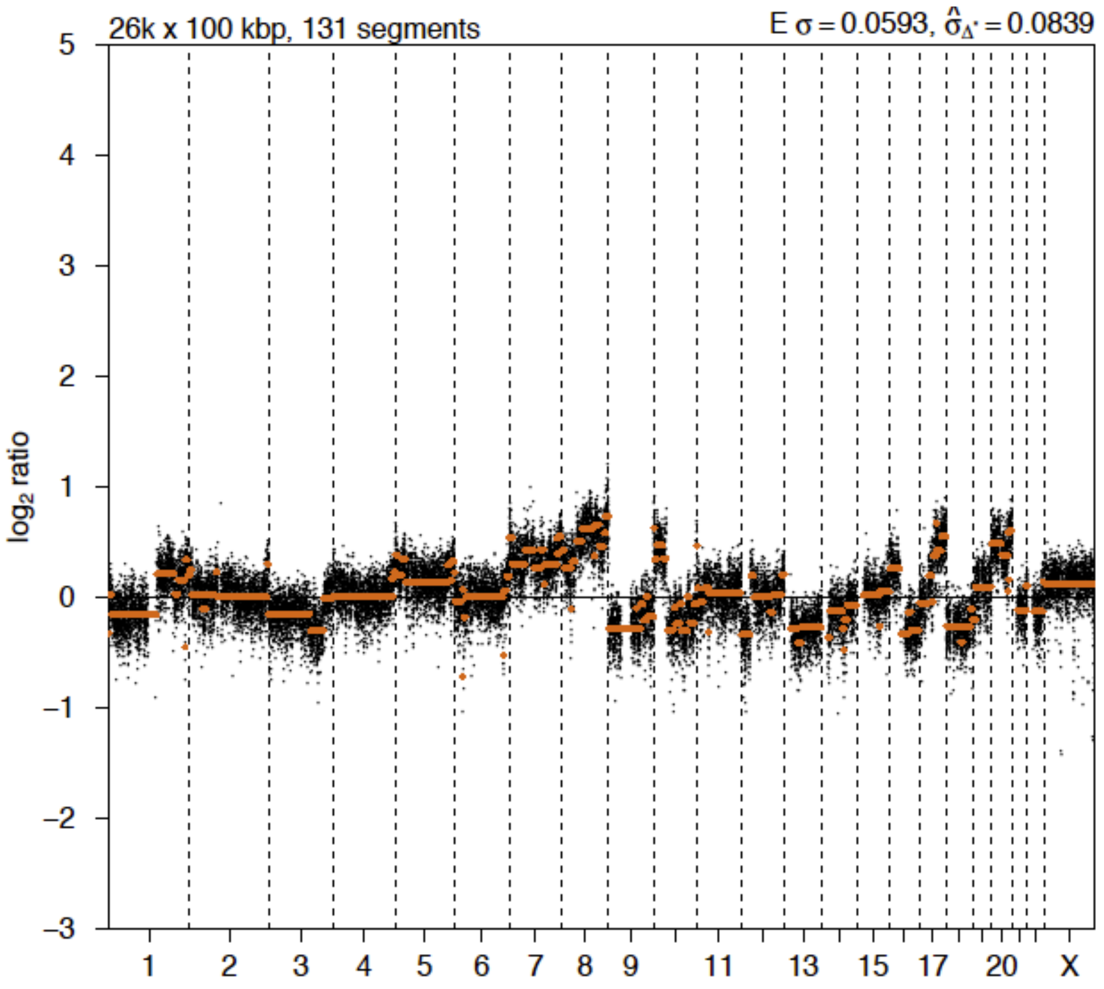

# P061

| Syn/Meta | Time from 1st surgery to 2nd event (Months) | Side 2nd event | Histology 2nd event | Surgery | AdjuvanT Treatment Pri (RT/ HT) | ER Pri | ER 2nd event | Her2 Pri | Her2 2nd event | Grade Pri | Grade 2nd event | Quadrant 2nd event | Margins | Screening | Clonality P value Copy N | Clonality P value Panel seq | Clonality P value WES | Final verdict |
|---|---|---|---|---|---|---|---|---|---|---|---|---|---|---|---|---|---|---|
| metachronous | 41 | Ipsilateral | IDC with DCIS | lumpectomy | None | + | + | - | - | 2 | 2 | NA | Clear | symptomatic | 0.00032457 | 0.003 | NA | Related |

### Primary event

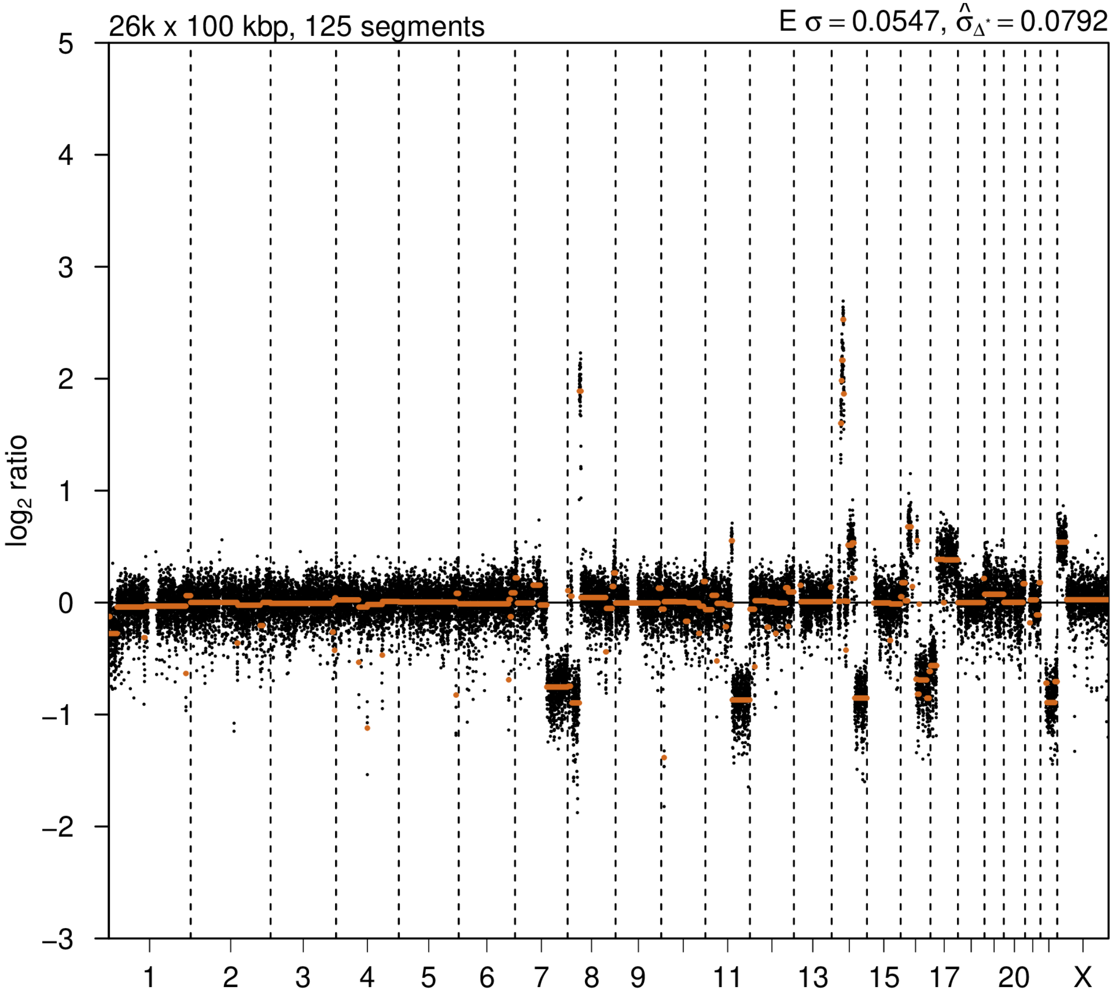


### 2nd event

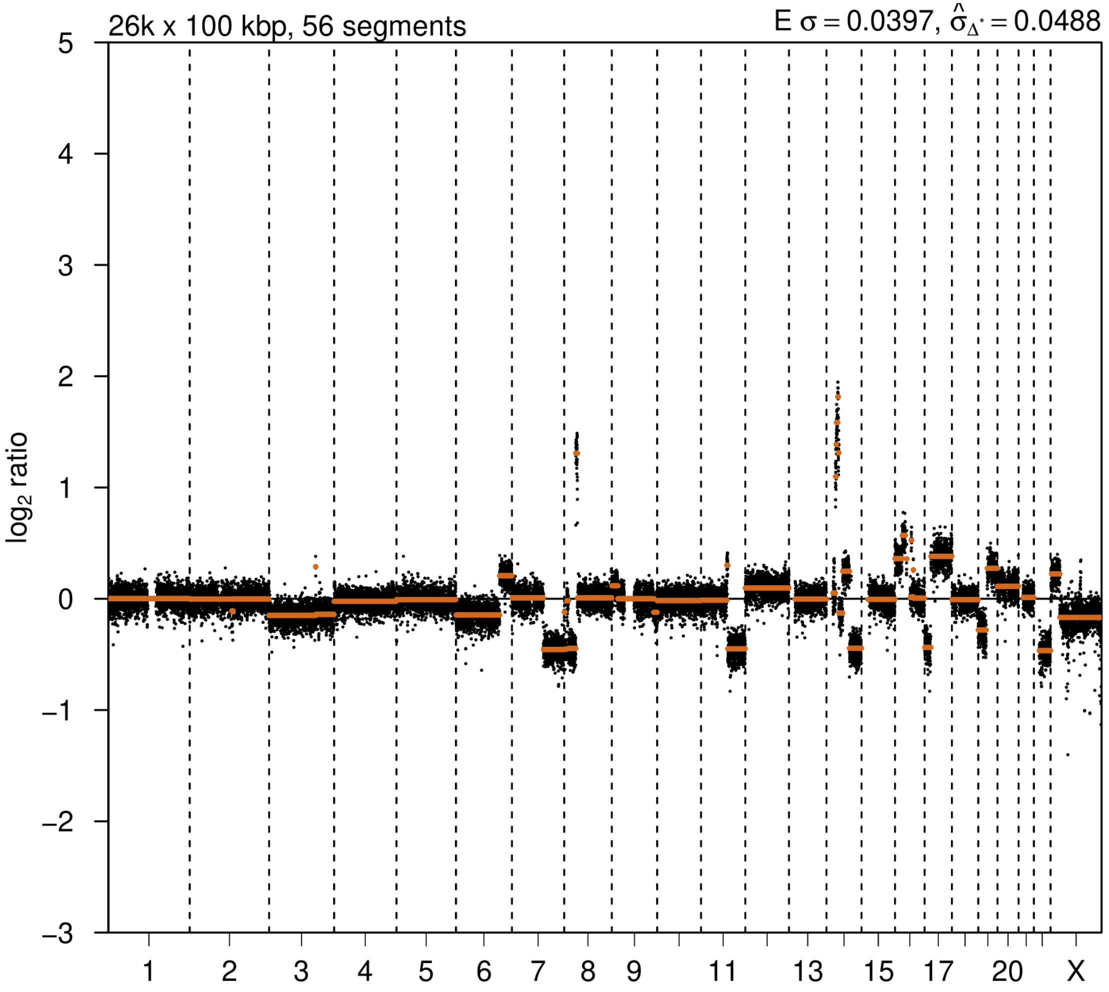

# P063

| Syn/Meta | Time from 1st surgery to 2nd event (Months) | Side 2nd event | Histology 2nd event | Surgery | AdjuvanT Treatment Pri (RT/ HT) | ER Pri | ER 2nd event | Her2 Pri | Her2 2nd event | Grade Pri | Grade 2nd event | Quadrant 2nd event | Margins | Screening | Clonality P value Copy N | Clonality P value Panel seq | Clonality P value WES | Final verdict |
|---|---|---|---|---|---|---|---|---|---|---|---|---|---|---|---|---|---|---|
| metachronous | 37 | Ipsilateral | IDC with DCIS | lumpectomy | None | + | + | - | - | 3 | 1 | NA | involved | screen-detected | 0.000324 57 | NA | NA | Related |

### Primary event

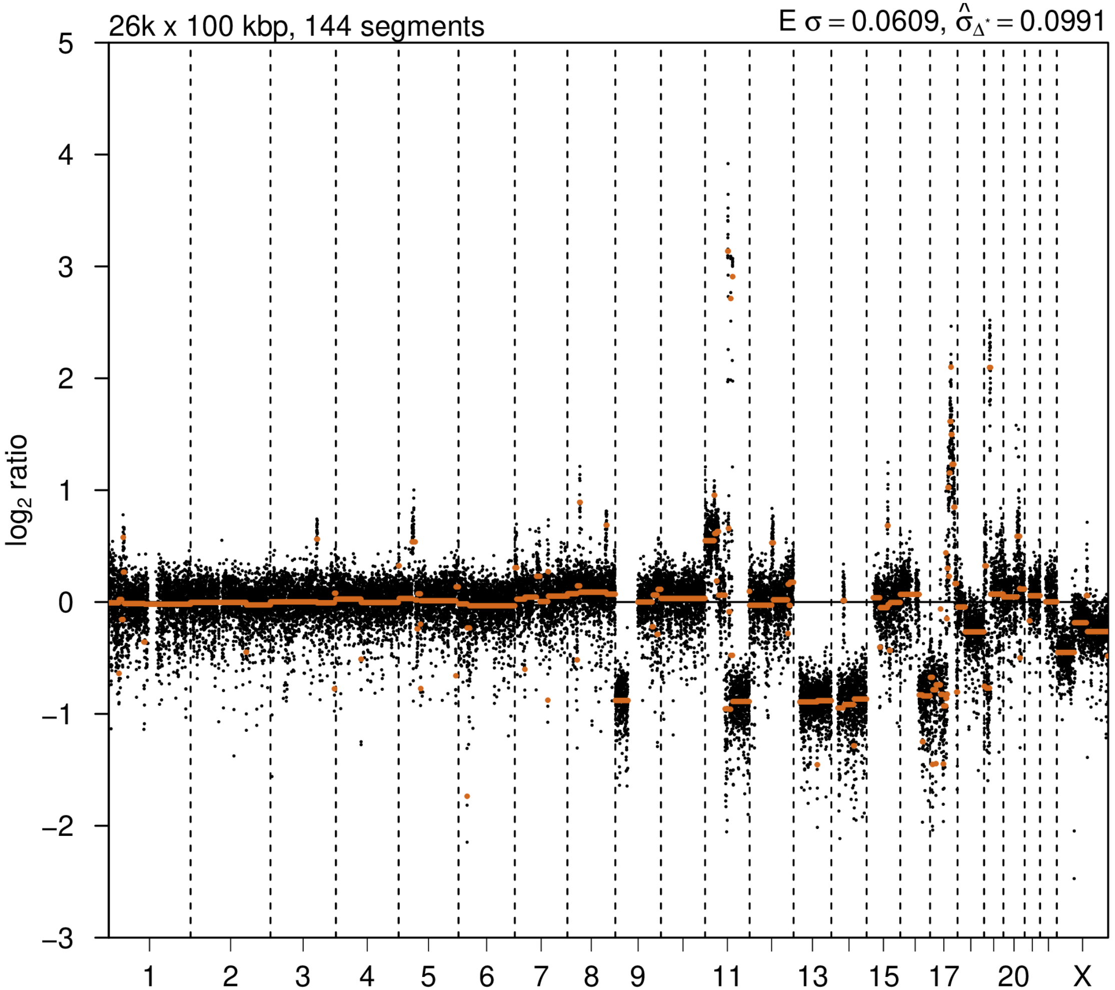


### 2nd event

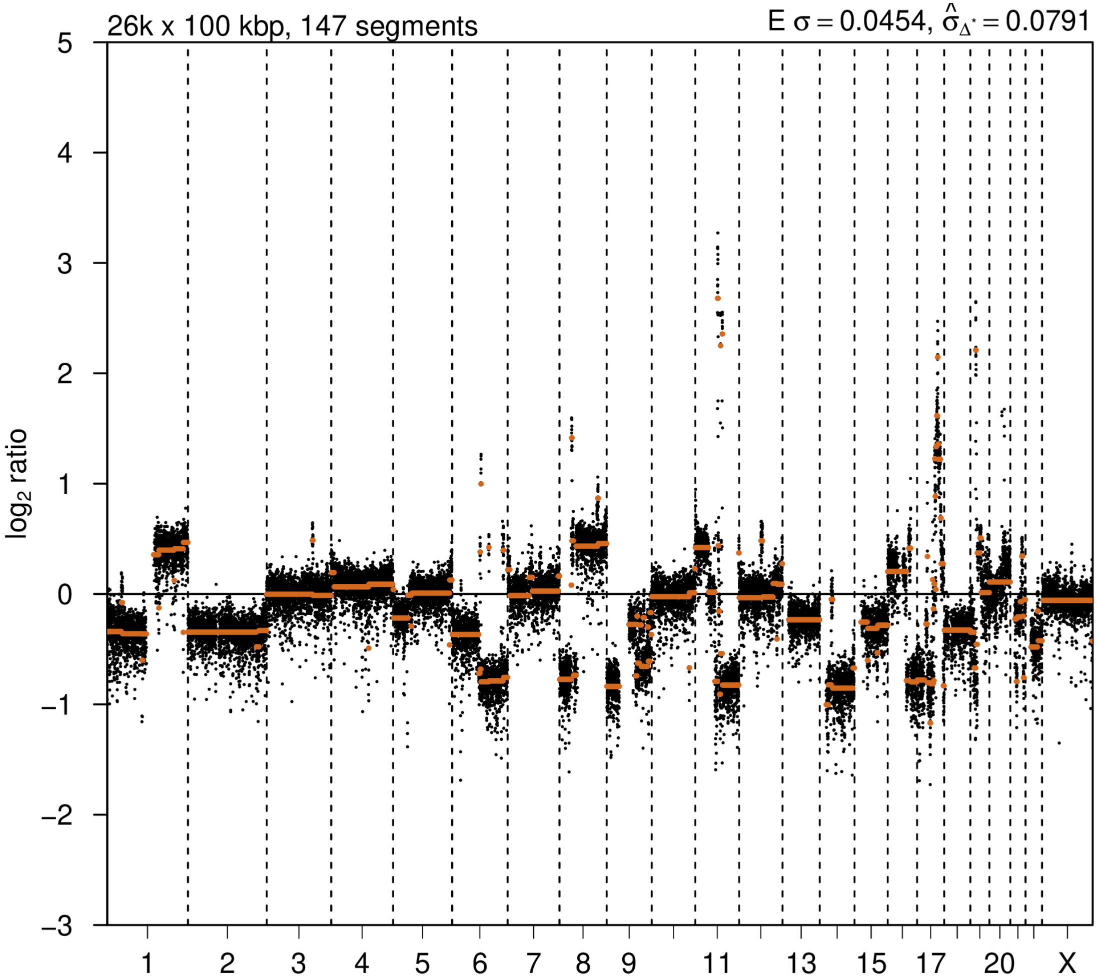

# P065

| Syn/Meta | Time from 1st surgery to 2nd event (Months) | Side 2nd event | Histology 2nd event | Surgery | AdjuvanT Treatment Pri (RT/ HT) | ER Pri | ER 2nd event | Her2 Pri | Her2 2nd event | Grade Pri | Grade 2nd event | Quadrant 2nd event | Margins | Screening | Clonality P value Copy N | Clonality P value Panel seq | Clonality P value WES | Final verdict |
|---|---|---|---|---|---|---|---|---|---|---|---|---|---|---|---|---|---|---|
| metachronous | 44 | Ipsilateral | IDC with DCIS | lumpectomy | None | - | - | + | + | 2 | 1 | at or adjacent to primary | involved | screen-detected | 0.000324 57 | NA | NA | Related |

### Primary event

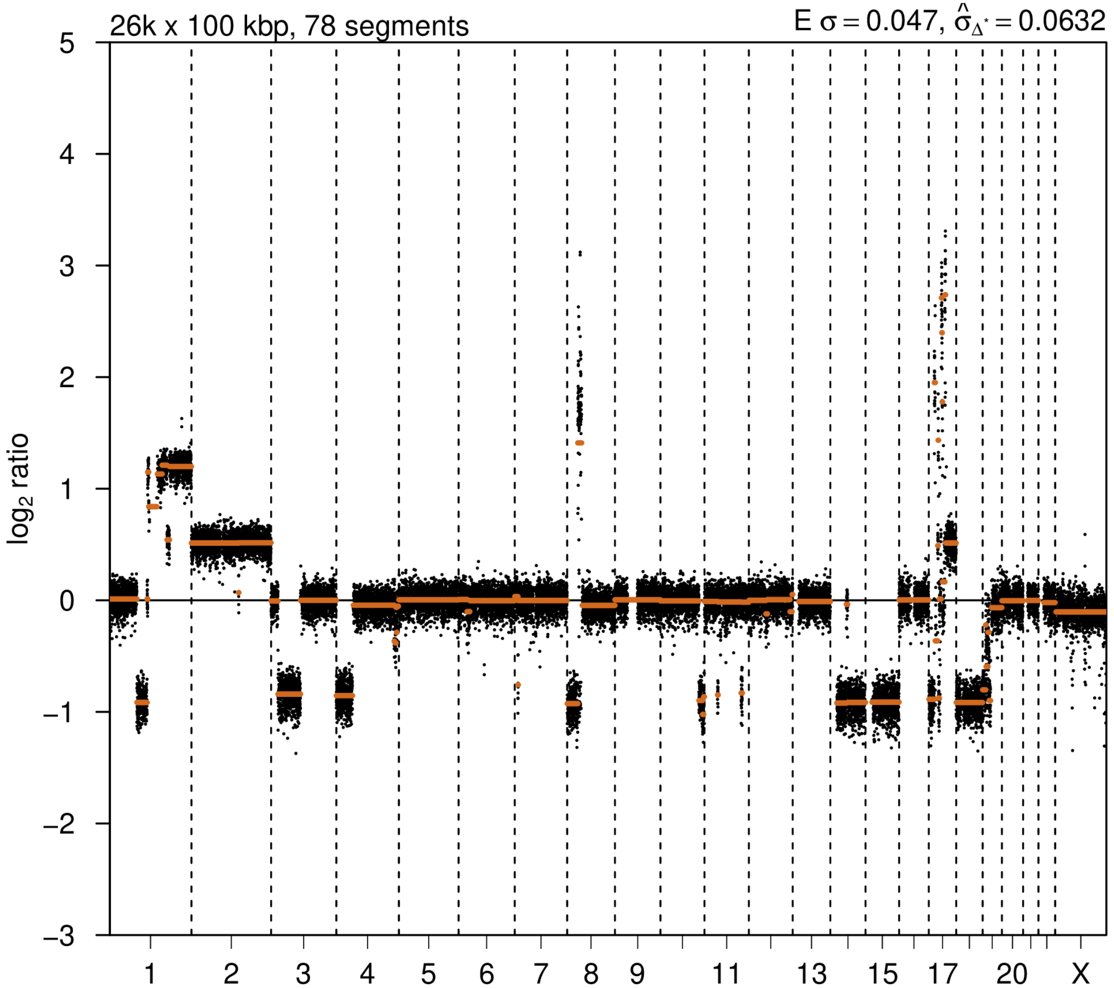


### 2nd event

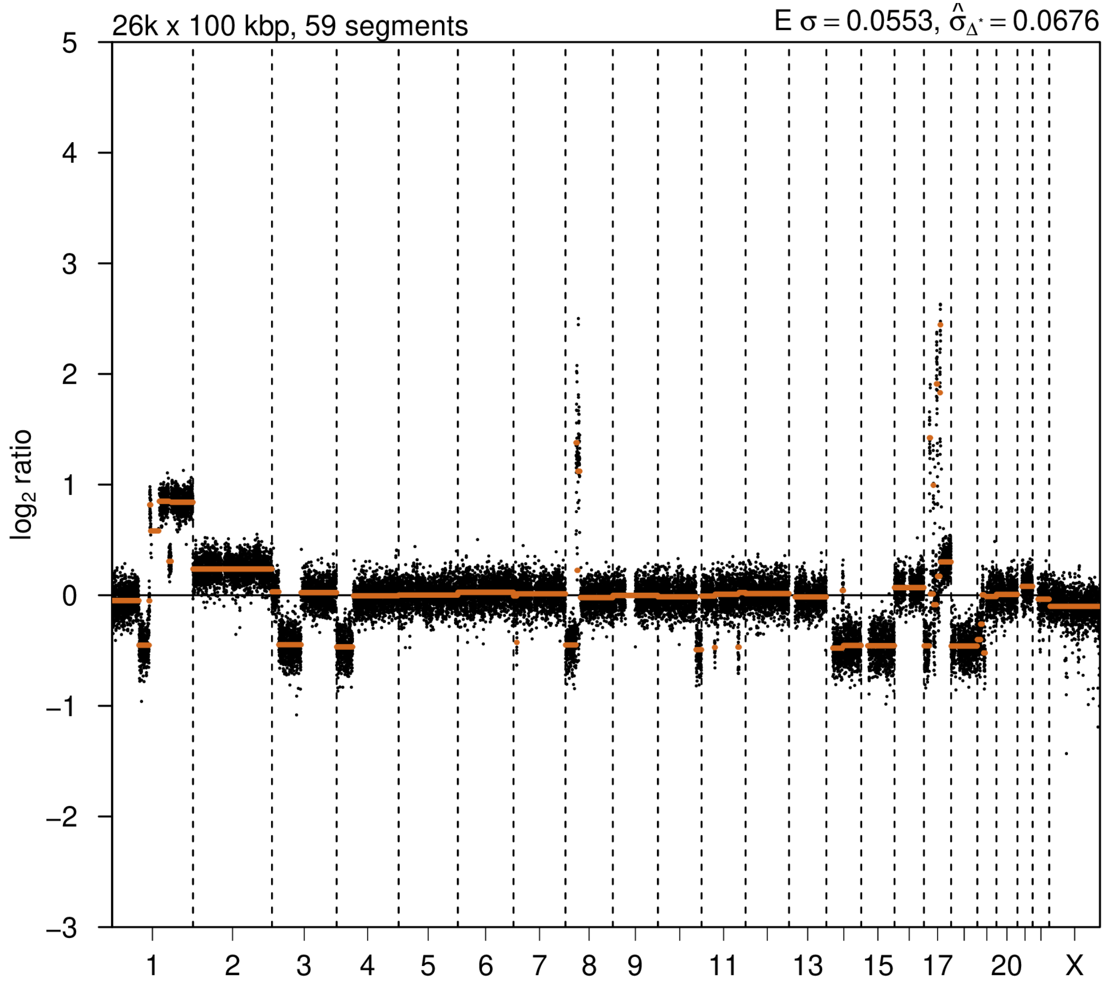

# P066

| Syn/Meta | Time from 1st surgery to 2nd event (Months) | Side 2nd event | Histology 2nd event | Surgery | AdjuvanT Treatment Pri (RT/ HT) | ER Pri | ER 2nd event | Her2 Pri | Her2 2nd event | Grade Pri | Grade 2nd event | Quadrant 2nd event | Margins | Screening | Clonality P value Copy N | Clonality P value Panel seq | Clonality P value WES | Final verdict |
|---|---|---|---|---|---|---|---|---|---|---|---|---|---|---|---|---|---|---|
| metachronous | 50 | Ipsilateral | IDC with DCIS | lumpectomy | None | + | + | - | - | 1 | 2 | at or adjacent to primary | Clear | screen-detected | 0.000324 57 | Single mutation - shared | NA | Related |

### Primary event

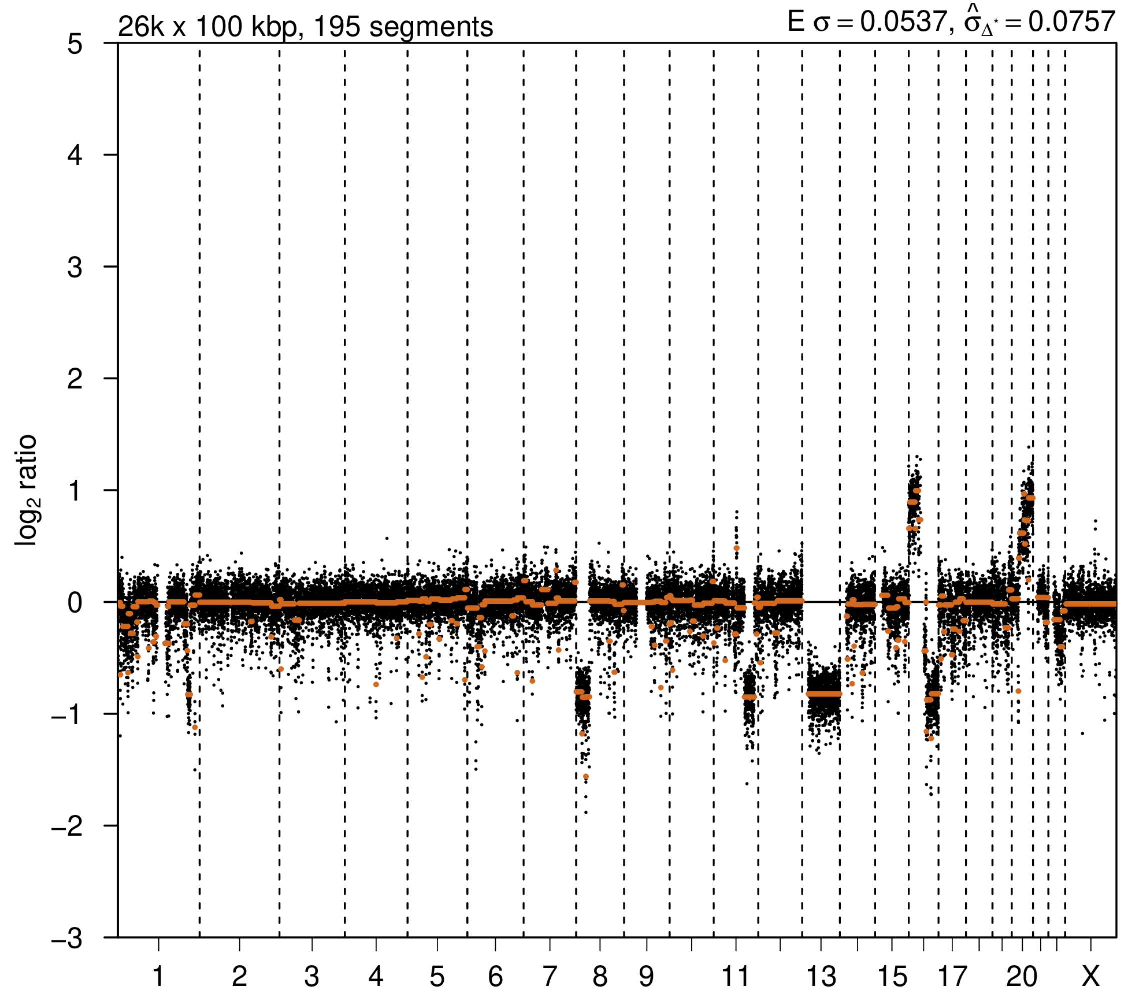


### 2nd event

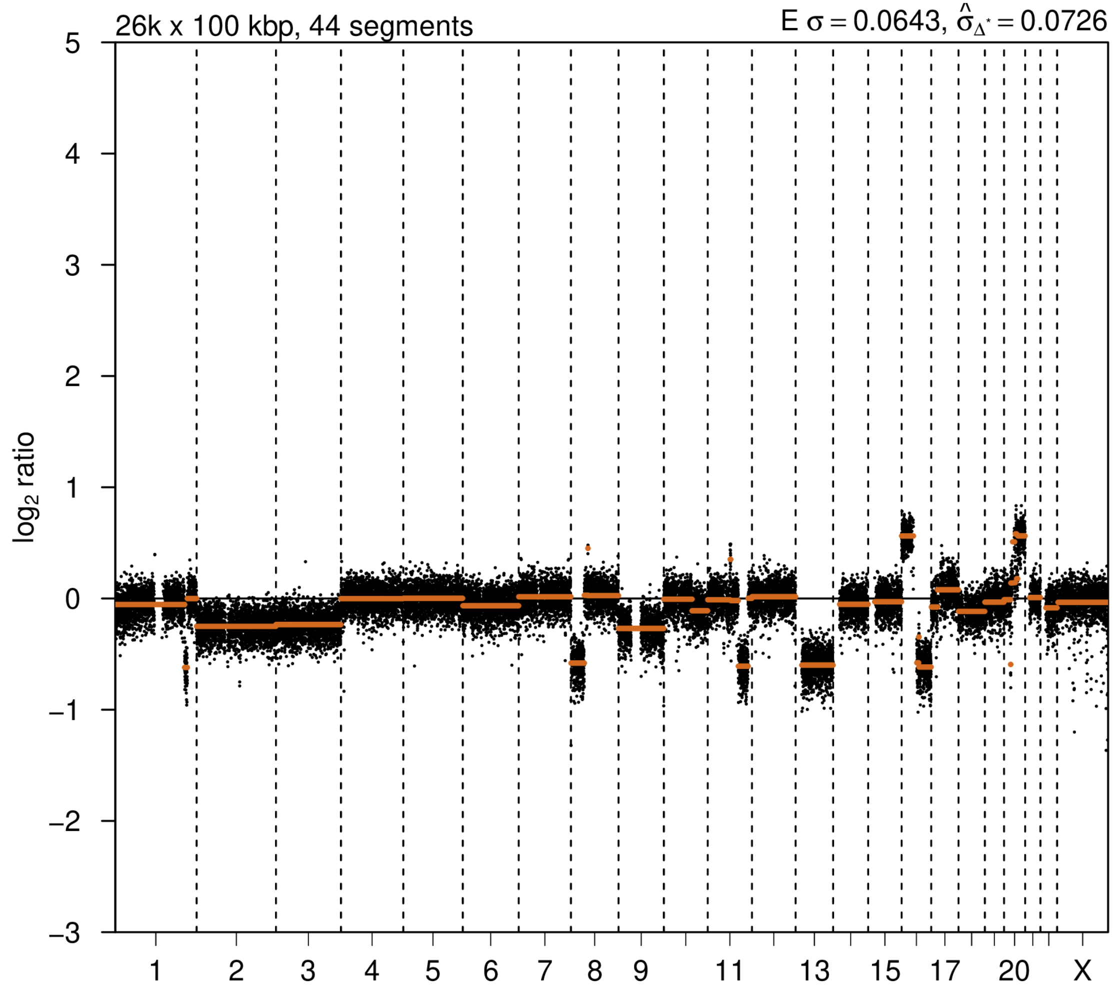

# P067

| Syn/Meta | Time from 1st surgery to 2nd event (Months) | Side 2nd event | Histology 2nd event | Surgery | AdjuvanT Treatment Pri (RT/ HT) | ER Pri | ER 2nd event | Her2 Pri | Her2 2nd event | Grade Pri | Grade 2nd event | Quadrant 2nd event | Margins | Screening | Clonality P value Copy N | Clonality P value Panel seq | Clonality P value WES | Final verdict |
|---|---|---|---|---|---|---|---|---|---|---|---|---|---|---|---|---|---|---|
| metachronous | 33 | Ipsilateral | IDC with DCIS | lumpectomy | None | + | + | - | - | 2 | 2 | at or adjacent to primary | NA | symptomatic | 0.00032457 | Single mutation - shared | NA | Related |

### Primary event

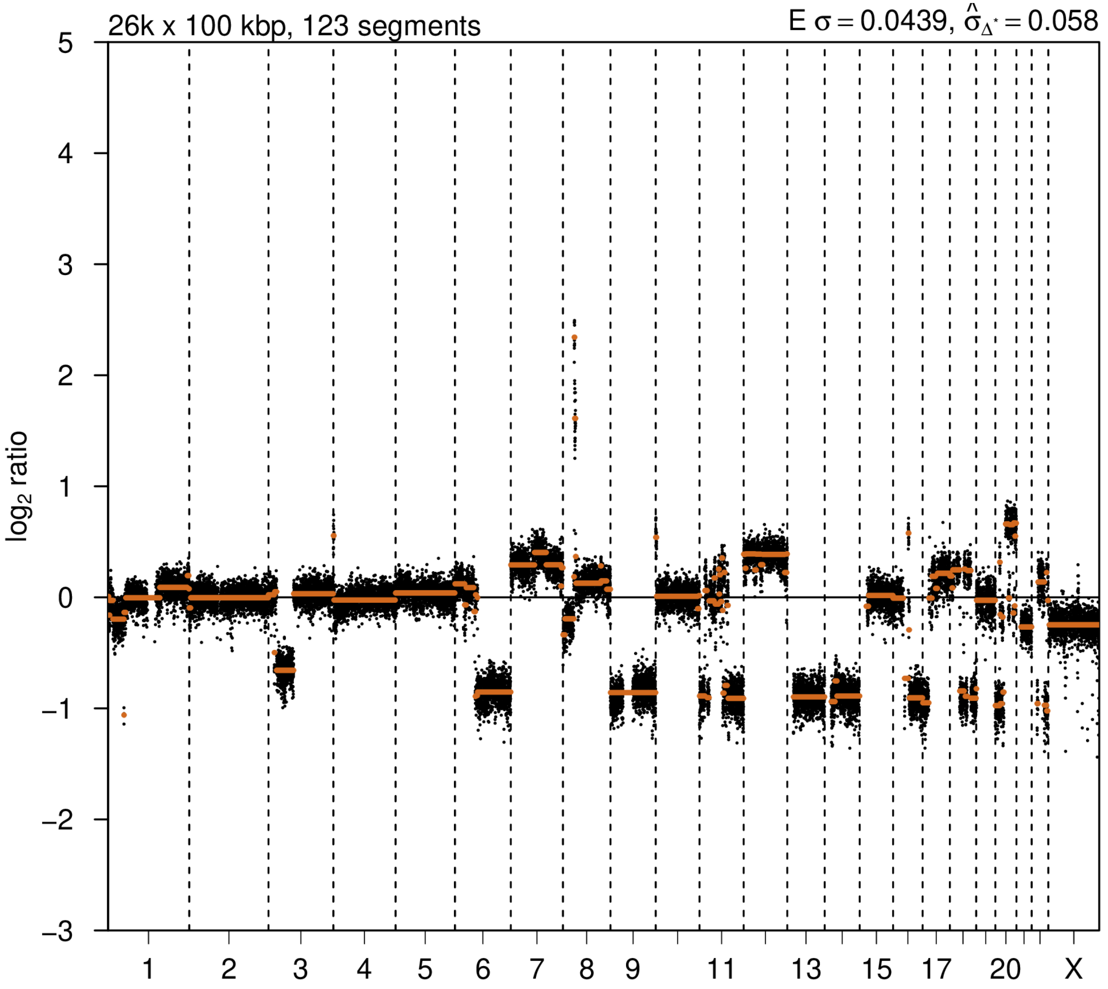


### 2nd event

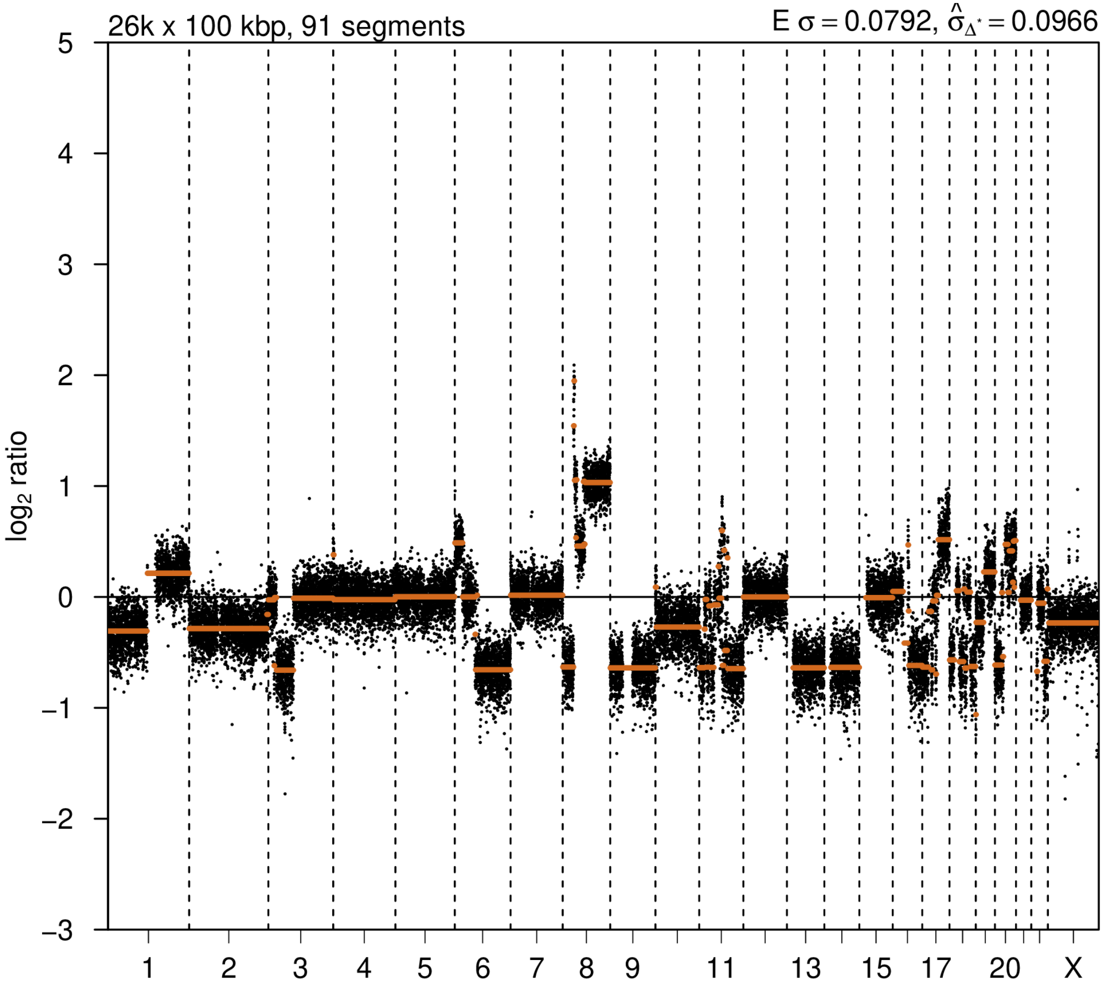

# P068

| Syn/Meta | Time from 1st surgery to 2nd event (Months) | Side 2nd event | Histology 2nd event | Surgery | AdjuvanT Treatment Pri (RT/ HT) | ER Pri | ER 2nd event | Her2 Pri | Her2 2nd event | Grade Pri | Grade 2nd event | Quadrant 2nd event | Margins | Screening | Clonality P value Copy N | Clonality P value Panel seq | Clonality P value WES | Final verdict |
|---|---|---|---|---|---|---|---|---|---|---|---|---|---|---|---|---|---|---|
| metachronous | 50 | Ipsilateral | IDC with DCIS | lumpectomy | None | + | + | - | - | 2 | 2 | at or adjacent to primary | Clear | screen-detected | 0.000324 57 | NA | 0.00075244 5 | Related |

### Primary event

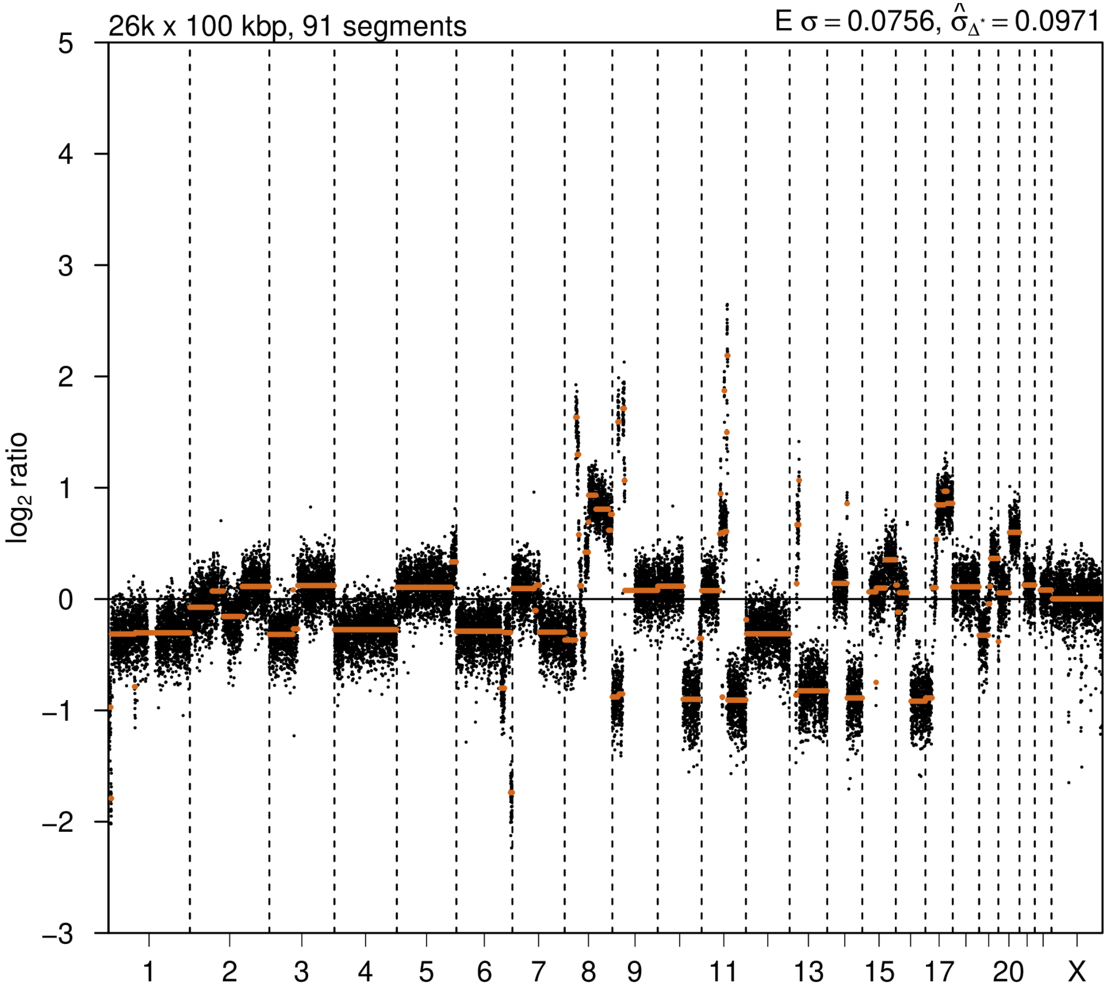


### 2nd event

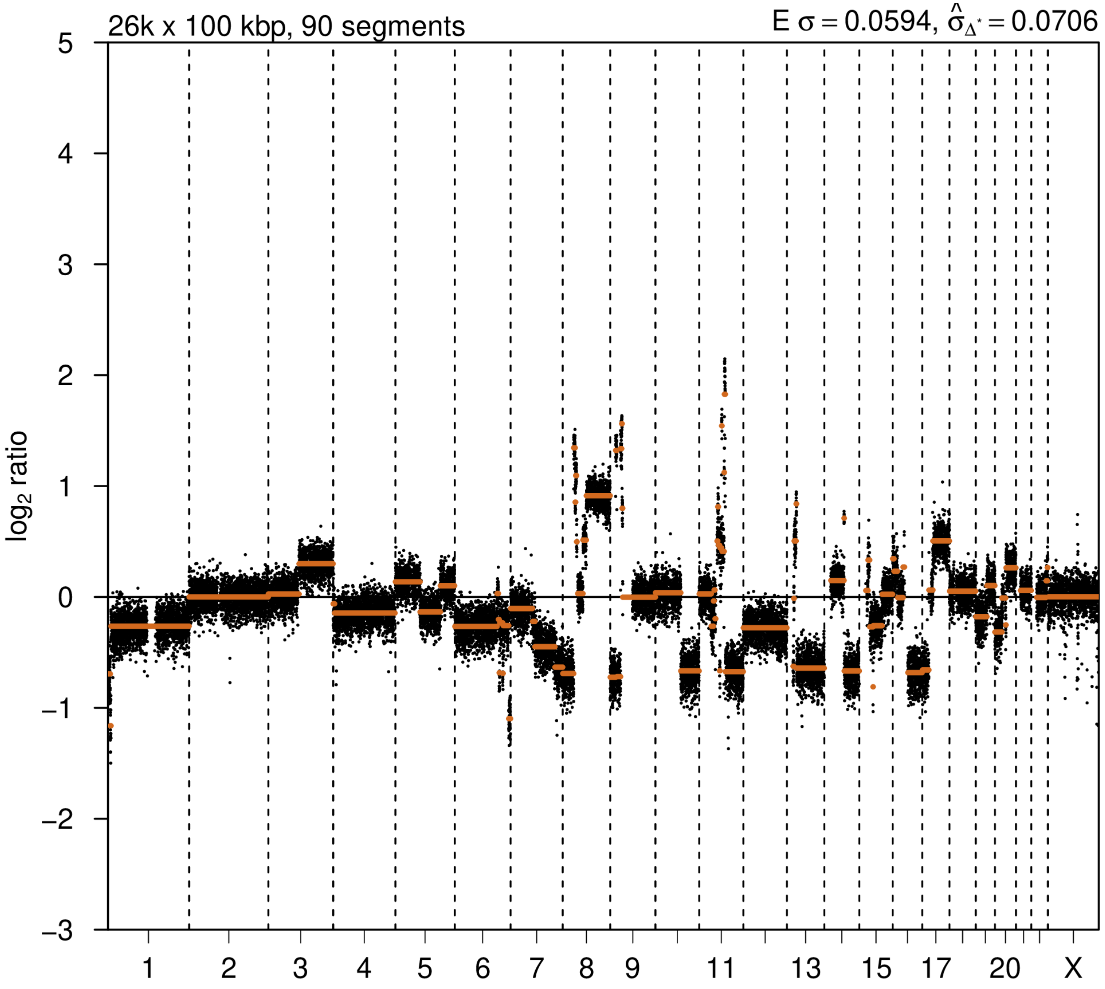

# P069

| Syn/Meta | Time from 1st surgery to 2nd event (Months) | Side 2nd event | Histology 2nd event | Surgery | AdjuvanT Treatment Pri (RT/ HT) | ER Pri | ER 2nd event | Her2 Pri | Her2 2nd event | Grade Pri | Grade 2nd event | Quadrant 2nd event | Margins | Screening | Clonality P value Copy N | Clonality P value Panel seq | Clonality P value WES | Final verdict |
|---|---|---|---|---|---|---|---|---|---|---|---|---|---|---|---|---|---|---|
| metachronous | 35 | Ipsilateral | IDC no DCIS | lumpectomy | None | + | + | + | + | 3 | 3 | at or adjacent to primary | Clear | symptomatic | 0.00032457 | 0.003 | NA | Related |

### Primary event

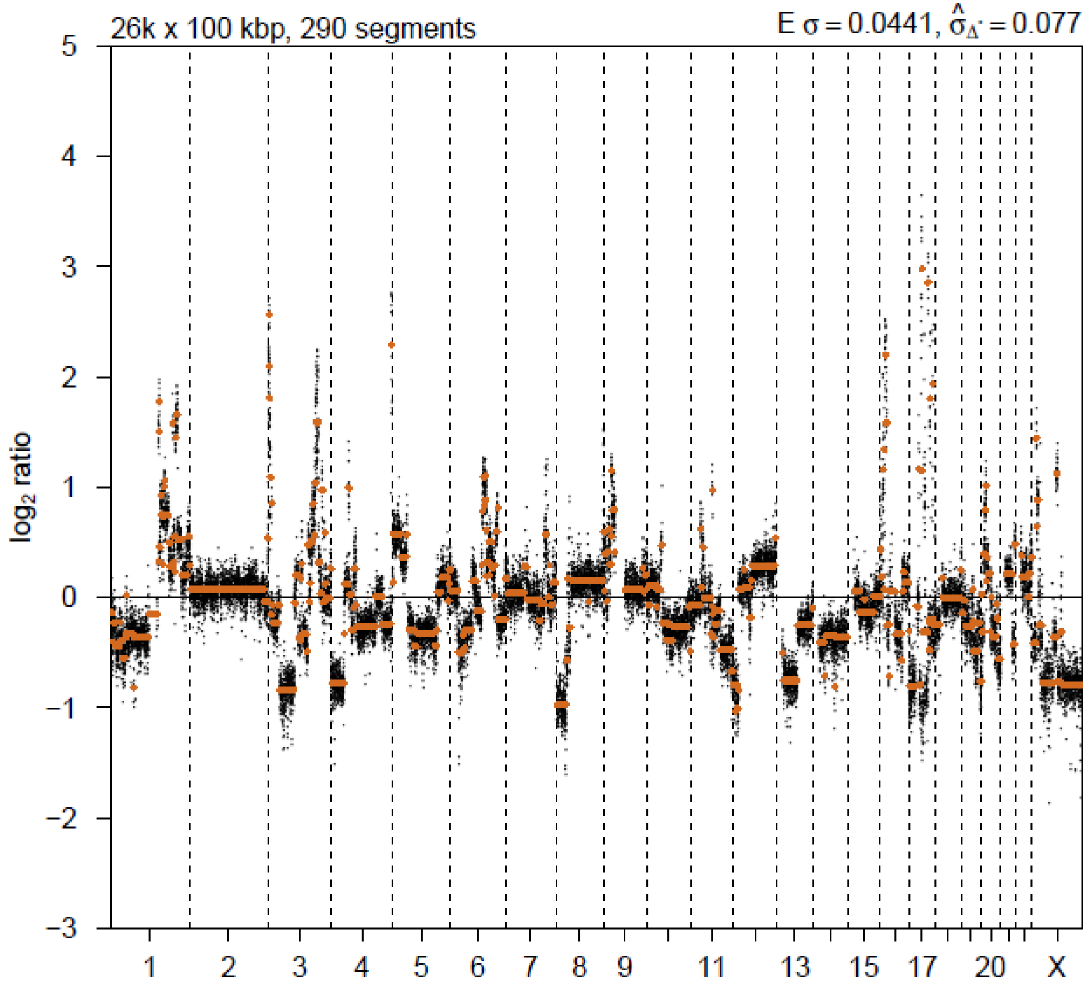


### 2nd event

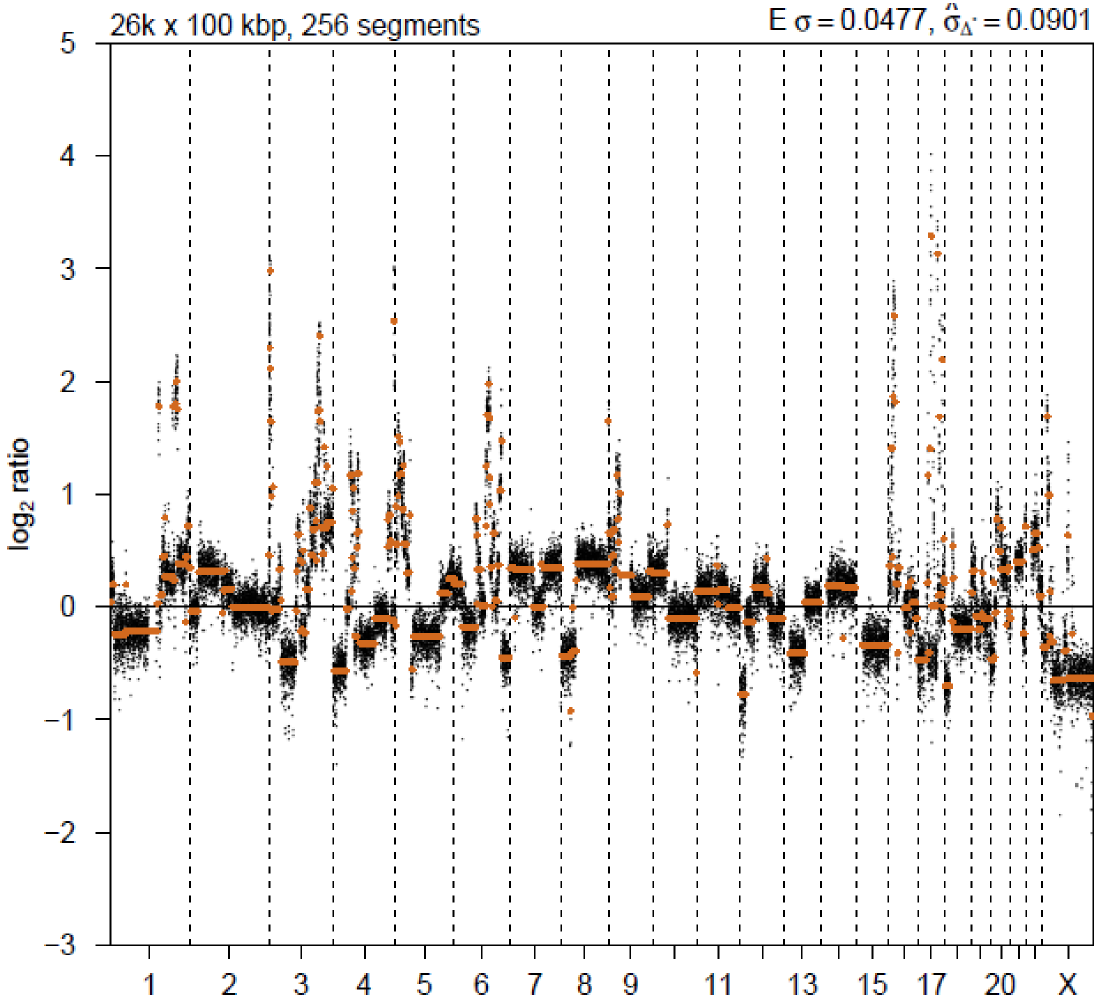

# P072

| Syn/Meta | Time from 1st surgery to 2nd event (Months) | Side 2nd event | Histology 2nd event | Surgery | AdjuvanT Treatment Pri (RT/ HT) | ER Pri | ER 2nd event | Her2 Pri | Her2 2nd event | Grade Pri | Grade 2nd event | Quadrant 2nd event | Margins | Screening | Clonality P value Copy N | Clonality P value Panel seq | Clonality P value WES | Final verdict |
|---|---|---|---|---|---|---|---|---|---|---|---|---|---|---|---|---|---|---|
| metachronous | 55 | Ipsilateral | IDC with DCIS | lumpectomy | None | + | + | + | + | 3 | 3 | at or adjacent to primary | NA | symptomatic | 0.00292113 | NA | NA | Related |

### Primary event

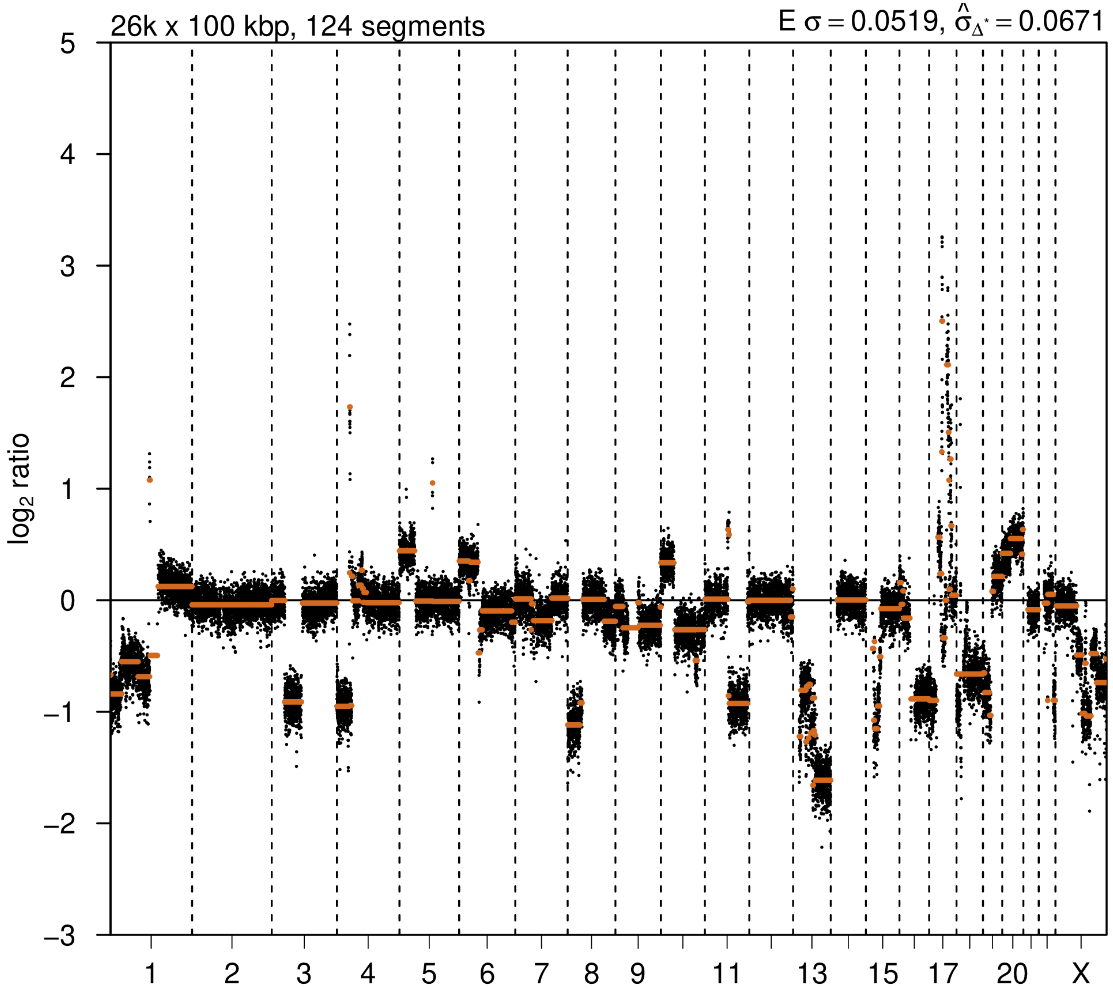


### 2nd event

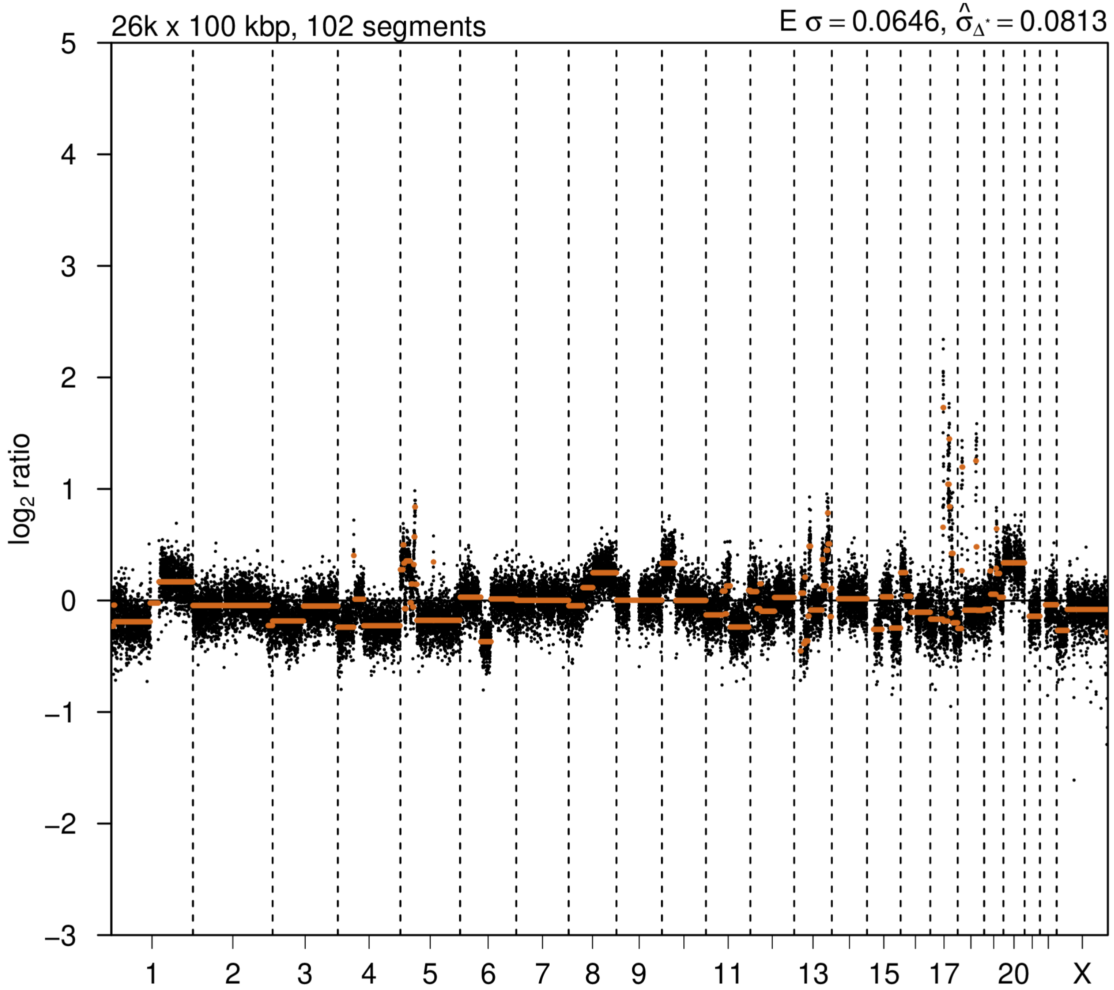

# P073

| Syn/Meta | Time from 1st surgery to 2nd event (Months) | Side 2nd event | Histology 2nd event | Surgery | AdjuvanT Treatment Pri (RT/ HT) | ER Pri | ER 2nd event | Her2 Pri | Her2 2nd event | Grade Pri | Grade 2nd event | Quadrant 2nd event | Margins | Screening | Clonality P value Copy N | Clonality P value Panel seq | Clonality P value WES | Final verdict |
|---|---|---|---|---|---|---|---|---|---|---|---|---|---|---|---|---|---|---|
| metachronous | 36 | Ipsilateral | IDC with DCIS | lumpectomy | None | - | - | + | + | 3 | 3 | at or adjacent to primary | Clear | screen-detected | 0.000324 57 | NA | NA | Related |

### Primary event

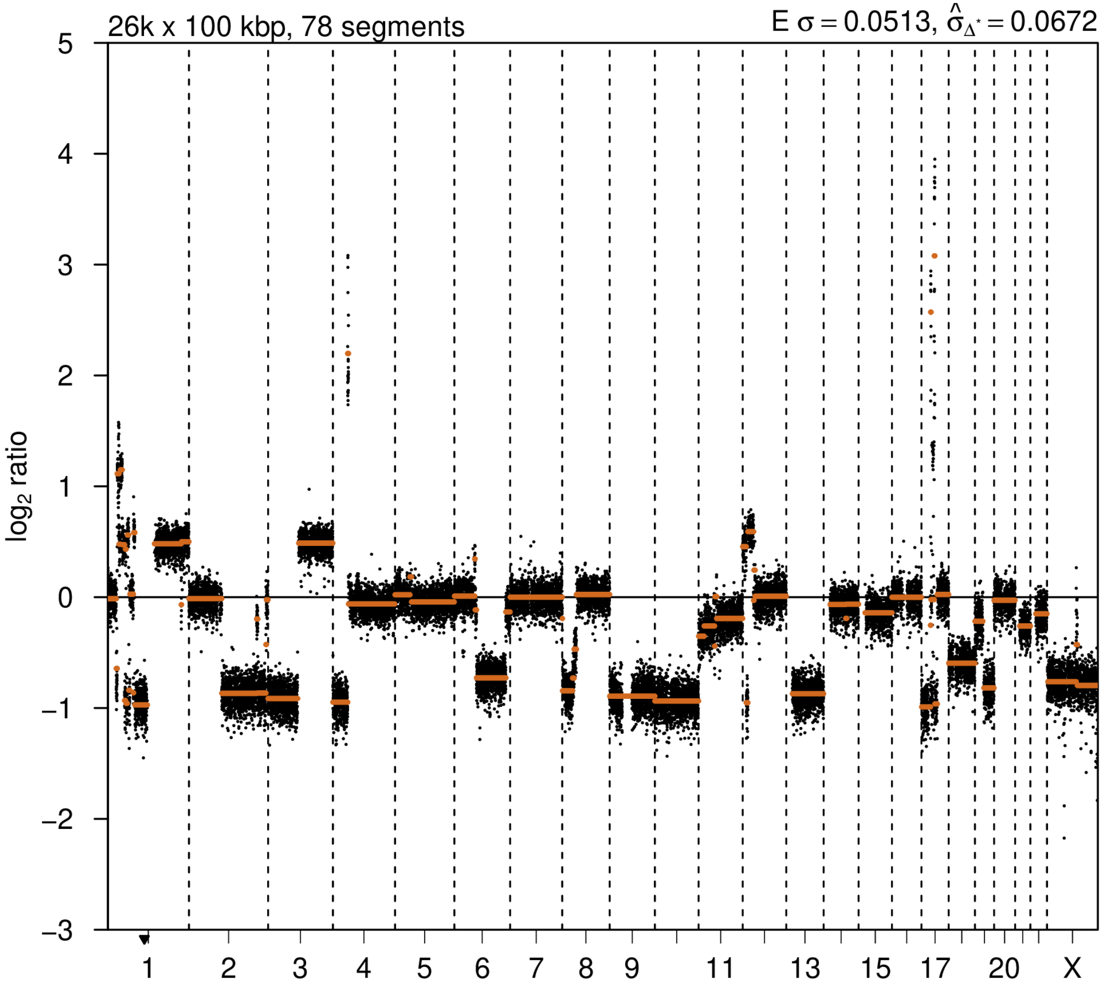


### 2nd event

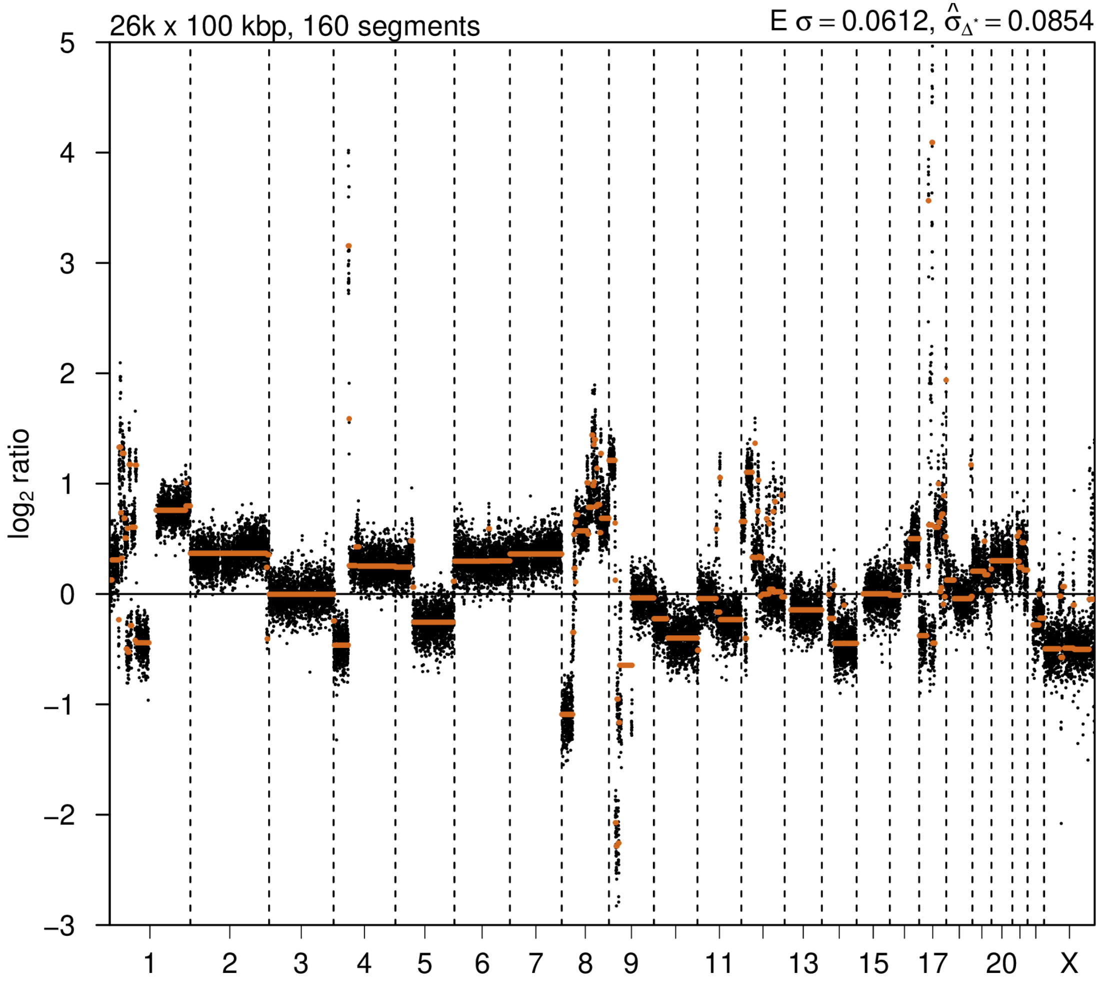

# P082

| Syn/Meta | Time from 1st surgery to 2nd event (Months) | Side 2nd event | Histology 2nd event | Surgery | AdjuvanT Treatment Pri (RT/ HT) | ER Pri | ER 2nd event | Her2 Pri | Her2 2nd event | Grade Pri | Grade 2nd event | Quadrant 2nd event | Margins | Screening | Clonality P value Copy N | Clonality P value Panel seq | Clonality P value WES | Final verdict |
|---|---|---|---|---|---|---|---|---|---|---|---|---|---|---|---|---|---|---|
| metachronous | 11 | Ipsilateral | IDC with DCIS | lumpectomy | None | + | + | + | - | 2 | 2 | distant from primary | involved | symptomatic | 0.00032457 | Single mutation - shared | NA | Related |

### Primary event

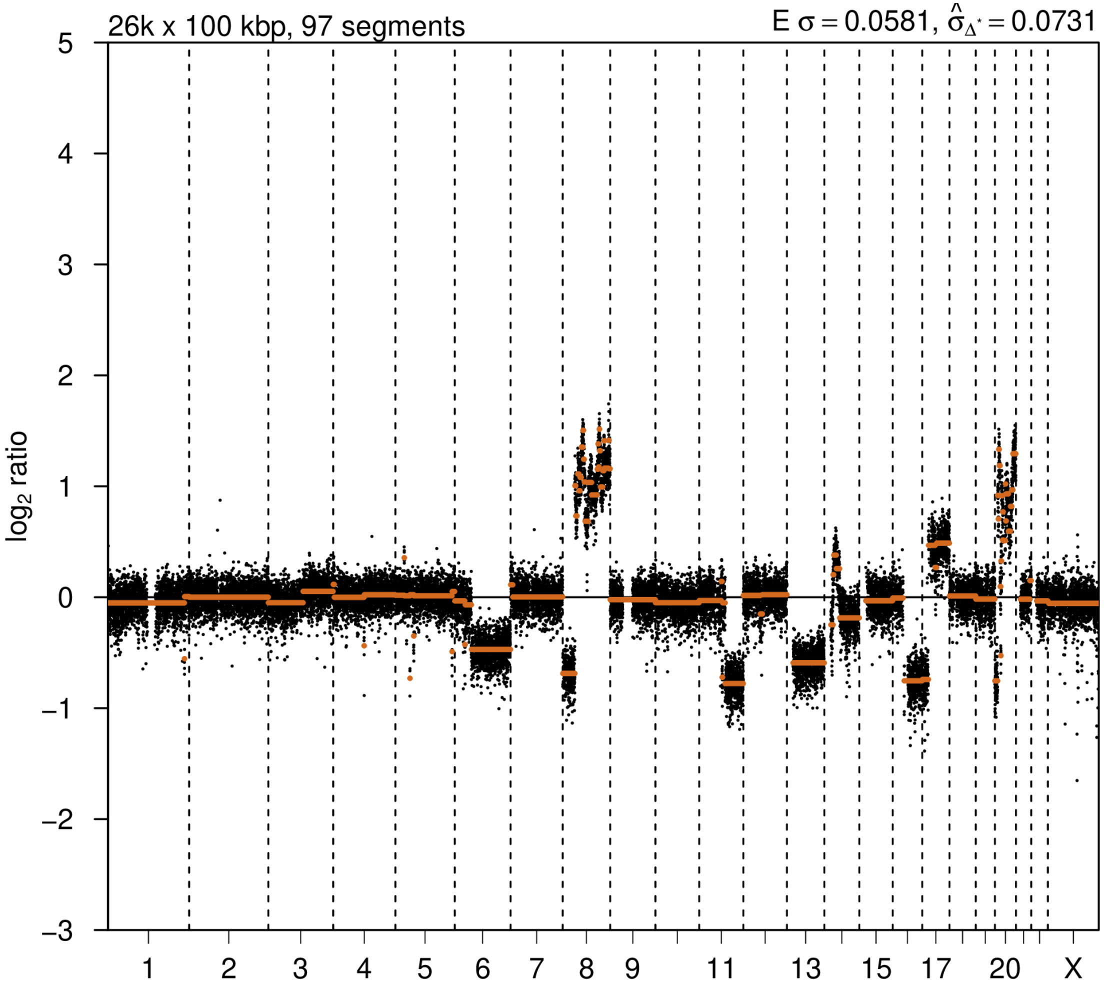


### 2nd event

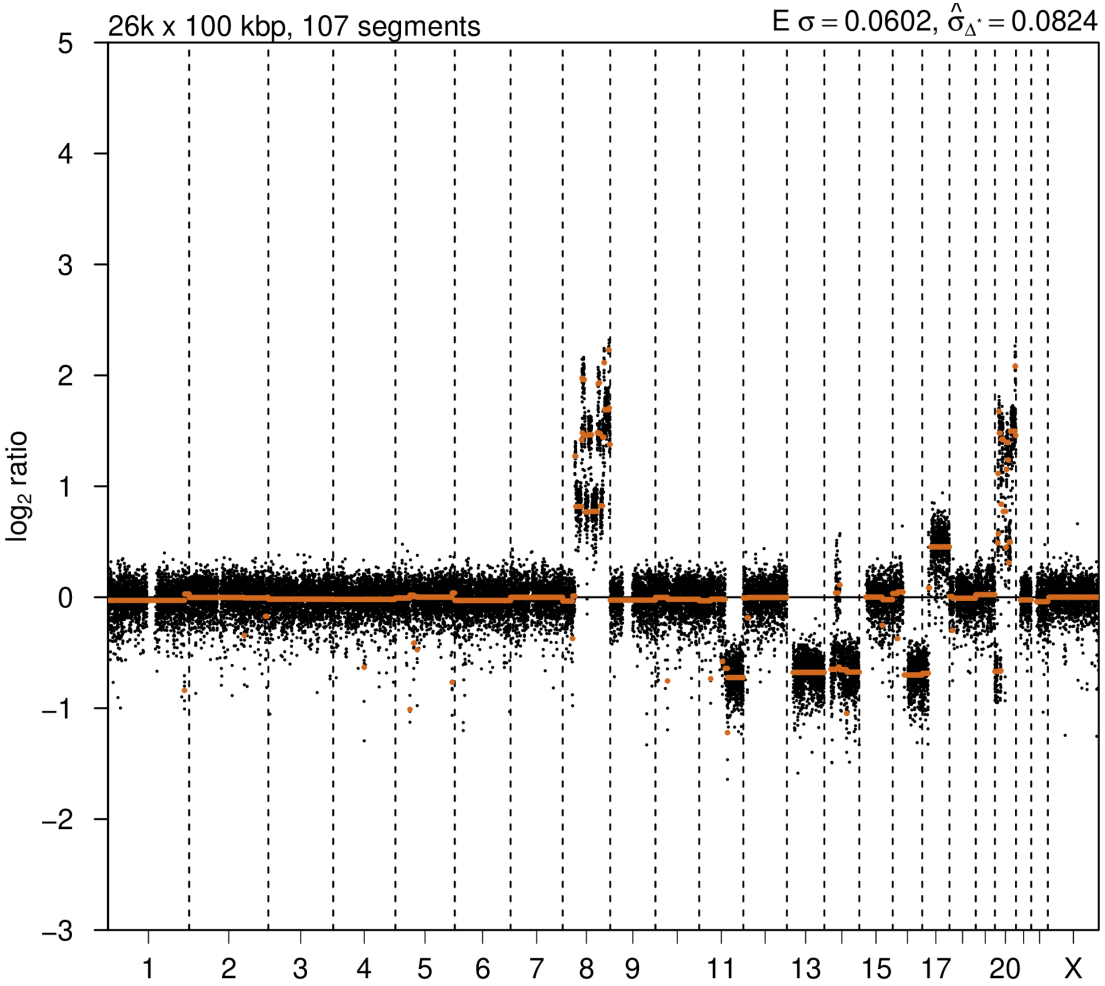

# P083

| Syn/Meta | Time from 1st surgery to 2nd event (Months) | Side 2nd event | Histology 2nd event | Surgery | AdjuvanT Treatment Pri (RT/ HT) | ER Pri | ER 2nd event | Her2 Pri | Her2 2nd event | Grade Pri | Grade 2nd event | Quadrant 2nd event | Margins | Screening | Clonality P value Copy N | Clonality P value Panel seq | Clonality P value WES | Final verdict |
|---|---|---|---|---|---|---|---|---|---|---|---|---|---|---|---|---|---|---|
| metachronous | 13 | Ipsilateral | IDC with DCIS | lumpectomy | None | - | - | + | + | 3 | 3 | distant from primary | Clear | symptomatic | 0.00064914 | 0.003 | NA | Related |

### Primary event

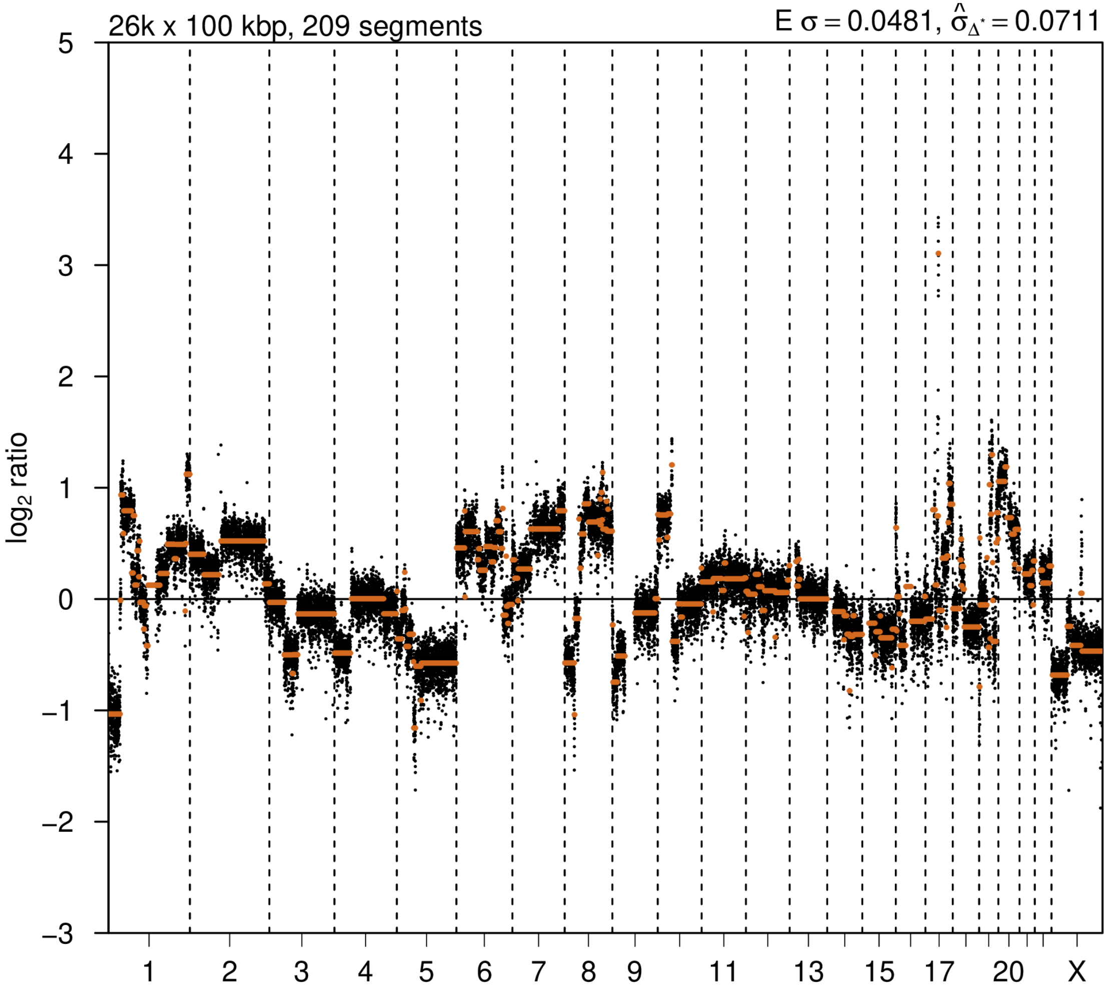


### 2nd event

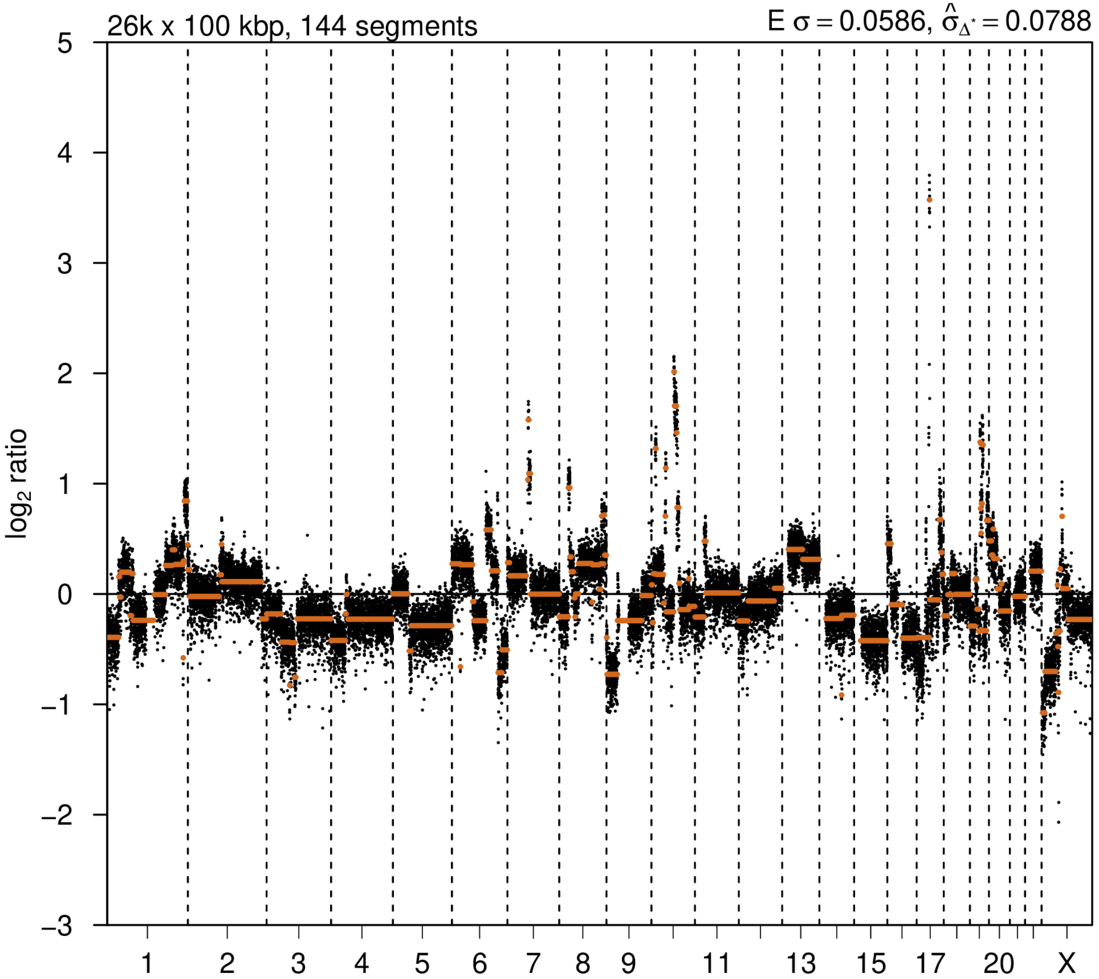

# P084

| Syn/Meta | Time from 1st surgery to 2nd event (Months) | Side 2nd event | Histology 2nd event | Surgery | AdjuvanT Treatment Pri (RT/ HT) | ER Pri | ER 2nd event | Her2 Pri | Her2 2nd event | Grade Pri | Grade 2nd event | Quadrant 2nd event | Margins | Screening | Clonality P value Copy N | Clonality P value Panel seq | Clonality P value WES | Final verdict |
|---|---|---|---|---|---|---|---|---|---|---|---|---|---|---|---|---|---|---|
| metachronous | 13 | Ipsilateral | IDC no DCIS | lumpectomy | None | NA | - | NA | - | 3 | 3 | distant from primary | Clear | NA | 0.007789 679 | NA | NA | Related |

### Primary event

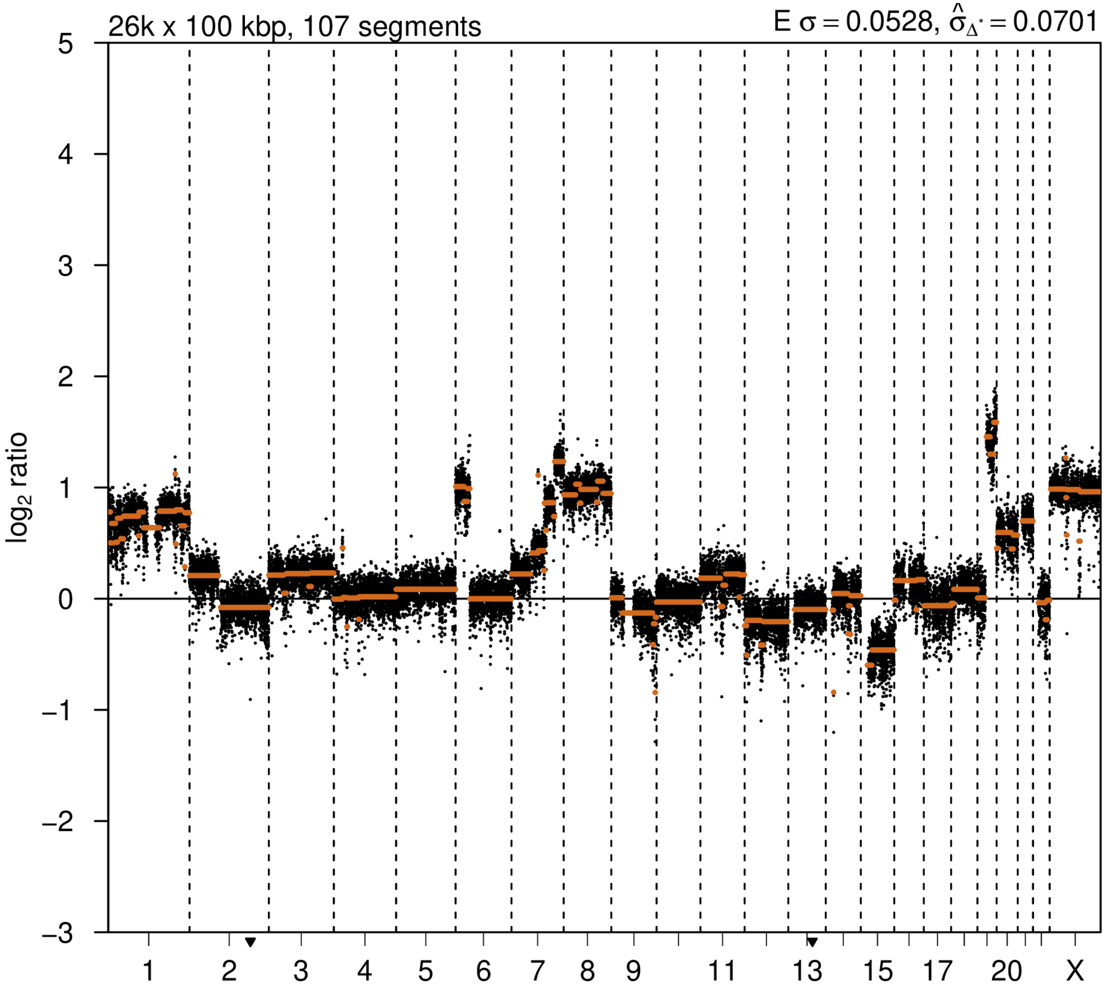


### 2nd event

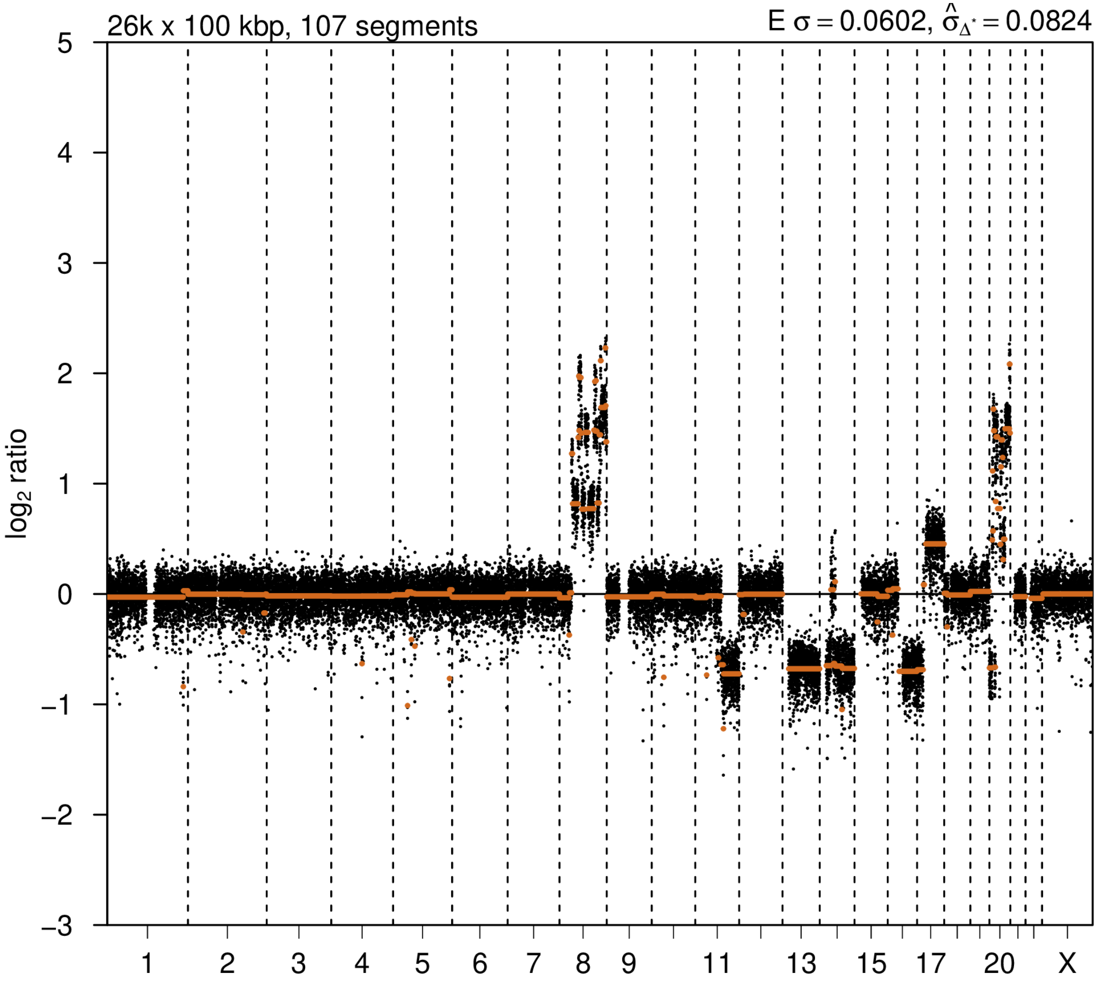

# P085

| Syn/Meta | Time from 1st surgery to 2nd event (Months) | Side 2nd event | Histology 2nd event | Surgery | AdjuvanT Treatment Pri (RT/ HT) | ER Pri | ER 2nd event | Her2 Pri | Her2 2nd event | Grade Pri | Grade 2nd event | Quadrant 2nd event | Margins | Screening | Clonality P value Copy N | Clonality P value Panel seq | Clonality P value WES | Final verdict |
|---|---|---|---|---|---|---|---|---|---|---|---|---|---|---|---|---|---|---|
| metachronous | 26 | Ipsilateral | IDC with DCIS | lumpectomy | None | - | - | + | + | 2 | 2 | NA | Clear | screen-detected | 0.000324 57 | 0.013 | NA | Related |

**Primary event**

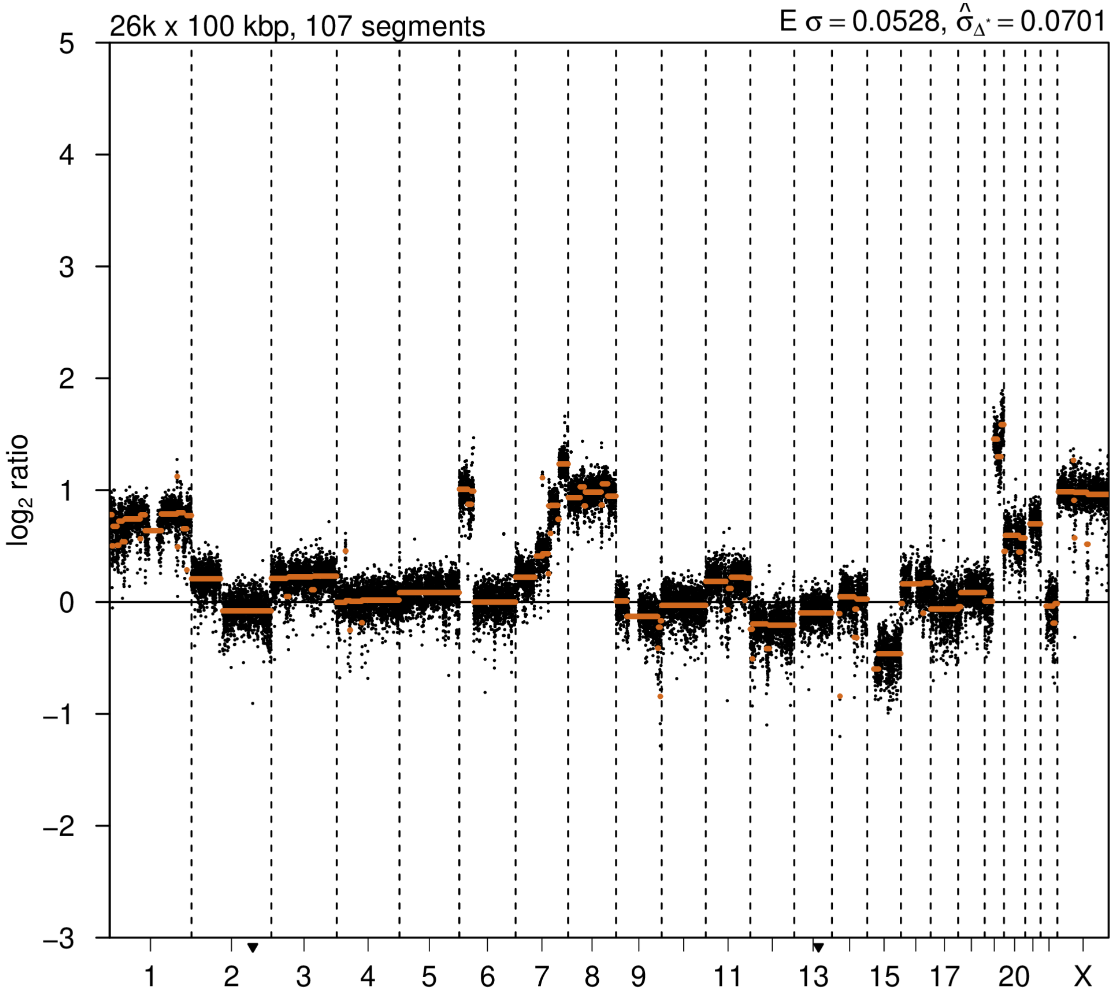


**2nd event**

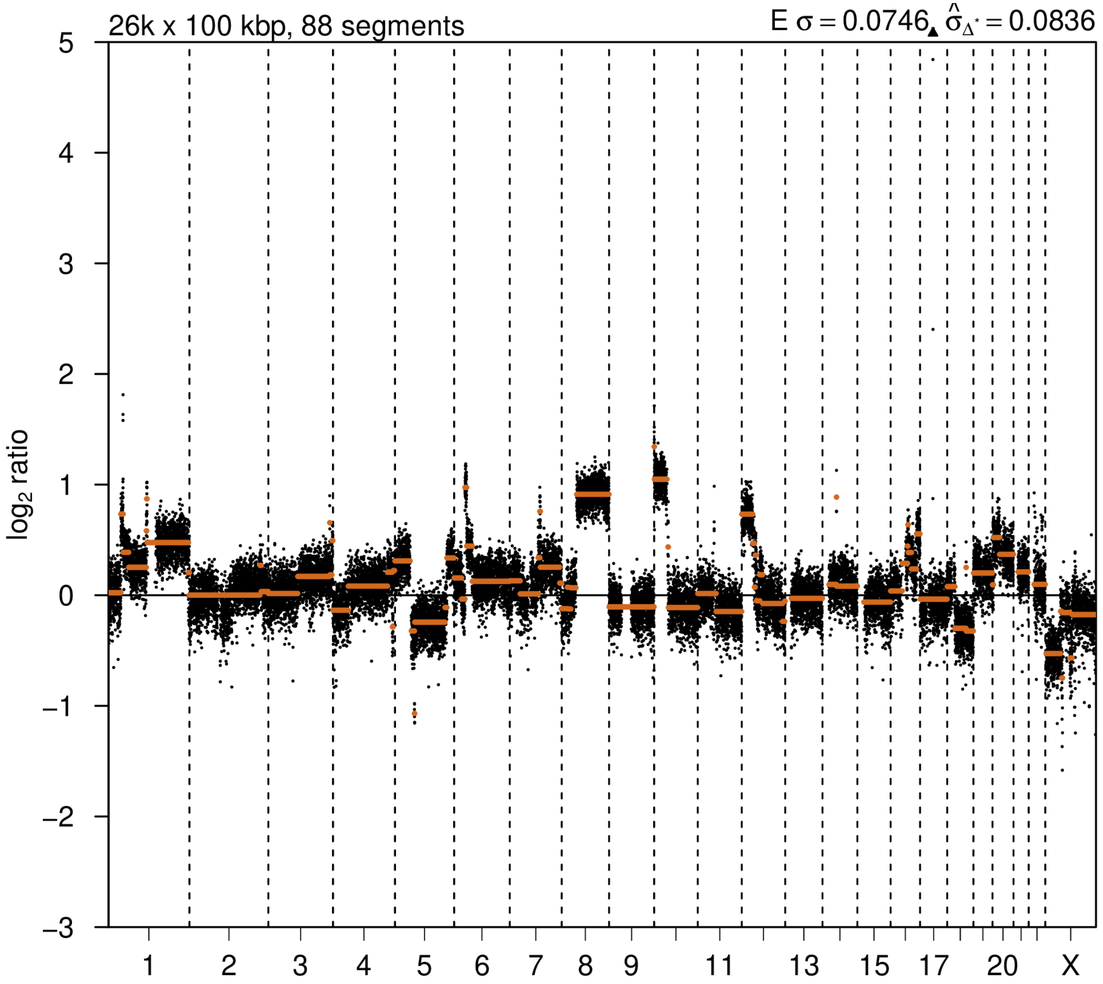

# P086

| Syn/Meta | Time from 1st surgery to 2nd event (Months) | Side 2nd event | Histology 2nd event | Surgery | AdjuvanT Treatment Pri (RT/ HT) | ER Pri | ER 2nd event | Her2 Pri | Her2 2nd event | Grade Pri | Grade 2nd event | Quadrant 2nd event | Margins | Screening | Clonality P value Copy N | Clonality P value Panel seq | Clonality P value WES | Final verdict |
|---|---|---|---|---|---|---|---|---|---|---|---|---|---|---|---|---|---|---|
| metachronous | 27 | Ipsilateral | IDC with DCIS | lumpectomy | None | - | - | + | - | 2 | 2 | NA | NA | screen-detected | 0.107757222 | 0.003 | NA | Related |

### Primary event

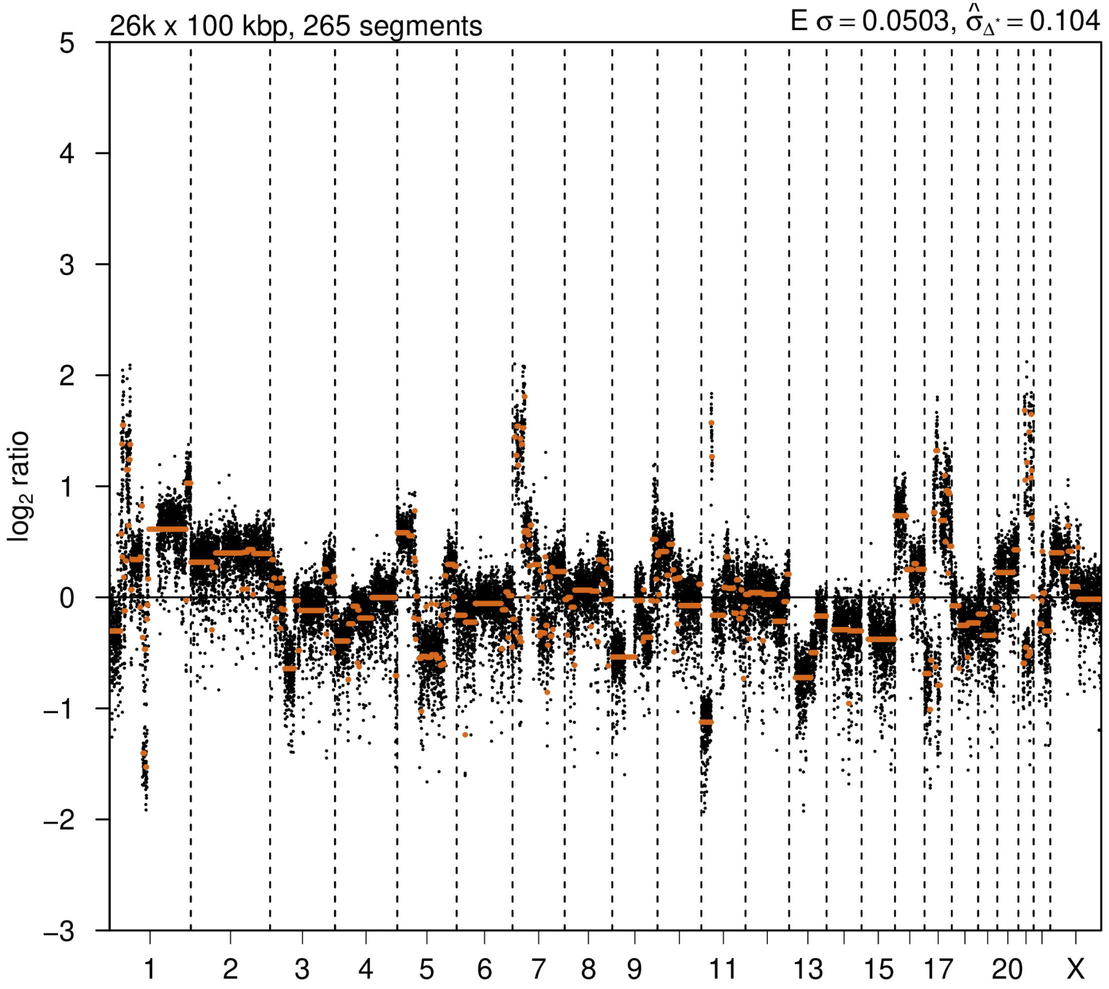


### 2nd event

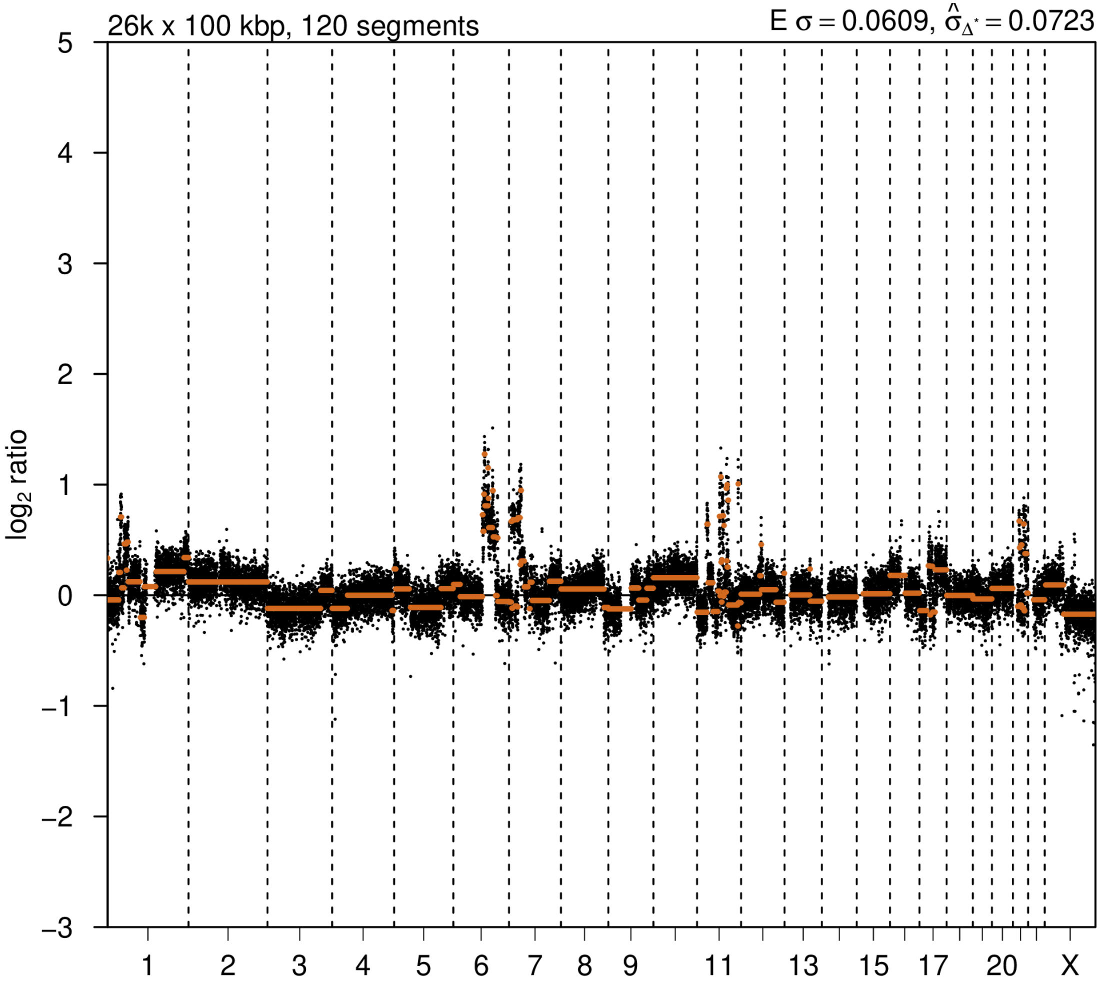

# P088

| Syn/Meta | Time from 1st surgery to 2nd event (Months) | Side 2nd event | Histology 2nd event | Surgery | AdjuvanT Treatment Pri (RT/ HT) | ER Pri | ER 2nd event | Her2 Pri | Her2 2nd event | Grade Pri | Grade 2nd event | Quadrant 2nd event | Margins | Screening | Clonality P value Copy N | Clonality P value Panel seq | Clonality P value WES | Final verdict |
|---|---|---|---|---|---|---|---|---|---|---|---|---|---|---|---|---|---|---|
| metachronous | 28 | Ipsilateral | IDC with DCIS | lumpectomy | None | + | + | - | - | 2 | 3 | NA | Clear | symptomatic | 0.00032457 | Single mutation - shared | NA | Related |

### Primary event

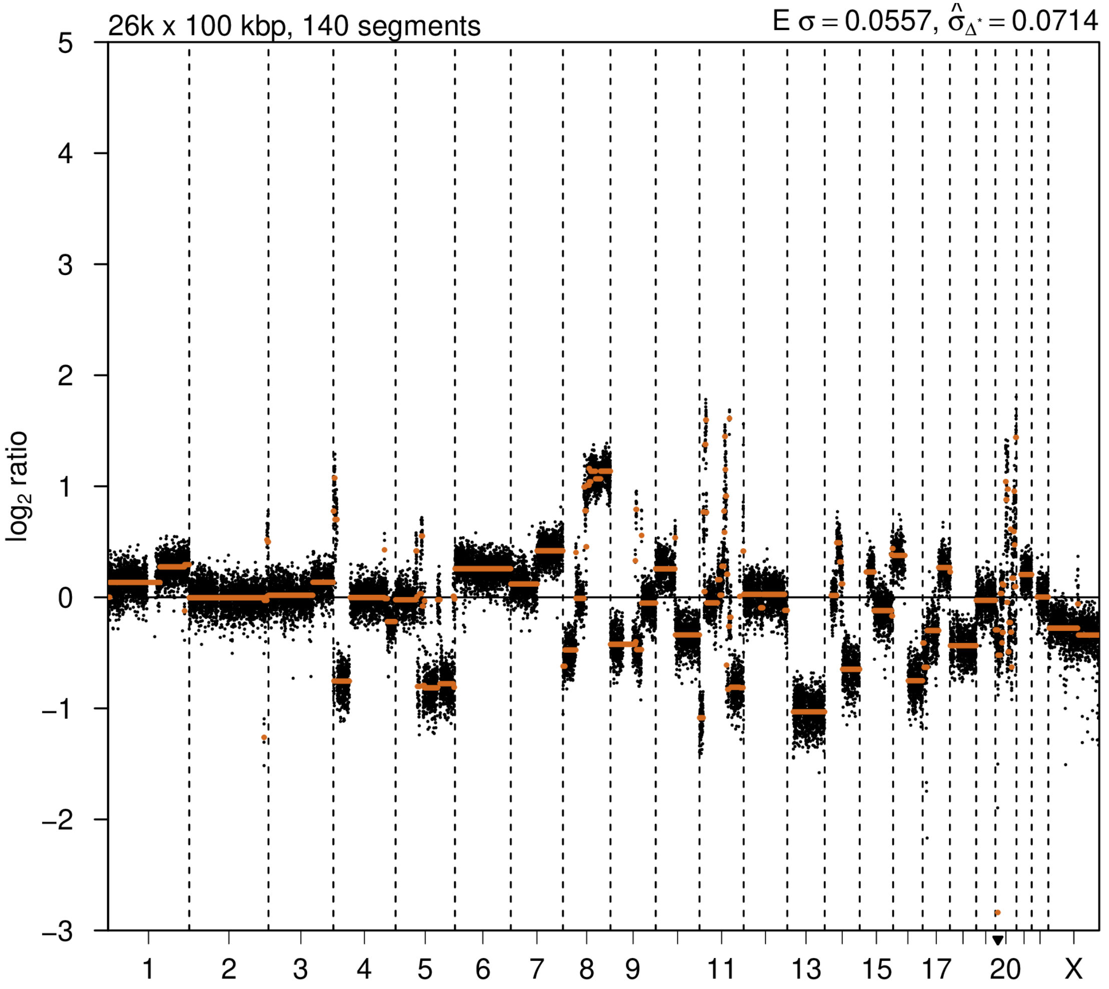


### 2nd event

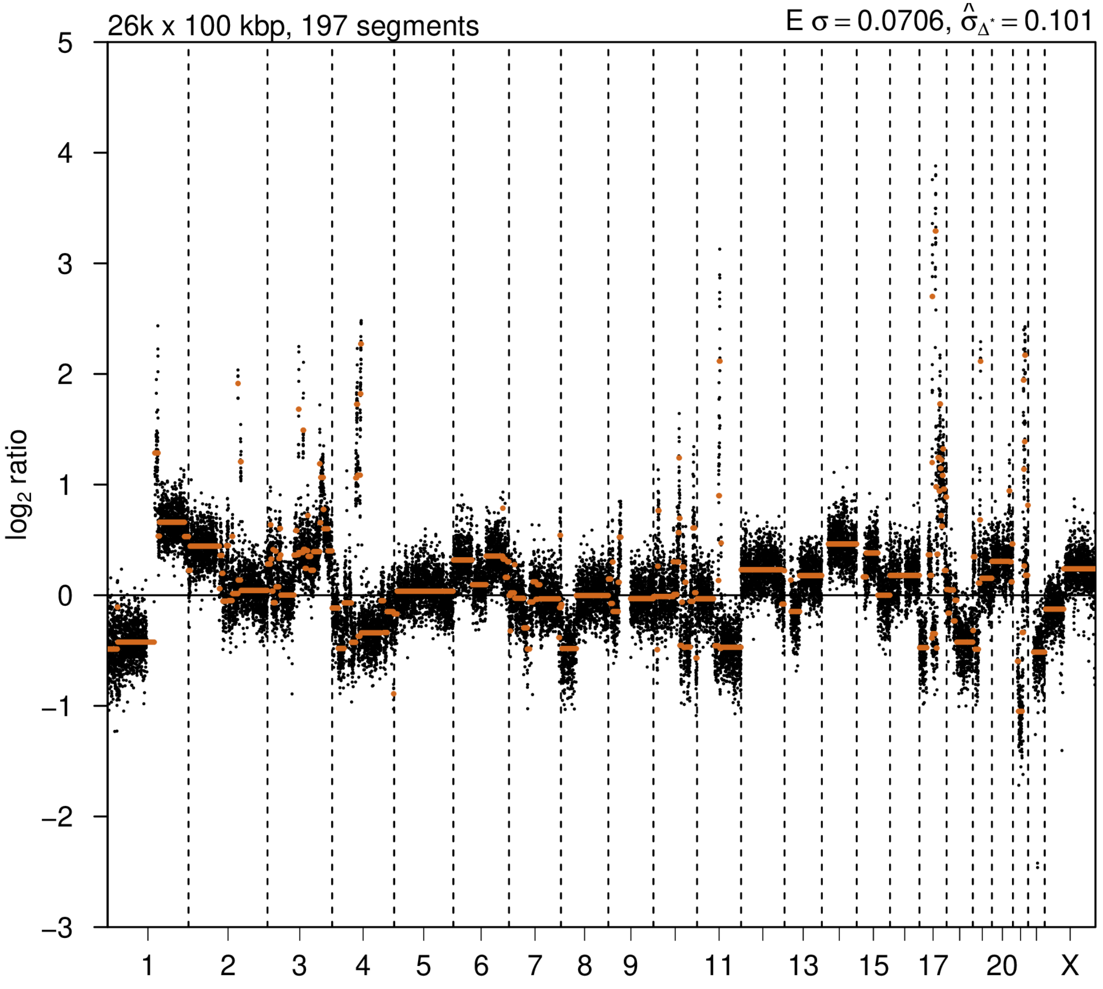

# P094

| Syn/Meta | Time from 1st surgery to 2nd event (Months) | Side 2nd event | Histology 2nd event | Surgery | AdjuvanT Treatment Pri (RT/ HT) | ER Pri | ER 2nd event | Her2 Pri | Her2 2nd event | Grade Pri | Grade 2nd event | Quadrant 2nd event | Margins | Screening | Clonality P value Copy N | Clonality P value Panel seq | Clonality P value WES | Final verdict |
|---|---|---|---|---|---|---|---|---|---|---|---|---|---|---|---|---|---|---|
| metachronous | 29 | Ipsilateral | IDC no DCIS | lumpectomy | None | + | + | - | - | 2 | 2 | at or adjacent to primary | Clear | symptomatic | 0.00032457 | NA | NA | Related |

### Primary event

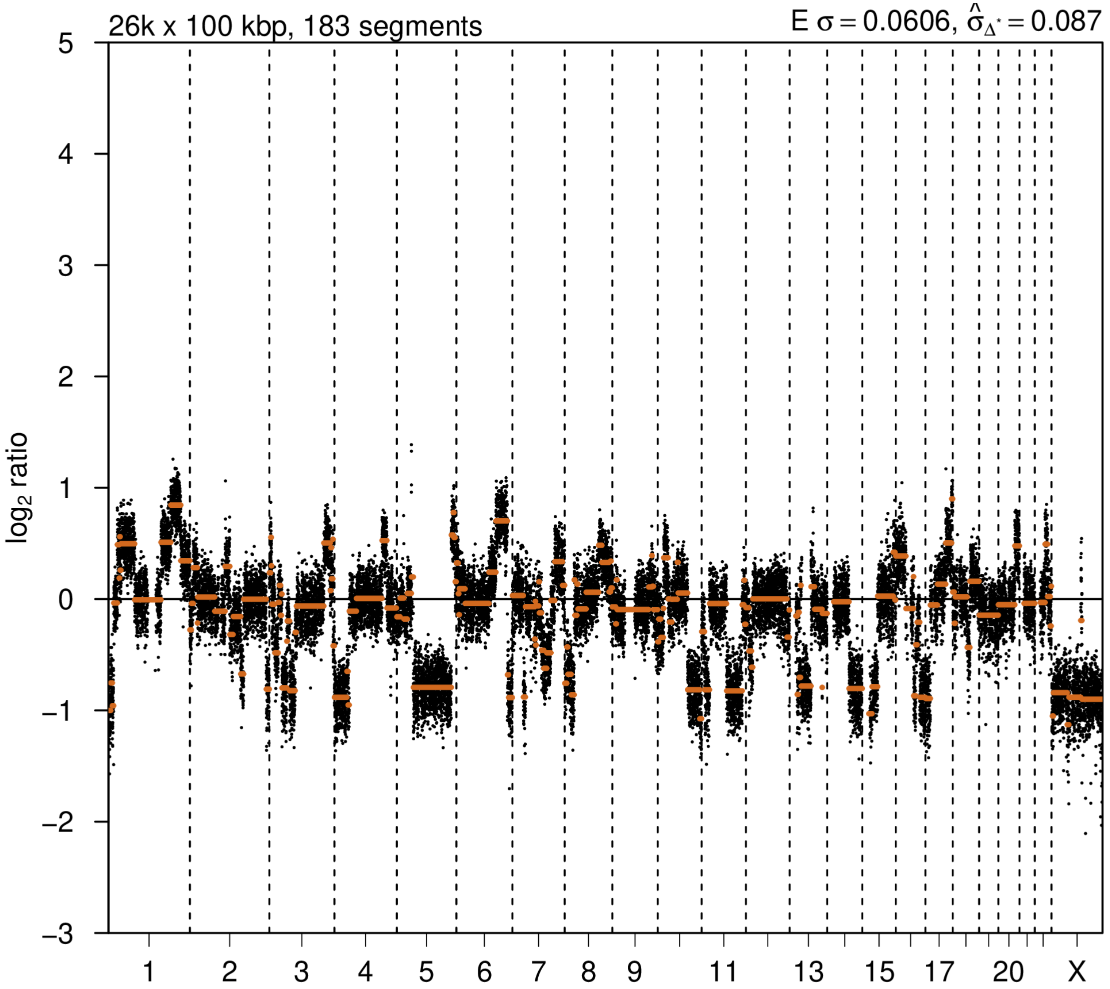


### 2nd event

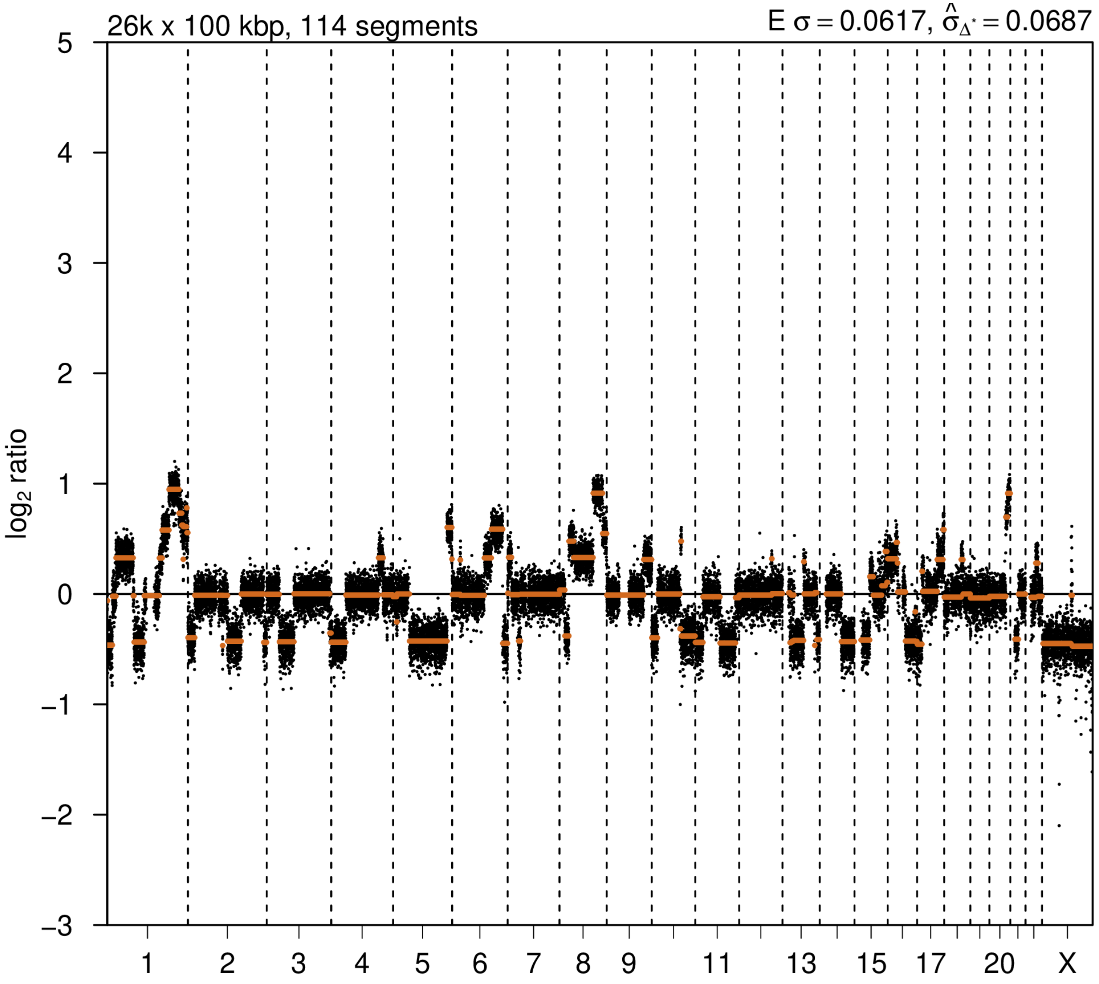

# P095

| Syn/Meta | Time from 1st surgery to 2nd event (Months) | Side 2nd event | Histology 2nd event | Surgery | AdjuvanT Treatment Pri (RT/ HT) | ER Pri | ER 2nd event | Her2 Pri | Her2 2nd event | Grade Pri | Grade 2nd event | Quadrant 2nd event | Margins | Screening | Clonality P value Copy N | Clonality P value Panel seq | Clonality P value WES | Final verdict |
|---|---|---|---|---|---|---|---|---|---|---|---|---|---|---|---|---|---|---|
| metachronous | 25 | Ipsilateral | IDC with DCIS | lumpectomy | None | + | + | - | - | 2 | 1 | at or adjacent to primary | Clear | screen-detected | 0.000324 57 | 0.006 | NA | Related |

### Primary event

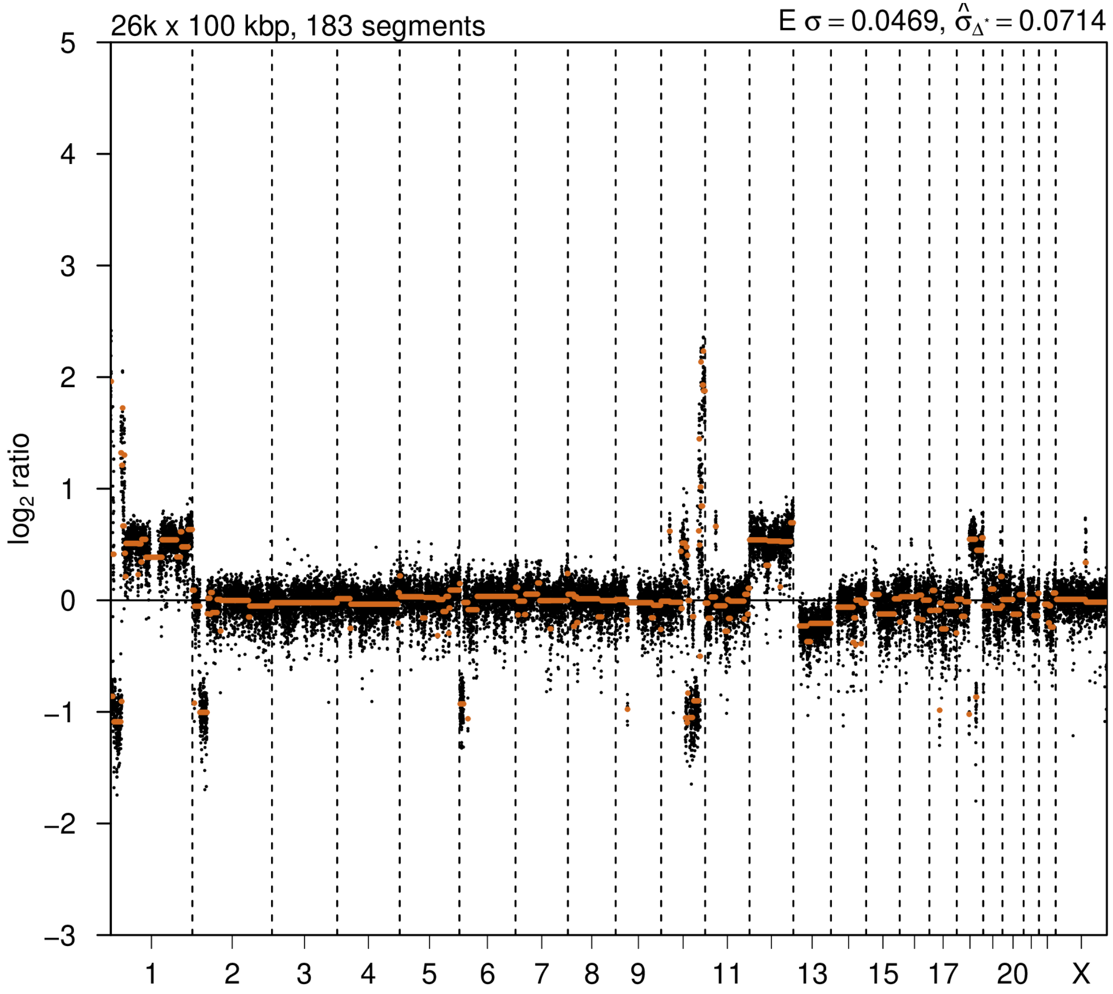


### 2nd event

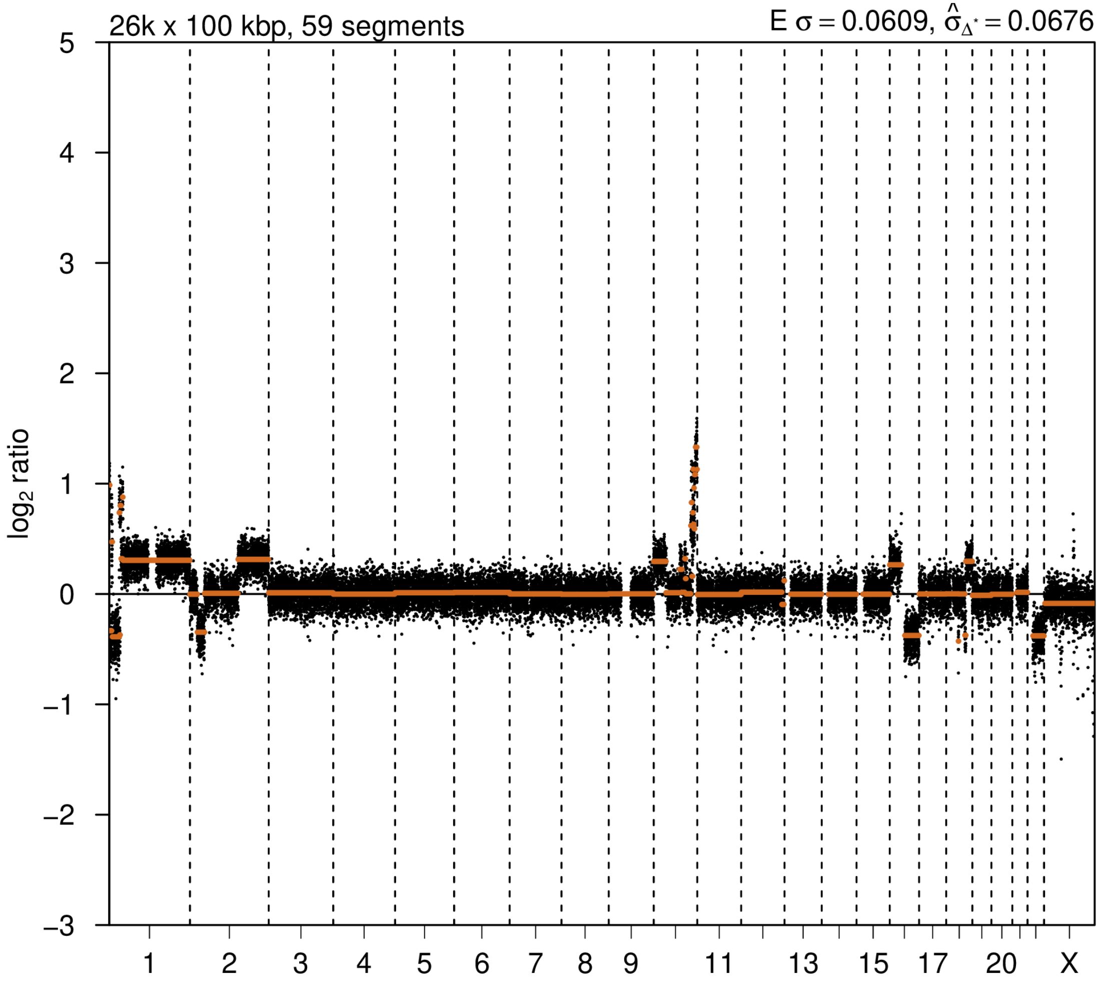

# P096

| Syn/Meta | Time from 1st surgery to 2nd event (Months) | Side 2nd event | Histology 2nd event | Surgery | AdjuvanT Treatment Pri (RT/ HT) | ER Pri | ER 2nd event | Her2 Pri | Her2 2nd event | Grade Pri | Grade 2nd event | Quadrant 2nd event | Margins | Screening | Clonality P value Copy N | Clonality P value Panel seq | Clonality P value WES | Final verdict |
|---|---|---|---|---|---|---|---|---|---|---|---|---|---|---|---|---|---|---|
| metachronous | 23 | Ipsilateral | IDC with DCIS | lumpectomy | None | + | + | - | - | 2 | 3 | at or adjacent to primary | Clear | symptomatic | 0.00032457 | NA | 0.000752445 | Related |

### Primary event

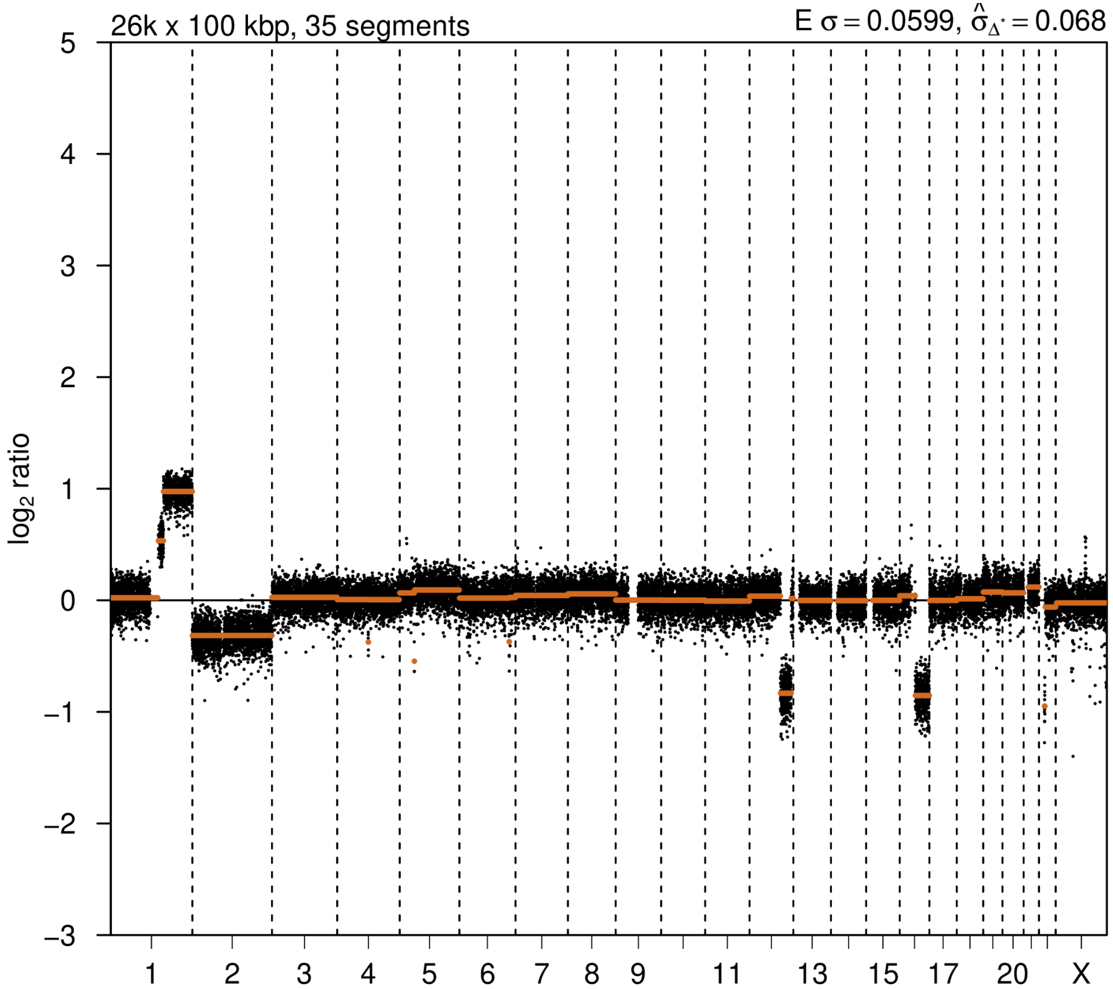


### 2nd event

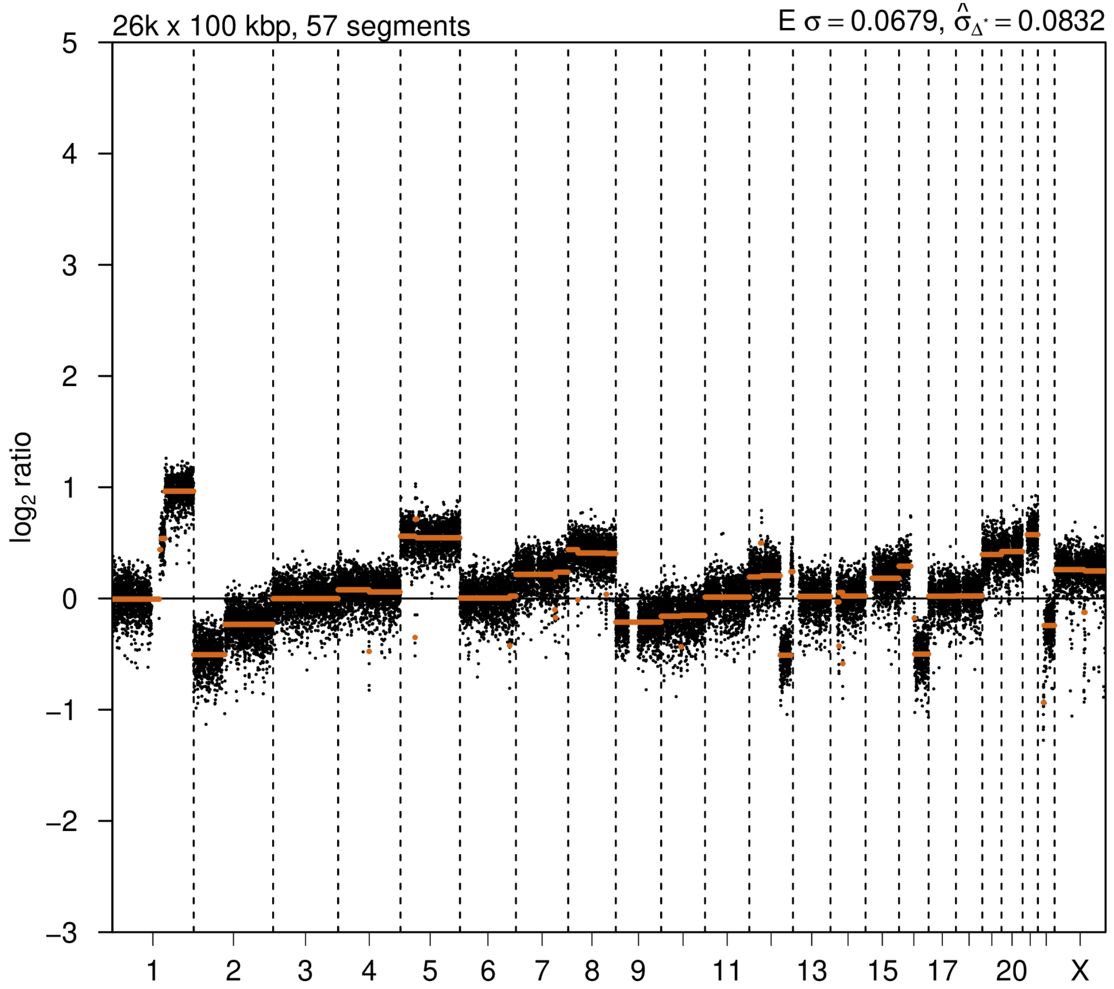

# P102

| Syn/Meta | Time from 1st surgery to 2nd event (Months) | Side 2nd event | Histology 2nd event | Surgery | AdjuvanT Treatment Pri (RT/ HT) | ER Pri | ER 2nd event | Her2 Pri | Her2 2nd event | Grade Pri | Grade 2nd event | Quadrant 2nd event | Margins | Screening | Clonality P value Copy N | Clonality P value Panel seq | Clonality P value WES | Final verdict |
|---|---|---|---|---|---|---|---|---|---|---|---|---|---|---|---|---|---|---|
| metachronous | 184 | Ipsilateral | IDC no DCIS | lumpectomy | None | + | + | - | - | 1 | 2 | NA | NA | symptomatic | 0.003570269 | Single mutation - shared | 1 | Equivocal |

### Primary event

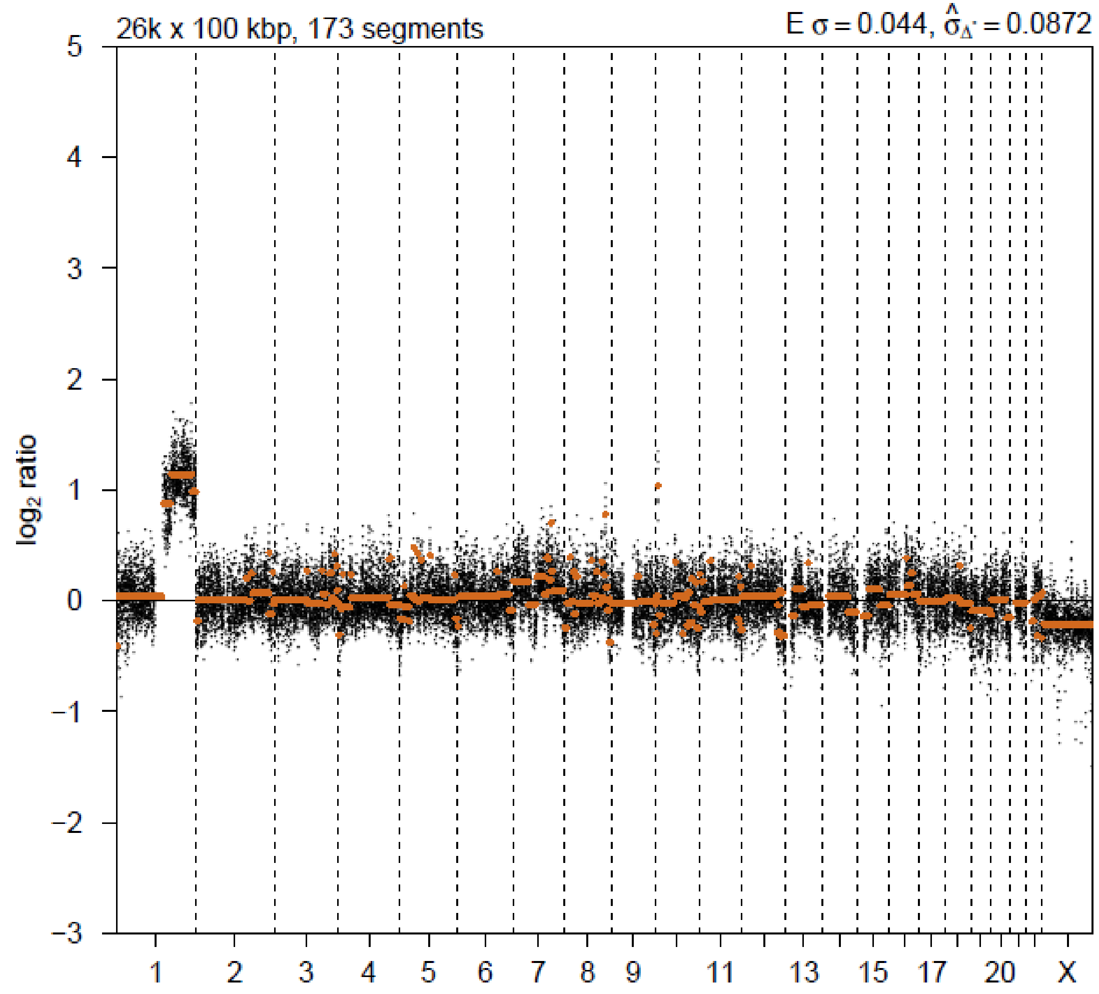


### 2nd event

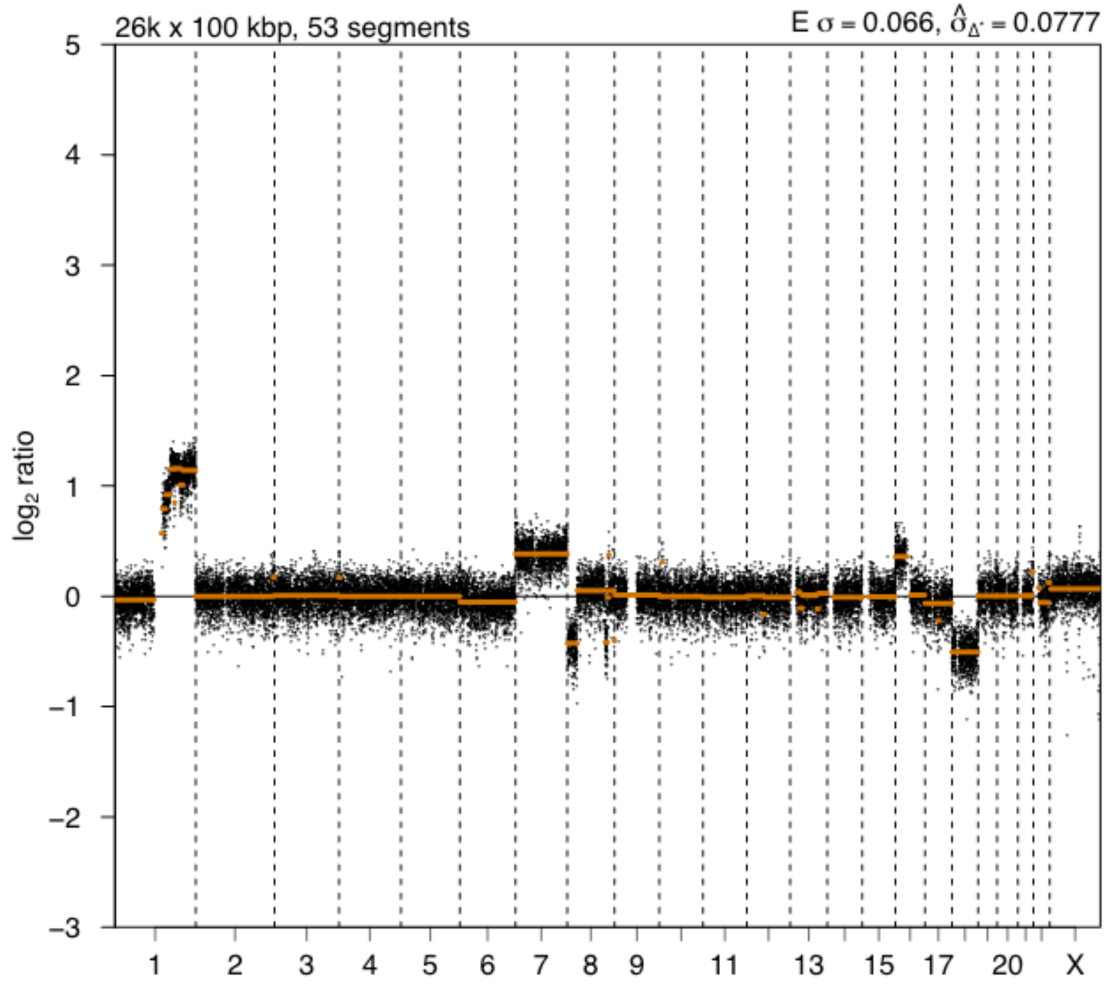

# P103

| Syn/Meta | Time from 1st surgery to 2nd event (Months) | Side 2nd event | Histology 2nd event | Surgery | AdjuvanT Treatment Pri (RT/ HT) | ER Pri | ER 2nd event | Her2 Pri | Her2 2nd event | Grade Pri | Grade 2nd event | Quadrant 2nd event | Margins | Screening | Clonality P value Copy N | Clonality P value Panel seq | Clonality P value WES | Final verdict |
|---|---|---|---|---|---|---|---|---|---|---|---|---|---|---|---|---|---|---|
| metachronous | 119 | Ipsilateral | IDC with DCIS | lumpectomy | None | + | + | - | - | 2 | 3 | distant from primary | Clear | symptomatic | 0.214540734 | 0.025 | NA | Equivocal |

### Primary event

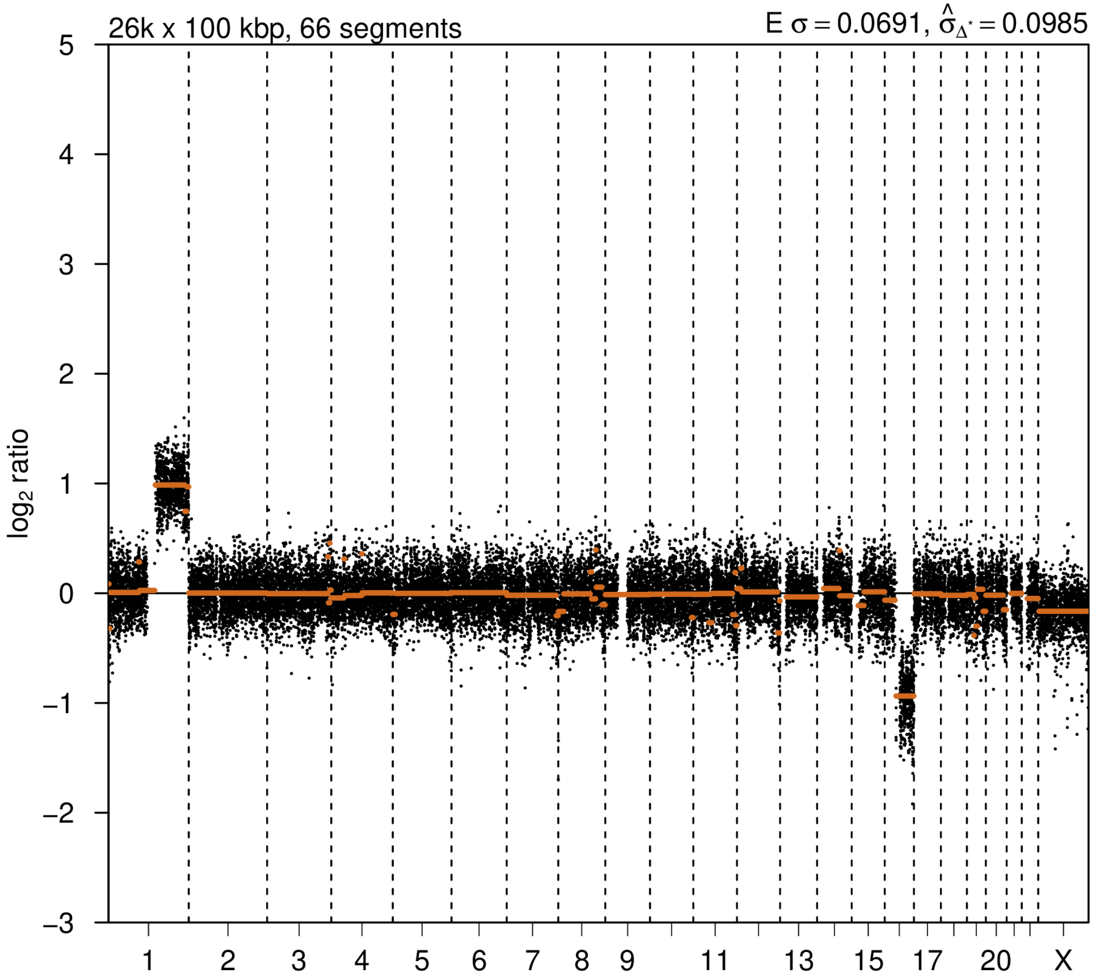


### 2nd event

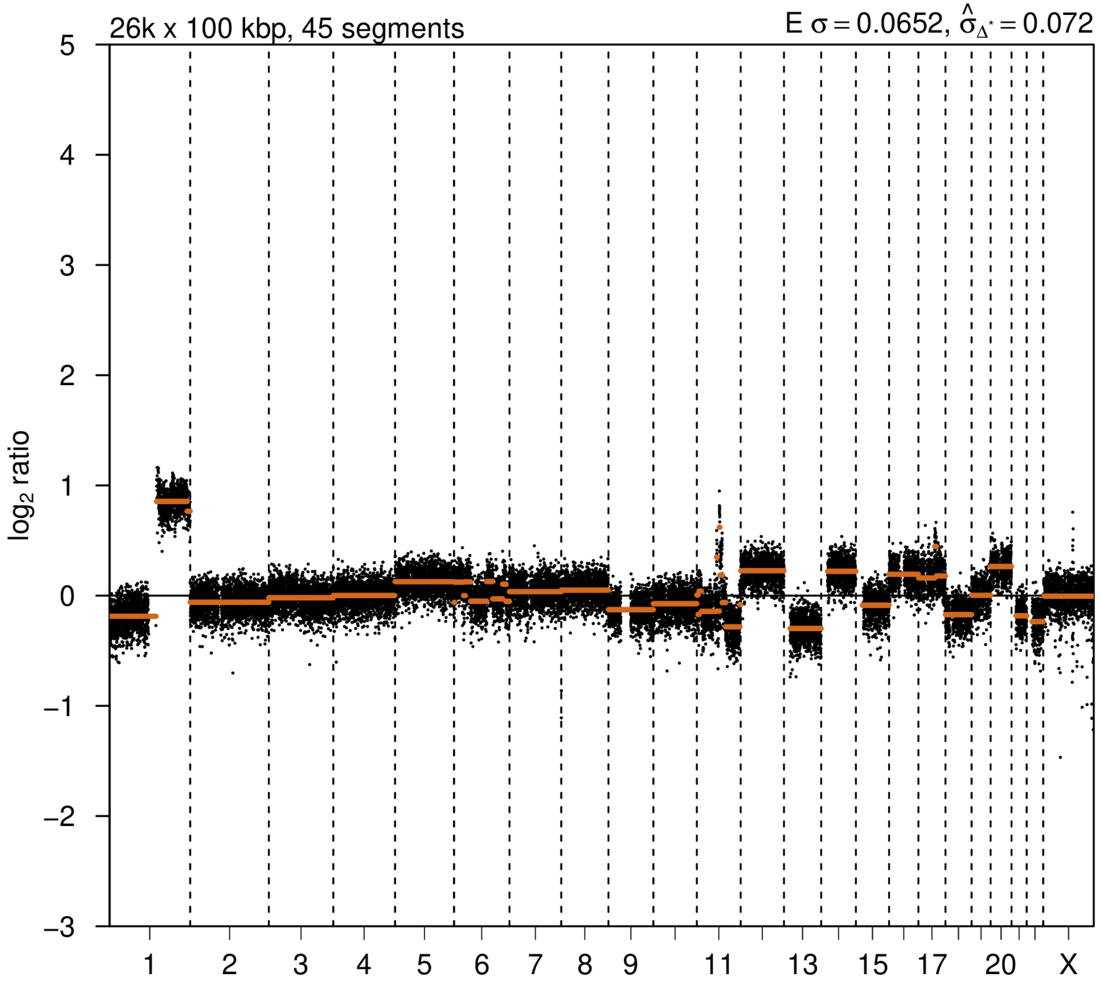

# P104

| Syn/Meta | Time from 1st surgery to 2nd event (Months) | Side 2nd event | Histology 2nd event | Surgery | AdjuvanT Treatment Pri (RT/ HT) | ER Pri | ER 2nd event | Her2 Pri | Her2 2nd event | Grade Pri | Grade 2nd event | Quadrant 2nd event | Margins | Screening | Clonality P value Copy N | Clonality P value Panel seq | Clonality P value WES | Final verdict |
|---|---|---|---|---|---|---|---|---|---|---|---|---|---|---|---|---|---|---|
| metachronous | 104 | Ipsilateral | IDC with DCIS | lumpectomy | None | + | + | - | - | 1 | 1 | at or adjacent to primary | Clear | symptomatic | 0.045439792 | 0.02 | NA | Equivocal |

### Primary event

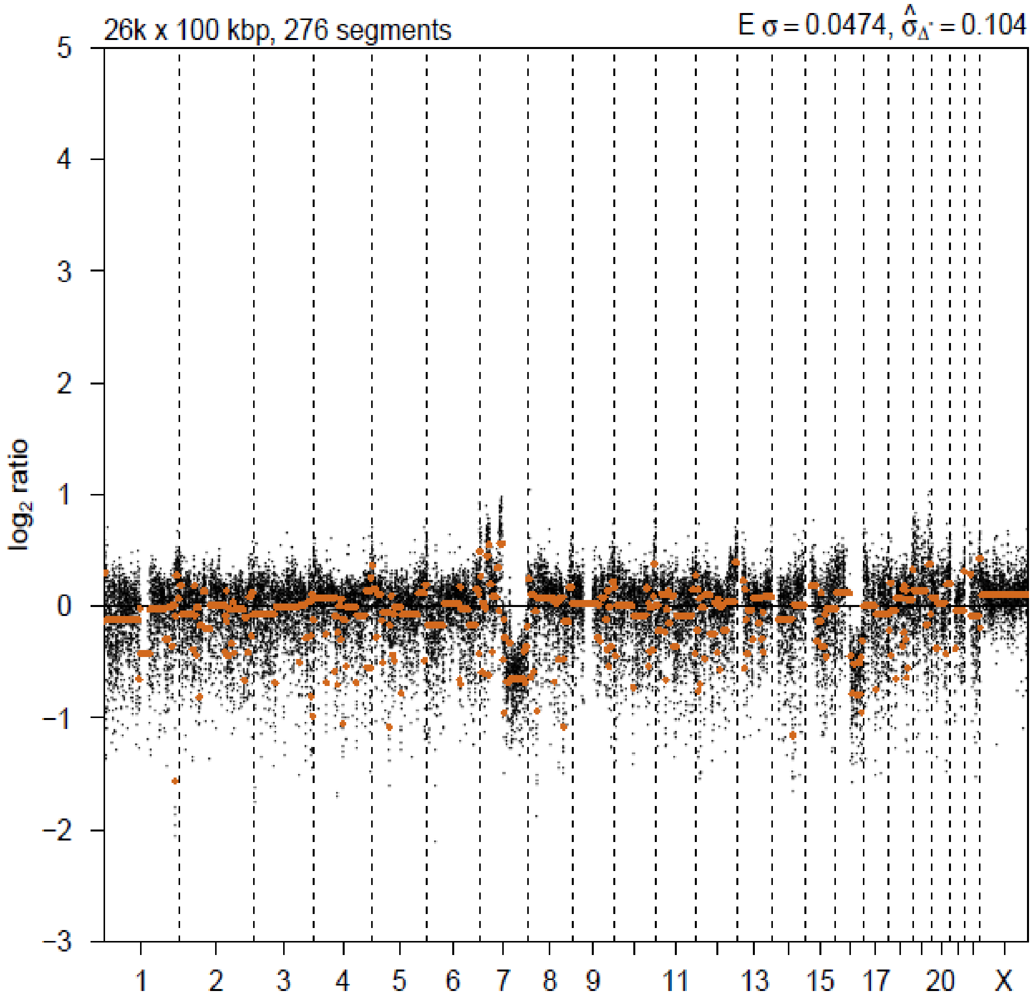


### 2nd event

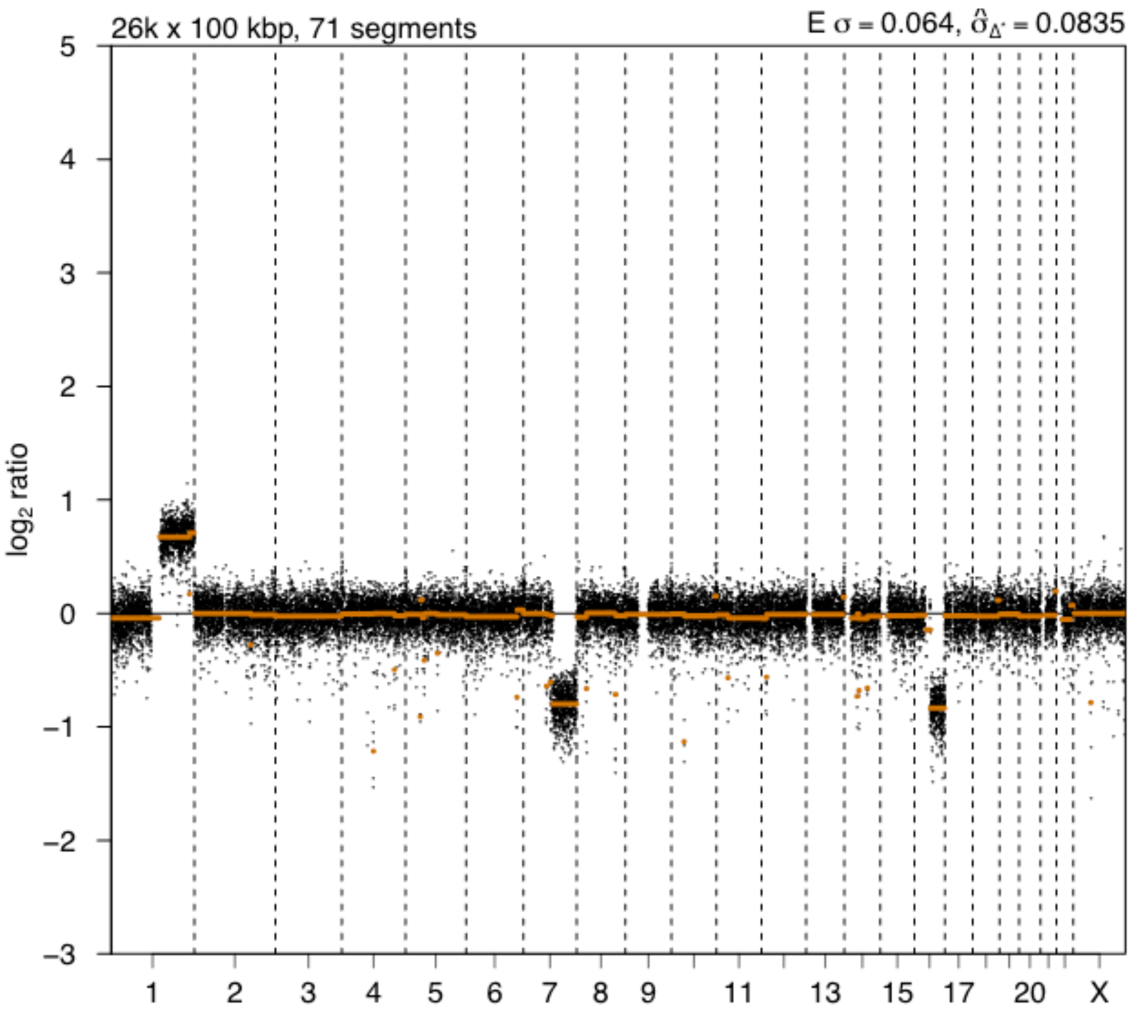

# P105

| Syn/Meta | Time from 1st surgery to 2nd event (Months) | Side 2nd event | Histology 2nd event | Surgery | AdjuvanT Treatment Pri (RT/ HT) | ER Pri | ER 2nd event | Her2 Pri | Her2 2nd event | Grade Pri | Grade 2nd event | Quadrant 2nd event | Margins | Screening | Clonality P value Copy N | Clonality P value Panel seq | Clonality P value WES | Final verdict |
|---|---|---|---|---|---|---|---|---|---|---|---|---|---|---|---|---|---|---|
| metachronous | 90 | Ipsilateral | IDC no DCIS | lumpectomy | None | + | + | - | - | 2 | 3 | at or adjacent to primary | Clear | screen-detected | 0.127880558 | Single mutation - shared | 1 | Equivocal |

### Primary event

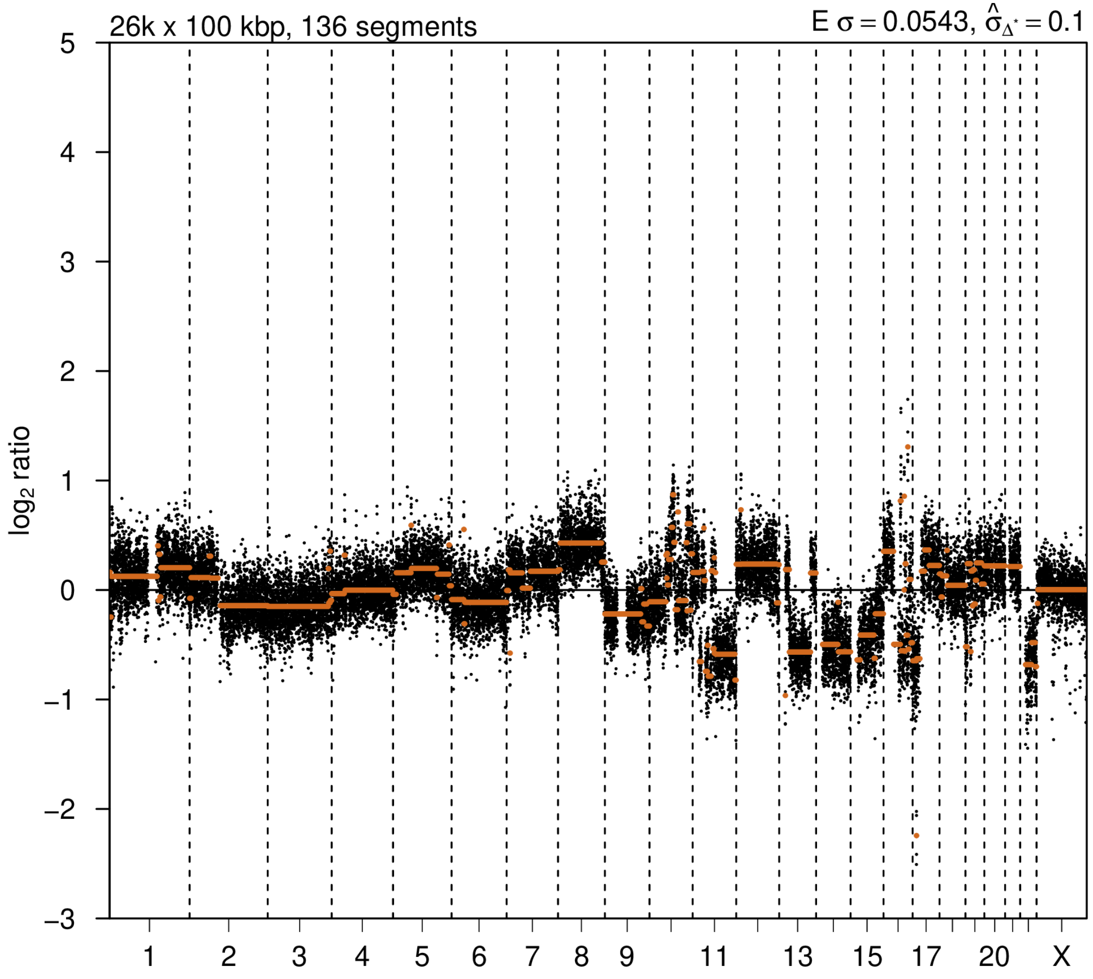


### 2nd event

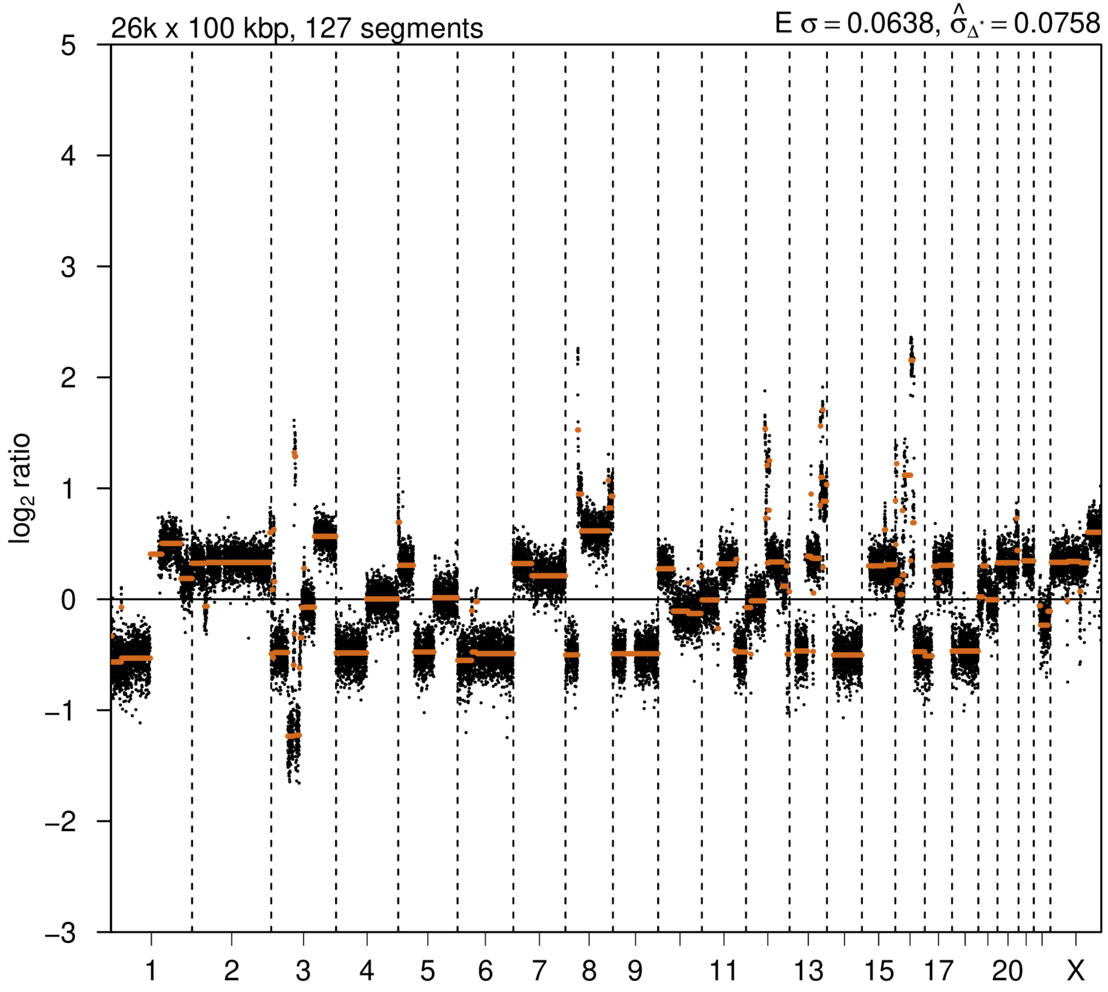

# P106

| Syn/Meta | Time from 1st surgery to 2nd event (Months) | Side 2nd event | Histology 2nd event | Surgery | AdjuvanT Treatment Pri (RT/ HT) | ER Pri | ER 2nd event | Her2 Pri | Her2 2nd event | Grade Pri | Grade 2nd event | Quadrant 2nd event | Margins | Screening | Clonality P value Copy N | Clonality P value Panel seq | Clonality P value WES | Final verdict |
|---|---|---|---|---|---|---|---|---|---|---|---|---|---|---|---|---|---|---|
| metachronous | 80 | Ipsilateral | IDC with DCIS | lumpectomy | None | + | + | - | - | 2 | 1 | at or adjacent to primary | involved | screen-detected | 0.271989614 | 0.013 | NA | Equivocal |

### Primary event

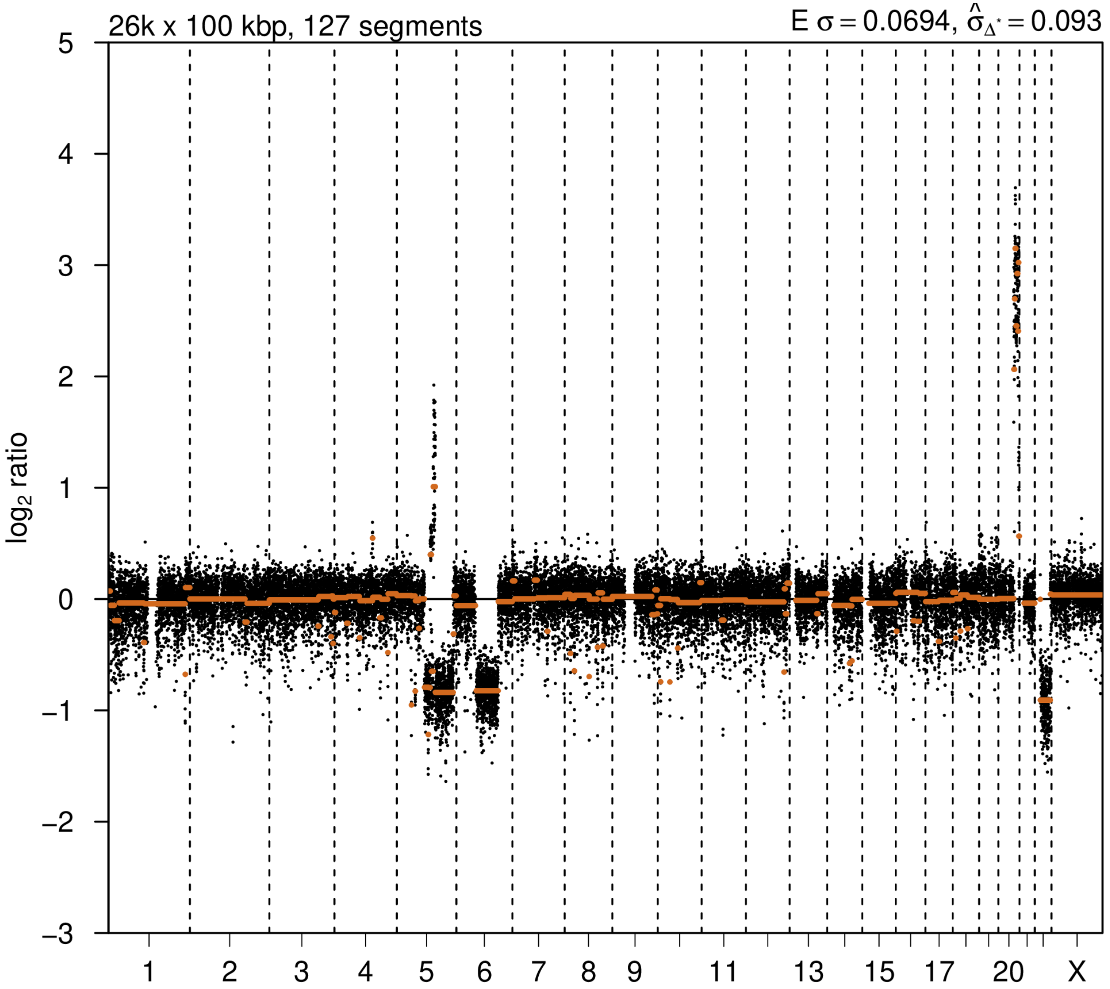


### 2nd event

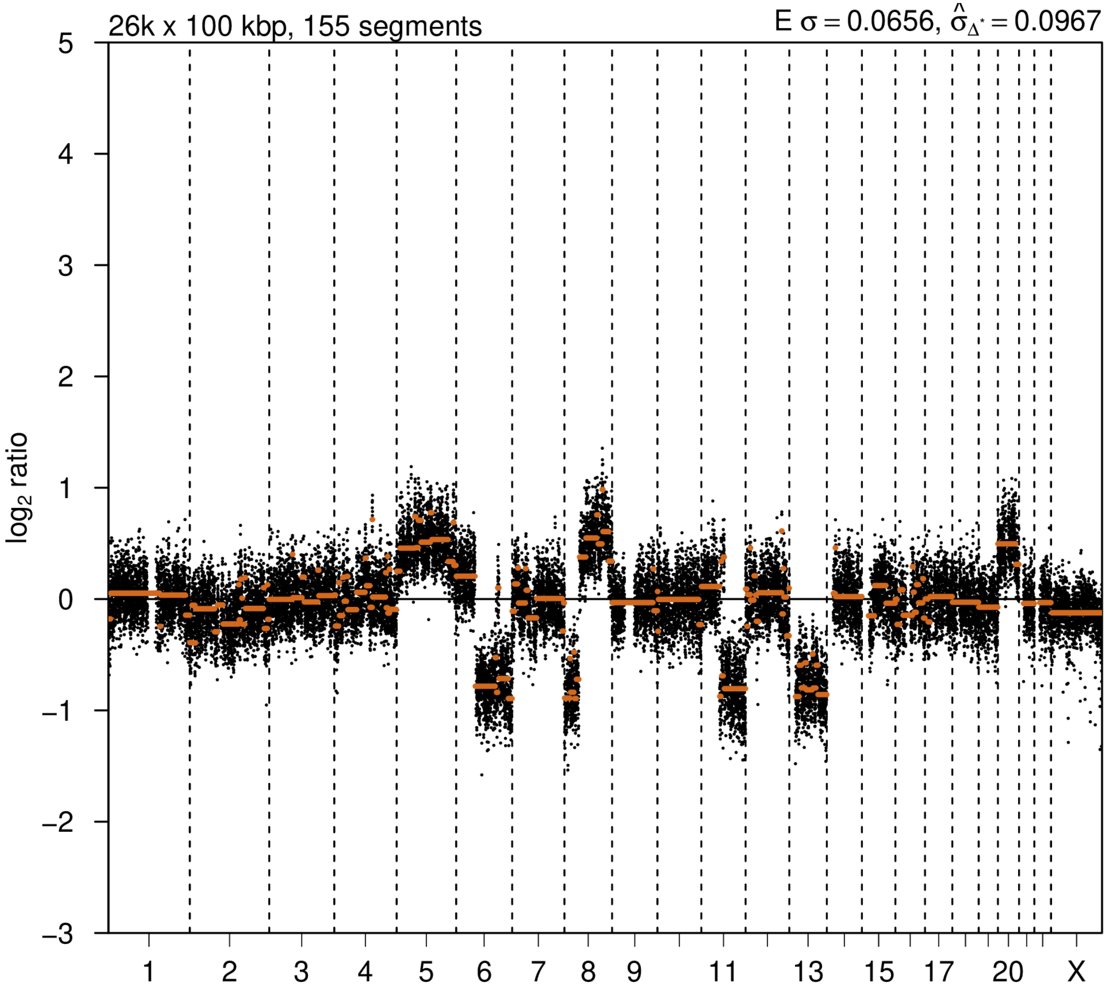

# P113

| Syn/Meta | Time from 1st surgery to 2nd event (Months) | Side 2nd event | Histology 2nd event | Surgery | AdjuvanT Treatment Pri (RT/ HT) | ER Pri | ER 2nd event | Her2 Pri | Her2 2nd event | Grade Pri | Grade 2nd event | Quadrant 2nd event | Margins | Screening | Clonality P value Copy N | Clonality P value Panel seq | Clonality P value WES | Final verdict |
|---|---|---|---|---|---|---|---|---|---|---|---|---|---|---|---|---|---|---|
| metachronous | 173 | Ipsilateral | IDC with DCIS | lumpectomy | None | - | + | + | + | 2 | 2 | distant from primary | Clear | NA | 0.163907822 | NA | NA | Unrelated |

### Primary event

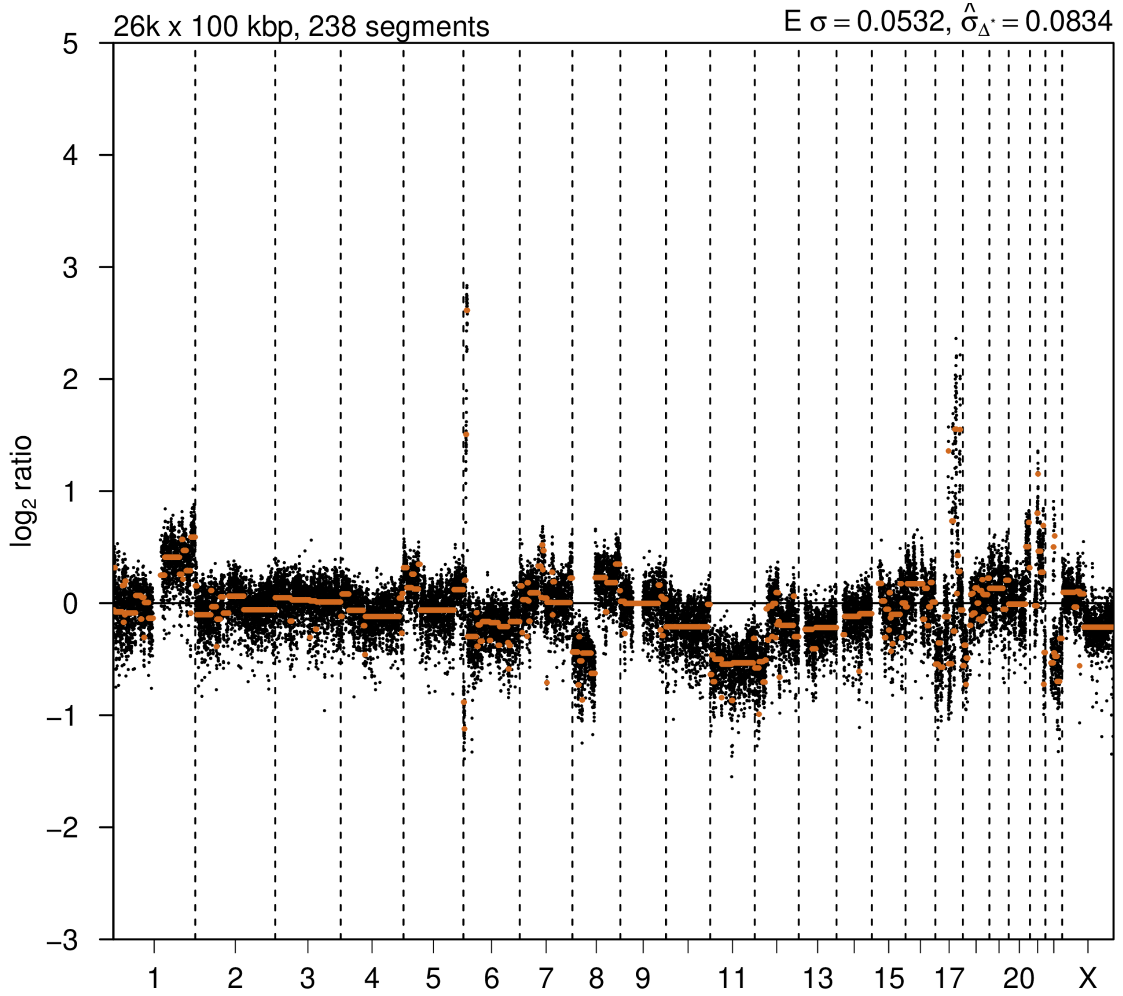


### 2nd event

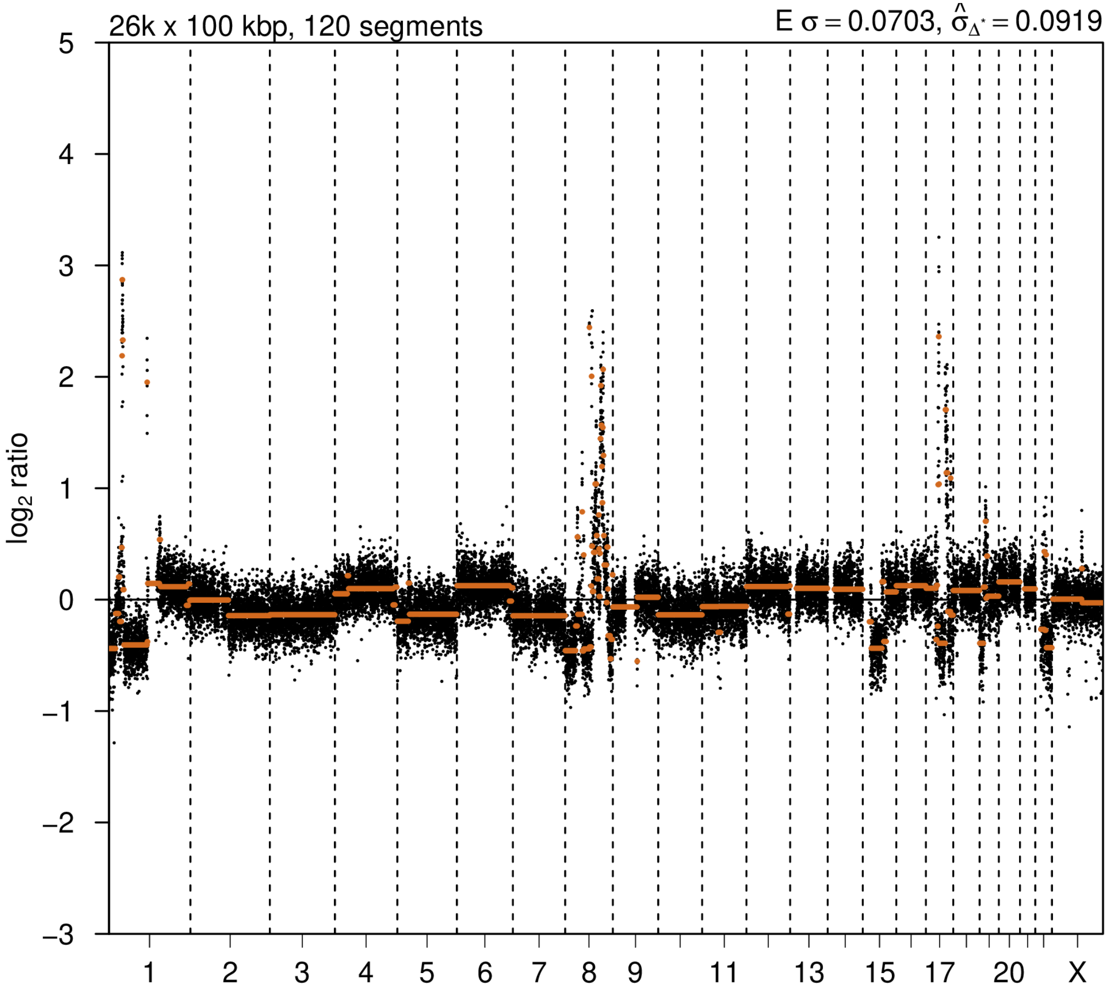

# P114

| Syn/Meta | Time from 1st surgery to 2nd event (Months) | Side 2nd event | Histology 2nd event | Surgery | AdjuvanT Treatment Pri (RT/ HT) | ER Pri | ER 2nd event | Her2 Pri | Her2 2nd event | Grade Pri | Grade 2nd event | Quadrant 2nd event | Margins | Screening | Clonality P value Copy N | Clonality P value Panel seq | Clonality P value WES | Final verdict |
|---|---|---|---|---|---|---|---|---|---|---|---|---|---|---|---|---|---|---|
| metachronous | 165 | Ipsilateral | IDC with DCIS | lumpectomy | None | + | + | - | - | 2 | 3 | distant from primary | Clear | symptomatic | 0.310613437 | NA | NA | Unrelated |

### Primary event

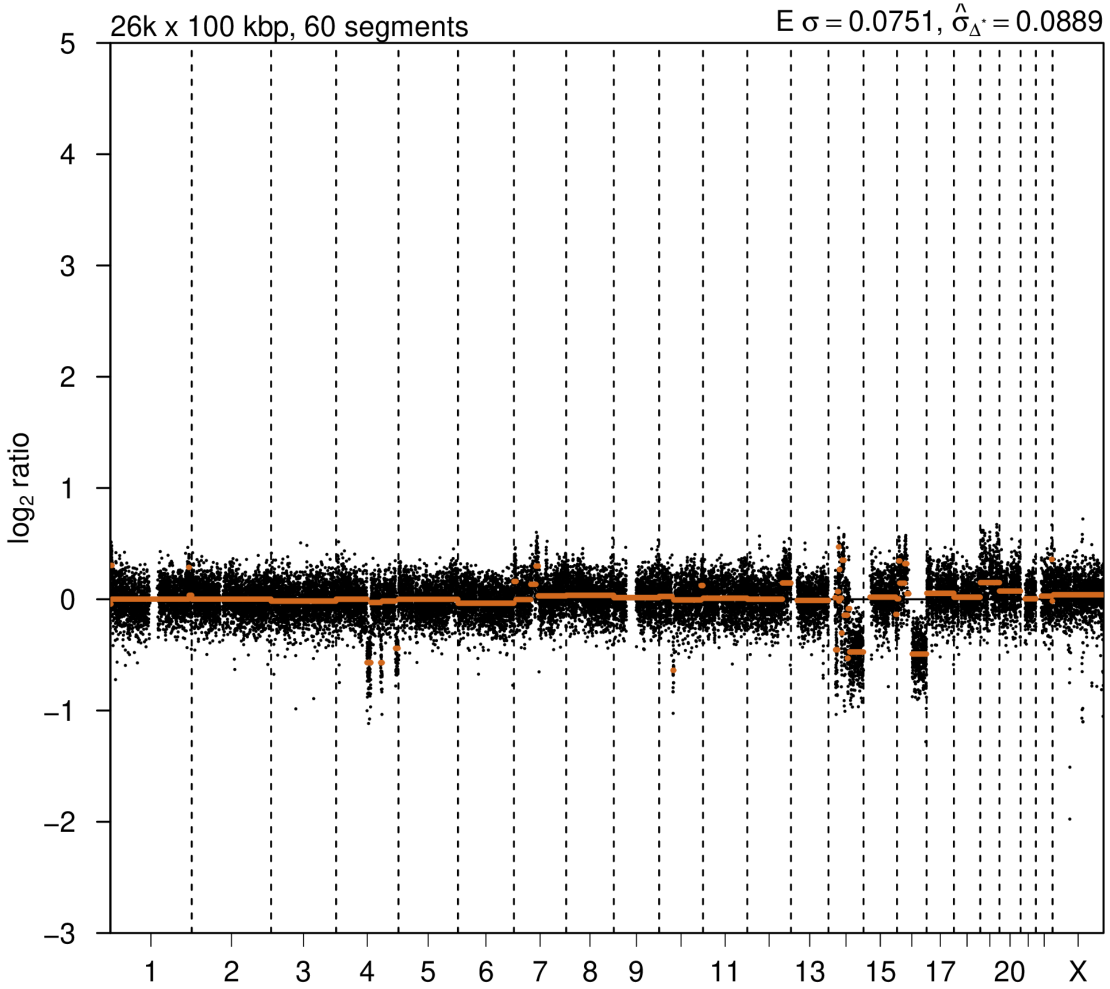


### 2nd event

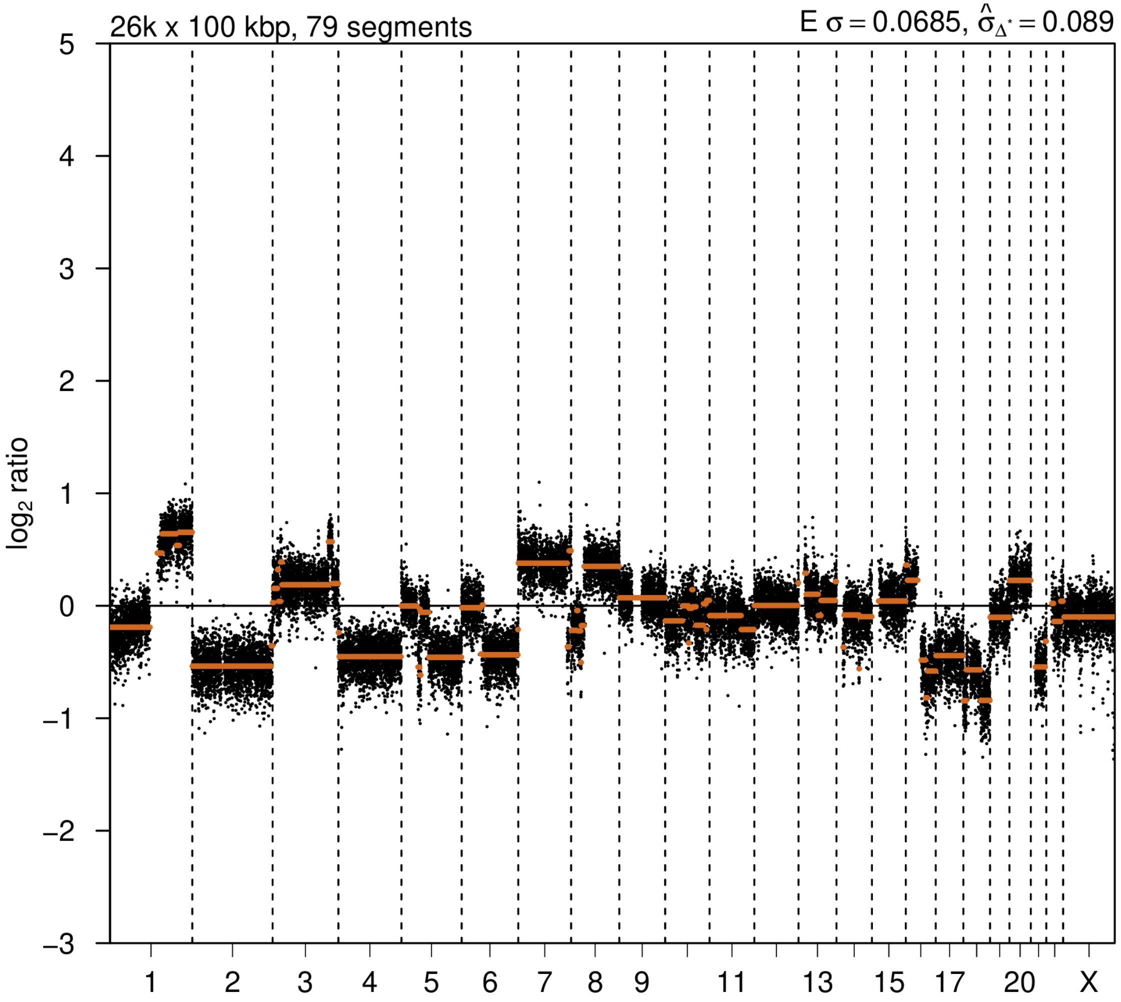

# P117

| Syn/Meta | Time from 1st surgery to 2nd event (Months) | Side 2nd event | Histology 2nd event | Surgery | AdjuvanT Treatment Pri (RT/ HT) | ER Pri | ER 2nd event | Her2 Pri | Her2 2nd event | Grade Pri | Grade 2nd event | Quadrant 2nd event | Margins | Screening | Clonality P value Copy N | Clonality P value Panel seq | Clonality P value WES | Final verdict |
|---|---|---|---|---|---|---|---|---|---|---|---|---|---|---|---|---|---|---|
| metachronous | 105 | Ipsilateral | IDC with DCIS | lumpectomy | None | + | - | - | - | 2 | 3 | NA | Clear | symptomatic | 0.317429406 | NA | NA | Unrelated |

### Primary event

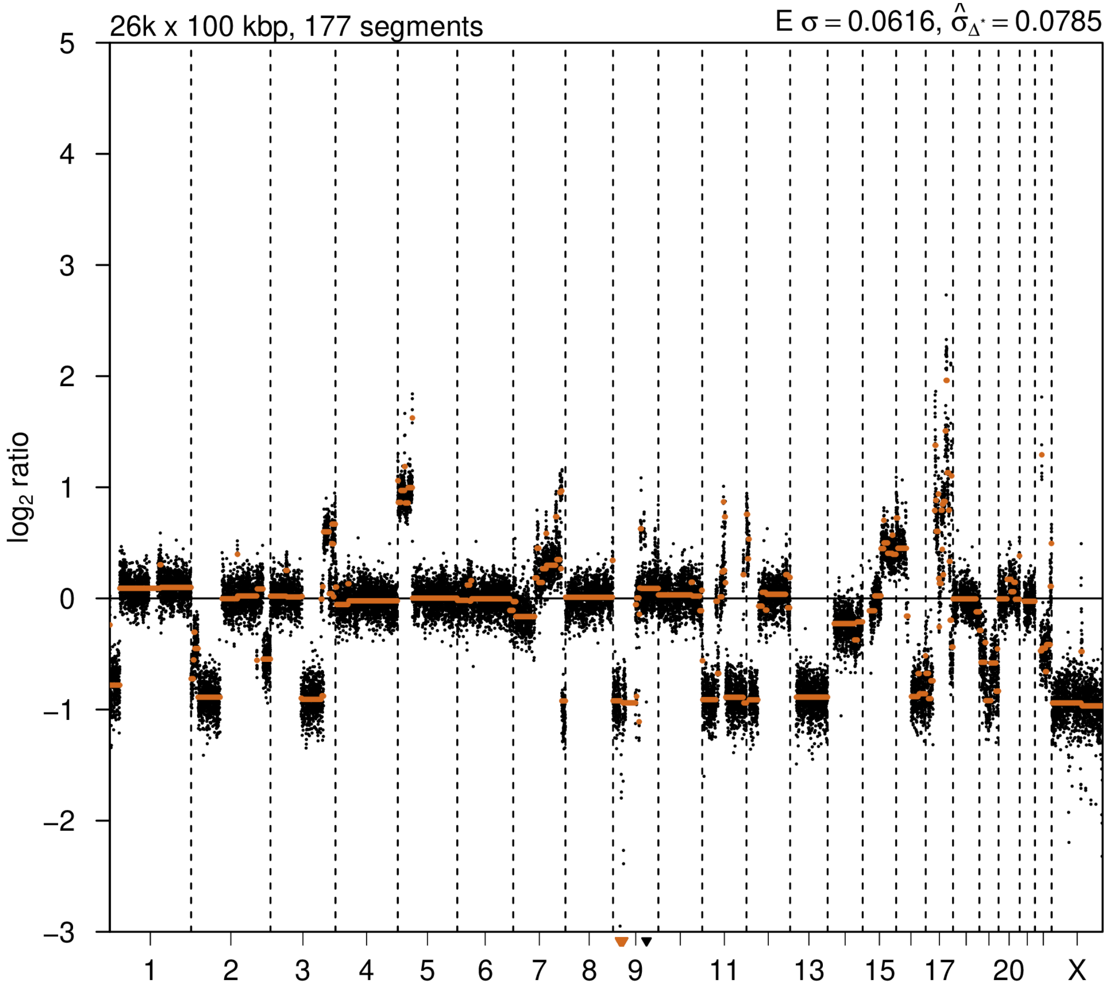


### 2nd event

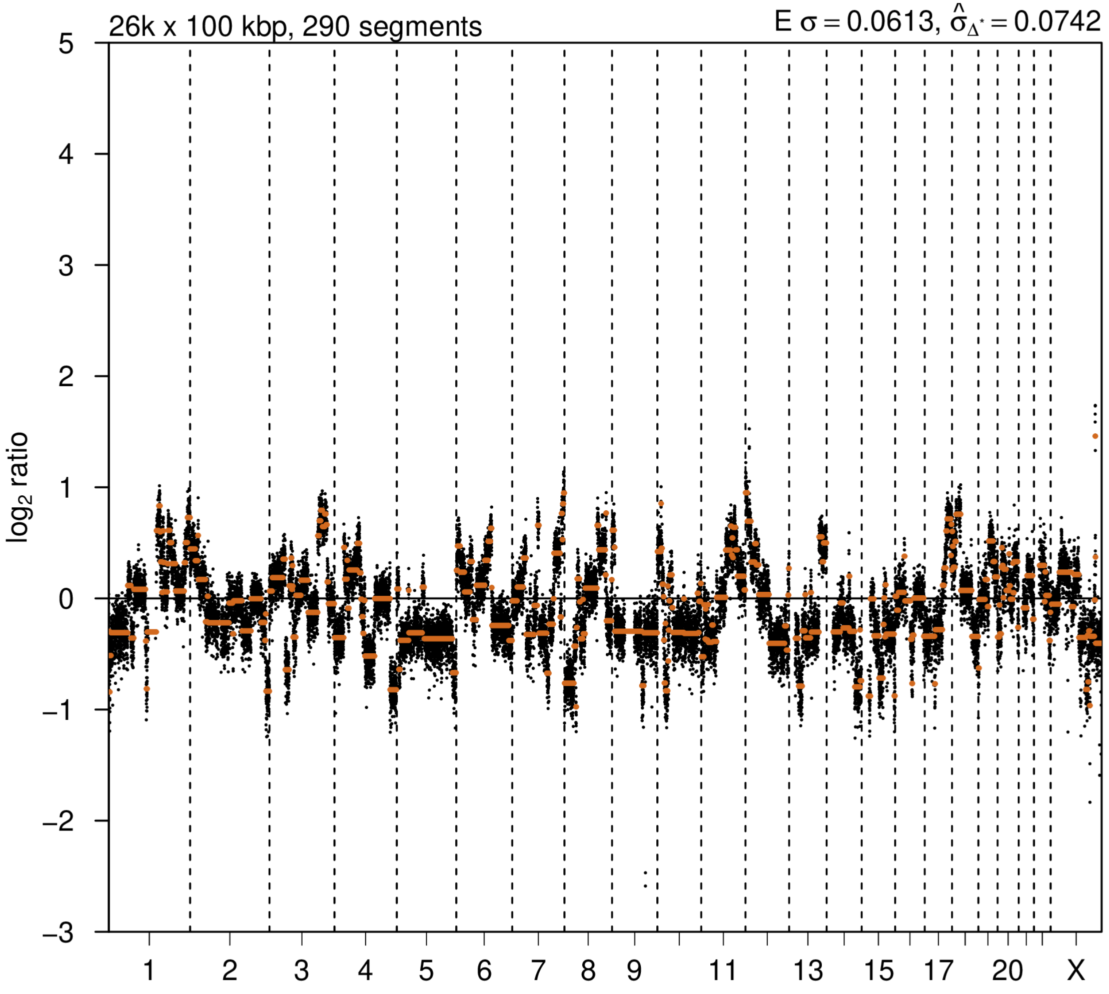

# P119

| Syn/Meta | Time from 1st surgery to 2nd event (Months) | Side 2nd event | Histology 2nd event | Surgery | AdjuvanT Treatment Pri (RT/ HT) | ER Pri | ER 2nd event | Her2 Pri | Her2 2nd event | Grade Pri | Grade 2nd event | Quadrant 2nd event | Margins | Screening | Clonality P value Copy N | Clonality P value Panel seq | Clonality P value WES | Final verdict |
|---|---|---|---|---|---|---|---|---|---|---|---|---|---|---|---|---|---|---|
| metachronous | 66 | Ipsilateral | IDC with DCIS | lumpectomy | None | + | - | + | - | 2 | 3 | at or adjacent to primary | Clear | screen-detected | 0.214540734 | NA | NA | Unrelated |

### Primary event

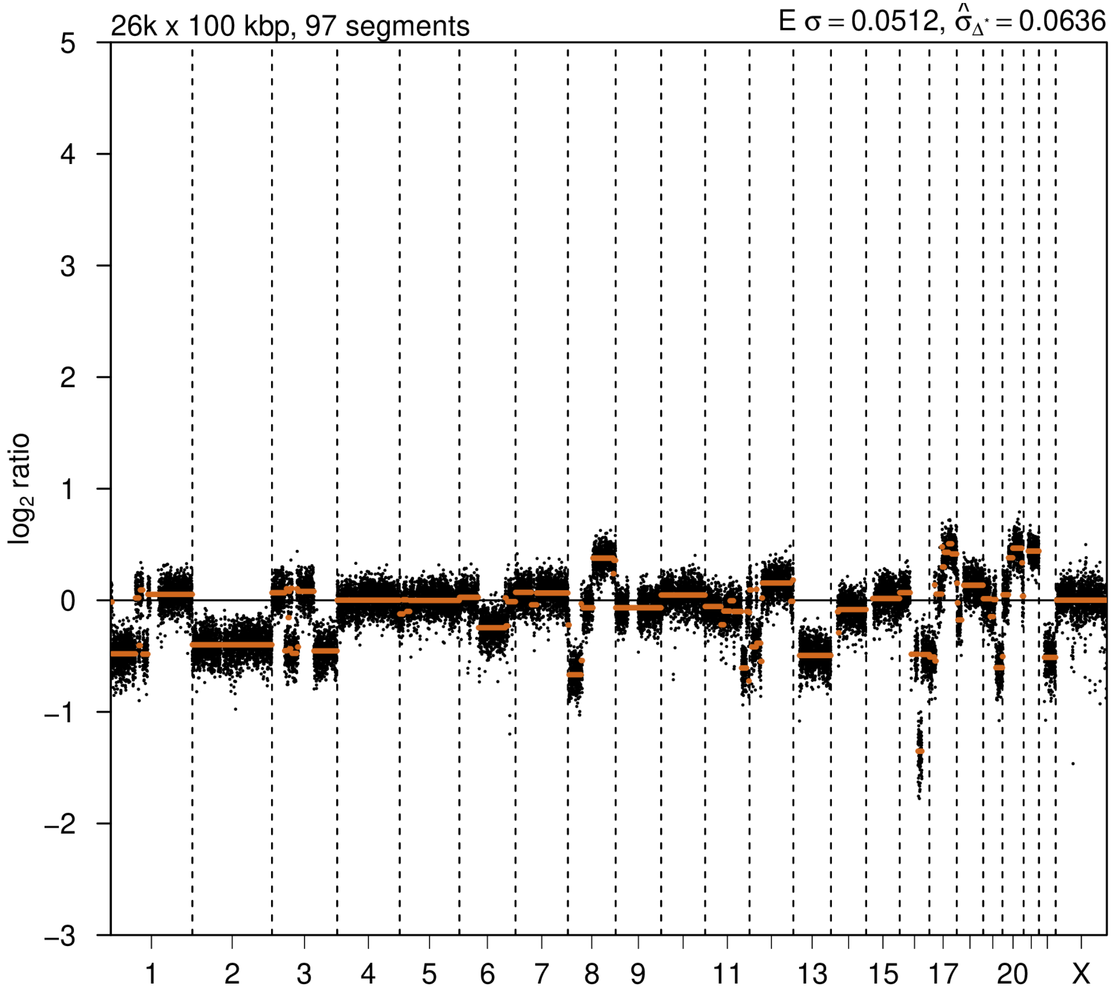


### 2nd event

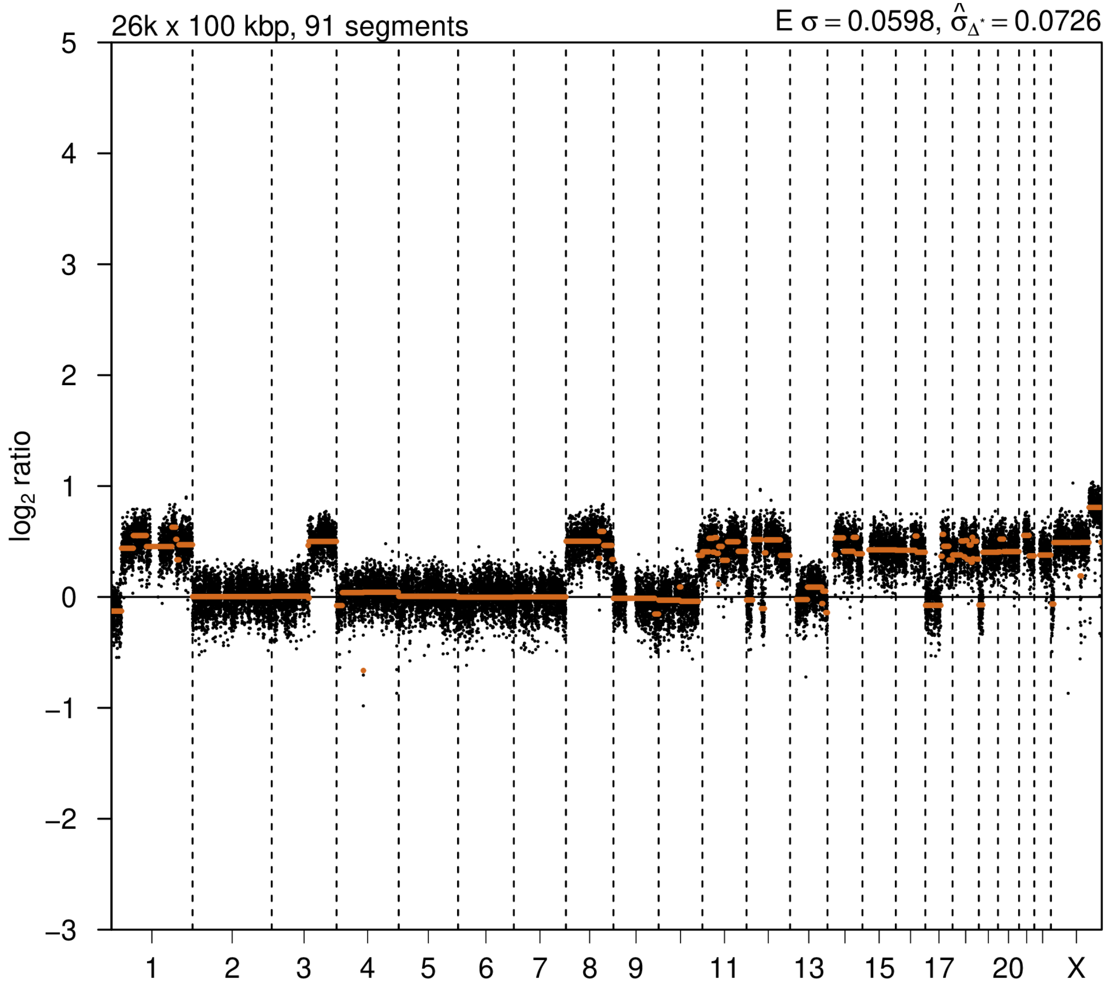

# P120

| Syn/Meta | Time from 1st surgery to 2nd event (Months) | Side 2nd event | Histology 2nd event | Surgery | AdjuvanT Treatment Pri (RT/ HT) | ER Pri | ER 2nd event | Her2 Pri | Her2 2nd event | Grade Pri | Grade 2nd event | Quadrant 2nd event | Margins | Screening | Clonality P value Copy N | Clonality P value Panel seq | Clonality P value WES | Final verdict |
|---|---|---|---|---|---|---|---|---|---|---|---|---|---|---|---|---|---|---|
| metachronous | 64 | Ipsilateral | IDC no DCIS | lumpectomy | None | + | + | - | - | 1 | 1 | at or adjacent to primary | Clear | screen-detected | 0.148003 895 | NA | NA | Unrelated |

### Primary event

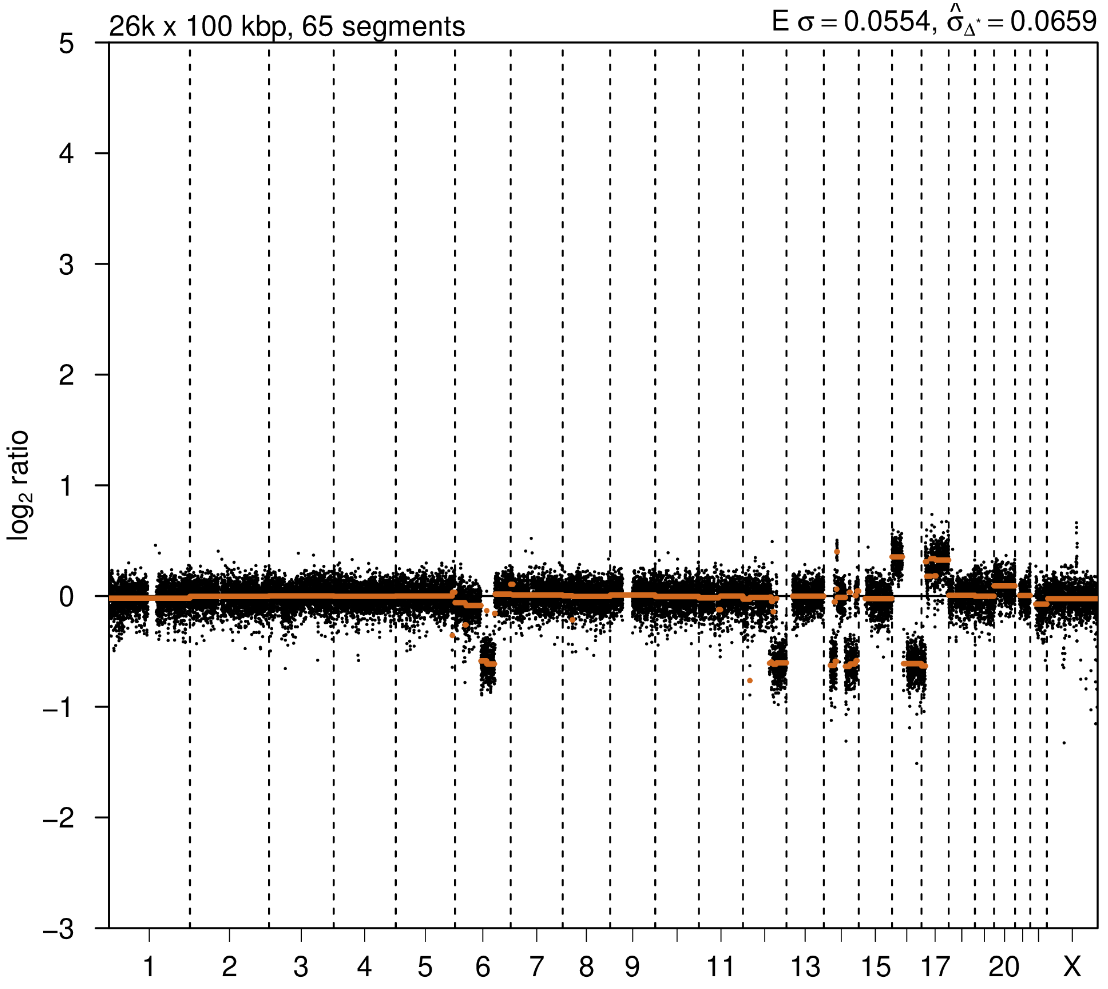


### 2nd event

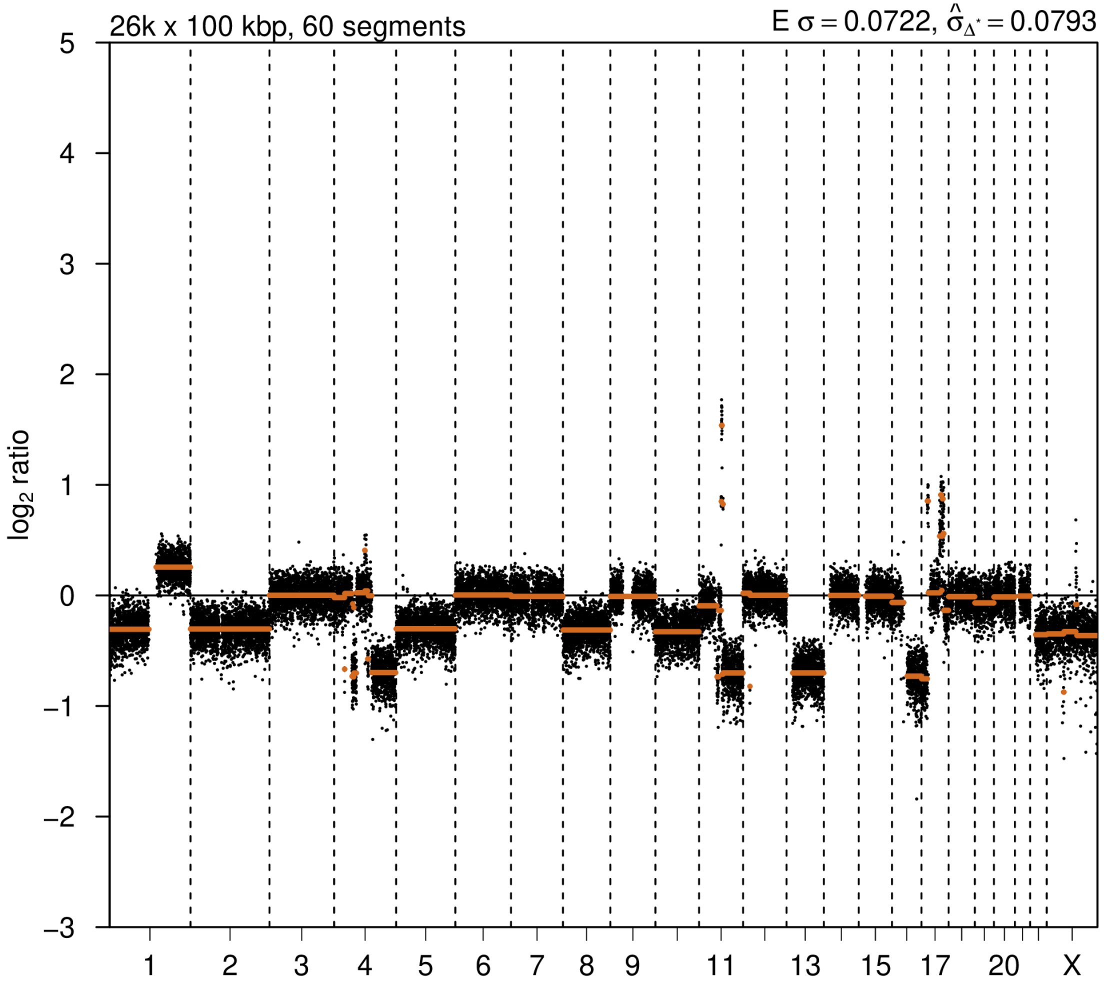

# P121

| Syn/Meta | Time from 1st surgery to 2nd event (Months) | Side 2nd event | Histology 2nd event | Surgery | AdjuvanT Treatment Pri (RT/ HT) | ER Pri | ER 2nd event | Her2 Pri | Her2 2nd event | Grade Pri | Grade 2nd event | Quadrant 2nd event | Margins | Screening | Clonality P value Copy N | Clonality P value Panel seq | Clonality P value WES | Final verdict |
|---|---|---|---|---|---|---|---|---|---|---|---|---|---|---|---|---|---|---|
| metachronous | 85 | Ipsilateral | IDC no DCIS | lumpectomy | None | + | - | - | + | 1 | 2 | at or adjacent to primary | involved | screen-detected | 0.804933 463 | 1 | NA | Unrelated |

### Primary event

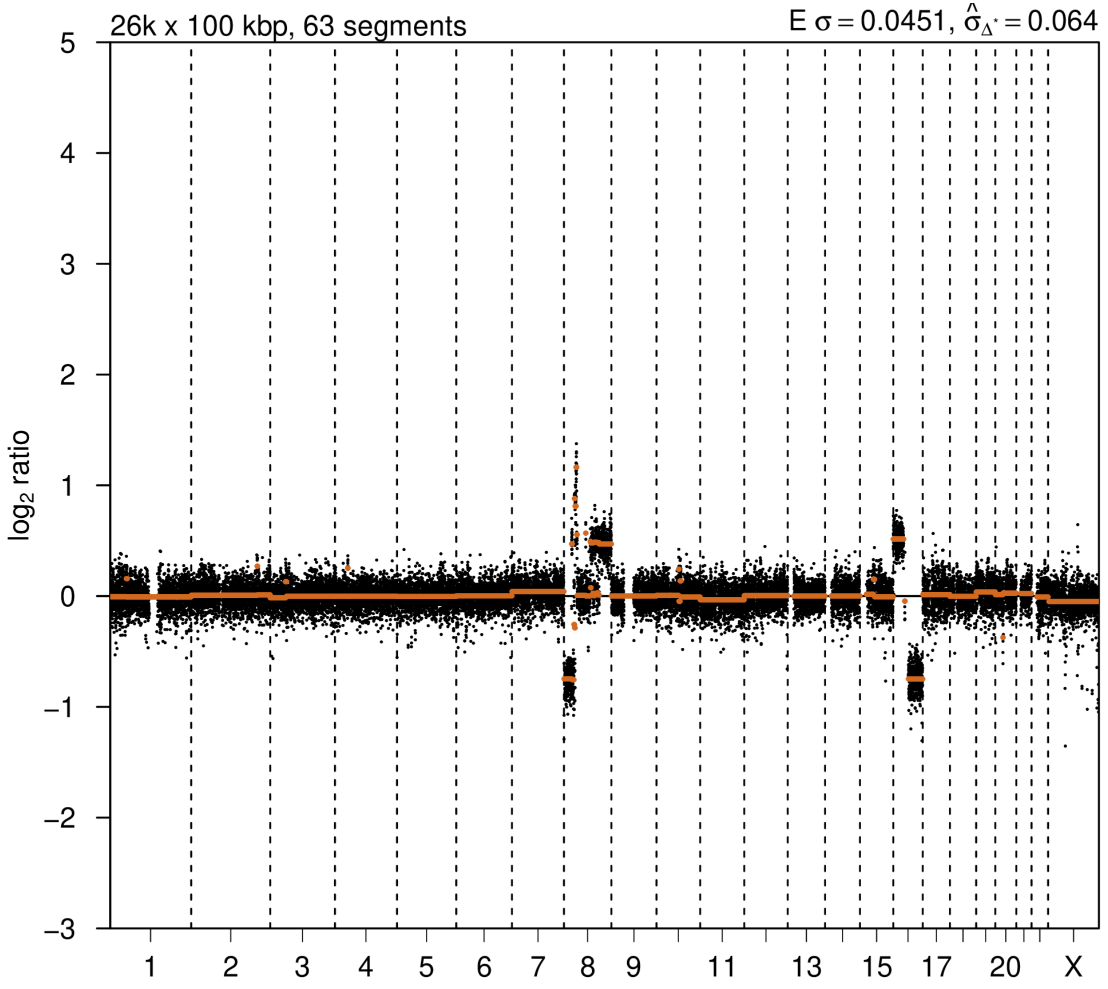


### 2nd event

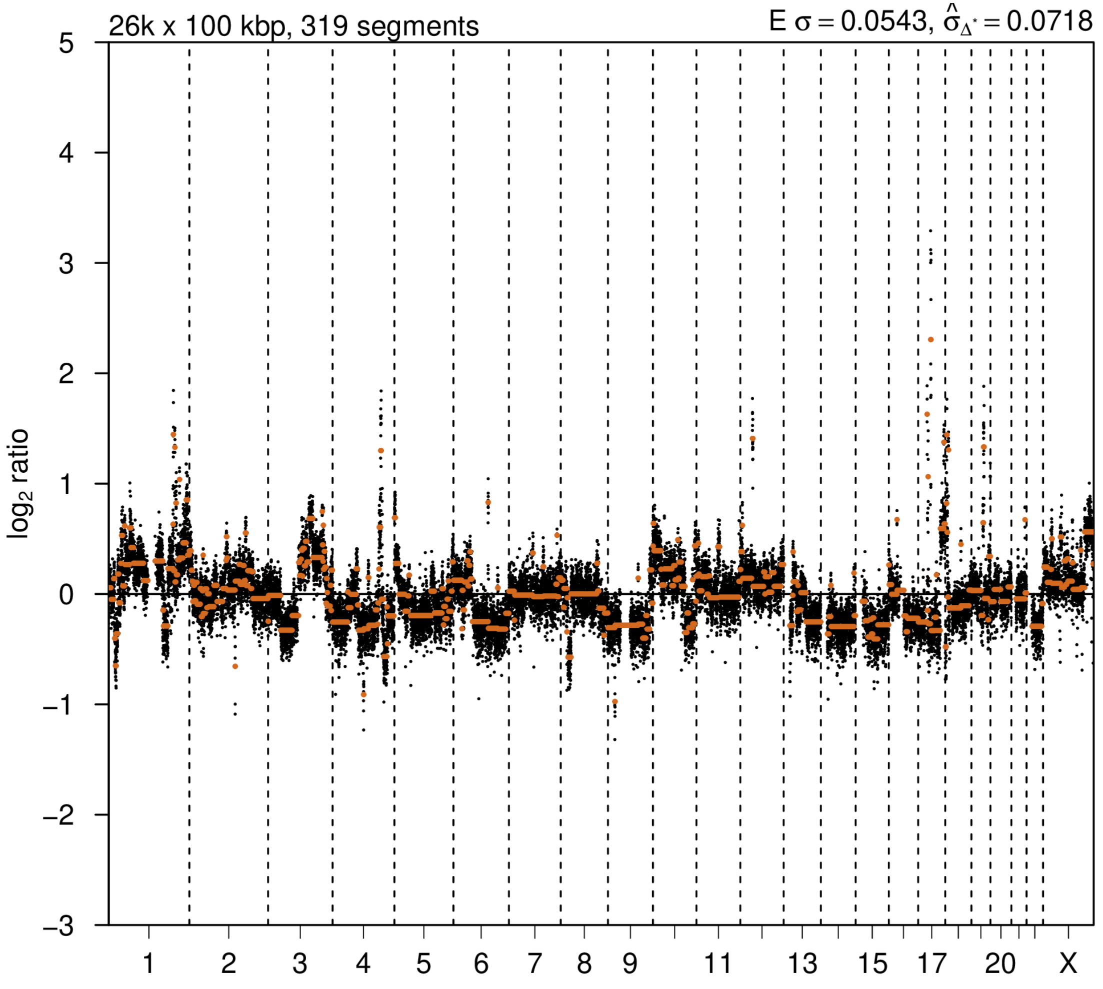

# P123

| Syn/Meta | Time from 1st surgery to 2nd event (Months) | Side 2nd event | Histology 2nd event | Surgery | AdjuvanT Treatment Pri (RT/ HT) | ER Pri | ER 2nd event | Her2 Pri | Her2 2nd event | Grade Pri | Grade 2nd event | Quadrant 2nd event | Margins | Screening | Clonality P value Copy N | Clonality P value Panel seq | Clonality P value WES | Final verdict |
|---|---|---|---|---|---|---|---|---|---|---|---|---|---|---|---|---|---|---|
| metachronous | 36 | Ipsilateral | IDC no DCIS | lumpectomy | None | + | + | - | - | 1 | 3 | distant from primary | Clear | screen-detected | 0.331061344 | NA | NA | Unrelated |

### Primary event

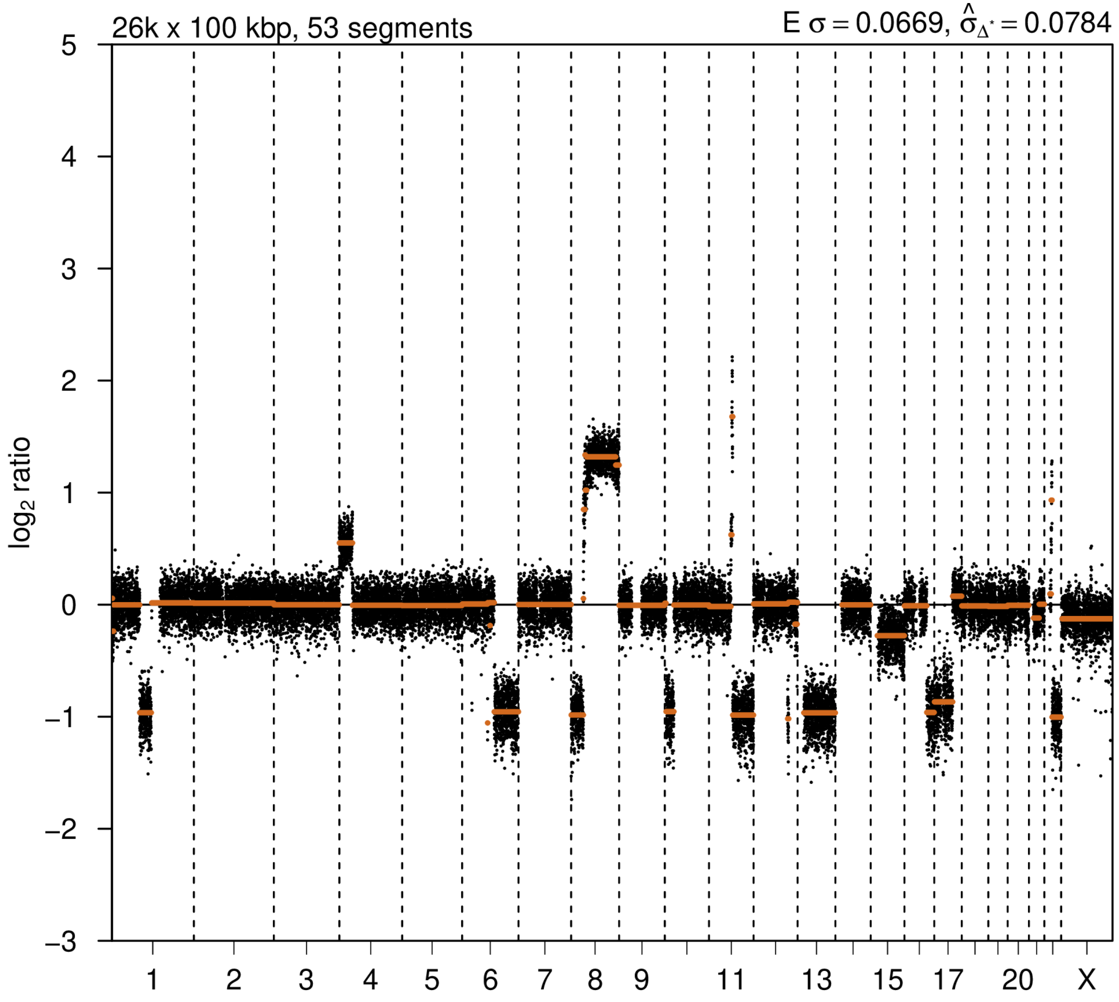


### 2nd event

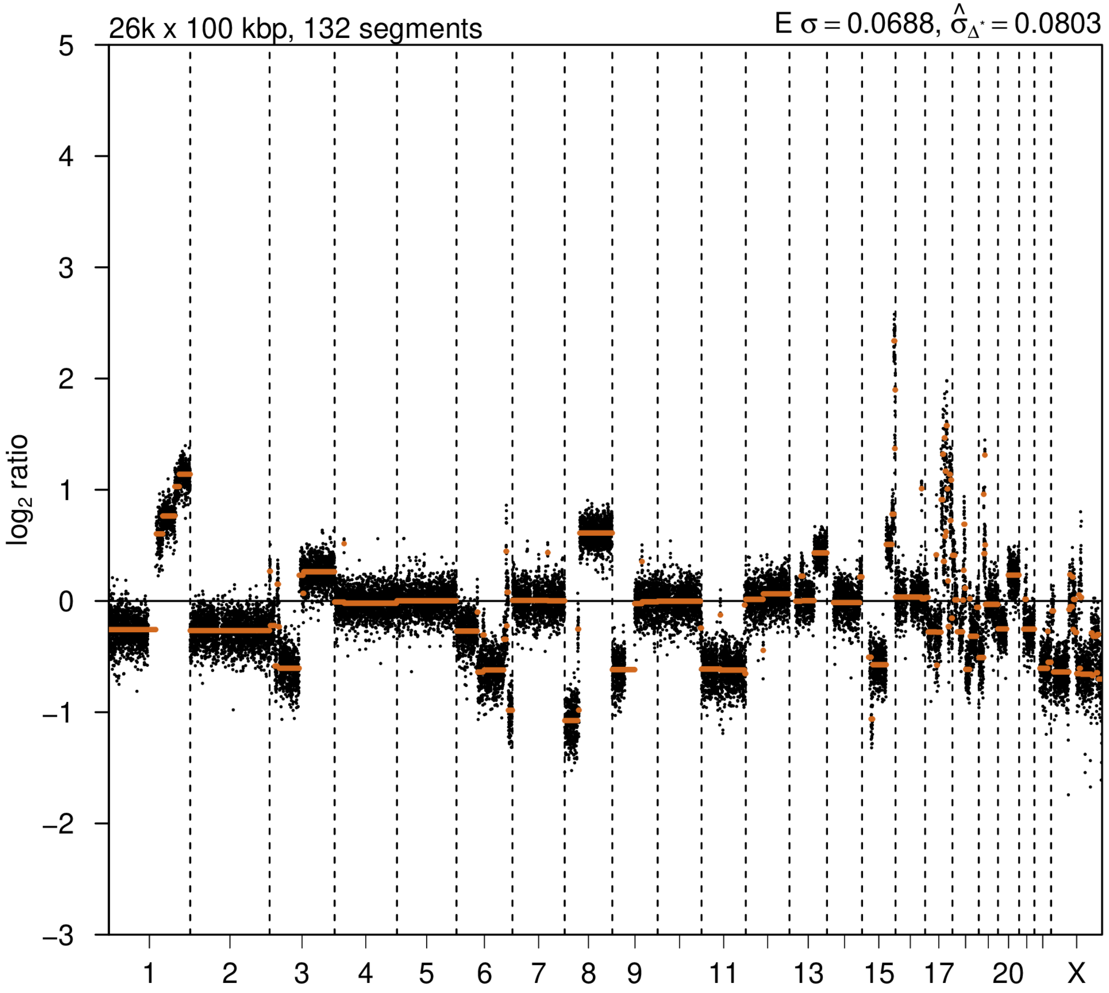

# P125

| Syn/Meta | Time from 1st surgery to 2nd event (Months) | Side 2nd event | Histology 2nd event | Surgery | AdjuvanT Treatment Pri (RT/ HT) | ER Pri | ER 2nd event | Her2 Pri | Her2 2nd event | Grade Pri | Grade 2nd event | Quadrant 2nd event | Margins | Screening | Clonality P value Copy N | Clonality P value Panel seq | Clonality P value WES | Final verdict |
|---|---|---|---|---|---|---|---|---|---|---|---|---|---|---|---|---|---|---|
| metachronous | 52 | Ipsilateral | IDC with DCIS | lumpectomy | None | + | + | - | - | 2 | 1 | NA | Clear | symptomatic | 0.64459591 | 1 | NA | Unrelated |

### Primary event

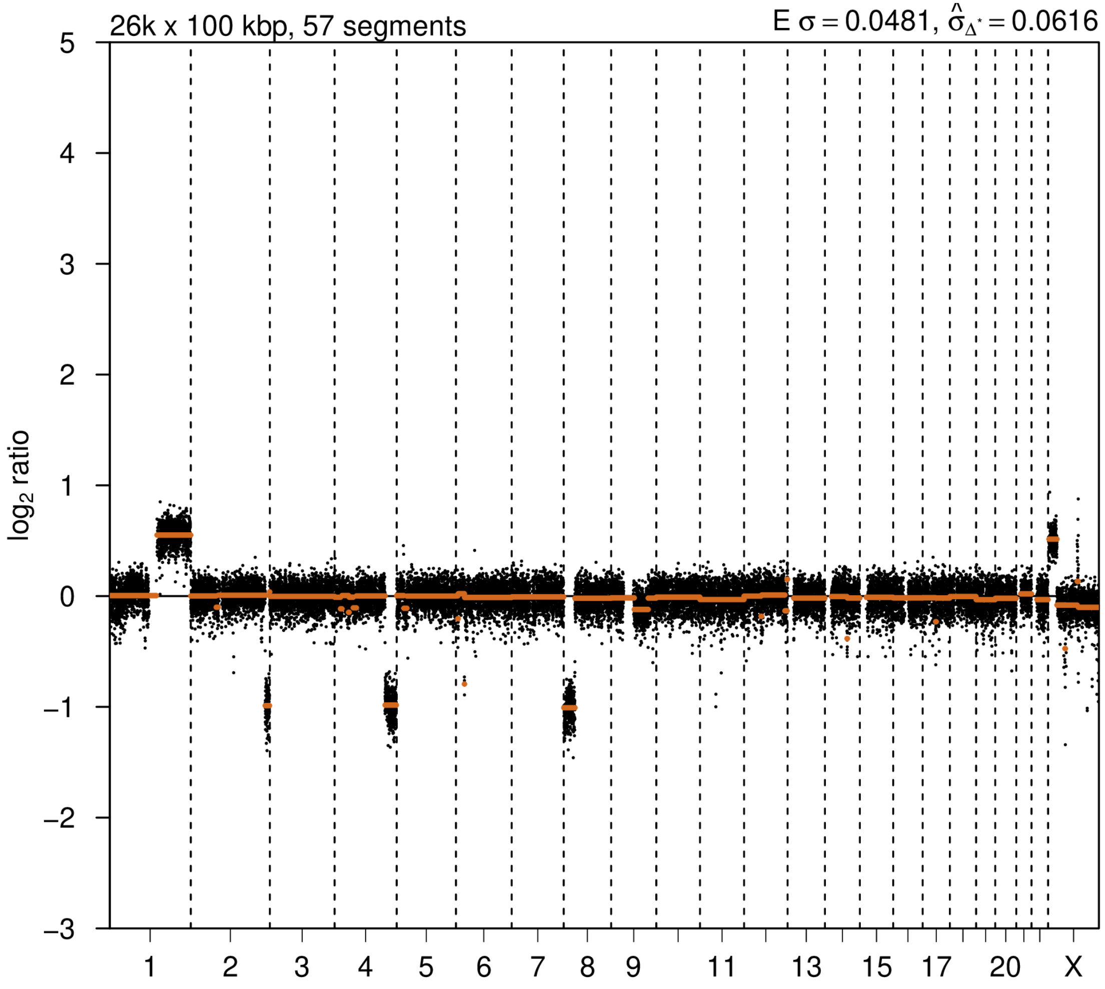


### 2nd event

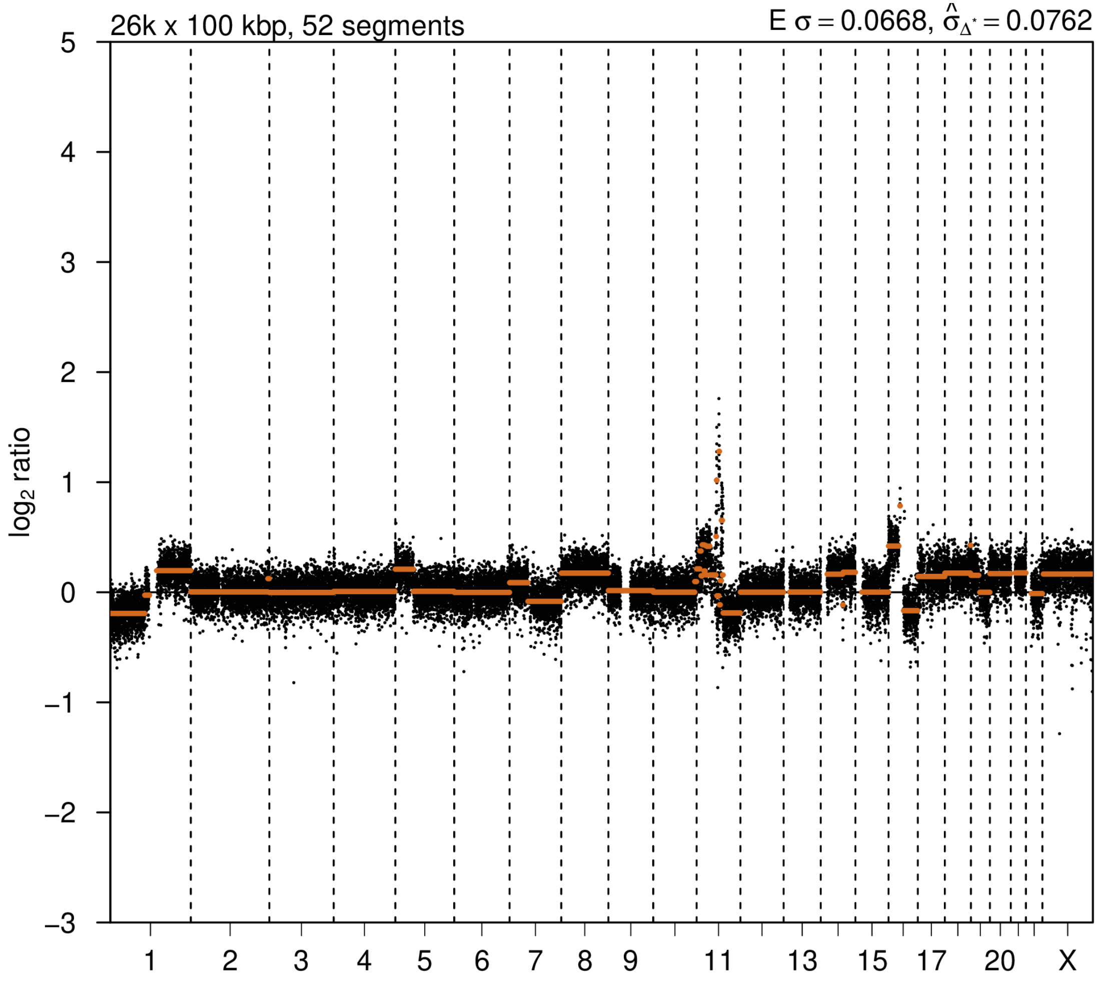

# P126

| Syn/Meta | Time from 1st surgery to 2nd event (Months) | Side 2nd event | Histology 2nd event | Surgery | AdjuvanT Treatment Pri (RT/ HT) | ER Pri | ER 2nd event | Her2 Pri | Her2 2nd event | Grade Pri | Grade 2nd event | Quadrant 2nd event | Margins | Screening | Clonality P value Copy N | Clonality P value Panel seq | Clonality P value WES | Final verdict |
|---|---|---|---|---|---|---|---|---|---|---|---|---|---|---|---|---|---|---|
| metachronous | 55 | Ipsilateral | IDC with DCIS | lumpectomy | None | - | + | + | - | 3 | 3 | at or adjacent to primary | Clear | screen-detected | 0.931191 172 | NA | NA | Unrelated |

### Primary event

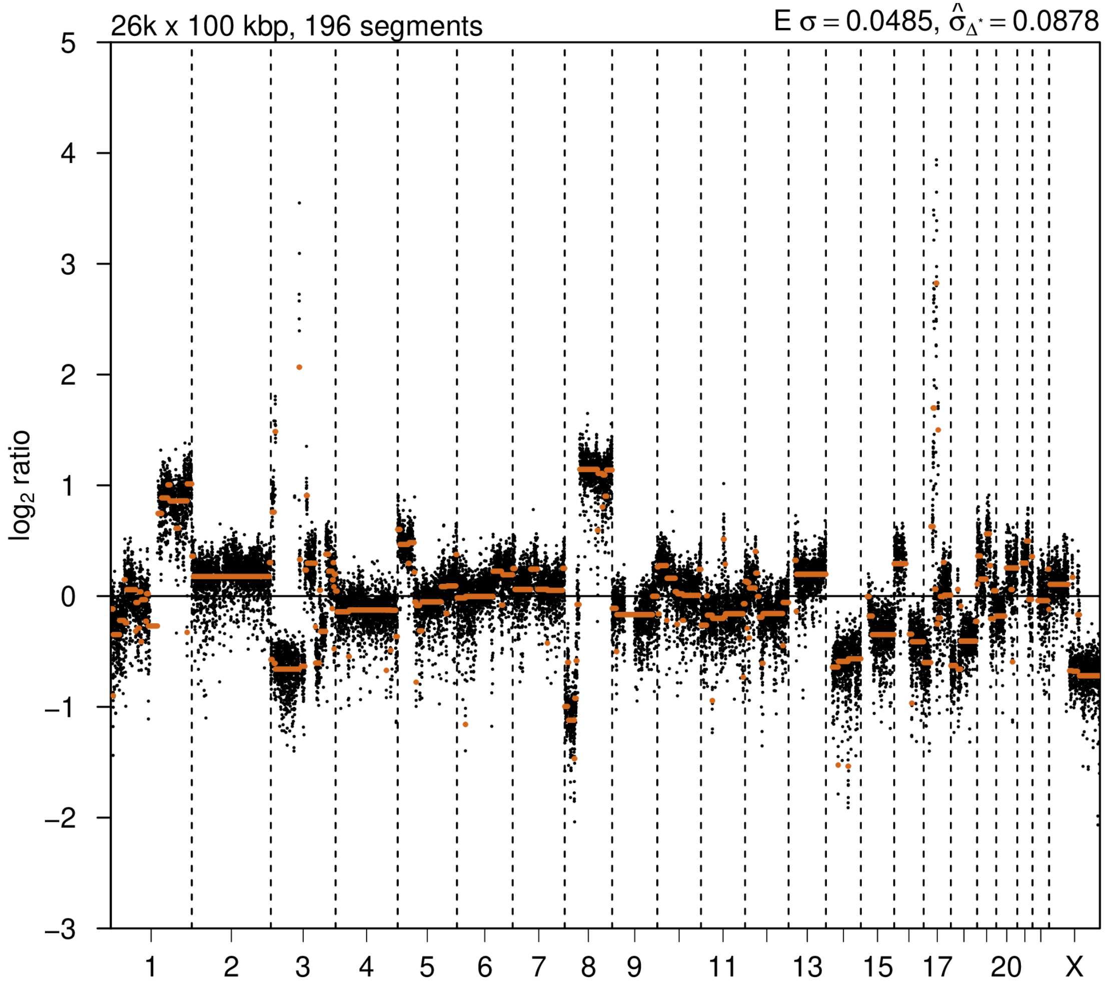


### 2nd event

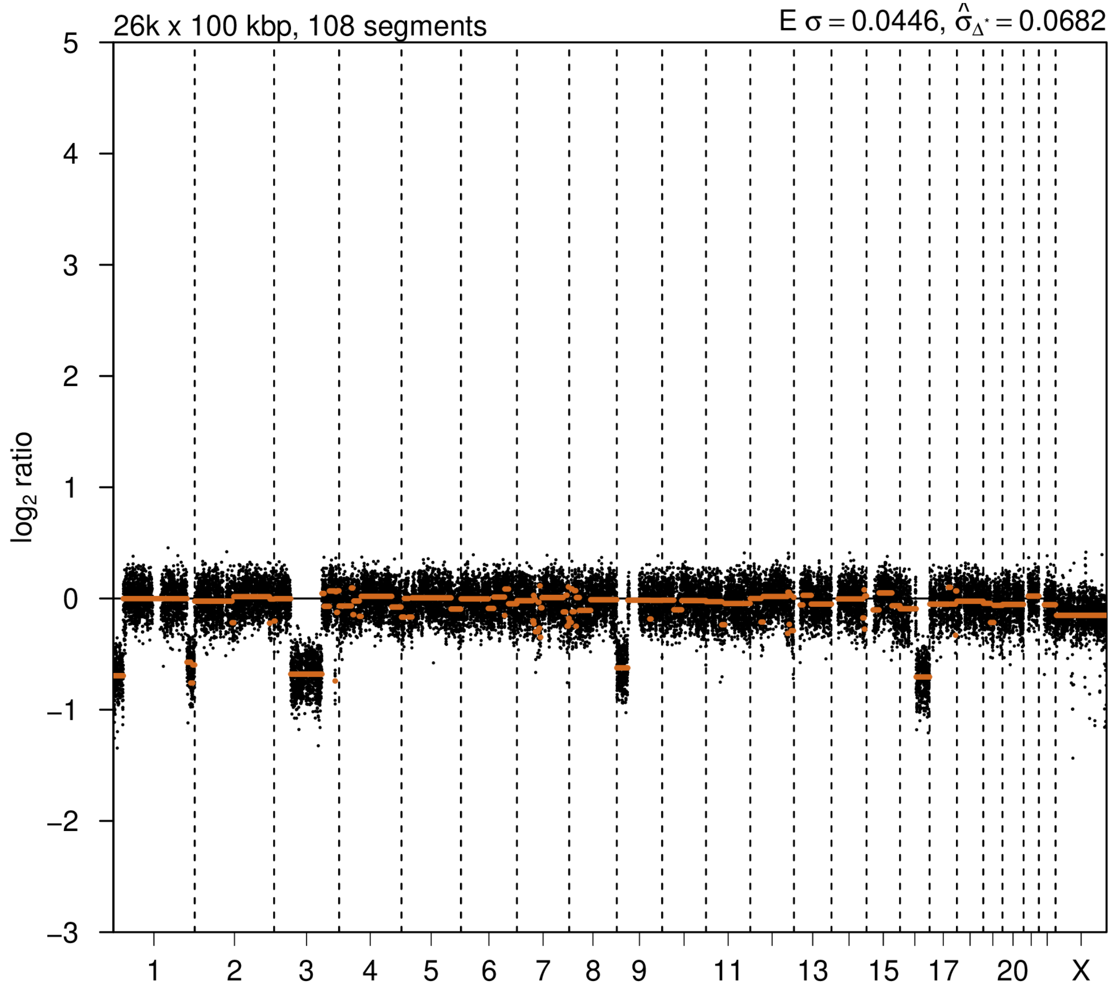

# P129

| Syn/Meta | Time from 1st surgery to 2nd event (Months) | Side 2nd event | Histology 2nd event | Surgery | AdjuvanT Treatment Pri (RT/ HT) | ER Pri | ER 2nd event | Her2 Pri | Her2 2nd event | Grade Pri | Grade 2nd event | Quadrant 2nd event | Margins | Screening | Clonality P value Copy N | Clonality P value Panel seq | Clonality P value WES | Final verdict |
|---|---|---|---|---|---|---|---|---|---|---|---|---|---|---|---|---|---|---|
| metachronous | 23 | Ipsilateral | IDC with DCIS | lumpectomy | None | - | - | + | - | 3 | 2 | NA | Clear | screen-detected | 0.581953911 | 1 | 1 | Unrelated |

**Primary event**

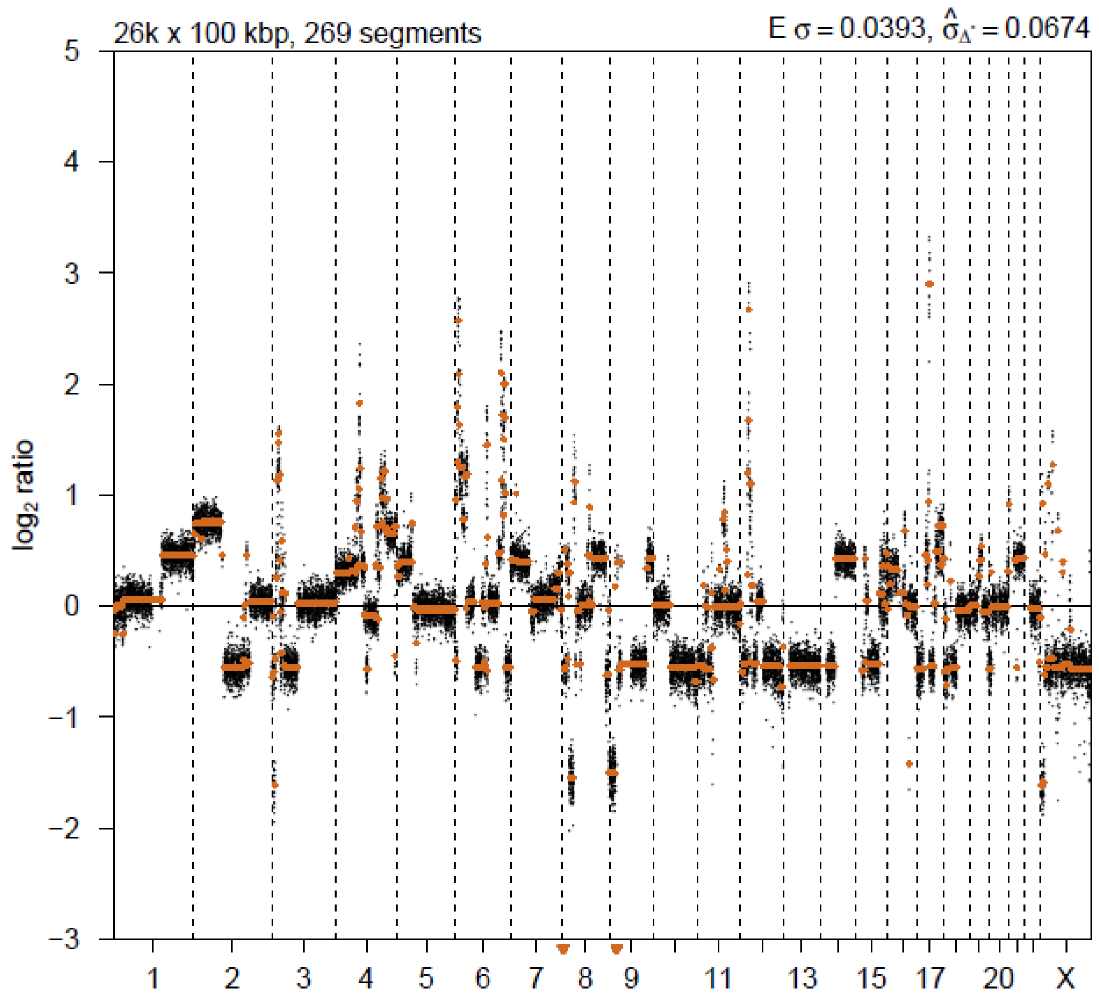


**2nd event**

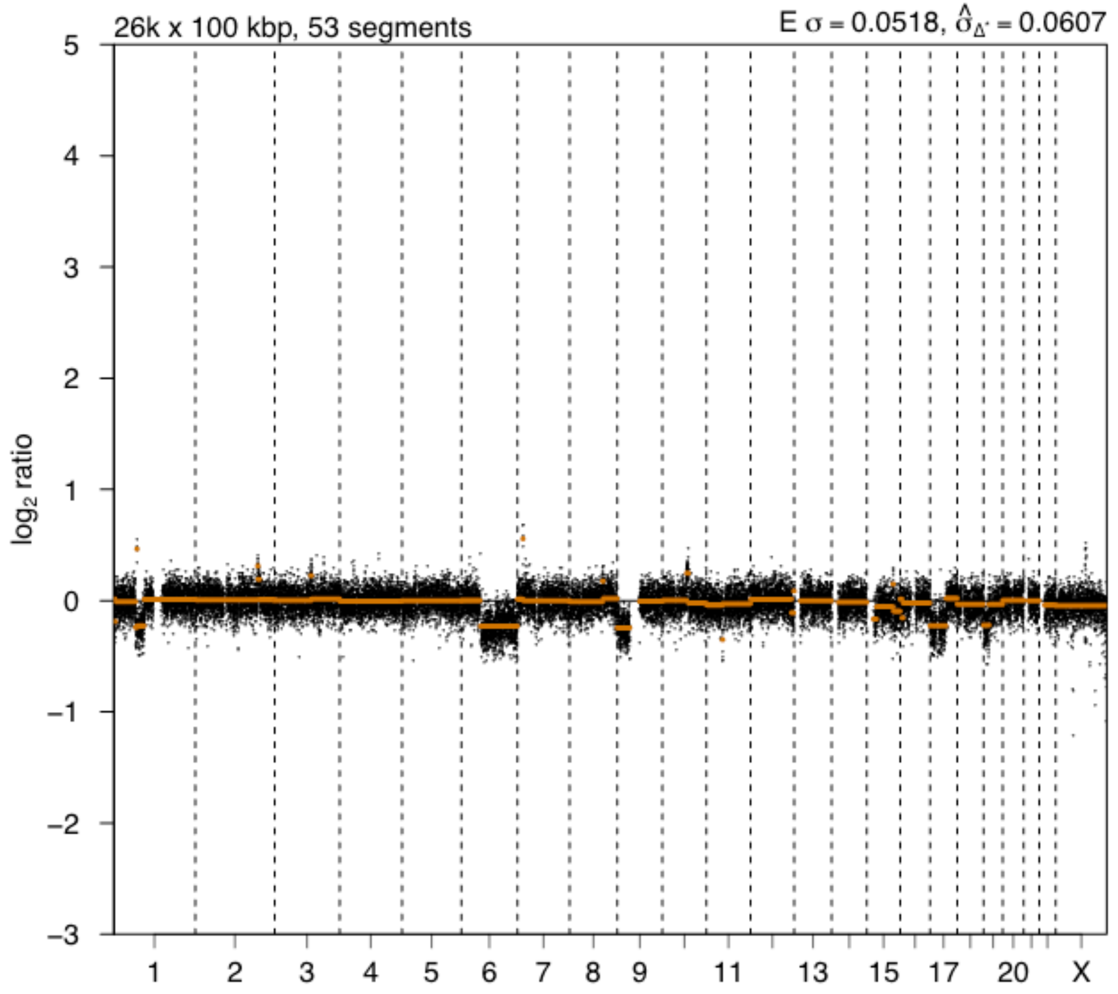
