## Supplementary File 2 for "Genomic profiling defines variable clonal relatedness between invasive breast cancer and primary ductal carcinoma *in situ*"

**Supplementary File 2 - PanelSeq performance and filtering strategy for NKI samples**

*Performance PanelSeq*

Figure SF1 and SF2 show a representative example of the performance of the custom 53-gene panel. The graphs contain the data of 40 DCIS samples.

| **A.** | 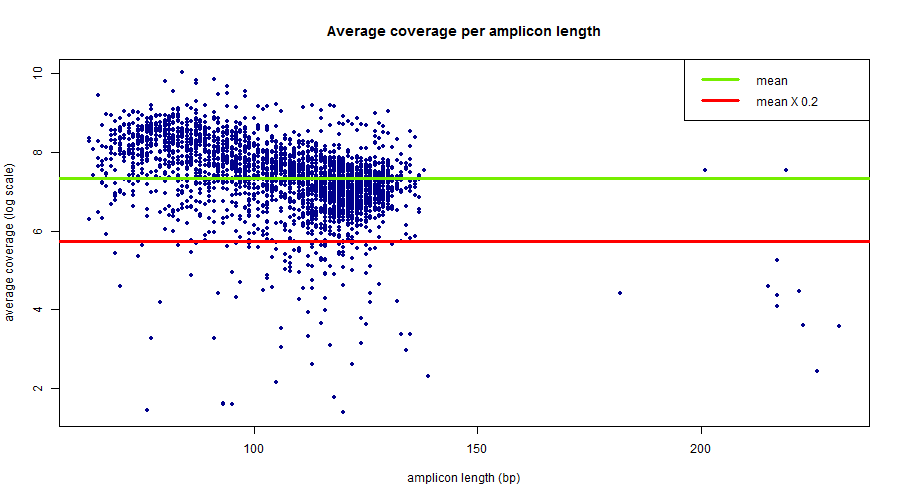 |
| --- | --- |
| **B.** | 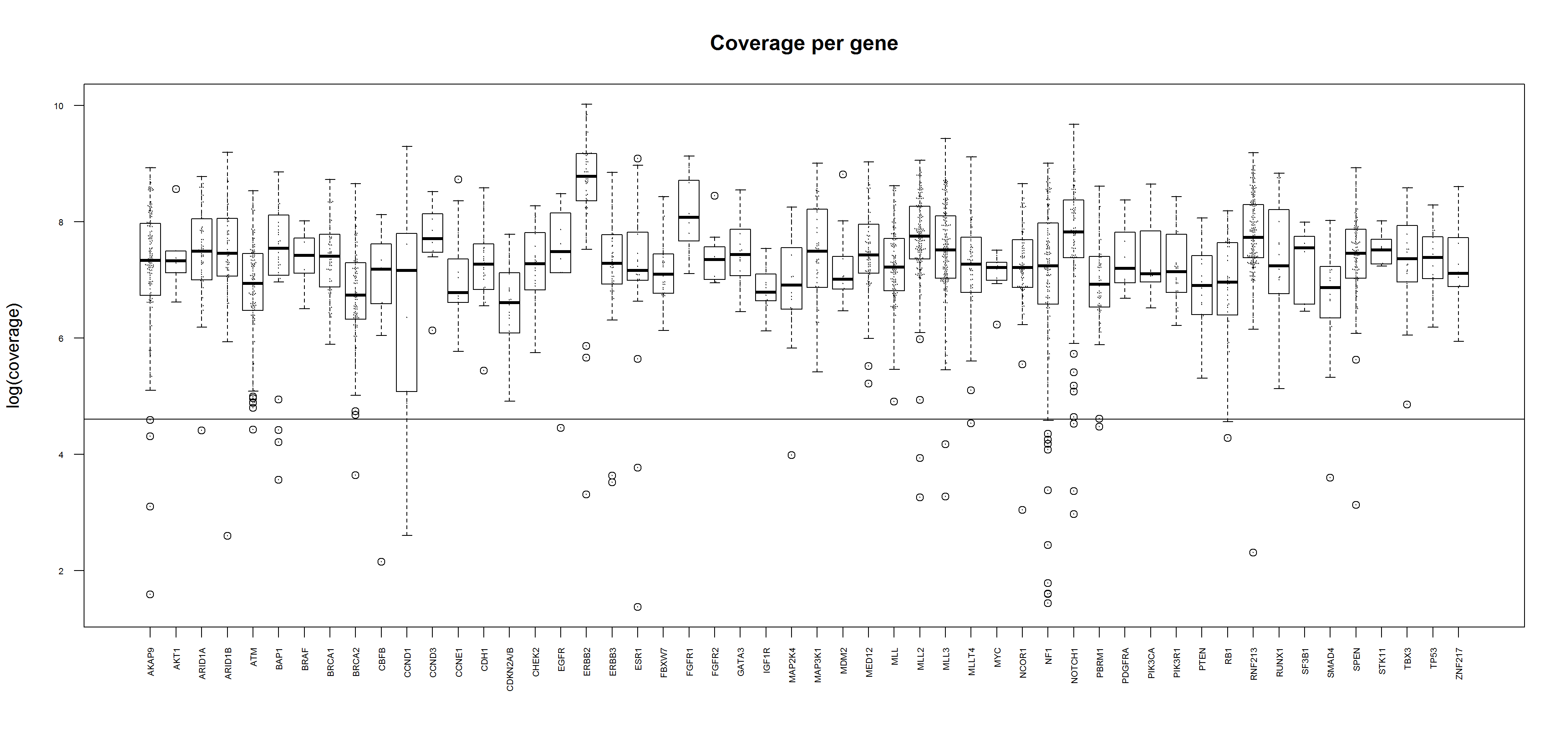 |
| **Figure SF1.** Performance of the custom 53-gene panel. **A.** Amplicon coverage by amplicon length; **B.** Amplicon coverage per gene. Black line represents a coverage of 100X. Average coverage >2000X. | |

*Filtering strategy PanelSeq data*

To narrow down the IonTorrent sequencing data to the true somatic variants found in the DCIS and IBC samples, the following steps were taken. First, we included variants with a variant allele frequency >5%, coverage >100x, and a quality (QUAL) of >1000 (variants of QUAL <1000 were mostly low frequency variants, see Figure SF2A). Variants not found in GNOMAD and GoNL and found in somatic ClinVar were included. Figure SF2B). Variants found in <90% of the samples and located within the coding sequence of the genome were included. All included somatic variants were merged to one variant list. This list of variants was subjected to a final check via manual assessment in IGV. Lastly, the positions of variants called solely in the IBC lesions were checked in the matching DCIS lesion and vice versa, to reassure the absence of this variant in the paired sample.

| **A.** | 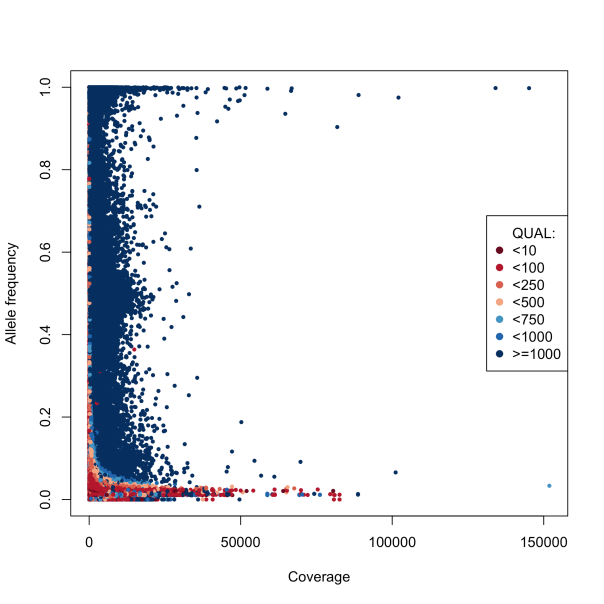 | **B.** | 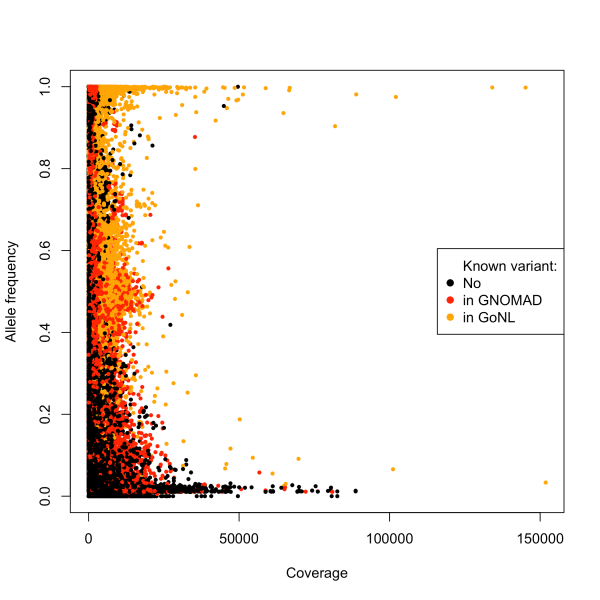 |
| --- | --- | --- | --- |
| **Figure SF2.** Visualization of variants selected for exclusion. **A.** Variants with a quality (QUAL) of <1000 were excluded (all colors except dark blue). **B.** Variants found in in GNOMAD (red) and GoNL (orange) were excluded. | | | |
